# Optimization of Pharmacist Medication Management and Mortality in the Intensive Care Unit

**DOI:** 10.64898/2025.12.31.25342855

**Authors:** Susan E. Smith, Kelli R. Henry, Mojdeh S. Heavner, Zhetao Chen, Xianyan Chen, John W. Devlin, David J. Murphy, Greg S. Martin, Marisha Burden, Brian Murray, Andrea Sikora, Optimizing Pharmacist Team-Integration for ICU Patient Management (OPTIM) Investigator Team (see supplemental author list)

## Abstract

**Rationale:** Medication-related morbidity due to inappropriate prescribing, delays in appropriate treatment, and adverse drug events contributes to ICU patient mortality. Comprehensive medication management (CMM) is a care standard provided by pharmacists in collaboration with the interprofessional team. Optimizing ICU pharmacist workload via the pharmacist-to-patient ratio while ensuring daily CMM may reduce mortality.

**Research Question:** Is ICU pharmacist staffing, measured by the pharmacist-to-patient ratio and absence of comprehensive medication management (CMM), associated with in-hospital mortality among critically ill adults?

**Study Design and Methods:** Adults admitted to an ICU from 64 centers (in the United States, Jordan, and Saudi Arabia) were enrolled in a multicenter observational study that collected patient and team staffing data from August 2023-January 2025. The primary outcome was in-hospital mortality. The primary exposure was the patient-level pharmacist-to-patient ratio averaged over the ICU stay. A secondary exposure was the absence of CMM for at least 1 day. Multivariable generalized estimating equations (GEE) were used to estimate associations with in-hospital mortality, accounting for clustering by center and adjusting for patient-, ICU-, and hospital-level covariates.

**Results:** The 28,795 patients enrolled had a median (IQR) pharmacist-to-patient ratio of 1:17 (13-23). For every one patient increase in the pharmacist-to-patient ratio, the odds of mortality increased by 1% (Odds Ratio (OR) 1.01, 95% Confidence Interval (CI) 1.00-1.01, p=0.04). Patients without pharmacist CMM for 1 day had an increased risk of mortality of 20% (OR 1.20, 95% CI 1.03-1.40, p=0.02). The odds of hospital mortality were lower in a pharmacist-to-patient ratio of 1:7-15 compared to 1:16-46 (OR 1.10, 95% CI 1.00-1.22).

**Interpretation:** Increasing pharmacist-to-patient ratios and the absence of pharmacist CMM every day were both associated with an increased risk of in-hospital mortality.

## Background

Medication-related morbidity due to inappropriate prescribing, polypharmacy, delays in appropriate treatment, and adverse drug events (ADEs) are major contributors to mortality in acutely ill hospitalized patients.^1^ Medication errors occur in 6.5% of hospitalized adults.^2^ The Institute of Medicine found hospital deaths due to preventable medication causes exceed deaths attributed to motor vehicle accidents, with associated costs up to $50 billion.^3^ All of this underscores the importance of intentional medication management in acutely ill hospitalized patients.^4–11^

Comprehensive medication management (CMM) provided by clinical pharmacists integrated with interprofessional teams is a well-established intervention for acutely ill hospitalized patients that has become the standard of care across the spectrum of care given it improves time to appropriate drug treatment, prevents medication errors and ADEs, and reduces mortality.^12–15^ Defined as **“**a patient-centered approach to optimize medication use and improve patient health outcomes…that ensures patients’ medications (both prescription and nonprescription) are individually assessed to determine each has an appropriate indication, is effective and achieving defined patient and/or clinical goals, is safe given comorbidities and other medications being taken, and that the patient is able to take the medication as intended and adhere to the prescribed regimen,” CMM is intended as a cognitive service aimed at individualizing medication therapy to maximize benefit and minimize harm.^16^ Multiple interprofessional societies have endorsed recommendations affirming pharmacists as essential members of the healthcare team to provide CMM, specifically citing the importance of reasonable patient care volumes and attendance on interprofessional rounds to improve CMM quality.^12,14,17–19^

Healthcare professional workload and delivered care quality are inter-related: nurse and physician staffing practices that result in high workloads are associated with decreased care quality and increased medication errors and ADEs.^20–27^ Because CMM is a cognitive service that occurs in the context of interprofessional rounds, pharmacist staffing practices have the potential to affect the quality of CMM delivered.^28,29^ However, the impact of clinical pharmacist staffing practices on patient-centered outcomes remains largely untested. With most medication errors and ADEs being preventable,^8,30,31^ and unit-based, clinical pharmacist patient staffing being modifiable by administrators, establishing the pharmacist-to-patient staffing ratio that is associated with improved patient outcomes is important to determine.

Patients managed in the intensive care unit (ICU) have the highest rate of medication errors and ADEs among acutely ill hospitalized patients.^32–36^ Rapidly fluctuating end-organ function and the elevated complexity of medication regimens make the ICU the ideal setting to evaluate the role of clinical pharmacist staffing as a mitigating factor to CMM delivery and patient-centered outcomes. Prior evaluations of ICU pharmacist staffing practices have identified a low pharmacist-to-patient ratio and the attendance of pharmacists on interprofessional rounds (where CMM primarily takes place) as quantitative factors associated with improved outcomes; however, these evaluations failed to consider crucial elements, including underlying patient severity of illness, medication therapy intensity, and the composition of the ICU interprofessional team.^13,28,29,37,38^

Thus, we aimed to establish the optimal ICU pharmacist-to-patient staffing ratio through a rigorous evaluation of patient, institutional, and team factors. Our goal was to identify quantitative staffing indicators associated with optimized patient-centered outcomes to inform improvements in clinical practice and future interventional trials. As conceptualized in **Supplementary Appendix eFigure 1**, our primary hypothesis was that lower patient volume per ICU-based pharmacist (described in terms of the pharmacist-to-patient ratio) is associated with improved survival.

**Figure 1.**
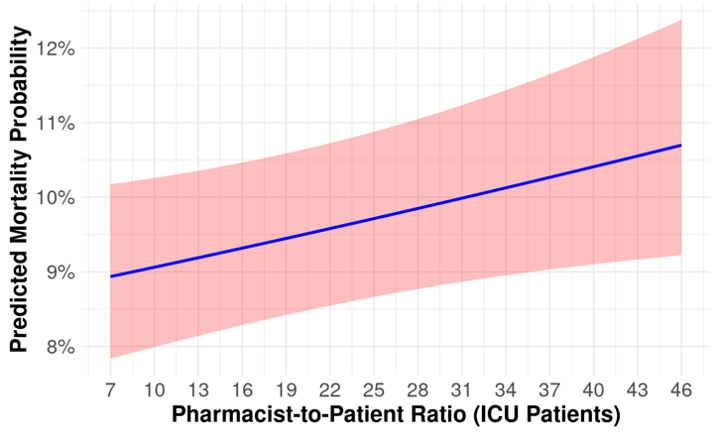
Consort Diagram. ICU: intensive care unit, LOS: length of stay, MV: mechanical ventilation

## Methods

### Study Design and Oversight

The Optimizing Pharmacist Team-Integration for ICU patient Management (OPTIM) study was a multicenter, observational cohort study conducted in adult intensive care units (ICUs). The protocol was previously published and is available online.^39^ The study used both prospective and retrospective data collection strategies to evaluate the association between clinical pharmacist staffing practices and patient-centered outcomes. The study design and reporting followed the Strengthening the Reporting of Observational Studies in Epidemiology (STROBE) guidelines for cross-sectional studies (see **Supplementary Appendix)**.^40^

A waiver of informed consent was granted for ICU patient enrollment, and a partial waiver of consent was granted for pharmacist enrollment. All institutions completed a data use agreement and IRB per institutional requirements. Study procedures adhered to the ethical principles outlined in the Helsinki Declaration of 1975.^41^ REDCap (Research Electronic Data Capture) tools hosted at the University of Georgia, with technical development by the Vanderbilt University Medical Center Data Coordinating Core, were used to collect and manage study data.^42,43^

### Study Population

A total of 64 sites were included (62 in the United States, 1 in Saudia Arabia, and 1 in Jordan). All adult ICU patients were eligible for inclusion if their ICU stay was greater than or equal to 24 hours. Patients were excluded if a transition to comfort measures occurred within the first ICU day. Pharmacists at participating institutions whose primary clinical responsibility was the care of adult ICU patients during day shifts and were not currently enrolled in a training program completed data collection. Recruitment was conducted through targeted outreach, including personalized emails to postgraduate year two (PGY2) critical care pharmacy residency program directors and via professional organization listservs and communities (i.e., the American College of Clinical Pharmacy Critical Care Practice and Research Network and the Society of Critical Care Medicine Clinical Pharmacy and Pharmacology Section). Methods for recruitment and retention were also published.^44^

### Data Collection and Study Variables

Data collection occurred in three phases that resulted in data for a total of 476 raw variables. A full list of collected and calculated variables is provided in the **Supplementary Appendix.**

*Phase 1:* A survey to capture institution characteristics (e.g., region, institution type), pharmacy department staffing information (e.g., number of daytime ICU pharmacists present, ICU rounding practices), and ICU team characteristics (e.g., which professions participate in daily rounds) was completed by each participating institution. A survey assessing pharmacist characteristics (e.g., degree(s), board certification(s), years since terminal training) was completed by each participating pharmacist.

*Phase 2:* For 100 consecutive days between August 2023 and August 2024, pharmacists prospectively enrolled each ICU patient they were assigned to provide CMM who met enrollment criteria. Patients were included on the first day they received CMM but had retrospective data collection for any prior days that they were admitted to the ICU but did not receive CMM. On each day of the ICU admission the pharmacist collected both patient data and pharmacist staffing data. Patient data included, but was not limited to, the pharmacist-to-patient ratio, whether the patient received CMM care, the nurse-to-patient ratio, and the composition of the medical prescribers [i.e., attending physicians, resident(s), fellow(s), advanced practice provider(s)]. Pharmacist staffing data included the number of interprofessional rounding teams assigned to the pharmacist, number of ICU and non-ICU patients assigned, and other relevant staffing and workload indicators. The approach used to calculate the daily ICU patient-specific pharmacist-to-patient ratio is described in detail in the **Supplementary Appendix eMethods 1**. If a participating pharmacist was not working on a given day, they retrospectively recorded the workload variables of the pharmacist who was working that day (or indicated that there was no CMM provided that day) on their next day of work. Further details about data collection, including the weekend process is described in **Supplementary Appendix eMethods 2**.

*Phase 3:* Retrospective data collection from the electronic health record (EHR) for each study patient was performed through January 2025 and included baseline demographics, the severity of illness using the Sequential Organ Failure Assessment (SOFA) score^45^ and the medication regimen complexity-intensive care unit (MRC-ICU) score^28^ during the first 24 hours of the ICU admission, and the defined outcome data.

### Quality Control and Data Processing

Prior to analysis, extensive data cleaning and quality control were undertaken to ensure high quality data. Two rounds of data cleaning focused on: (1) identifying missing data and non-sensical data (e.g., date variable entered with a month of 14 or day of 60); and (2) identifying outliers (e.g., pharmacist-to-patient ratio of 1000). These data “errors” were sent back to participating pharmacists for review and correction. After these quality control measures, data were then processed to calculate summary variables at the patient and pharmacist level, including the patient-specific pharmacist-to-patient ratio, as detailed in the **Supplementary Appendix eMethods 3.**

### Outcomes, Exposures, and Covariates

The primary outcome was in-hospital mortality, defined as death during the acute hospitalization. Patients were followed through hospital discharge. Secondary outcomes included hospital length of stay (LOS), ICU LOS, and Ventilator-Free Days (VFDs). VFDs and probably of discharge are defined in **Supplementary Appendix eMethods 4**. The primary exposure was the average patient-specific, pharmacist-to-patient ratio during the ICU stay. The secondary exposure was the absence of CMM services (i.e., when no pharmacist was assigned to provide CMM to a patient for at least one day during their ICU stay. This variable was added to ensure our definition of daily CMM in the ICU was in line with multi-professionally endorsed 2025 Consensus Recommendations, which recommends that all patients admitted toa critical care service require daily care from a critical care pharmacist.^14^ Covariates included age, sex, SOFA score during first 24 hours of ICU stay, MRC-ICU score during first 24 hours of ICU stay, ICU type, hospital type, hospital Center for Medicare & Medicare Services (CMS) quality star rating, pharmacist coverage during first 24 hours of ICU stay, daily nurse-to-patient ratio, the proportion of teams the pharmacist rounded with each day among the total ICU medical teams assigned to the pharmacist, percentage of ICU days with no pharmacist on rounds, and the percentage of days where the ICU medical team consisted of only the attending physician.

### Statistical Analysis

The study was powered for the prespecified hypothesis that a lower pharmacist-to-ICU patient ratio (i.e., fewer patients per individual pharmacist) would be associated with improved survival in ICU patients. Assuming a baseline ICU mortality rate of 20% at a pharmacist-to-patient ratio of 1:15 (the recommended ratio per national guidance)^14^ and estimating a 1.5% increase in mortality for every 3 additional patients added to the ICU pharmacist CMM workload and that other covariates would explain 50% of this ratio, a sample size of 9,500 patients was deemed necessary to detect an association between the pharmacist-to-patient ratio and hospital mortality with an alpha level 0.05 and power 80%.^39^

The statistical analysis methodology is described in detail in the **Supplementary Appendix eMethods 5-9**. Missing data were handled by multiple imputation (10 datasets), and models were fit within each dataset and combined using Rubin’s rules to yield pooled coefficients and standard errors (**Supplementary Appendix eMethods 5**). For the primary analysis, multivariable generalized estimating equation (GEE) models with a logit link were fitted to estimate odds ratios (ORs) and 95% confidence intervals (CIs) for ICU mortality while accounting for within-cluster correlation (**Supplementary Appendix eMethods 6 & 7**). Sub-group sensitivity analyses are provided in the **Supplementary Appendix** included: (1) a complete-case analysis without any data imputation (**eTables 12-14 and eFigures 21-29**) (2) prespecified subgroup analyses including data from academic medical centers (vs. other hospital types) patients only (**eTables 21-26 and eFigures 40-49**), medical ICU patients only (**eTables 16-20 and eFigures 30-39**), invasively ventilated patients only (**eTables 32-34**), and an analysis including only variables from the first day of the ICU stay (**eTables 27-31**) and (3) an analysis using pharmacist-to-patient ratio (all patients) as opposed to the pharmacist-to-patient ratio (ICU patients only) (**eTable 7**). Additionally, propensity score matching was conducted, including the calculation of an e-value (**eMethods 10-12**). These additional analyses were chosen due to the wide inclusion criteria and heterogeneity of patients included in the main analysis (including institution type, severity of illness, ICU type, etc.) and the hypothesis that pharmacist-to-patient ratio may have a more significant impact on patient outcomes in certain patient populations. All models were adjusted for relevant patient- and ICU-level confounders, including institutional characteristics, patient severity of illness, ICU type, and ICU team rounding practices. All analyses were performed with the statistical software R, version 4.3.0.

## Results

### Baseline characteristics

A total of 33,464 patients were enrolled, with 4,669 excluded from analysis due to age <18 years old (n=1,685), ICU LOS <1 day (n=2,981), and missing mortality data (n=3) (**Figure 1**). A total of 28,795 patients were analyzed, representing a median (IQR) of 223.5 (129.0-311.0) patients enrolled per pharmacist (**Figure 1**). The 213 participating ICU pharmacists from 64 institutions collected data for a total of 15,888 ICU days [median days per pharmacist (interquartile range: IQR); 88.5 (45.3-108.8). The response rate for the institution demographic survey and the ICU pharmacist demographic survey was 100%. Almost all (97.3%) of the patients had a daily form completed for each day of their ICU admission. A total of 4,378 patients (15.2%) had missing data; a complete case sensitivity analysis is provided in the **Supplementary Appendix (eTables 12-14)**, using the 24,420 patients with no missing data.

The majority of pharmacists held a Doctor of Pharmacy (98.6%), were board certified in critical care (72.8%), and had completed two years of residency training (postgraduate year 1 and postgraduate year 2, 67.9%) (**Table 1**). Pharmacists reported spending a mean of 63.6% of their time on direct patient care duties, 18.7% on education duties, 14.5% on administration duties, and 7.4% on operational duties; additional pharmacist characteristics are reported in **eTable 1**.

**Table 1.** Pharmacist characteristics.

| Characteristic | N=213, n (%) |
| --- | --- |
| Degree(s) earned, n (%) |  |
| Doctor of Pharmacy | 210 (98.6) |
| Bachelor of Science in Pharmacy | 24 (11.3) |
| Master of Science | 16 (7.5) |
| Doctor of Philosophy | 1 (0.5) |
| Other | 56 (26.3) |
| Highest level of postgraduate training, n (%) |  |
| Postgraduate Year 2 (PGY2) | 144 (67.6) |
| Postgraduate Year 1 (PGY1) | 47 (22.1) |
| Fellowship | 3 (1.4) |
| No Postgraduate Training | 18 (8.5) |
| Active board certification, n (%) |  |
| Board Certification as a Critical Care Pharmacist (BCCCP) | 155 (72.8) |
| Board Certification as a Pharmacotherapy Specialist (BCPS) | 78 (36.6) |
| Board Certification in other Pharmacy Specialties | 19 (8.9) |
| No Active Board Certification | 18 (8.5) |
| Years since graduation from highest training program, mean (SD) | 8.3 (7.1) |
| Type of ICU most commonly practiced in, n (%) |  |
| Medical | 59 (27.7) |
| Surgical/Trauma | 35 (16.4) |
| Mixed Medical-Surgical | 33 (15.5) |
| Cardiothoracic/Cardiac | 27 (12.7) |
| Neurosurgery/Neurology | 14 (6.6) |
| Other | 28 (13.1) |
| Not specified | 17 (8.0) |

ICU: intensive care unit; SD: standard deviation

The average patient age was 61.2 ±17.0 years, and 12,155 (42.2%) were female (**Table 2**). Pharmacist-to-patient ratios were similar in alive patients (mean ± standard deviation (SD) 19.5 ± 10.2) compared to the deceased group (mean ± SD: 19.6 ± 10.2).76.5% of patients received CMM by a pharmacist every day of ICU admission, with fewer patients receiving CMM in the deceased group compared to the alive group (72.5% vs. 77.2%, p<0.001). **eTable 2** summarizes institutional and ICU characteristics. The Supplementary Appendix provides demographic tables split by patients who received CMM every day and those who did not as well as additional data on which patients were more likely to not receive CMM (**eFigure 1 and eTables 3 &4**)

**Table 2.** Patient demographics, intensity of pharmacist coverage and outcomes.

| Variable | Overall<br>(28,795) | Deceased<br>(4,224) | Alive<br>(24,571) | p-value |
| --- | --- | --- | --- | --- |
| **Primary Variable** |  |  |  |  |
| Pharmacist-to-patient ratio (ICU patients) | 19.5±10.2 | 19.6±10.2 | 19.5±10.2 | 0.70 |
| **Secondary Variable** |  |  |  |  |
| Absence of CMM for at least 1 day of ICU stay | 6760 (23.5) | 1160 (27.5) | 5600 (22.8) | <0.001 |
| **Co-variables** |  |  |  |  |
| Age, years | 61.2±17.0 | 65.5±15.5 | 60.5±17.1 | <0.001 |
| Sex, female | 12,155 (42.2) | 1,770 (41.9) | 10,385 (42.3) | 0.50 |
| SOFA Score* | 5.2±4.1 | 8.7±4.4 | 4.6±3.8 | <0.001 |
| MRC-ICU Score* | 11.4±6.6 | 14.6±7.1 | 10.9±6.4 | <0.001 |
| ICU admission day of the week |  |  |  | 0.01 |
| Monday | 4,634 (16.1) | 669 (15.8) | 3,965 (16.1) |  |
| Tuesday | 4,599 (16.0) | 640 (15.2) | 3,959 (16.1) |  |
| Wednesday | 4,364 (15.2) | 615 (14.6) | 3,749 (15.3) |  |
| Thursday | 4,386 (15.2) | 623 (14.8) | 3,763 (15.3) |  |
| Friday | 3,852 (13.4) | 558 (13.2) | 3,294 (13.4) |  |
| Saturday | 3,408 (11.8) | 563 (13.3) | 2,845 (11.5) |  |
| Sunday | 3,549 (12.3) | 556 (13.2) | 2,993 (12.2) |  |
| ICU type† |  |  |  | <0.001 |
| Medical | 8,850 (30.7) | 1,766 (41.8) | 7,084 (28.8) |  |
| Surgical/Trauma | 3,927 (13.6) | 426 (10.1) | 3,501 (14.3) |  |
| Surgical | 2,560 (8.9) | 284 (6.7) | 2,276 (9.3) |  |
| Cardiothoracic | 2,629 (9.1) | 199 (4.7) | 2,430 (9.9) |  |
| Cardiac | 2,498 (8.7) | 405 (9.6) | 2,093 (8.5) |  |
| Neurosurgery/Neurology | 3,886 (13.5) | 465 (11.0) | 3,421 (13.9) |  |
| Mixed Medical-Surgical | 4,195 (14.6) | 664 (15.7) | 3,531 (14.4) |  |
| Pediatric | 35 (0.1) | 2 (0.05) | 33 (0.1) |  |
| Burn | 185 (0.6) | 11 (0.3) | 174 (0.7) |  |
| Other | 27 (0.1) | 2 (0.05) | 25 (0.1) |  |
| Hospital type |  |  |  | 0.02 |
| Academic Medical Center | 19,243 (66.8) | 2,868 (67.9) | 16,375 (66.6) |  |
| Community - Teaching | 7,006 (24.3) | 1,034 (24.5) | 5,972 (24.3) |  |
| Community - Non-teaching | 2,398 (8.3) | 306 (7.2) | 2,092 (8.5) |  |
| Government/VA/Military | 148 (0.5) | 16 (0.4) | 132 (0.5) |  |
| Hospital CMS star rating |  |  |  | 0.20 |
| 2 | 6,345 (22.6) | 870 (21.7) | 5,475 (22.7) |  |
| 3 | 14,958 (53.3) | 2,151 (53.7) | 12,807 (53.2) |  |
| 4 | 6,354 (22.6) | 931 (23.3) | 5,423 (22.5) |  |
| 5 | 424 (1.5) | 50 (1.2) | 374 (1.6) |  |
| ICU pharmacist coverage 1<sup>st</sup> 24 hours‡ |  |  |  | 0.01 |
| CMM delivered on interprofessional rounds | 19,547 (67.9) | 2,885 (68.3) | 16,662 (67.8) |  |
| CMM delivered not on interprofessional rounds | 3,995 (13.9) | 527 (12.5) | 3,468 (14.1) |  |
| Abbreviated CMM delivered | 2,087 (7.2) | 296 (7.0) | 1,791 (7.3) |  |
| CMM not delivered (absence of CMM) | 3,016 (10.5) | 489 (11.6) | 2,527 (10.3) |  |
| Unknown | 142 (0.5) | 26 (0.6) | 116 (0.5) |  |
| Nurse-to-patient ratio (average), 1:X |  |  |  | <0.001 |
| ≤1 | 1,785 (6.4) | 461 (11.2) | 1,324 (5.6) |  |
| 1-2 | 2,970 (10.7) | 696 (16.9) | 2,274 (9.6) |  |
| 2 | 22,388 (80.5) | 2,878 (69.8) | 19,510 (82.3) |  |
| >2 | 674 (2.4) | 87 (2.1) | 587 (2.5) |  |
| Percent of assigned ICU teams pharmacist rounded with§ | 54 (33) | 56 (31) | 54 (33) | <0.001 |
| Percent days, no pharmacist or pharmacist trainee | 32.2 (34.1) | 31.1 (30.7) | 32.40 (34.6) | 0.60 |
| Percent days, only attending physician □ | 8.8 (26.7) | 8.7 (25.9) | 8.90 (26.8) | 0.30 |
| **Outcomes** |  |  |  |  |
| Hospital mortality | 4,224 (14.7) | 4,224 (100) | 0 | <0.001 |
| Hospital LOS, days | 12.3±17.0 | 14.1±19.6 | 12.0±16.5 | <0.001 |
| ICU LOS, days | 5.8±9.6 | 8.7±12.8 | 5.3±8.8 | <0.001 |
| Duration of mechanical ventilation, days | 2.4±18.3 | 5.1±8.0 | 1.9±19.4 | <0.001 |

Data presented as mean (±SD) or n (%)
CMM: comprehensive medication management, CMS: Center for Medicare & Medicaid Services, ICU: intensive care unit, LOS: length of stay, MRC-ICU: medication regimen complexity- intensive care unit, SOFA: sequential organ failure assessment, VA: Veterans Affairs
Full details of missingness are available in the **Supplementary Appendix**.
\*worst score during the first 24 hours of ICU stay
†Inclusion criteria specified ≥18 years of age; however, some institutions allowed patients ≥18 years of age in the pediatric ICU
‡CMM delivered on interprofessional rounds: the pharmacist attended multidisciplinary rounds and verbally provided CMM (including review of medications and recommendations); CMM delivered not on interprofessional rounds: the pharmacist provided by CMM (reviewed medications and provided recommendations) but did not attend interdisciplinary rounds; Abbreviated CMM delivered: the pharmacist provided abbreviated CMM which includes brief medication review but does not include full patient review (e.g. progress notes, results) and recommendations were provided outside of rounds; CMM not delivered (absence of CMM): the patient received no medication review from a pharmacist outside of pharmacokinetic monitoring and prospective medication order verification
§Percent of assigned ICU teams pharmacist rounded is calculated by total number of ICU medical teams the pharmacist attended and provided CMM on interprofessional rounds for divided by total number of ICU medical teams providing care for patients that the pharmacist was assigned to provide CMM for
□ Only attending physician refers to composition of the primary medical team. On these days, the patient received care from only the attending physician and did not receive care from any medical residents, fellows, or advanced practice providers.

### Hospital mortality

A total of 4,224 (14.7%) of the patients died during the hospital stay. For every one ICU patient increase that the pharmacist was assigned to provide CMM to (i.e., a one patient increase in pharmacist-to-patient ratio), the odds of mortality increased by 1.0% (OR 1.01, 95% CI 1.00-1.01, p=0.04), after adjusting for 15 different co-variates (**Table 3**). This relationship is depicted in **Figure 2**. Patients who had at least one day with no CMM had an increase in mortality of 20% (adjusted OR 1.20; 95% CI 1.03-1.40; p=0.02), after adjusting for 15 different co-variates.

**Figure 2.**
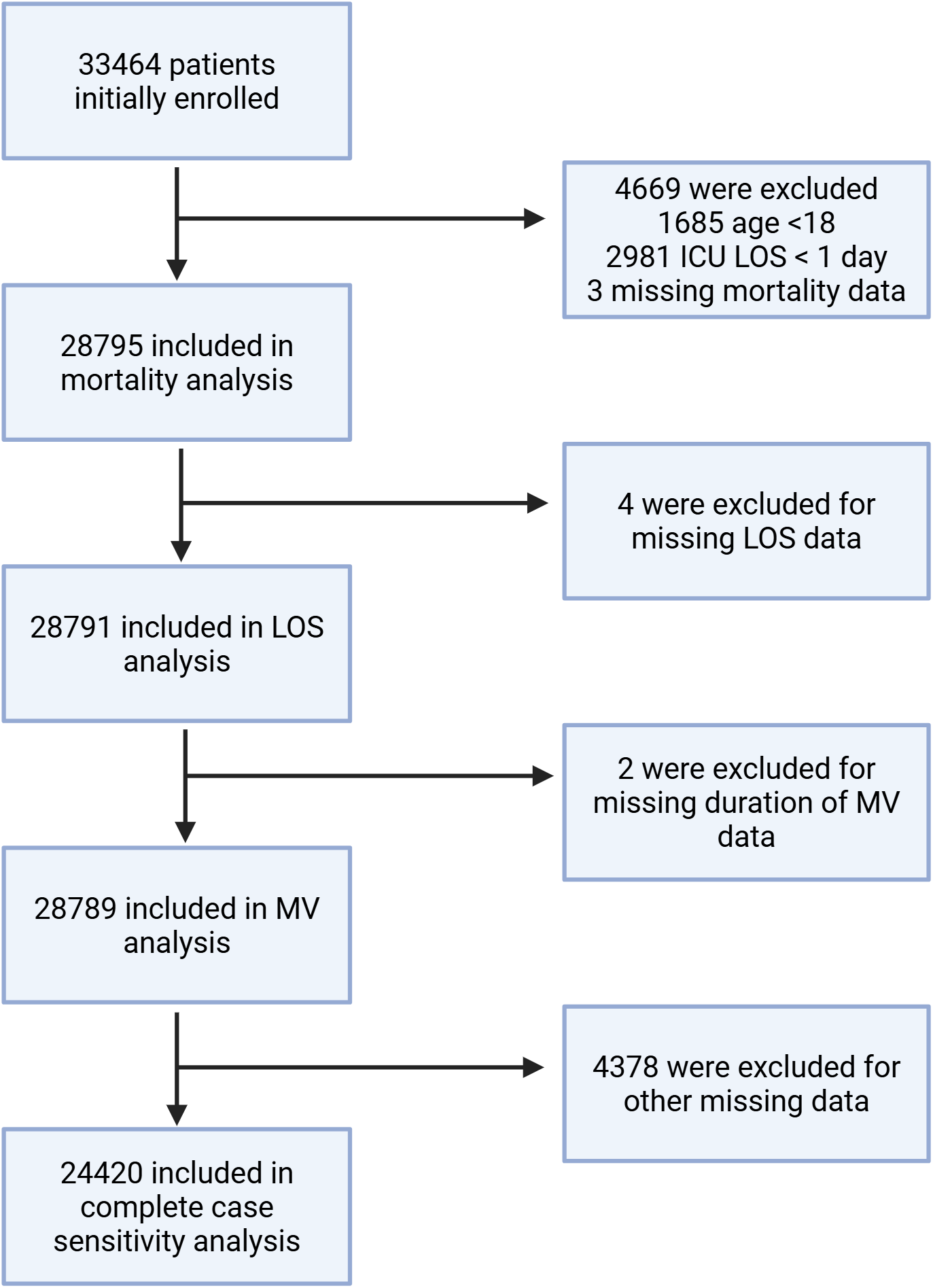
Predicted Mortality Probability by Pharmacist-to-Patient Ratio from Logistic Regression. ICU: intensive care unit 95% of the patients included in this study had an average pharmacist-to-patient ratio between 1:7 and 1:46 Pharmacist-to-patient ratio depicted as 1:X, with X being represented on the X axis

**Table 3.**
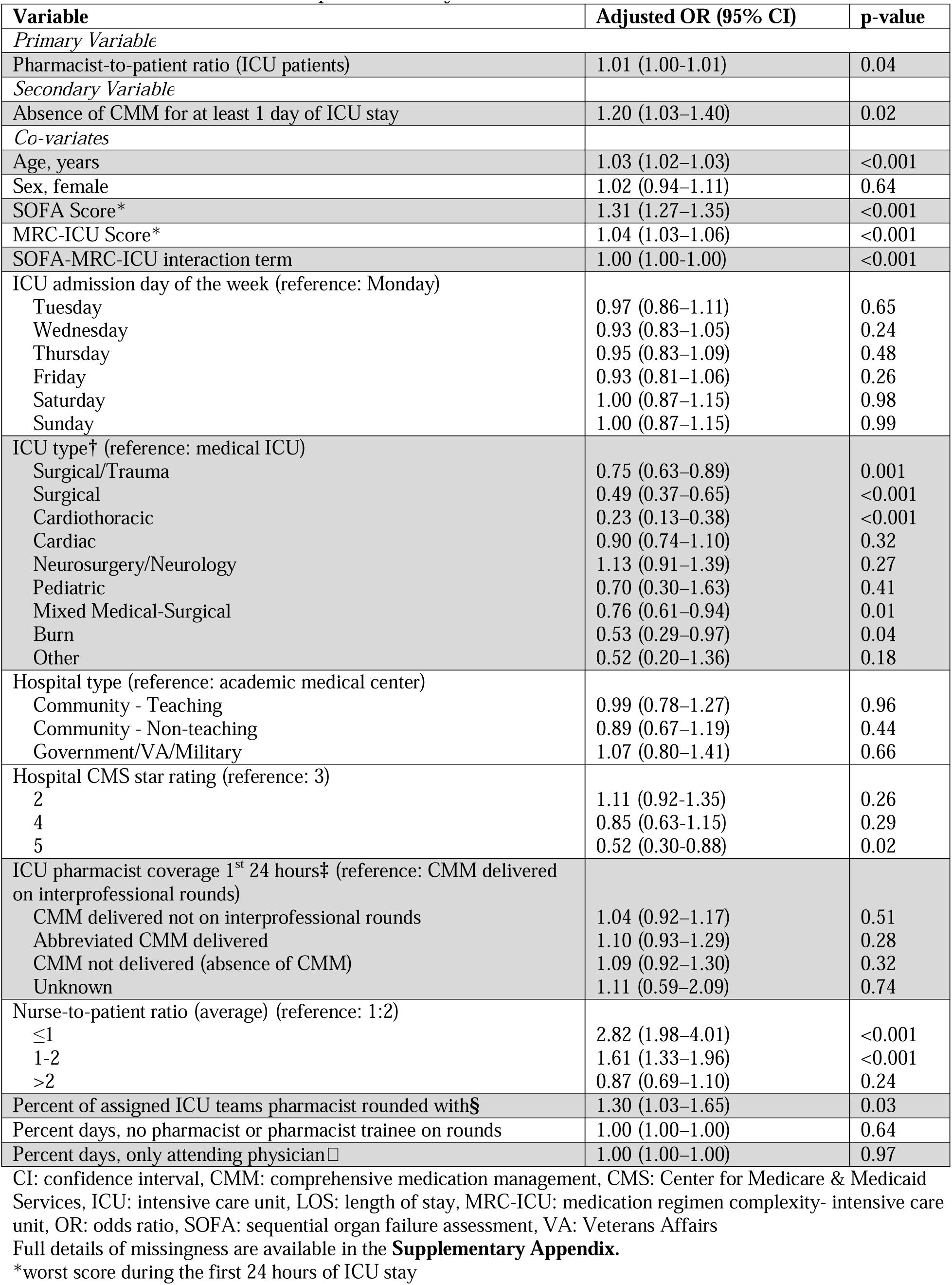
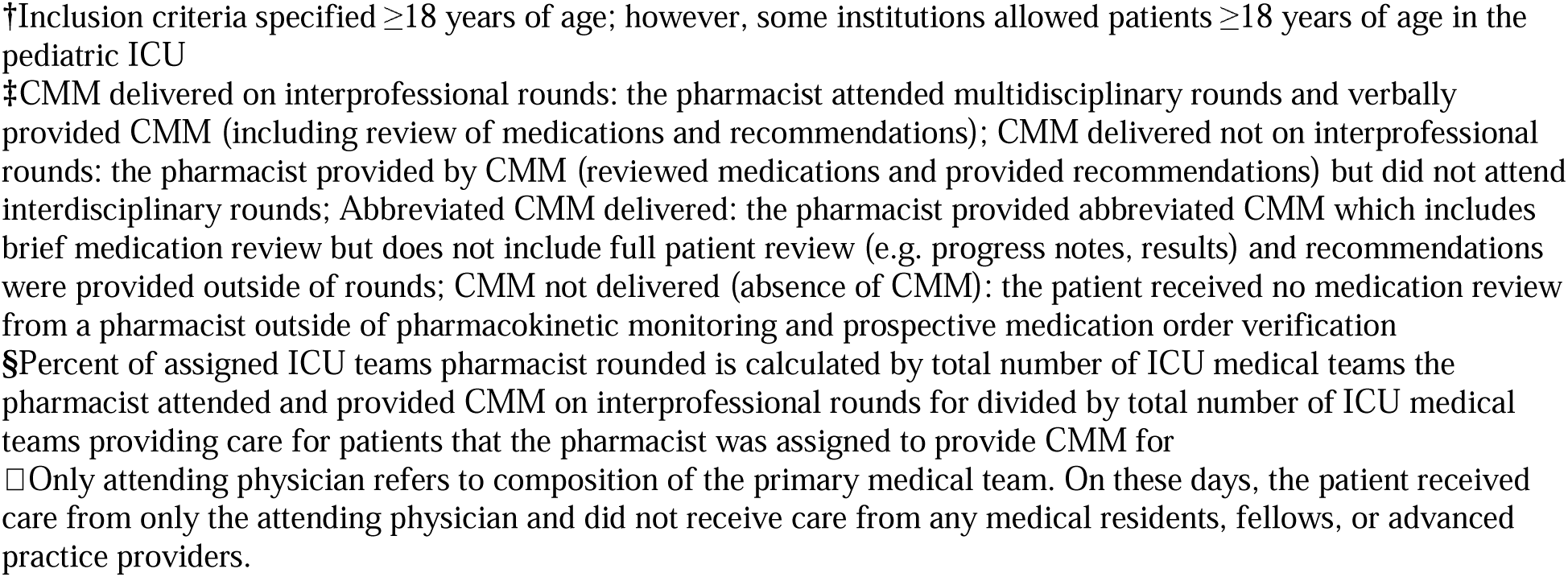
GEE model for in-hospital mortality.

| Variable | Adjusted OR (95% CI) | p-value |
| --- | --- | --- |
| *Primary Variable* |  |  |
| Pharmacist-to-patient ratio (ICU patients) | 1.01 (1.00–1.01) | 0.04 |
| *Secondary Variable* |  |  |
| Absence of CMM for at least 1 day of ICU stay | 1.20 (1.03–1.40) | 0.02 |
| *Co-variables* |  |  |
| Age, years | 1.03 (1.02–1.03) | <0.001 |
| Sex, female | 1.02 (0.94–1.11) | 0.64 |
| SOFA Score* | 1.31 (1.27–1.35) | <0.001 |
| MRC-ICU Score* | 1.04 (1.03–1.06) | <0.001 |
| SOFA-MRC-ICU interaction term | 1.00 (1.00–1.00) | <0.001 |
| ICU admission day of the week (reference: Monday) |  |  |
| Tuesday | 0.97 (0.86–1.11) | 0.65 |
| Wednesday | 0.93 (0.83–1.05) | 0.24 |
| Thursday | 0.95 (0.83–1.09) | 0.48 |
| Friday | 0.93 (0.81–1.06) | 0.26 |
| Saturday | 1.00 (0.87–1.15) | 0.98 |
| Sunday | 1.00 (0.87–1.15) | 0.99 |
| ICU type† (reference: medical ICU) |  |  |
| Surgical/Trauma | 0.75 (0.63–0.89) | 0.001 |
| Surgical | 0.49 (0.37–0.65) | <0.001 |
| Cardiothoracic | 0.23 (0.13–0.38) | <0.001 |
| Cardiac | 0.90 (0.74–1.10) | 0.32 |
| Neurosurgery/Neurology | 1.13 (0.91–1.39) | 0.27 |
| Pediatric | 0.70 (0.30–1.63) | 0.41 |
| Mixed Medical-Surgical | 0.76 (0.61–0.94) | 0.01 |
| Burn | 0.53 (0.29–0.97) | 0.04 |
| Other | 0.52 (0.20–1.36) | 0.18 |
| Hospital type (reference: academic medical center) |  |  |
| Community - Teaching | 0.99 (0.78–1.27) | 0.96 |
| Community - Non-teaching | 0.89 (0.67–1.19) | 0.44 |
| Government/VA/Military | 1.07 (0.80–1.41) | 0.66 |
| Hospital CMS star rating (reference: 3) |  |  |
| 2 | 1.11 (0.92–1.35) | 0.26 |
| 4 | 0.85 (0.63–1.15) | 0.29 |
| 5 | 0.52 (0.30–0.88) | 0.02 |
| ICU pharmacist coverage 1<sup>st</sup> 24 hours‡ (reference: CMM delivered on interprofessional rounds) |  |  |
| CMM delivered not on interprofessional rounds | 1.04 (0.92–1.17) | 0.51 |
| Abbreviated CMM delivered | 1.10 (0.93–1.29) | 0.28 |
| CMM not delivered (absence of CMM) | 1.09 (0.92–1.30) | 0.32 |
| Unknown | 1.11 (0.59–2.09) | 0.74 |
| Nurse-to-patient ratio (average) (reference: 1:2) |  |  |
| ≤1 | 2.82 (1.98–4.01) | <0.001 |
| 1-2 | 1.61 (1.33–1.96) | <0.001 |
| >2 | 0.87 (0.69–1.10) | 0.24 |
| Percent of assigned ICU teams pharmacist rounded with§ | 1.30 (1.03–1.65) | 0.03 |
| Percent days, no pharmacist or pharmacist trainee on rounds | 1.00 (1.00–1.00) | 0.64 |
| Percent days, only attending physician□ | 1.00 (1.00–1.00) | 0.97 |

CI: confidence interval, CMM: comprehensive medication management, CMS: Center for Medicare & Medicaid Services, ICU: intensive care unit, LOS: length of stay, MRC-ICU: medication regimen complexity- intensive care unit, OR: odds ratio, SOFA: sequential organ failure assessment, VA: Veterans Affairs
Full details of missingness are available in the **Supplementary Appendix**.
\*worst score during the first 24 hours of ICU stay
†Inclusion criteria specified $\geq 18$ years of age; however, some institutions allowed patients $\geq 18$ years of age in the pediatric ICU
‡CMM delivered on interprofessional rounds: the pharmacist attended multidisciplinary rounds and verbally provided CMM (including review of medications and recommendations); CMM delivered not on interprofessional rounds: the pharmacist provided by CMM (reviewed medications and provided recommendations) but did not attend interdisciplinary rounds; Abbreviated CMM delivered: the pharmacist provided abbreviated CMM which includes brief medication review but does not include full patient review (e.g. progress notes, results) and recommendations were provided outside of rounds; CMM not delivered (absence of CMM): the patient received no medication review from a pharmacist outside of pharmacokinetic monitoring and prospective medication order verification
§Percent of assigned ICU teams pharmacist rounded is calculated by total number of ICU medical teams the pharmacist attended and provided CMM on interprofessional rounds for divided by total number of ICU medical teams providing care for patients that the pharmacist was assigned to provide CMM for
□ Only attending physician refers to composition of the primary medical team. On these days, the patient received care from only the attending physician and did not receive care from any medical residents, fellows, or advanced practice providers.

Similar results were found in most sensitivity analyses (**Supplementary Appendix**), including propensity score matching. In a propensity score matching analysis without replacement, a higher pharmacist ratio of 1:16 to 1:46 compared to 1:7 to 1:15 was associated with an increased adjusted OR for mortality of 1.10 (95% CI 1.00-1.22) with an E-value of 1.43. In this analysis, not receiving CMM every day compared to receiving CMM every day was associated with an increased adjusted OR for mortality of 1.28 (95% CI 1.09-1.51) with an E-value of 1.88 (**eTable 11**). Estimated ORs for mortality during hospital admission for the ICU pharmacist-to-patient ratio, the absence of CMM for at least 1 ICU day, and each of the 15 co-variates are reported in **Table 3**. Unadjusted ORs are reported in **eTable 5.** Secondary outcomes including hospital LOS, ICU LOS, and VFDs are reported in **eTable 8** and follow similar trends. Increasing pharmacist-to-patient ratio was associated with decreased hazard of discharge alive from the hospital and ICU (adjusted hazard ratio (HR) [95% CI]: 0.99 [0.99-1.00] and 0.99 [0.98-0.99]) and decreased hazard of extubation alive (adjusted HR [95% CI]: 0.99 [0.99-1.00]). Similar results were seen with absence of CMM for at least 1 day of ICU stay, with adjusted hazard ratios of 0.71, 0.60, and 0.81 for hazard of discharge alive from the hospital, hazard of discharge alive from the ICU, and hazard of extubation alive, respectively.

## Discussion

Pharmacists providing CMM in the ICU improve patient outcomes including survival.^13,28,37,38,46,47^ In this large, multi-center evaluation of ICU adults, a relationship between increasing pharmacist-to-patient ratio and increasing odds of mortality was observed suggesting, like it has been established for ICU physicians and nurses,^20–27^ that ICU pharmacist workload is inversely related to patient outcome. Our study also highlights the importance of ICU pharmacists delivering CMM every day given the absence of a pharmacist delivering CMM for even one day is associated with an increased risk of hospital mortality by 20%. These results align with those from a meta-analysis of 14 studies that observed that a pharmacist presence on interprofessional ICU rounds reduced mortality by 22% and ADEs by 74%.^17,37,48–52^ These results are also supported by a recent causal inference analysis in a single-center ICU study that demonstrated that increased CMM care by pharmacists reduced mortality and ICU length of stay, after adjusting for relevant covariates.^13^

The associations between pharmacist staffing and mortality observed in this analysis have plausible mechanistic relationships supporting the modest effect size observed.^13,53^ E-values calculated during propensity score matching were consistent with these modest effect sizes, but could indicate that unmeasured confounding may explain the observed signal. Medications are causal agents for patient outcomes because they are a primary treatment for disease (e.g., antibiotics for sepsis) and major contributors to adverse outcomes. ICU patients experience an average of 1.7 medication errors per day and suffer ADEs ranging from 13.8 to 116.8 per 1,000 patient-days.^5,6,54,55^ These ADEs are associated with almost double the risk of mortality during a single ICU stay.^5,6,54,55^ CMM is a direct intervention intended to maximize the benefit and reduce the harm of medications and has been shown to reduce mortality in ICU patients and has been cited as an important component to team-based care.^13^ Importantly, the relationship observed in this study was in the context of top predictors and contributors to hospital mortality for patients admitted to the ICU, including severity of illness, age, and admission ICU.^56–58^ Finally, indicators for the quality of CMM have been shown to be influenced by pharmacist staffing in several multi-center, observational trials.^28,29,59^ While confounding cannot be ruled out given the study design, the present associations observed do have the strength of falling along a plausible causal pathway and importantly add to the growing body of evidence about the influence of ICU healthcare worker staffing on patient outcomes.^13,28,56,60–62^

The results of this study may play in important role in supporting guidelines regarding team-based care and may also support future interventional analyses aimed at confirming these relationships. The potential for impact of such studies could be substantial because pharmacists who provide this service are not consistently available: in a large national survey of pharmacist services, one-third of ICUs did not have direct pharmacist services (a proxy that indicates minimal delivery of CMM care), and even when pharmacists were available, 84.4% of ICUs lacked pharmacist participation in interprofessional rounds (where CMM has been found to be most impactful) on weekends.^63^ Other surveys have shown that CMM occurs under high patient volumes, and taken together, it suggests that pharmacist staffing as it relates to optimized CMM may be a modifiable factor with a large population for potential impact. Identifying practice-changing interventions for ICU patients that improve survival is notoriously difficult, and this study may support more resource-intensive analyses to confirm the observed associations.

Our study has several limitations. Given the observational design, despite rigorous regression methodology, the risk of confounding from other variables in the setting of a heterogenous patient population cannot be ruled out. Additionally, absence of CMM was typically noted at institutions without weekend pharmacist coverage; absence of CMM on a weekend may be correlated with other institutional confounders such as overall worse quality of care provided on weekends by decreased staffing or availability of timely services (e.g. procedures, imaging).

Given the voluntary nature of enrollment, selection bias for pharmacists who felt they had more time to commit to a time-intensive data collection process, potentially indicating hospitals with overall more resources and higher quality of care, is possible. Additionally, exclusion of patients who died or transferred from the ICU during the first 24 hours may have removed the highest and lowest acuity patients from this analysis, as indicated by the lower mortality noted in this study compared to the ICU literature.

Our study has several strengths. This is the largest study of ICU team staffing factors and pharmacist-to-patient ratio to date. The prospective design accompanied by use of a rigorous data collection tool ensured granular, accurate data collection with an overall data missingness rate less than 1%. This study maximized the feasibility of generating a large, externally generalizable cohort capable of supporting rigorous analysis against the known strengths of a randomized controlled trial, in which the resource-intensive nature of hiring pharmacists specifically for this intervention (and coordinators to ensure a controlled ratio) would have likely limited this study to a smaller handful of ICUs, making the results less generalizable.

## Conclusion

In this cohort study of ICU patients, both an increasing ICU pharmacist-to-patient ratio and a patient not receiving CMM every day were associated with reduced survival. Pharmacist staffing poses a modifiable, structural factor that may improve the survival of ICU patients. Our results suggesting patient outcome is more favorable when critical care pharmacists provide CMM every day need to be reproduced in future, prospective interventional trials.

## IRB Approval Information

The protocol was reviewed and approved by the Institutional Review Board at the University of Georgia (April 11, 2023).

## Conflicts of Interest

The authors have no conflicts of interest. A list of relevant disclosures are provided in the Supplementary Appendix.

## Funding

Funding through the Agency for Healthcare Research and Quality for Drs. Sikora and Smith was provided through R21HS028485 and R01HS029009. Funding through the University of Maryland, Baltimore, Institute for Clinical & Translational Research voucher program was provided to Dr. Heavner. Funding through the ASHP Research and Education Foundation was provided through a research grant to Dr. Heavner. Funding through the Board of Pharmacy Specialties was provided through a research grant to Dr. Smith. Funding through the American College of Clinical Pharmacy Critical Care Practice & Research Network was provided through research grants to Drs. Sikora, Henry, and Murray. The funders had no role in the design and conduct of the study; collection, management, analysis, and interpretation of the data; preparation, review, or approval of the manuscript; and decision to submit the manuscript for publication.

## Data Availability

All data produced in the present study are available upon reasonable request to the authors

## Acknowledgement

All authors contributed to the research and manuscript and meet ICJME authorship criteria. SES had full access to all of the data in the study and takes responsibility for the integrity of the data and the accuracy of the data analysis, including and especially any adverse effects. SES, KRH, BM, and AS contributed substantially to the study design, data analysis and interpretation, and the writing of the manuscript. ZC and XC conducted the statistical analysis and interpretation and participated in writing the first draft of the manuscript. JWD, DJM, MB, and GSM provided study design oversight and provided feedback on data interpretation and the manuscript. The OPTIM Investigator Team completed data collection and provided feedback on data interpretation and the manuscript.

## Notes

### Competing Interest Statement

Authors with conflicts of interest are listed below. If an author is not listed they reported no conflicts of interest. Marisha Burden- Dr. Burden reports funding from the Agency for Healthcare Research and Quality, the National Institute for Occupational Health and Safety, University of Colorado Innovations digiSPARK award, Med IQ, and the American Medical Association not related to this work. Dr. Burden contributed to the development of GrittyWork, a digital workforce application, and a registered trademark of the University of Colorado not related to this work. Ashley Hawthorne- Speakers Bureau for Vericel Corporation

### Clinical Protocols

https://journals.lww.com/ccejournal/fulltext/2023/09000/optimizing_pharmacist_team_integration_for_icu.3.aspx

### Author Declarations

The protocol was reviewed and approved by the Institutional Review Board at the University of Georgia (PROJECT00007120, April 11, 2023). A waiver of informed consent was granted for ICU patient enrollment, and a partial waiver of consent was granted for pharmacist enrollment.

### Summary of Updates

Added additional reference information for how to navigate the supplemental material.

