## Supplemental Appendix for "Optimization of Pharmacist Medication Management and Mortality in the Intensive Care Unit"

This appendix has been provided by the authors to give readers additional information about the work.

Sarah A.H. Adams, PharmD, Piedmont Athens Regional

Sarah K. Adie, PharmD, FACC, University of Michigan

Maria O. Agunsoye, PharmD, Abbott Northwestern Hospital

Muhammad Y. Akbik, PharmD, Baptist Memorial Hospital-Golden Triangle

May Alanazi, PharmD, Princess Noura bint Abdulrahman University College of Pharmacy

Abdullah M. Alhammad, PharmD, Department of Clinical Pharmacy, College of Pharmacy, King Saud University, Riyadh, Saudi Arabia

Enas Alkurdi, PharmD, King Hussein Cancer Center

Shannon J. Allcron, PharmD

Whitney S. Anderson, PharmD, Premier Health - Miami Valley Hospital

Eve Anderson, PharmD, Indiana University Academic Health Center

Christopher A. Anderson, PharmD, Indiana University Health

Katherine L. Artman, PharmD, University of Mississippi Medical Center

Mark Awad, PharmD, Sutter Memorial Medical Center Modesto

Wedad Bilal Awad, PharmD, King Hussein Cancer Center

Leslie D. Banuelos, PharmD, University of Maryland Medical Center, Cedars-Sinai Medical Center

Alyson T. Basting, PharmD, Indiana University Health Adult Academic Health Center

Jennifer L. Bauer, PharmD, OhioHealth-Riverside Methodist Hospital

Katherine F. Beach, PharmD, Atrium Health Wake Forest Baptist Medical Center

Erin Beauclair, PharmD, Avera

Angel M. Becker, PharmD, Abbott Northwestern Hospital

Rachel M. Belcher, PharmD, Onvida Health

Andrew T. Bennett, PharmD, University of Georgia College of Pharmacy, Wellstar MCG Health

Amanda Bernarde, PharmD, University of Alabama at Birmingham Hospital

Jessica M. Biggs, PharmD, University of Maryland Medical Center

Kara L. Birrer, PharmD, FCCP, Orlando Health Orlando Regional Medical Center

Kaitlin M. Blotske, PharmD, University of Colorado School of Medicine, University of Colorado School of Pharmacy

Christopher Bollinger, PharmD, University of North Carolina

Allison N. Boyd, PharmD, FCCM, Eskenazi Health - Department of Pharmacy

Trisha N. Branan, PharmD, FCCM, University of Georgia College of Pharmacy

Jessica M. Brochu, PharmD, Henry Ford Hospital

Michelle R.H. Brownstein, PharmD, Cleveland Clinic Weston

Quang Bui, PharmD, UCSF Benioff Children's Hospital Oakland

Simona O. Butler, PharmD, University of Michigan Health

Stacy L. Campbell-Bright, PharmD, UNC Healthcare

Christina L. Candeloro, PharmD, Roswell Park Comprehensive Cancer Center

Irene Capistrano, PharmD, Orlando Health Bayfront Hospital

Breanna L. Carter, PharmD, MBA, Erlanger, UTHSC College of Medicine-Chattanooga, Department of Surgery.

Gianna Lauren Casal, PharmD, Massachusetts General Hospital

Alyssa M. Castillo, PharmD, Texas A&M University Irma Lerma Rangel College of Pharmacy

Michael E. Chao, PharmD, UNC Health

Aaron Chase, PharmD, Medical University of South Carolina

Joshua Chestnutt, PharmD, Piedmont Columbus Regional Midtown

Genevieve Cheung, PharmD, Sharp HealthCare

Stephanie R. L. Ciapala, PharmD, Cleveland Clinic, Department of Pharmacy

Angela Clark, PharmD, University of Michigan Health

Kathryn A. Connor, PharmD, FCCP, St. John Fisher University Wegmans School of Pharmacy, The University of Rochester Medical Center

Jenna M. Corvelli, PharmD, University of Rochester Medical Center

Emma Covington, PharmD, UVA Health

Jenna F. Cox, PharmD, FCCM, Prisma Health Richland Hospital, University of South Carolina College of Pharmacy

Shawnee N. Daniel-McCalla, PharmD, University of Maryland Medical Center

Hannah Davis, PharmD, University of North Carolina Medical Center

Aubrey A. Defayette, PharmD, Roswell Park Comprehensive Cancer Center

Tilyn N. DiGiacomo, PharmD, MSHIA, University of Alabama at Birmingham Hospital

Serena A. Dine, PharmD, FCCM, Indiana University Health, Adult Academic Health Center

Sydney R. Dobson Kochheiser, PharmD, Indiana University Health

Cortney R. Dodson, PharmD, Prisma Health

Kimberly L. Doerhoff, PharmD, MPA, SSM Health DePaul Hospital

Candiss Dominick, PharmD, Upper Chesapeake Medical Center

Logan J. Doriety, PharmD, McLeod Regional Medical Center

Sabrina Dunham, PharmD, University of Michigan Health

Kyle Dvoracek, PharmD, Avera McKennan Hospital and University Health Center, Sioux Falls, SD (Current institution: Forrest General Hospital, Hattiesburg, MS)

Emily Dye, PharmD, The University of Alabama at Birmingham

Megan E. Feeney, PharmD, Boston Medical Center

Christy Forehand, PharmD, FCCM, Piedmont Augusta

Neal S. Fox, PharmD, Premier Health Miami Valley Hospital

Amber D. Fraley, PharmD, Wellstar MCG Health

Samantha L. Gauthier, PharmD, University of Kentucky Healthcare

Kayla Giang, PharmD, VA San Diego Healthcare System, UC San Diego Skaggs School of Pharmacy and Pharmaceutical Sciences, University of the Pacific Thomas J. Long School of Pharmacy.

Alexis Glenn, PharmD, University of Maryland School of Pharmacy

Caroline D. Gresham, PharmD, Piedmont Healthcare

Kristin M. Griebe, PharmD, Henry Ford Hospital, Department of Pharmacy

Razelle Grimes, PharmD, HonorHealth Deer Valley Hospital and Medical Center

Ariel H. Haber, PharmD, Cleveland Clinic Florida

Rola A. Halabi, PharmD, MedStar Washington Hospital Center

Rachel Hall, BS, University of Maryland School of Pharmacy

Kaylee Hall, PharmD, University of Mississippi Medical Center

Christian D. Hauser, PharmD, Indiana University Health Adult Academic Health Center

Ashley Hawthorne, PharmD, Auburn University Harrison College of Pharmacy

Tanner L. Hedrick, PharmD, UNC Health

Brennan Herrmann, PharmD, University of Mississippi Medical Center

McKenzie J. Hodges, PharmD, Piedmont Columbus Regional Midtown

Alana K. Holliman, PharmD, University of Georgia College of Pharmacy

Caleb Hoover, PharmD, Premier Health Miami Valley Hospital

Erin A. Houry, PharmD, SSM Health DePaul Hospital

GwangYee J. Hu, PharmD, Rutgers University

Wan-Ting Huang, PharmD, UC San Diego Health

Nicole E. Hume, PharmD, University of Kentucky

Kyle R. Humphreys, PharmD, UAB Hospital

Megan Ingebrigtson, PharmD, University of Michigan Health

Stephanie Janusz, PharmD, Premier Health-Miami Valley Hospital

Katelyn Jimison, PharmD, Atrium Health Wake Forest Baptist

Kayla E. John, PharmD, UNC Medical Center

Sara R. Jones, PharmD, Children's of Mississippi

Kevin Josey, PhD, MS, Colorado School of Public Health

Nareg Kaltakdjian, PharmD, University of Georgia College of Pharmacy

Jana L. Kelly, PharmD, McLeod Regional Medical Center

Lauren C. Kennedy, PharmD, Barnes Jewish Hospital

Rebecca P. Kessinger, PharmD, Baylor St. Luke's Texas Medical Center

Alley Killian, PharmD, Emory Healthcare

Natalie Kong, PharmD, Lankenau Medical Center

Christian L. Kressin, PharmD, University of Kentucky HealthCare

Christian E. Kroll, PharmD, University of Iowa Health Care

Abigail M. Kurtz, PharmD, Sutter Memorial Medical Center

Megan Lail, PharmD, McLeod Regional Medical Center

Kaitlin M. Landolf, PharmD, University of Maryland School of Pharmacy

Ellen Lee, PharmD, Kadlec Regional Medical Center

Jennifer Lee, PharmD, Inova Fairfax Medical Campus

Kyla Leon, PharmD, University of Mississippi Medical Center

Matthew Li, PharmD, MHA, Westchester Medical Center

Mark J. Lin, PharmD, UCSF Benioff Children's Hospital Oakland

Nicole Lu, PharmD, Community Regional Medical Center, Fresno CA

Whitney J. Ly, PharmD, University of North Carolina Medical Center

Katharine L. Madding, PharmD, Premier Health Miami Valley Hospital

Olivia Marchionda, PharmD, Cleveland Clinic

Angelica Marques, PharmD, McLeod Regional Medical Center

Whitney Mays, PharmD, Children's of Mississippi

Bradford L. McDaniel, PharmD, MBA, Carilion Clinic

Allyson M. McIntire, PharmD, Franciscan Health Indianapolis

Brian McKinzie, PharmD, University of North Carolina Medical Center

Ana McLean, PharmD, University of Georgia College of Pharmacy

Mary McNeely, PharmD, Abbott Northwestern Hospital - Allina Health

Alyssa S. Meester, PharmD, The Ohio State University Wexner Medical Center

Arjay Mendoza, PharmD, VA San Diego Healthcare System

Kailey R. Meyer, PharmD, Avera McKennan Hospital & University Health Center

Ashley E. Milkovits, PharmD, Carilion Clinic

Stephanie Millan, PharmD, University of Georgia

James T. Miller, PharmD, University of Michigan Health

Christopher Miller, PharmD, MedStar Washington Hospital Center

Keri L. Mills, PharmD, Baptist Memorial Hospital Golden Triangle, DCH Health System Regional Campus

Corinne Murphy, PharmD, Piedmont Columbus Regional

Neha D. Naik, PharmD, MBA, Emory Healthcare

Lama H. Nazer, PharmD, FCCM, King Hussein Cancer Center

Andrea M. Nei, PharmD, FCCM, Mayo Clinic

Jared W. Netley, PharmD, University of Wisconsin Hospitals and Clinics

Jennifer D. Nguyen, PharmD, UCSF Benioff Children's Hospital Oakland

Kelly T. Nguyen, PharmD, University of Georgia College of Pharmacy

Christian Nicolosi, PharmD, University of Maryland

Nicole M. Palm, PharmD, FCCM, Cleveland Clinic

Komal Pandya, PharmD, MBA, FCCM, University of Kentucky

Kristine A. Parbuoni, PharmD, University of Maryland School of Pharmacy

Sara E. Parli, PharmD, UK HealthCare, Department of Pharmacy Services, University of Kentucky College of Pharmacy, Department of Pharmacy Practice and Science

Shyam Patel, PharmD, Boston Medical Center, Department of Pharmacy, Boston University Chobanian & Avedisian School of Medicine, Department of Emergency Medicine

Akta S. Patel, PharmD, McLeod Regional Medical Center

Kerilyn Petrucci, PharmD, UChicago Medicine

Brian V. Phan, PharmD, United Therapeutics Corporation

Kara E. Phillips, PharmD, University of Georgia College of Pharmacy, Northside Hospital

Stephanie T. Proctor, PharmD, Kadlec Regional Medical Center

Caitlin J. Quibeuf, PharmD, MUSC Health

Stephen H. Rappaport, PharmD, FCCM, University of Rochester Medical Center

Marianne E. Ray, PharmD, University of Mississippi College of Pharmacy

Paige E. Reese, PharmD, Children's Hospital of Georgia

Stephanee L. Rhoades, PharmD, ProMedica Toledo Hospital

Tim Robinson, PharmD, Wellstar MCG

Christine B. Rojas, BS, University of Maryland School of Pharmacy

Klayton M. Ryman, PharmD, UT Southwestern Medical Center

Alicia J. Sacco, PharmD, FCCM, Mayo Clinic Arizona

Mallorie N. Saling, PharmD, Jackson Health System

Robert A. Sbertoli, PharmD, SSM Health Saint Louis University Hospital

Madeline A. Scarbrough, PharmD, UT Southwestern Medical Center

Addy E. Schoening, PharmD, Abbott Northwestern Hospital - Allina Health

Janet Shin, PharmD, UCSF Benioff Children's Hospital Oakland

Brandon M. Smith, PharmD, Prisma Health Richland

Maya Smith, PharmD, University of Maryland School of Pharmacy

Zachary R. Smith, PharmD, FCCP, FCCM, Henry Ford Hospital, Department of Pharmacy

Brooke A. Smith, PharmD, Wellstar MCG Health

Alyssa Sonchaiwanich, PharmD, Mayo Clinic Rochester

Jillian K. Songstad, PharmD, Sanford USD Medical Center

Katherine M. Spezzano, PharmD, MBA, University of Kentucky

Gillian Steiger, PharmD, Massachusetts General Hospital

Donna Steinbacher, PharmD, UNC Health

Melissa Sterling, PharmD, Boston Medical Center

Joseph Stoldt, PharmD, IU Health

Margaret Street, PharmD, Baylor St. Luke's Medical Center

Maria K. Stubbs, BS, VA San Diego Healthcare System, UC San Diego Skaggs School of Pharmacy and Pharmaceutical Sciences, University of the Pacific Thomas J. Long School of Pharmacy

Jennifer Tawwater, PharmD, UT Southwestern Medical Center

Ashley N. Taylor, PharmD, Wellstar MCG Health

Dakota Taylor, PharmD, University of Mississippi Medical Center

Tori Thompson, PharmD, MercyOne Des Moines Medical Center

Kailee Toews, PharmD, University of California, San Francisco

Tu-Trinh Tran, PharmD

Abby Tyson, PharmD, OhioHealth Riverside Methodist Hospital

Sandra A. Valencia, PharmD, Providence Sacred Heart Medical Center

Meghna Vallabh, PharmD, Baylor St. Luke's Medical Center

Storm A. Van Wey, PharmD, Indiana University School of Medicine, Butler University College of Pharmacy and Health Sciences, Purdue University College of Pharmacy

Beth E. Varnes, PharmD, UAB Hospital

Vanessa Velazco, PharmD, Cleveland Clinic Florida

Arianna J. Vidger, PharmD, Indiana University Health

Ann W. Vu, PharmD, Community Regional Medical Center

Stephanie C. Waldrep, PharmD, University of Alabama at Birmingham Hospital

Jessica A. Ward, PharmD, Cleveland Clinic

Nathaniel B. Wayne, PharmD, Wellstar MCG Health, University of Georgia College of Pharmacy

Lori Wetmore, PharmD, IU Health

Jessica A. Whitten, PharmD, Eskenazi Health

Alexandra M. Wiegand, PharmD, UK HealthCare Department of Pharmacy Services

Sarah K. Williford, PharmD, University of North Carolina Medical Center

Sharon L. Wilson, PharmD, University of Maryland Medical Center

Kevin Wohlfarth, PharmD, ProMedica Toledo Hospital

Douglas R. Wylie, PharmD, Chiesi USA

Siu Yan A. Yeung, PharmD, University of Maryland Medical Center

Jae H. Yook, PharmD, Piedmont Athens Regional

Connie H. Yoon, PharmD, OhioHealth Riverside Methodist Hospital

### List of acknowledgements

REDCap Consortium at Vanderbilt

Pharmaceutical Research Computing (PRC) center at University of Maryland School of Pharmacy

Society of Critical Care Medicine (SCCM)

American College of Clinical Pharmacy (ACCP)

Diana Aguilar

Ghadah Alajmi

Julia Alexander

Emily Austin

Cesar Bejarano-Garcia

Mary Blair

James Braun

Nicholas Bravo

Garrett Brown

Jennifer Bui

Joshua Campbell

Carlette Cavenaugh

Sarah Chiu

Patrick Costello

Samantha Delibert

Sam Dewitt

Megan Dorsey

Courtney Feagin

Shelby Fideler

Emily George

Kristen Giles

Renae Gozelski

Aileen Gregorio-Corallo

Liana Ha

Andrea Hankins

Gresham Hindman

Elizabeth Hodges

Cory Johnson

Lama Kanawati

Nadine Kanyana

Tara Kennell

Alexa Luboff

Isabel Mangaoang

Carolyn Martz

Taylor McCart

Jamie McCarthy

Hannah McMurrin

Emily Miller

Makenna Moll

Peter Moran

Rebecca Morgan

Amoreena Most

Zach Muller

Nicole Newton

Justin Petrovic

Inna Perinskaya

Nicole Pfeffer

Leslie Phillips

Lisa Pickmans

Laura Provost

Marinna Raqueno

Jenna Schwartz

Shaleen Singh

Kelcy Sorsensen

Inderpal Srai

Samori Swygert

Kari Taggart

Farrah Tavakoli

Melissa Thompson Bastin

Noelle Vo

Todd Walroth

Hailey Wang

Brian Watson

Ashley Wischmeyer

Laura Witt

Shelby Young

Denisse Garcia Zavala

Kara Zacholski

Qingrong Laura Zhang

### Reporting of Observational Studies in Epidemiology (STROBE) Checklist

|  | Item No | Recommendation | Page No |
| --- | --- | --- | --- |
| **Title and abstract** | 1 | (*a*) Indicate the study’s design with a commonly used term in the title or the abstract | 1-2 |
|  |  | (*b*) Provide in the abstract an informative and balanced summary of what was done and what was found |  |
| Introduction | | | |
| Background/rationale | 2 | Explain the scientific background and rationale for the investigation being reported | 3 |
| Objectives | 3 | State specific objectives, including any prespecified hypotheses | 3 |
| Methods | | | |
| Study design | 4 | Present key elements of study design early in the paper | 4, Supplemental |
| Setting | 5 | Describe the setting, locations, and relevant dates, including periods of recruitment, exposure, follow-up, and data collection | 4 |
| Participants | 6 | (*a*) Give the eligibility criteria, and the sources and methods of selection of participants. Describe methods of follow-up | 4 |
|  |  | (*b*) For matched studies, give matching criteria and number of exposed and unexposed | Supplemental propensity score matching analysis |
| Variables | 7 | Clearly define all outcomes, exposures, predictors, potential confounders, and effect modifiers. Give diagnostic criteria, if applicable | 4-5 |
| Data sources/ measurement | 8* | For each variable of interest, give sources of data and details of methods of assessment (measurement). Describe comparability of assessment methods if there is more than one group | 4-5 |
| Bias | 9 | Describe any efforts to address potential sources of bias | 5-6 |
| Study size | 10 | Explain how the study size was arrived at | 5 |
| Quantitative variables | 11 | Explain how quantitative variables were handled in the analyses. If applicable, describe which groupings were chosen and why | 5 |
| Statistical methods | 12 | (*a*) Describe all statistical methods, including those used to control for confounding | 5-6 |
|  |  | (*b*) Describe any methods used to examine subgroups and interactions | Supplemental |
|  |  | (*c*) Explain how missing data were addressed | Supplemental |
|  |  | (*d*) If applicable, explain how loss to follow-up was addressed | Supplemental |
|  |  | (*e*) Describe any sensitivity analyses | Supplemental |
| Results | | |  |
| Participants | 13* | (a) Report numbers of individuals at each stage of study—eg numbers potentially eligible, examined for eligibility, confirmed eligible, included in the study, completing follow-up, and analysed | 6, Figure 1 |
|  |  | (b) Give reasons for non-participation at each stage | Figure 1 |
|  |  | (c) Consider use of a flow diagram | Figure 1 |
| Descriptive data | 14* | (a) Give characteristics of study participants (eg demographic, clinical, social) and information on exposures and potential confounders | 6 |
|  |  | (b) Indicate number of participants with missing data for each variable of interest | Supplemental |
|  |  | (c) Summarise follow-up time (eg, average and total amount) | 6 |
| Outcome data | 15* | Report numbers of outcome events or summary measures over time | 6, Tables 2 and 3 |

| Main results | 16 | (*a*) Give unadjusted estimates and, if applicable, confounder-adjusted estimates and their precision (eg, 95% confidence interval). Make clear which confounders were adjusted for and why they were included | 7, Table 3 |
| --- | --- | --- | --- |
|  |  | (*b*) Report category boundaries when continuous variables were categorized | 7 |
|  |  | (*c*) If relevant, consider translating estimates of relative risk into absolute risk for a meaningful time period | 7 |
| Other analyses | 17 | Report other analyses done—eg analyses of subgroups and interactions, and sensitivity analyses | Supplemental |
| Discussion | | | |
| Key results | 18 | Summarise key results with reference to study objectives | 8 |
| Limitations | 19 | Discuss limitations of the study, taking into account sources of potential bias or imprecision. Discuss both direction and magnitude of any potential bias | 8-9 |
| Interpretation | 20 | Give a cautious overall interpretation of results considering objectives, limitations, multiplicity of analyses, results from similar studies, and other relevant evidence | 8-9 |
| Generalisability | 21 | Discuss the generalisability (external validity) of the study results | 8-9 |
| Other information | | | |
| Funding | 22 | Give the source of funding and the role of the funders for the present study and, if applicable, for the original study on which the present article is based | 1 |

*Give information separately for exposed and unexposed groups.

**Note:** An Explanation and Elaboration article discusses each checklist item and gives methodological background and published examples of transparent reporting. The STROBE checklist is best used in conjunction with this article (freely available on the Web sites of PLoS Medicine at http://www.plosmedicine.org/, Annals of Internal Medicine at http://www.annals.org/, and Epidemiology at http://www.epidem.com/). Information on the STROBE Initiative is available at http://www.strobe-statement.org.

### DISCLOSURES

Authors with conflicts of interest are listed below. If an author is not listed, they reported no conflicts of interest.

Marisha Burden- Dr. Burden reports funding from the Agency for Healthcare Research and Quality, the National Institute for Occupational Health and Safety, University of Colorado Innovations digiSPARK award, Med-IQ, and the American Medical Association not related to this work.  Dr. Burden contributed to the development of GrittyWork, a digital workforce application, and a registered trademark of the University of Colorado not related to this work.

Ashley Hawthorne- Speaker’s Bureau for Vericel Corporation

### Additional Methods

#### eMethods 1. Definition of Pharmacist-to-Patient Ratio

The daily pharmacist-to-patient ratio was calculated for each ICU patient and averaged over the course of their ICU stay. For example, if a patient had a 3-day ICU length of stay and the pharmacist caring for the patient also covered 12, 8, and 14 ICU patients across these three days, then the patient-specific pharmacist-to-patient ratio would be: (12+8+14)/3 = 11.3 (ratio: 1:11). The daily pharmacist-patient ratio (all patients) was also calculated each day and included all patients (ICU as well as floor or step-down patients) that the pharmacist was assigned to provide CMM for. Since most pharmacists had the majority of their workload from providing CMM to ICU patients, the main analysis includes pharmacist-to-patient ratio (ICU patients) but an analysis of pharmacist-to-patient ratio (all patients) is provided in **Supplementary Appendix eTable 7**. If the patient did not receive CMM, that day did not count towards their calculation of the pharmacist-to-patient ratio, as any attempt to quantify the effective ratio for no CMM would have been pure speculation. Thus, not receiving CMM was treated as a separate variable for analysis to distinguish these patients, as the care they received is likely not truly reflected by the ratio alone. For example, if a patient had a 4-day ICU length of stay and the pharmacist caring for the patient also covered 12, 8, and 14 ICU patients across the first 3 days and the patient received no CMM on the fourth day, then the patient-specific pharmacist-to-patient ratio would be: (12+8+14)/3 = 11.3 (ratio: 1:11).

#### eMethods 2. Data Collection – Weekends and Non-OPTIM-participating Pharmacists

3.2% of the institutions had no pharmacist clinical coverage (CMM) on weekends; however, the majority of missing CMM occurred on weekends (see eTable 3). For these institutions, patients who were admitted during a weekend had their data retrospectively recorded on Monday, indicating that they received no CMM during the weekend. Patients who had no CMM were identified typically through either daily printing or saving of the hospital census or through the data collection tool in REDCap where patients were marked as “incomplete” until it was documented that they had been discharged from the ICU. While the pharmacists who participated typically included all the pharmacists who covered a particular unit so there was no lack of continuity, in cases where a non-OPTIM pharmacist was covering, an OPTIM-pharmacist would enter the workload of the non-OPTIM pharmacist for each patient through using the unit census list for that day. If they were able to answer additional questions (like which specific patients had rounding, if the patients were new patients for the non-OPTIM pharmacist, etc.) they would enter those as well, and if unable to answer, they would mark those data points as unknown.

#### eMethods 3. Data Processing and Quality Control

A series of novel variables were calculated using preexisting data from the OPTIM REDCap. Novel variables were calculated that required critical care pharmacist (CCP) and patient data to be matched and that also account for different CCPs caring for the same patient on different days of their ICU admission. Variables to be calculated from the preexisting data include things like key workload metrics calculated at both the patient and CCP level (e.g., patient-specific CCP-to-patient ratio vs. pharmacist-specific CCP-to-patient ratio).

Importantly, calculation of these variables required form matching using rigorous, data science methods and trained data analysts. In the preexisting REDCap tool, each patient had an outcomes form and had multiple daily forms corresponding to each day that they were admitted to the ICU. The daily patient forms provided information specific to the patient’s ICU day (e.g., what was the ICU team makeup for this specific patient on this day, was this specific patient rounded on this day, which CCP covered the patient this day). Similarly, each CCP had a demographics form and multiple CCP daily forms for each day that they worked during the 100-day period of prospective data collection. Each daily patient form must be matched with the daily pharmacist form for the corresponding CCP who covered the patient on that specific day. This matching provided information on pharmacist workload. Since a patient may be covered by several different CCPs over the course of their ICU stay, they also may be matched with several CCP demographic forms as well as an institution demographic form. In total, each patient will have the following forms, where *X* = number of distinct CCPs who covered the patient during their ICU stay and *Y* = number of ICU days:

$$No. of REDCap forms for each patient=institution demographic form+ICU team form+\left( X*CCP demographic forms \right)+\left( Y*CCP daily forms \right)+(Y*patient daily forms)+outcomes form$$

For example, a patient who had 5 ICU days and was covered by 3 different pharmacists on those days would have a total of 16 forms

$$No. of REDCap forms for patient=1+1+\left( 3*1 \right)+\left( 5*1 \right)+\left( 5*1 \right)+1=16$$

#### eMethods 4. Definition of Ventilator Free Days and Probability of Discharge

We defined 28-day ventilator-free days (VFD) as the number of days from day 0 (first ventilation) to day 28 during which a patient was alive and free of mechanical ventilation. Patients who died before day 28 or remained ventilated through day 28 were assigned VFD = 0. For patients extubated and later re-intubated within 28 days, only days not receiving ventilation counted toward VFD.

Modeling length of stay (LOS) as time to discharge alive with a competing risk of death directly targets the clinical and operational question: “How quickly are patients discharged, accounting for the fact that some die before discharge?” Standard survival or linear LOS models can be misleading because death truncates LOS and creates informative censoring: patients who die are not “long-stayers,” they are non-discharged. The Fine–Gray hazards approach estimates the cumulative incidence of discharge, which (i) treats death appropriately as a competing event rather than censoring it, (ii) yields subdistribution hazard ratios (SHR) that map to an absolute, time-indexed probability of discharge, and (iii) aligns with bed-flow and staffing decisions grounded in probabilities of discharge over time. Practically, SHR >1 indicates a higher rate of discharge (shorter LOS) after adjusting for covariates, while SHR <1 indicates prolonged stays. This framework avoids survivor bias inherent in analyzing raw LOS among only survivors, preserves an interpretable link to absolute risk, and complements hazards models by answering the policy-relevant question about discharge likelihood in the presence of death.

#### eMethods 7. Handling of Missing Data

Missing covariates were addressed under a missing-at-random (MAR) assumption using multiple imputation by chained equations (m = 10). The imputation included all analysis covariates, and the outcomes themselves were not imputed. Continuous variables were imputed with predictive mean matching (k = 5) with logical bounds; binary variables with logistic regression; and multi-category variables with polytomous regression. Derived variables were passively imputed to maintain algebraic consistency. Each analysis model (GEE for mortality, Fine-Gray for LOS) was fit within each imputed dataset, and point estimates and variances were pooled with Rubin’s rules. Convergence diagnostics, distributions of imputed vs observed values, and the fraction of missing information were reviewed and were consistent with the MAR assumption. As sensitivity analyses, we repeated all models in the complete-case cohort and explored pattern-mixture (delta) adjustments to assess robustness to plausible departures from MAR.

#### eMethods 6. Modeling for the Primary Analysis

We modeled In-Hospital mortality using multivariable generalized estimating equations (GEE) with a logit link to account for clustering of patients within ICUs. The primary variable was the patient-level average pharmacist-to-patient ratio, parameterized per one additional ICU patient per pharmacist; for interpretability we also analyzed a prespecified threshold contrast (fewer patients, ≤1:15, vs more patients, 1:15.1 to 1:46) to derive absolute effects. Results are presented as adjusted odds ratios (OR) with 95% confidence intervals (CI) and two-sided p-values.

ICU and hospital LOS were analyzed as time to discharge alive using Fine-Gray subdistribution hazards regression, which estimates covariate effects on the cumulative incidence of discharge in the presence of competing death. For ICU LOS, time originated at ICU admission and the event was ICU discharge alive with ICU death as the competing event; for hospital LOS, time originated at hospital admission, and the event was hospital discharge alive with In-Hospital death as the competing event. The effect was summarized as the subdistribution hazard ratio per one additional ICU patient per pharmacist, with 95% CI and two-sided P values; by construction, values greater than 1 indicate a higher rate of discharge (shorter stays) and values less than 1 indicate a lower rate of discharge (longer stays). As complementary analyses, we repeated all models in the complete-case cohort and in prespecified subgroups.

#### eMethods 7. Modeling for Analysis Adjusting for Baseline Co-variates

We adjusted for prespecified patient- and ICU-level baseline covariates selected *a priori* based on clinical relevance and prior literature. Patient-level covariates included age, sex, and severity of illness (SOFA and MRC-ICU scores). ICU-level covariates included ICU type† (e.g., medical, surgical, mixed), institutional type (e.g., academic, community, government) as well as Center for Medicare & Medicaid Services (CMS) star quality ratings, and team rounding practices (e.g., percentage of teams rounding with a pharmacist). All covariates were coded to reflect baseline status at or within the first 24 hours of ICU admission. Continuous variables were retained on their native scales, and categorical variables used clinically meaningful levels. To limit overfitting and preserve interpretability, we did not use data-driven variable selection; instead, the same covariate set was applied across mortality and length-of-stay models and used for marginal standardization when estimating adjusted risks and absolute risk differences.

#### eMethods 8. Collinearity Analysis

To assess multicollinearity among predictors included in the GEE mortality model, a correlation matrix and variance inflation factors (VIF) of predictors included in the mortality model were created. These are reported in the supplementary appendix.

#### eMethods 9. Propensity Score Matching

Propensity score matching was employed to reduce confounding in observational study by balancing baseline characteristics between groups. The propensity score was defined as the conditional probability of receiving the exposure given observed baseline covariates and was estimated using a multivariable logistic regression model. All covariates described above were included in the propensity score model as potential confounders, encompassing demographic characteristics, clinical variables, and relevant baseline measures.

Individuals in the treatment and control groups were then matched based on similar propensity scores using a nearest-neighbor matching algorithm with a predefined caliper, without replacement. This approach creates a pseudo-randomized sample in which the distribution of measured covariates is more comparable across groups. Balance between groups after matching was assessed by standardized mean differences, with values less than 0.1 indicating adequate covariate balance. The matched cohort was subsequently used for mortality outcome analyses.

#### eMethods 10. E-Value Methods

We used E-values to assess the robustness of the observed associations to potential unmeasured confounding. The E-value represents the minimum strength of association, on the risk ratio scale, that an unmeasured confounder would need to have with both the exposure and the outcome, conditional on the measured covariates, to fully explain away the observed effect estimate. For a risk ratio (RR) greater than 1, the E-value was calculated as $RR +\sqrt{RR \times(RR -1)}$. For protective associations (RR < 1), the E-value was computed using the reciprocal of the risk ratio. Larger E-values indicate that stronger unmeasured confounding would be required to explain away the observed association, whereas smaller E-values suggest greater sensitivity to unmeasured confounding. This approach provides a quantitative assessment of sensitivity to unmeasured confounding in observational studies.

#### eMethods 11. E-Value Results

In general, matching with replacement allows each treated unit to be paired with its closest available control, which can improve local match quality and reduce potential bias, particularly in regions of the propensity score distribution where suitable controls are sparse. This advantage, however, comes at the cost of increased variance, as a smaller subset of control units may be reused multiple times, thereby reducing the effective sample size. Matching without replacement, by contrast, distributes control units more evenly and can yield smaller variance and narrower confidence intervals, although it may introduce bias if control units are locally exhausted and treated units are forced to match to more distant controls. In settings with a large sample size, high-quality matches can still be achieved even without replacement, while also preserving an intuitive study design, maintaining a larger effective sample size, and achieving lower variance. Viewed through this lens, our empirical results align well with these expected trade-offs. Under matching without replacement, both exposures were significantly associated with mortality, with adjusted odds ratios of 1.10 (95% CI: 1.00-1.22) for the pharmacist-to-patient ratio and 1.28 (95% CI: 1.09-1.51) for CMM every day of ICU stay, accompanied by relatively narrow confidence intervals and E-values of 1.43 and 1.88, respectively. Overall, the E-values are consistent with the modest effect sizes observed in this study and are comparable to prior ICU-based observational studies. Additionally, our study provides extensive covariate adjustment and good balance achieved through PSM.

### Supplemental Tables

#### eTable 1. Additional Pharmacist Demographics

|  | N=213 |
| --- | --- |
| FTE Allocation (as perceived by the pharmacist), mean (SD)  Direct Patient Care  Operational Duties  Teaching/Education  Administration | 63.6 (18.9)  7.4 (12.6)  18.7 (13.7)  14.5 (16.4) |
| Order Verification, Rounding Service, n (%)  0%  1-25%  26-75%  75-99%  100%  Not applicable- do not have this type of patient | 7 (3.3) 51 (24.1)  47 (22.2)  72 (34.0)  34 (16.0)  1 (0.5) |
| Order Verification, Nonrounding Service, n (%)  0%  1-25%  26-75%  75-99%  100%  Not applicable- do not have this type of patient | 48 (22.6)  79 (37.3)  36 (17.0)  22 (10.4)  15 (7.1)  12 (5.7) |
| Order Verification, non-ICU patients, n (%)  0%  1-25%  26-75%  75-99%  100%  Not applicable- do not have this type of patient | 89 (42.0)  61 (28.8)  7 (3.3)  9 (4.3)  9 (4.3)  37 (17.5) |

FTE: full-time equivalent, ICU: intensive care unit; SD: standard deviation

#### eTable 2. Institutional Demographics

|  | N=64, n (%) |
| --- | --- |
| Hospital Type  Academic Medical Center  Community - Teaching  Community - Non-teaching  Government/VA/Military | 39 (60.9)  16 (25.0)  8 (12.5)  1 (1.6) |
| Hospital Beds  <200  200-400  401-600  >600 | 2 (3.1)  17 (16.6)  14 (21.9)  31 (48.4) |
| Hospital CMS Star Rating  2  3  4  5  Missing/N/A* | 12 (20.3)  34 (57.6)  11 (18.6)  2 (3.4)  5 (7.8) |
| ICU Beds  <26  26-50  51-75  76-100  101-150  >150 | 6 (9.4)  13 (20.3)  9 (14.1)  6 (9.4)  10 (15.6)  20 (31.3) |
| Region  Northeast  Southeast  Midwest  Northwest  Southwest  International | 11 (17.2)  20 (31.3)  16 (25.0)  4 (6.3)  11 (17.2)  2 (3.1) |
| Adult Trauma Center Type  Level I  Level II  Level III  Level IV  Level V  N/A | 33 (51.6)  14 (21.9)  6 (9.4)  1 (1.6)  0  10 (15.6) |
| Highest Level of Stroke Designation  Comprehensive Stroke Center  Thrombectomy-Capable Stroke Center  Primary Stroke Center  Acute Stroke-Ready Hospital  Basic Stroke Certification  No Stroke Certifications or Designations | 40 (62.5)  4 (6.25)  16 (25)  1 (1.56)  0  3 (4.69) |
| Electronic Medical Record  Epic  Cerner  Meditech  Sunrise Clinical Manager  VA  Other | 50 (78.1)  10 (15.6)  1 (1.6)  1 (1.6)  1 (1.6)  1 (1.6) |
| Typical Weekend Pharmacist Coverage  CMM delivered on interprofessional rounds for all ICU patients  CMM with bedside rounds for some ICU patients  CMM delivered not on interprofessional rounds  Abbreviated CMM No CMM  Missing | 7 (11.3)  11 (17.7)  6 (9.7)  36 (58.1)  2 (3.2)  2 (3.1) |

ICU: intensive care unit, N/A: not applicable, CMM: comprehensive medication management, CMS: Center for Medicare & Medicaid Services, VA: Veterans Affairs

*2 centers do not have a CMS star rating as they are located outside of the United States (US). 2 US hospitals included also did not have a CMS star rating per the publicly available CMS website as of April 2026.

#### eFigure 1. Distribution of lack of CMM

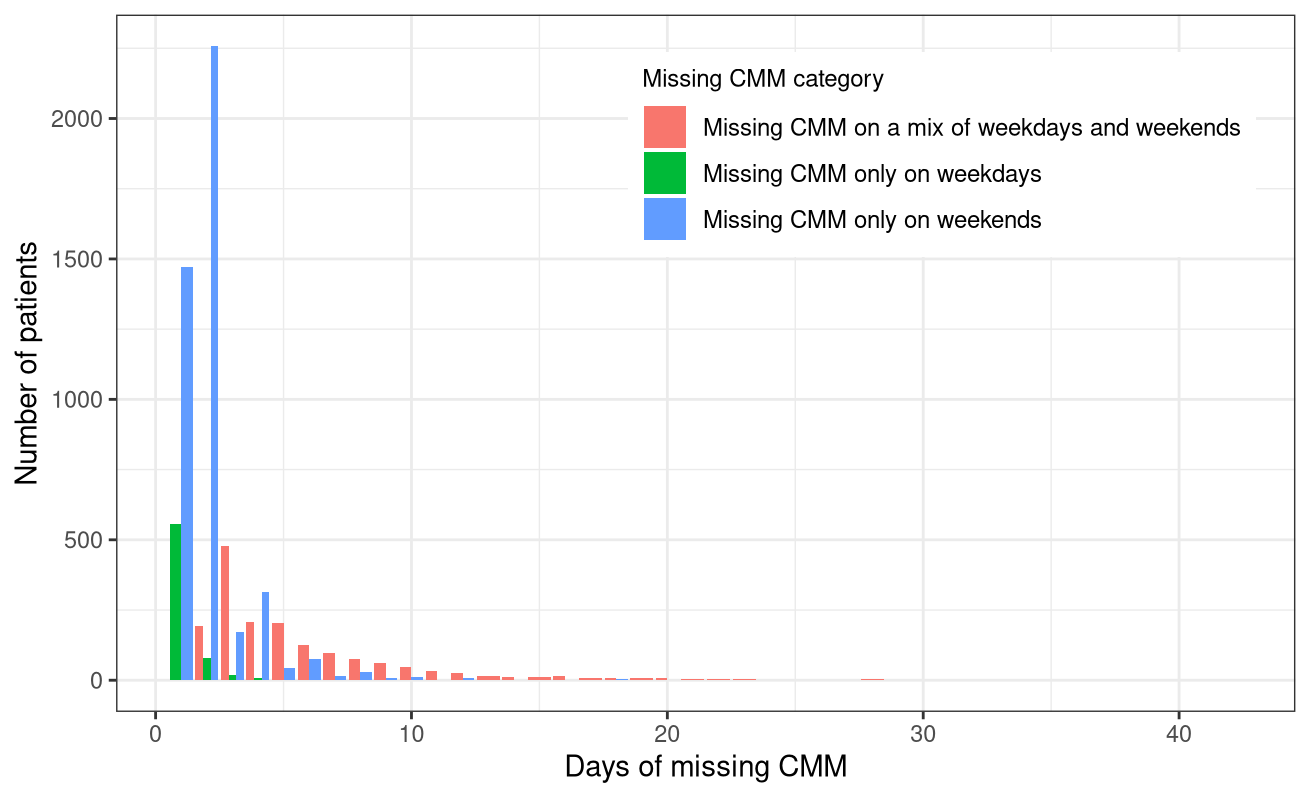

CMM: comprehensive medication management

#### eTable 3. Missed CMM Demographics

|  | n (%) |
| --- | --- |
| No CMM on day 1 of admission | 3016 (40.9) |
| No CMM on last day of admission | 554 (7.5) |
| At least one missing CMM on a weekend | 6069 (82.2) |
| Missing CMM only on weekends | 4406 (59.7) |

Percentages calculated based off denominator of 7379 patients who had at least 1 day of missing CMM

#### eTable 4. Demographics split by lack of CMM

| **Variable** | **Overall**  **(28,795)** | **CMM Every Day (21,416)** | **No CMM at least 1 day (7,379)** | **p-value** |
| --- | --- | --- | --- | --- |
| ***Primary Variable*** | | | |  |
| Pharmacist-to-patient ratio (ICU patients) | 19.6±10.2 | 20.4±10.8 | 17.1±7.8 | <0.001 |
| ***Secondary Variable*** | | | | |
| Absence of CMM for at least 1 day of ICU stay | 7,379 (25.6) | 0 (0) | 7,379 (100) | ---- |
| ***Co-variates*** | | | | |
| Age, years | 61.2±17.0 | 61.2±17.0 | 61.3±16.9 | 0.40 |
| Sex, female | 12,144 (42.2) | 9,139 (42.7) | 3,005 (40.7) | 0.01 |
| SOFA Score* | 5.2±4.1 | 5.1±4.1 | 5.7±4.3 | <0.001 |
| MRC-ICU Score* | 11.4±6.6 | 11.2±6.7 | 12.0±6.5 | <0.001 |
| ICU admission day of the week  Monday  Tuesday  Wednesday  Thursday  Friday  Saturday  Sunday | 4,634 (16.1)  4,599 (16.0)  4,364 (15.2)  4,386 (15.2)  3,852 (13.4)  3,408 (11.8)  3,549 (12.3) | 3,917 (18.3)  3,801 (17.8)  3,351 (15.6)  3,149 (14.7)  2,478 (11.6)  2,058 (9.6)  2,659 (12.4) | 717 (9.7)  798 (10.8)  1,013 (13.7)  1,237 (16.8)  1,374 (18.6)  1,350 (18.3)  890 (12.1) | <0.001 |
| ICU type**†**  Medical  Surgical/Trauma  Surgical  Cardiothoracic  Cardiac  Neurosurgery/Neurology  Mixed Medical-Surgical  Pediatric  Burn  Other | 8,850 (30.7)  3,927 (13.6)  2,560 (8.9)  2,629 (9.1)  2,498 (8.7)  3,886 (13.5)  4,195 (14.6)  35 (0.1)  185 (0.6)  27 (0.1) | 7,214 (33.7)  2,585 (12.1)  1,926 (9.0)  1,776 (8.3)  1,868 (8.7)  3,078 (14.4)  2,772 (12.9)  23 (0.1)  156 (0.7)  17 (0.1) | 1,636 (22.2)  1,342 (18.2)  634 (8.6)  853 (11.6)  630 (8.5)  808 (11.0)  1,423 (19.3)  12 (0.2)  29 (0.4)  10 (0.1) | <0.001 |
| Hospital type  Academic Medical Center  Community - Teaching  Community - Non-teaching  Government/VA/Military | 19,243 (66.8)  7,006 (24.3)  2,398 (8.3)  148 (0.5) | 15,250 (71.2)  4,581 (21.4)  1,439 (6.7)  146 (0.7) | 3,993 (54.1)  2,425 (32.9)  959 (13.0)  2 (0) | <0.001 |
| Hospital CMS star rating  2  3  4  5 | 6,345 (22.6)  14,958 (53.3)  6,354 (22.6)  424 (1.5) | 4,657 (22.4)  10,432 (50.1)  5,297 (25.5)  424 (2.0) | 1,688 (23.2)  4,526 (62.2)  1,057 (14.5)  0 (0) | <0.001 |
| ICU pharmacist coverage 1^st^ 24 hours**‡**  CMM delivered on interprofessional rounds  CMM delivered not on interprofessional rounds  Abbreviated CMM delivered  CMM not delivered (absence of CMM)  Unknown | 19,547 (67.9)  3,995 (13.9)  2,087 (7.3)  3,016 (10.5)  142 (0.5) | 16,103 (75.2)  3,365 (15.7)  1,830 (8.5)  0 (0)  112 (0.5) | 3,444 (46.7)  630 (8.5)  257 (3.5)  3,016 (40.9)  30 (0.4) | <0.001 |
| Nurse-to-patient ratio (average), 1:X  ≤1  1-2  2  >2 | 1,785 (6.4)  2,970 (10.7)  22,388 (80.5)  674 (2.4) | 1,227 (5.9)  2,033 (9.8)  16,941 (81.9)  472 (2.3) | 558 (7.8)  937 (13.1)  5,447 (76.2)  202 (2.8) | <0.001 |
| Percent of assigned ICU teams pharmacist rounded with**§** | 54.3±32.6 | 53.4±32.9 | 56.8±31.6 | <0.001 |
| Percent days, no pharmacist or pharmacist trainee | 32.2±34.1 | 25.9±33.6 | 50.9±28.1 | <0.001 |
| Percent days, only attending physician**⁂** | 8.8±26.7 | 8.0±25.8 | 11.6±28.9 | <0.001 |
| ***Outcomes*** | | | | |
| Hospital mortality | 4,224 (14.7) | 2,659 (12.4) | 1,267 (17.2) | <0.001 |
| Hospital LOS, days | 12.3±17.0 | 11.3±15.3 | 15.3±20.9 | <0.001 |
| ICU LOS, days | 5.8±9.6 | 5.0±8.2 | 8.3±12.4 | <0.001 |
| Duration of mechanical ventilation, days | 2.4±18.3 | 2.0±20.7 | 3.5±7.3 | <0.001 |

Data presented as mean (±SD) or n (%)

CMM: comprehensive medication management, CMS: Center for Medicare & Medicaid Services, ICU: intensive care unit, LOS: length of stay, MRC-ICU: medication regimen complexity- intensive care unit, SOFA: sequential organ failure assessment, VA: Veterans Affairs

Full details of missingness are available in the **Supplementary Appendix.**

*worst score during the first 24 hours of ICU stay

**†**Inclusion criteria specified ≥18 years of age; however, some institutions allowed patients ≥18 years of age in the pediatric ICU

**‡**CMM delivered on interprofessional rounds: the pharmacist attended multidisciplinary rounds and verbally provided CMM (including review of medications and recommendations); CMM delivered not on interprofessional rounds: the pharmacist provided by CMM (reviewed medications and provided recommendations) but did not attend interdisciplinary rounds; Abbreviated CMM delivered: the pharmacist provided abbreviated CMM which includes brief medication review but does not include full patient review (e.g. progress notes, results) and recommendations were provided outside of rounds; CMM not delivered (absence of CMM): the patient received no medication review from a pharmacist outside of pharmacokinetic monitoring and prospective medication order verification

**§**Percent of assigned ICU teams pharmacist rounded is calculated by total number of ICU medical teams the pharmacist attended and provided CMM on interprofessional rounds for divided by total number of ICU medical teams providing care for patients that the pharmacist was assigned to provide CMM for

**⁂**Only attending physician refers to composition of the primary medical team. On these days, the patient received care from only the attending physician and did not receive care from any medical residents, fellows, or advanced practice providers.

#### eTable 5. Unadjusted and adjusted odds ratios for GEE for primary outcome of mortality

| **Mortality Model (N=28,795)** | | | | |
| --- | --- | --- | --- | --- |
| **Variable** | **Univariable** | | **Multivariable** | |
|  | **Odds Ratio (CI)** | **P-value** | **Odds Ratio (CI)** | **P-value** |
| *Primary Variable* | | | | |
| Pharmacist-to-patient ratio (ICU patients) | 1.00 (1.00–1.01) | 0.40 | 1.01 (1.00-1.01) | 0.04 |
| *Secondary Variable* | | | | |
| Absence of CMM for at least 1 day of ICU stay | 1.60 (1.36–1.87) | <0.001 | 1.20 (1.03–1.40) | 0.02 |
| *Co-variates* | | | | |
| Age, years | 1.02 (1.02–1.02) | <0.001 | 1.03 (1.02–1.03) | <0.001 |
| Sex, female | 0.98 (0.91–1.05) | 0.58 | 1.02 (0.94–1.11) | 0.64 |
| SOFA Score* | 1.25 (1.23–1.27) | <0.001 | 1.31 (1.27–1.35) | <0.001 |
| MRC-ICU Score* | 1.09 (1.08–1.10) | <0.001 | 1.04 (1.03–1.06) | <0.001 |
| SOFA-MRC-ICU interaction term | 1.008 (1.007–1.008) | <0.001 | 1.00 (1.00-1.00) | <0.001 |
| ICU admission day of the week (reference: Monday) | - | - | - | - |
| Tuesday | 0.96 (0.85–1.07) | 0.45 | 0.97 (0.86–1.11) | 0.65 |
| Wednesday | 0.97 (0.86–1.09) | 0.62 | 0.93 (0.83–1.05) | 0.24 |
| Thursday | 0.98 (0.86–1.10) | 0.68 | 0.95 (0.83–1.09) | 0.48 |
| Friday | 1.01 (0.89–1.14) | 0.90 | 0.93 (0.81–1.06) | 0.26 |
| Saturday | 1.15 (1.01–1.31) | 0.04 | 1.00 (0.87–1.15) | 0.98 |
| Sunday | 1.08 (0.95–1.22) | 0.23 | 1.00 (0.87–1.15) | 0.99 |
| ICU type† (reference: medical ICU) | - | - | - | - |
| Surgical/Trauma | 0.54 (0.46–0.65) | <0.001 | 0.75 (0.63–0.89) | 0.001 |
| Surgical | 0.49 (0.38–0.64) | <0.001 | 0.49 (0.37–0.65) | <0.001 |
| Cardiothoracic | 0.36 (0.27–0.50) | <0.001 | 0.23 (0.13–0.38) | <0.001 |
| Cardiac | 0.74 (0.60–0.91) | 0.005 | 0.90 (0.74–1.10) | 0.32 |
| Neurosurgery/Neurology | 0.52 (0.44–0.61) | <0.001 | 1.13 (0.91–1.39) | 0.27 |
| Pediatric | 0.23 (0.12–0.46) | <0.001 | 0.70 (0.30–1.63) | 0.41 |
| Mixed Medical-Surgical | 0.75 (0.61–0.94) | 0.01 | 0.76 (0.61–0.94) | 0.01 |
| Burn | 0.32 (0.20–0.51) | <0.001 | 0.53 (0.29–0.97) | 0.04 |
| Other | 0.22 (0.08–0.58) | 0.003 | 0.52 (0.20–1.36) | 0.18 |
| Hospital type (reference: academic medical center) | - | - | - | - |
| Community - Teaching | 0.98 (0.76–1.26) | 0.88 | 0.99 (0.78–1.27) | 0.96 |
| Community - Non-teaching | 0.89 (0.64–1.24) | 0.49 | 0.89 (0.67–1.19) | 0.44 |
| Government/VA/Military | 0.68 (0.58–0.80) | <0.001 | 1.07 (0.80–1.41) | 0.66 |
| Hospital CMS star rating (reference: 3) | - | - | - | - |
| 2 | 0.92 (0.74–1.15) | 0.47 | 1.11 (0.92–1.35) | 0.26 |
| 4 | 1.06 (0.80–1.40) | 0.67 | 0.85 (0.63–1.15) | 0.29 |
| 5 | 0.87 (0.45–1.69) | 0.68 | 0.52 (0.30–0.88) | 0.02 |
| ICU pharmacist coverage 1st 24 hours‡ (reference: CMM delivered on interprofessional rounds) | - | - | - | - |
| CMM delivered not on interprofessional rounds | 0.93 (0.83–1.05) | 0.25 | 1.04 (0.92–1.17) | 0.51 |
| Abbreviated CMM delivered | 1.04 (0.90–1.19) | 0.61 | 1.10 (0.93–1.29) | 0.28 |
| CMM not delivered (absence of CMM) | 1.14 (0.97–1.33) | 0.1 | 1.09 (0.92–1.30) | 0.32 |
| Unknown | 1.32 (0.71–2.45) | 0.38 | 1.11 (0.59–2.09) | 0.74 |
| Nurse-to-patient ratio (average)(reference: 1:2) | - | - | - | - |
| ≤1 | 3.41 (2.56–4.55) | <0.001 | 2.82 (1.98–4.01) | <0.001 |
| 1-2 | 2.43 (2.13–2.78) | <0.001 | 1.61 (1.33–1.96) | <0.001 |
| >2 | 0.95 (0.70–1.29) | 0.76 | 0.87 (0.69–1.10) | 0.24 |
| Percent of assigned ICU teams pharmacist rounded with§ | 1.10 (0.87–1.38) | 0.42 | 1.30 (1.03–1.65) | 0.03 |
| Percent days, no pharmacist or pharmacist trainee | 1.00 (1.00–1.00) | 0.79 | 1.00 (1.00–1.00) | 0.64 |
| Percent days, only attending physician⁂ | 1.00 (1.00–1.00) | 0.08 | 1.00 (1.00–1.00) | 0.97 |

CI: confidence interval, CMM: comprehensive medication management, CMS: Center for Medicare & Medicaid Services, ICU: intensive care unit, MRC-ICU: medication regimen complexity- intensive care unit, OR: odds ratio, SOFA: sequential organ failure assessment, VA: Veterans Affairs

*worst score during the first 24 hours of ICU stay

**†**Inclusion criteria specified ≥18 years of age; however, some institutions allowed patients ≥18 years of age in the pediatric ICU

**‡**CMM delivered on interprofessional rounds: the pharmacist attended multidisciplinary rounds and verbally provided CMM (including review of medications and recommendations); CMM delivered not on interprofessional rounds: the pharmacist provided by CMM (reviewed medications and provided recommendations) but did not attend interdisciplinary rounds; Abbreviated CMM delivered: the pharmacist provided abbreviated CMM which includes brief medication review but does not include full patient review (e.g. progress notes, results) and recommendations were provided outside of rounds; CMM not delivered (absence of CMM) (absence of CMM): the patient received no medication review from a pharmacist outside of pharmacokinetic monitoring and prospective medication order verification

**§**Percent of assigned ICU teams pharmacist rounded is calculated by total number of ICU medical teams the pharmacist attended and provided CMM on interprofessional rounds for divided by total number of ICU medical teams providing care for patients that the pharmacist was assigned to provide CMM for

**⁂**Only attending physician refers to composition of the primary medical team. On these days, the patient received care from only the attending physician and did not receive care from any medical residents, fellows, or advanced practice providers.

#### eTable 6. Patient Demographics with Missing Data Reported

|  | Overall  (28,795) | Deceased  (4,224) | Alive  (24,571) | p-value |
| --- | --- | --- | --- | --- |
| ***Primary Variable*** | | | |  |
| Pharmacist-to-patient ratio (ICU patients only) | 19.5±10.2 | 19.5±10.2 | 19.6±10.2 | 0.7 |
| Missing data | 453 (1.6) | 45 (1.1) | 408 (1.7) |  |
| ***Secondary Variable*** | | | | |
| Absence of CMM for at least 1 day of ICU stay | 21416 (74.4) | 2957 (70.0) | 18459 (75.1) | <0.001 |
| ***Co-variates*** | | | | |
| Age, years | 61.2±17.0 | 65.5±15.5 | 60.±17.12 | <0.001 |
| Sex, female | 12,155 (42.2) | 1,770 (41.9) | 10,374 (42.2) | 0.5 |
| Missing data | 1 (<0.1) | 0 | 1 (<0.1%) |  |
| SOFA Score* | 5.2±4.1 | 8.7±4.4 | 4.6±3.8 | <0.001 |
| Missing data | 35 (0.1) | 4 (<0.1) | 31 (0.1) |  |
| MRC-ICU Score* | 11.4±6.6 | 14.6±7.1 | 10.9±6.4 | <0.001 |
| Missing Data | 4 (<0.1) | 0 | 4 (<0.1) |  |
| ICU admission day of the week  Monday  Tuesday  Wednesday  Thursday  Friday  Saturday  Sunday | 4,634 (16.1)  4,599 (16.0)  4,364 (15.2)  4,386 (15.2)  3,852 (13.4)  3,408 (11.8)  3,549 (12.3) | 669 (15.8)  640 (15.2)  615 (14.6)  623 (14.7)  558 (13.2)  563 (13.3)  556 (13.2) | 3,965 (16.1)  3,959 (16.1)  3,749 (15.3)  3,763 (15.3)  3,294 (13.4)  2,845 (11.6)  2,993 (12.2) | 0.01 |
| Missing Data | 3 (<0.1) | 0 | 3 (<0.1) |  |
| Dialysis | 3371 (11.7) | 1161 (27.5) | 2210 (9.0) | <0.001 |
| Missing data | 5 (0.1) | 2 (<0.1) | 3 (<0.1) |  |
| ECMO | 324 (1.1) | 144 (3.4) | 180 (0.7) | <0.001 |
| Missing Data | 3 (<0.1) | 1 (<0.1) | 2 (<0.1) |  |
| Mechanical circulatory support | 13972 (48.5) | 3307 (78.3) | 10665 (43.4) | <0.001 |
| Missing Data | 10 (<0.1) | 2 (<0.1) | 8 (<0.1) |  |
| Mechanical ventilation | 14150 (49.1) | 3183 (75.4) | 10967 (44.6) | <0.001 |
| Missing Data | 2 (<0.1) | 0 | 2 (<0.1) |  |
| ICU type†  Medical  Surgical/Trauma  Surgical  Cardiothoracic  Cardiac  Neurosurgery/Neurology  Pediatric  Mixed Medical-Surgical  Burn  Other | 8,850 (30.7)  3,927 (13.6)  2,560 (8.9)  2,629 (9.1)  2,498 (8.7)  3,886 (13.5)  35 (0.1)  4,195 (14.6)  185 (0.6)  27 (0.1) | 1,766 (41.8)  426 (10.1)  284 (6.7)  199 (4.7)  405 (9.6)  465 (11.0)  2 (0.05)  664 (15.7)  11 (0.3)  2 (<0.1) | 7,084 (28.8)  3,501 (14.3)  2,276 (9.3)  2,430 (9.9)  2,093 (8.5)  3,421(13.9)  33 (0.1)  3,531 (14.4)  174 (0.7)  25 (0.1) | <0.001 |
| Missing data | 3 (<0.1) | 0 | 3 (<0.1) |  |
| Hospital type  Academic Medical Center  Community - Teaching  Community - Non-teaching  Government/VA/Military | 19,243 (66.8)  7,006 (24.3)  2,398 (8.3)  148 (0.5) | 2,868 (67.9)  1,034 (24.5)  306 (7.2)  16 (0.4) | 16,375 (66.6)  5,972 (24.3)  2,092 (8.5)  132 (0.5) | 0.02 |
| Hospital CMS star rating  2  3  4  5 | 6,345 (22.6)  14,958 (53.3)  6,354 (22.6)  424 (1.5) | 870 (21.7)  2,151 (53.7)  931 (23.3)  50 (1.2) | 5,475 (22.7)  12,807 (53.2)  5,423 (22.5)  374 (1.6) | 0.20 |
| Missing data | 714 (2.5) | 222 (5.3) | 492 (2.0) |  |
| Pharmacist-to-patient ratio (all patients) | 27.4±22.6 | 26.4±19.5 | 27.6±23.1 | 0.08 |
| Missing data | 453 (1.6) | 45 (1.1) | 408 (1.7) |  |
| ICU pharmacist coverage 1^st^ 24 hours  CMM on interprofessional rounds  CMM delivered not on interprofessional rounds  Abbreviated CMM delivered only  CMM not delivered (absence of CMM)  Unknown | 19,547 (67.9)  3,995 (13.9)  2,087 (7.3)  3,016 (10.5)  141 (0.5) | 2,885 (68.3)  527 (12.5)  296 (7.0)  489 (11.6)  26 (0.6) | 16,662 (67.8)  3,468 (14.1)  1,791 (7.3)  2,527 (10.3)  116 (0.5) | 0.01 |
| Missing data | 8 (<0.1) | 1 (<0.1) | 7 (<0.1) |  |
| Nurse-to-patient ratio (average), 1:X  ≤1  1-2  2  >2 | 1,785 (6.4)  2,970 (10.7)  22,388 (80.5)  674 (2.4) | 461 (11.2)  696 (16.9)  2,878 (69.8)  87 (2.1) | 1,324 (5.6)  2,274 (9.6)  19,510 (82.3)  587 (2.5) | <0.001 |
| Missing data | 978 (3.4) | 102 (2.4) | 876 (3.6) |  |
| Percent of assigned ICU teams pharmacist rounded with§ | 54.3 (32.6) | 56.4 (31.5) | 53.9 (32.8) | <0.001 |
| Missing data | 518 (1.8) | 49 (1.2) | 469 (1.9) |  |
| Percent days, no pharmacist or pharmacist trainee | 32.2 (34.1) | 31.1 (30.7) | 32.4 (34.6) | 0.60 |
| Missing data | 1045 (3.6) | 227 (5.4) | 818 (3.3) |  |
| Percent days, only attending physician⁂ | 8.9 (26.7) | 8.7 (25.9) | 8.9 (26.8) | 0.30 |
| Missing data | 1768 (6.1) | 356 (8.4) | 1412 (5.7) |  |
| ***Outcomes*** | | | |  |
| Hospital mortality | 4,224 (14.7) | 4,224 (100) | 0 | <0.001 |
| Hospital LOS, days | 12.3±17.0 | 14.1±19.6 | 12.0±16.5 | <0.001 |
| Missing data | 4 (<0.1) | 0 | 4 (<0.1) |  |
| ICU LOS, days | 5.8±9.6 | 8.7±12.8 | 5.3±8.8 | <0.001 |
| ICU free days | 19.8±9.5 | 0±0 | 23.1±5.5 | <0.001 |
| Missing Data | 245 (0.9) | 187 (4.4) | 58 (0.2) |  |
| Duration of mechanical ventilation, days | 2.4±18.3 | 5.1±8.0 | 1.9±19.4 | <0.001 |
| Missing Data | 4 (<0.1) | 0 | 4 (<0.1) |  |
| VFDs | 22.5±10.1 | 0±0 | 26.4±4.1 | <0.001 |
| Missing Data | 4 (<0.1) | 0 | 4 (<0.1) |  |

Data presented as mean (±SD) or n (%)

CMM: comprehensive medication management, CMS: Center for Medicare & Medicaid Services, ECMO: extracorporeal membrane oxygenation, ICU: intensive care unit, LOS: length of stay, MRC-ICU: medication regimen complexity- intensive care unit, SOFA: sequential organ failure assessment, VA: Veterans Affairs, VFDs: ventilator free days

No missing data if unreported

*worst score during the first 24 hours of ICU stay

**†**Inclusion criteria specified ≥18 years of age; however, some institutions allowed patients ≥18 years of age in the pediatric ICU

**‡**CMM delivered on interprofessional rounds: the pharmacist attended multidisciplinary rounds and verbally provided CMM (including review of medications and recommendations); CMM delivered not on interprofessional rounds: the pharmacist provided by CMM (reviewed medications and provided recommendations) but did not attend interdisciplinary rounds; Abbreviated CMM delivered: the pharmacist provided abbreviated CMM which includes brief medication review but does not include full patient review (e.g. progress notes, results) and recommendations were provided outside of rounds; CMM not delivered (absence of CMM) (absence of CMM): the patient received no medication review from a pharmacist outside of pharmacokinetic monitoring and prospective medication order verification

**§**Percent of assigned ICU teams pharmacist rounded is calculated by total number of ICU medical teams the pharmacist attended and provided CMM on interprofessional rounds for divided by total number of ICU medical teams providing care for patients that the pharmacist was assigned to provide CMM for

**⁂**Only attending physician refers to composition of the primary medical team. On these days, the patient received care from only the attending physician and did not receive care from any medical residents, fellows, or advanced practice providers.

#### eTable 7. Unadjusted and adjusted odds ratios for GEE model for primary outcome of mortality using Pharmacist-to-all-patient ratio

| **Mortality Model (N=28,795)** | | | | |
| --- | --- | --- | --- | --- |
| **Variable** | **Univariable** | | **Multivariable** | |
|  | **Odds Ratio (CI)** | **P-value** | **Odds Ratio (CI)** | **P-value** |
| *Primary Variable* | | | | |
| Pharmacist-to-patient ratio (all patients) | 1.00 (1.00–1.00) | 0.31 | 1.00 (1.00–1.00) | 0.96 |
| *Secondary Variable* | | | | |
| Absence of CMM for at least 1 day of ICU stay | 1.59 (1.35–1.87) | <0.001 | 1.22 (1.03–1.45) | 0.02 |
| *Co-variates* | | | | |
| Age, years | 1.02 (1.02–1.02) | <0.001 | 1.03 (1.02–1.03) | <0.001 |
| Sex, female | 0.98 (0.91–1.05) | 0.58 | 1.02 (0.94–1.12) | 0.63 |
| SOFA Score* | 1.25 (1.23–1.27) | <0.001 | 1.31 (1.27–1.35) | <0.001 |
| MRC-ICU Score* | 1.09 (1.08–1.10) | <0.001 | 1.04 (1.03–1.06) | <0.001 |
| SOFA-MRC-ICU interaction term | 1.01 (1.01–1.01) | <0.001 | 1.00 (1.00–1.00) | <0.001 |
| ICU admission day of the week (reference: Monday) | - | - | - | - |
| Tuesday | 0.96 (0.85–1.07) | 0.45 | 0.97 (0.86–1.10) | 0.66 |
| Wednesday | 0.97 (0.86–1.09) | 0.62 | 0.93 (0.83–1.05) | 0.25 |
| Thursday | 0.98 (0.86–1.10) | 0.68 | 0.96 (0.84–1.10) | 0.55 |
| Friday | 1.01 (0.89–1.14) | 0.90 | 0.94 (0.82–1.08) | 0.35 |
| Saturday | 1.15 (1.01–1.31) | 0.04 | 1.01 (0.87–1.17) | 0.91 |
| Sunday | 1.08 (0.95–1.22) | 0.23 | 1.00 (0.87–1.15) | 0.98 |
| ICU type† (reference: medical ICU) | - | - | - | - |
| Surgical/Trauma | 0.54 (0.46–0.65) | <0.001 | 0.75 (0.63–0.90) | 0.002 |
| Surgical | 0.49 (0.38–0.64) | <0.001 | 0.49 (0.37–0.64) | <0.001 |
| Cardiothoracic | 0.36 (0.27–0.50) | <0.001 | 0.23 (0.13–0.38) | <0.001 |
| Cardiac | 0.74 (0.60–0.91) | 0.005 | 0.90 (0.74–1.10) | 0.31 |
| Neurosurgery/Neurology | 0.52 (0.44–0.61) | <0.001 | 1.12 (0.91–1.39) | 0.29 |
| Pediatric | 0.23 (0.12–0.46) | <0.001 | 0.70 (0.30–1.62) | 0.40 |
| Mixed Medical-Surgical | 0.75 (0.61–0.94) | 0.01 | 0.75 (0.60–0.94) | 0.01 |
| Burn | 0.32 (0.20–0.51) | <0.001 | 0.53 (0.29–0.96) | 0.04 |
| Other | 0.22 (0.08–0.58) | 0.003 | 0.52 (0.20–1.40) | 0.20 |
| Hospital type (reference: academic medical center) | - | - | - | - |
| Community - Teaching | 0.98 (0.76–1.26) | 0.88 | 0.99 (0.76–1.28) | 0.92 |
| Community - Non-teaching | 0.89 (0.64–1.24) | 0.49 | 0.88 (0.65–1.19) | 0.42 |
| Government/VA/Military | 0.68 (0.58–0.80) | <0.001 | 0.98 (0.74–1.30) | 0.90 |
| Hospital CMS star rating (reference: 3) |  |  |  |  |
| 2 | 0.92 (0.74–1.15) | 0.47 | 1.12 (0.92–1.35) | 0.26 |
| 4 | 1.06 (0.80–1.40) | 0.67 | 0.84 (0.61–1.15) | 0.27 |
| 5 | 0.87 (0.45–1.69) | 0.68 | 0.51 (0.30–0.88) | 0.02 |
| ICU pharmacist coverage 1st 24 hours‡ (reference: CMM delivered on interprofessional rounds) | - | - | - | - |
| CMM delivered not on interprofessional rounds | 0.93 (0.83–1.05) | 0.25 | 1.03 (0.92–1.16) | 0.61 |
| Abbreviated CMM delivered | 1.04 (0.90–1.19) | 0.61 | 1.12 (0.95–1.32) | 0.19 |
| CMM not delivered (absence of CMM) | 1.14 (0.97–1.33) | 0.1 | 1.03 (0.86–1.23) | 0.78 |
| Unknown | 1.32 (0.71–2.45) | 0.38 | 1.10 (0.59–2.06) | 0.75 |
| Nurse-to-patient ratio (average)(reference: 1:2) | - | - | - | - |
| ≤1 | 3.41 (2.56–4.55) | <0.001 | 2.78 (1.94–3.96) | <0.001 |
| 1-2 | 2.43 (2.13–2.78) | <0.001 | 1.61 (1.32–1.95) | <0.001 |
| >2 | 0.95 (0.70–1.29) | 0.76 | 0.87 (0.70–1.10) | 0.24 |
| Percent of assigned ICU teams pharmacist rounded with§ | 1.10 (0.87–1.38) | 0.42 | 1.24 (0.98–1.55) | 0.07 |
| Percent days, no pharmacist or pharmacist trainee on rounds | 1.00 (1.00–1.00) | 0.79 | 1.00 (1.00–1.00) | 0.94 |
| Percent days, only attending physician⁂ | 1.00 (1.00–1.00) | 0.08 | 1.00 (1.00–1.00) | 0.99 |

CMM: comprehensive medication management, CMS: Center for Medicare & Medicaid Services, ICU: intensive care unit, MRC-ICU: medication regimen complexity- intensive care unit, SOFA: sequential organ failure assessment, VA: Veterans Affairs
*worst score during the first 24 hours of ICU stay

**†**Inclusion criteria specified ≥18 years of age; however, some institutions allowed patients ≥18 years of age in the pediatric ICU

**‡**CMM delivered on interprofessional rounds: the pharmacist attended multidisciplinary rounds and verbally provided CMM (including review of medications and recommendations); CMM delivered not on interprofessional rounds: the pharmacist provided by CMM (reviewed medications and provided recommendations) but did not attend interdisciplinary rounds; Abbreviated CMM delivered: the pharmacist provided abbreviated CMM which includes brief medication review but does not include full patient review (e.g. progress notes, results) and recommendations were provided outside of rounds; CMM not delivered (absence of CMM) (absence of CMM): the patient received no medication review from a pharmacist outside of pharmacokinetic monitoring and prospective medication order verification

**§**Percent of assigned ICU teams pharmacist rounded is calculated by total number of ICU medical teams the pharmacist attended and provided CMM on interprofessional rounds for divided by total number of ICU medical teams providing care for patients that the pharmacist was assigned to provide CMM for

**⁂**Only attending physician refers to composition of the primary medical team. On these days, the patient received care from only the attending physician and did not receive care from any medical residents, fellows, or advanced practice providers.

This eTable includes the pharmacist-to-patient ratio using all patients that the pharmacist cared for (both floor and ICU patients) as opposed to the main analysis which used a pharmacist-to-patient ratio including only ICU patients.

#### eTable 8. Fine-Gray Sub-Distribution Hazards Regression for Hospital LOS, ICU LOS, and VFDs

|  | Hospital LOS  N=28791 | | ICU LOS  N=28791 | | VFD  N=28789 | |
| --- | --- | --- | --- | --- | --- | --- |
|  | adjusted Hazard Ratio (95% CI) | p-value | adjusted Hazard Ratio (95% CI) | p-value | adjusted Hazard Ratio (95% CI) | p-value |
| *Primary Variable* | | |  |  |  |  |
| Pharmacist-to-patient ratio (ICU patients) | 0.99 (0.99, 1.00) | <0.001 | 0.99 (0.98, 0.99) | <0.001 | 0.99 (0.99, 1.00) | <0.001 |
| *Secondary Variable* | | | | | | |
| Absence of CMM for at least 1 day of ICU stay | 0.71 (0.66, 0.77) | <0.001 | 0.60 (0.55, 0.66) | <0.001 | 0.81 (0.76, 0.85) | <0.001 |
| *Co-variates* | | | | | | |
| Age, years | 0.99 (0.99, 0.99) | <0.001 | 1.00 (1.00, 1.00) | 0.006 | 1.00 (0.99, 1.00) | <0.001 |
| Sex, female | 1.00 (0.97, 1.03) | 0.9 | 1.02 (0.99, 1.04) | 0.20 | 1.00 (0.97, 1.02) | 0.75 |
| SOFA Score* | 0.90 (0.89, 0.91) | <0.001 | 0.93 (0.92, 0.94) | <0.001 | 0.91 (0.90, 0.91) | <0.001 |
| MRC-ICU Score* | 0.98 (0.97, 0.98) | <0.001 | 0.98 (0.97, 0.98) | <0.001 | 0.97 (0.97, 0.97) | <0.001 |
| ICU admission day of the week (reference: Monday)  Tuesday  Wednesday  Thursday  Friday  Saturday  Sunday | 1.02 (0.98, 1.07)  1.03 (0.98, 1.07)  1.03 (0.98, 1.08)  0.97 (0.91, 1.03)  0.98 (0.91, 1.04)  0.96 (0.91, 1.01) | 0.33  0.26  0.20  0.27  0.47  0.15 | 1.02 (0.98, 1.07)  1.03 (0.99, 1.07)  1.04 (1.00, 1.09)  0.93 (0.88, 0.98)  0.93 (0.87, 1.00)  0.99 (0.94, 1.04) | 0.31  0.13  0.06  0.01  0.04  0.64 | 1.00 (0.96, 1.05)  1.01 (0.98, 1.04)  1.03 (0.99, 1.07)  0.99 (0.94, 1.03)  0.95 (0.90, 1.01)  0.95 (0.91, 1.00) | 0.85  0.59  0.21  0.56  0.09  0.04 |
| ICU type† (reference: medical ICU)  Surgical/Trauma  Surgical  Cardiothoracic  Cardiac  Neurosurgery/Neurology  Pediatric  Mixed Medical-Surgical  Burn  Other | 1.09 (0.93, 1.29)  1.03 (0.88, 1.21)  1.62 (1.34, 1.96)  1.11 (0.96, 1.28)  0.94 (0.82, 1.07)  1.20 (0.89, 1.62)  1.05 (0.92, 1.20)  0.82 (0.56, 1.19)  1.52 (0.86, 2.70) | 0.29  0.72  <0.001  0.17  0.36  0.23  0.44  0.30  0.15 | 0.98 (0.84, 1.14)  1.11 (0.92, 1.32)  1.25 (1.00, 1.55)  0.97 (0.82, 1.15)  0.82 (0.71, 0.95)  1.06 (0.82, 1.37)  1.30 (1.10, 1.54)  0.57 (0.46, 0.71)  1.37 (1.01, 1.85) | 0.83  0.27  0.05  0.73  0.007  0.65  0.003  <0.001  0.04 | 1.07 (1.00, 1.14)  1.27 (1.14, 1.41)  1.64 (1.44, 1.87)  1.11 (1.03, 1.19)  0.93 (0.86, 0.99)  1.14 (0.98, 1.33)  1.14 (1.05, 1.25)  1.05 (0.85, 1.29)  1.36 (1.16, 1.60) | 0.07  <0.001  <0.001  0.005  0.03  0.10  0.002  0.65  <0.001 |
| Hospital type (reference: academic medical center)  Community - Teaching  Community - Non-teaching  Government/VA/Military | 1.02 (0.90, 1.14)  1.82 (1.55, 2.13)  2.92 (2.17, 3.95) | 0.79  <0.001  <0.001 | 1.14 (1.02, 1.27)  0.82 (0.69, 0.98)  0.71 (0.58, 0.86) | 0.02  0.03  <0.001 | 0.99 (0.93, 1.05)  1.03 (0.94, 1.13)  1.08 (1.01, 1.16) | 0.64  0.57  0.03 |
| Hospital CMS star rating (reference: 3)  2  4  5 | 0.86 (0.78, 0.96)  0.82 (0.74, 0.92)  0.83 (0.70, 0.99) | 0.006  <0.001  0.04 | 1.09 (0.98, 1.22)  1.16 (1.03, 1.31)  1.41 (0.91, 2.17) | 0.13  0.02  0.12 | 1.01 (0.96, 1.07)  1.04 (0.97, 1.11)  1.12 (0.96, 1.30) | 0.66  0.25  0.14 |
| ICU pharmacist coverage 1st 24 hours‡ (reference: CMM delivered on interprofessional rounds)  CMM delivered not on interprofessional rounds  Abbreviated CMM delivered  CMM not delivered (absence of CMM)  Unknown | 0.96 (0.91, 1.02)  0.95 (0.87, 1.03)  1.12 (1.04, 1.21)  0.81 (0.56, 1.19) | 0.23  0.20  0.004  0.29 | 0.99 (0.94, 1.05)  1.00 (0.91, 1.09)  1.16 (1.07, 1.25)  0.66 (0.52, 0.85) | 0.72  0.99  <0.001  0.001 | 0.99 (0.94, 1.03)  1.00 (0.94, 1.06)  1.07 (1.00, 1.14)  0.79 (0.66, 0.95) | 0.59  0.90  0.03  0.01 |
| Nurse-to-patient ratio (average)(reference: 1:2)  ≤1  1-2  >2 | 0.73 (0.63, 0.85)  0.70 (0.65, 0.76)  0.91 (0.81, 1.02) | <0.001  <0.001  0.11 | 0.84 (0.57, 1.23)  0.67 (0.63, 0.72)  0.98 (0.81, 1.19) | 0.36  <0.001  0.86 | 0.78 (0.63, 0.96)  0.77 (0.73, 0.82)  1.04 (0.94, 1.14) | 0.02  <0.001  0.46 |
| Percent of assigned ICU teams pharmacist rounded with§ | 0.97 (0.84, 1.12) | 0.64 | 0.84 (0.72, 0.97) | 0.02 | 0.90 (0.84, 0.97) | 0.007 |
| Percent days, no pharmacist or pharmacist trainee on rounds | 1.00 (1.00, 1.00) | 0.18 | 1.00 (1.00, 1.00) | 0.003 | 1.00 (1.00, 1.00) | 0.02 |
| Percent days, only attending physician⁂ | 1.00 (1.00, 1.00) | 0.65 | 1.00 (1.00, 1.00) | 0.04 | 1.00 (1.00, 1.00) | 0.35 |

CI: confidence interval, CMM: comprehensive medication management, CMS: Center for Medicare & Medicaid Services, ICU: intensive care unit, LOS: length of stay, MRC-ICU: medication regimen complexity- intensive care unit, OR: odds ratio, SOFA: sequential organ failure assessment, VA: Veterans Affairs, VFD: ventilator free days

*worst score during the first 24 hours of ICU stay

**†**Inclusion criteria specified ≥18 years of age; however, some institutions allowed patients ≥18 years of age in the pediatric ICU

**‡**CMM delivered on interprofessional rounds: the pharmacist attended multidisciplinary rounds and verbally provided CMM (including review of medications and recommendations); CMM delivered not on interprofessional rounds: the pharmacist provided by CMM (reviewed medications and provided recommendations) but did not attend interdisciplinary rounds; Abbreviated CMM delivered: the pharmacist provided abbreviated CMM which includes brief medication review but does not include full patient review (e.g. progress notes, results) and recommendations were provided outside of rounds; CMM not delivered (absence of CMM) (absence of CMM): the patient received no medication review from a pharmacist outside of pharmacokinetic monitoring and prospective medication order verification

**§**Percent of assigned ICU teams pharmacist rounded is calculated by total number of ICU medical teams the pharmacist attended and provided CMM on interprofessional rounds for divided by total number of ICU medical teams providing care for patients that the pharmacist was assigned to provide CMM for

**⁂**Only attending physician refers to composition of the primary medical team. On these days, the patient received care from only the attending physician and did not receive care from any medical residents, fellows, or advanced practice providers.

This eTable examines likelihood of discharge, and odds ratios under 1 indicate decreased likelihood of discharge, whereas odds ratios greater than 1 indicate increased likelihood of discharge. As pharmacist-to-patient ratio increases, there is a decreased likelihood of discharge.

#### eTable 9. Univariable and Multivariable GEE Results for Mortality Using Secondary Variable of Patients Who Had CMM and Rounding Every Day of ICU Admission

|  | **Univariable** | | **Multivariable** | |
| --- | --- | --- | --- | --- |
| **Variable** | **Adjusted OR (95% CI)** | **p-value** | **Adjusted OR (95% CI)** | **p-value** |
| *Primary Variable* | | |  |  |
| Pharmacist-to-patient ratio (ICU patients) | 1.00 (1.00–1.01) | 0.4 | 1.00 (1.00–1.01) | 0.49 |
| *Secondary Variable* |  |  |  |  |
| CMM delivered on interprofessional rounds every day of ICU stay | 0.62 (0.55–0.71) | <0.001 | 0.72 (0.63–0.83) | <0.001 |
| *Co-variates* |  |  |  |  |
| Age, years | 1.02 (1.02–1.02) | <0.001 | 1.03 (1.02–1.03) | <0.001 |
| Sex, female | 0.98 (0.91–1.05) | 0.58 | 1.02 (0.94–1.12) | 0.58 |
| SOFA Score* | 1.25 (1.23–1.27) | <0.001 | 1.30 (1.26–1.34) | <0.001 |
| MRC-ICU Score* | 1.09 (1.08–1.10) | <0.001 | 1.04 (1.02–1.05) | <0.001 |
| SOFA-MRC-ICU interaction term | 1.01 (1.01–1.01) | <0.001 | 1.00 (1.00–1.00) | <0.001 |
| ICU admission day of the week (reference: Monday)  Tuesday  Wednesday  Thursday  Friday  Saturday  Sunday | 0.96 (0.85-1.07)  0.97 (0.86-1.09)  0.98 (0.86-1.10)  1.01 (0.89-1.14)  1.15 (1.01-1.31)  1.08 (0.95-1.22) | 0.45  0.62  0.68  0.90  0.04  0.23 | 0.97 (0.86-1.09)  0.91 (0.81-1.03)  0.93 (0.81-1.06)  0.91 (0.80-1.04)  0.99 (0.85-1.15)  1.00 (0.87-1.15) | 0.60  0.14  0.27  0.18  0.91  0.99 |
| ICU type† (reference: medical ICU)  Surgical/Trauma  Surgical  Cardiothoracic  Cardiac  Neurosurgery/Neurology  Pediatric  Mixed Medical-Surgical  Burn  Other | 0.54 (0.46-0.65)  0.49 (0.38–0.64)  0.36 (0.27–0.50)  0.74 (0.60–0.91)  0.52 (0.44–0.61)  0.23 (0.12–0.46)  0.75 (0.61–0.94)  0.32 (0.20–0.51)  0.22 (0.08–0.58) | <0.001  <0.001  <0.001  0.005  <0.001  <0.001  0.01  <0.001  0.003 | 0.73 (0.61–0.88)  0.47 (0.36–0.63)  0.23 (0.13–0.39)  0.89 (0.73–1.07)  1.09 (0.89–1.35)  0.70 (0.31–1.60)  0.73 (0.59–0.91)  0.52 (0.29–0.92)  0.51 (0.19–1.40) | <0.001  <0.001  <0.001  0.22  0.40  0.40  0.004  0.03  0.19 |
| Hospital type (reference: academic medical center)  Community - Teaching  Community - Non-teaching  Government/VA/Military | 0.98 (0.76–1.26)  0.89 (0.64–1.24) 0.68(0.58–0.80) | 0.88  0.49  <0.001 | 1.06 (0.82–1.35)  0.99 (0.74–1.32)  1.01 (0.76–1.33) | 0.67  0.93  0.97 |
| Hospital CMS star rating (reference: 3)  2  4  5 | 0.92 (0.74-1.15)  1.06 (0.80-1.69)  0.87 (0.45-1.69) | 0.47  0.67  0.68 | 1.12 (0.93-1.37)  0.86 (0.64-1.16)  0.56 (0.33-0.97) | 0.24  0.32  0.04 |
| ICU pharmacist coverage 1st 24 hours‡ (reference: CMM delivered on interprofessional rounds)  CMM delivered not on interprofessional rounds  Abbreviated CMM  CMM not delivered (absence of CMM)  Unknown | 0.93 (0.83–1.05)  1.04 (0.90–1.19)  1.14 (0.97–1.33)  1.32 (0.71–2.45) | 0.25  0.61  0.10  0.38 | 0.93 (0.84–1.04)  1.01 (0.87–1.19)  1.05 (0.89–1.25)  1.09 (0.59–2.03) | 0.21  0.87  0.54  0.77 |
| Nurse-to-patient ratio (average)(reference: 1:2)  ≤1  1-2  >2 | 3.41 (2.56–4.55)  2.43 (2.13–2.78)  0.95 (0.70–1.29) | <0.001  <0.001  0.76 | 2.90 (2.03–4.14)  1.59 (1.32–1.91)  0.84 (0.67–1.06) | <0.001<0.001  0.14 |
| Percent of assigned ICU teams pharmacist rounded with§ | 1.10 (0.87–1.38) | 0.42 | 1.47 (1.16–1.85) | 0.001 |
| Percent days, only attending physician⁂ | 1.00 (1.00–1.00) | 0.08 | 1.00 (1.00–1.00) | 0.94 |

CI: confidence interval, CMM: comprehensive medication management including pharmacist participation on interdisciplinary rounds, CMS: Center for Medicare & Medicaid Services, ICU: intensive care unit, MRC-ICU: medication regimen complexity- intensive care unit, OR: odds ratio, SOFA: sequential organ failure assessment, VA: Veterans Affairs

*worst score during the first 24 hours of ICU stay

**†**Inclusion criteria specified ≥18 years of age; however, some institutions allowed patients ≥18 years of age in the pediatric ICU

**‡**CMM delivered on interprofessional rounds: the pharmacist attended multidisciplinary rounds and verbally provided CMM (including review of medications and recommendations); CMM delivered not on interprofessional rounds: the pharmacist provided by CMM (reviewed medications and provided recommendations) but did not attend interdisciplinary rounds; Abbreviated CMM delivered: the pharmacist provided abbreviated CMM which includes brief medication review but does not include full patient review (e.g. progress notes, results) and recommendations were provided outside of rounds; CMM not delivered (absence of CMM) (absence of CMM): the patient received no medication review from a pharmacist outside of pharmacokinetic monitoring and prospective medication order verification

**§**Percent of assigned ICU teams pharmacist rounded is calculated by total number of ICU medical teams the pharmacist attended and provided CMM on interprofessional rounds for divided by total number of ICU medical teams providing care for patients that the pharmacist was assigned to provide CMM for

**⁂**Only attending physician refers to composition of the primary medical team. On these days, the patient received care from only the attending physician and did not receive care from any medical residents, fellows, or advanced practice providers.

#### eTable 10. Variance Inflation Factors (VIF) for Predictors Included in the Multivariable Model

| **Variable** | **VIF** |
| --- | --- |
| SOFA Score | 1.49 |
| MRC-ICU Score | 1.50 |
| Pharmacist-to-patient ratio (ICU patients) | 1.38 |
| Percent of assigned ICU teams pharmacist rounded with | 1.36 |
| Percent days, no pharmacist or pharmacist trainee | 1.53 |
| Percent days, only attending physician | 1.05 |
| Age | 1.04 |
| Absence of CMM for at least 1 day of ICU stay | 1.23 |

CMM: comprehensive medication management, ICU: intensive care unit, MRC-ICU: medication regimen complexity- intensive care unit, SOFA: sequential organ failure assessment

#### eTable 11. Propensity Score Matching Results Without Replacement

|  | Adjusted OR (95% CI) | E-value |
| --- | --- | --- |
| **Pharmacist-to-patient ratio of 1:15.1 to 1:46 compared to 1:7 to 1:15** | 1.10 (1.00, 1.22) | 1.43 |
| **CMM not delivered every day of ICU stay** | 1.28 (1.09, 1.51) | 1.88 |

CI: confidence interval, CMM: comprehensive medication management, ICU: intensive care unit, NNT: number needed to treat, OR: odds ratio

Row 1: matching based on pharmacist-to-patient ratio

Row 2: matching based on CMM every day of ICU stay

### Supplemental Figures

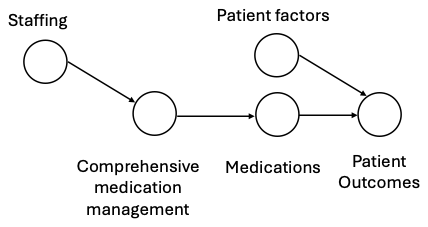

#### eFigure 2. Directed Acyclic Graph

The depicted directed acyclic graph (DAG) shows the causal relationship of ICU workforce staffing having a direct impact on comprehensive medication management (CMM) quality, which affects medications given to the patients, and medications (along with patient factors) have a direct causal effect on patient outcomes (due to both beneficial treatment effect and adverse drug effects).

Sikora, Andrea; Min, Wanyi; Devlin, John W.; Hu, Mengxuan; Murphy, David J.; Murray, Brian; Zhao, Bokai; Shen, Ye; Chen, Xianyan; Smith, Susan E.; Rowe, Sandra; Liu, Tianming; Li, Sheng. Medication Regimen Complexity-ICU (MRC-ICU) Investigator Team. Effect of Comprehensive Medication Management on Mortality in Critically Ill Patients. Critical Care Medicine 53(10):p e1995-e2004, October 2025. | DOI: 10.1097/CCM.0000000000006802

#### eFigure 3. Missingness

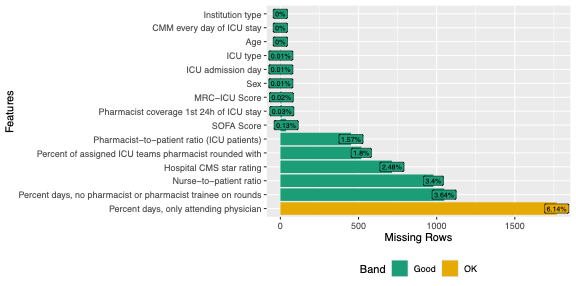

CMM: comprehensive medication management, CMS: Center for Medicare & Medicaid Services, ICU: intensive care unit, MRC-ICU: medication regimen complexity- intensive care unit, SOFA: sequential organ failure assessment

#### eFigure 4. Correlation Matrix of Predictors Included in the Mortality Model

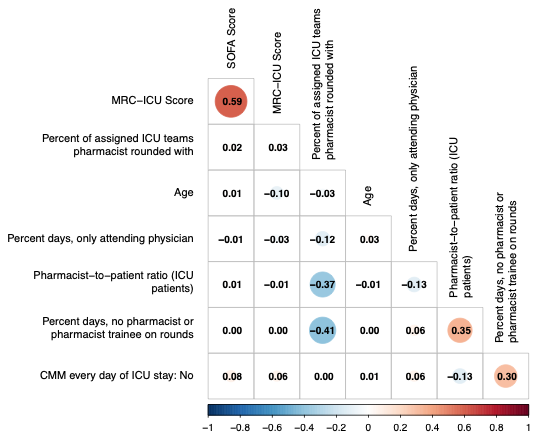

CMM: comprehensive medication management, ICU: intensive care unit, MRC-ICU: medication regimen complexity- intensive care unit, SOFA: sequential organ failure assessment

#### eFigure 5. Cumulative Incidence Function (CIF) for Probability of Discharge Alive

This plot depicts the likelihood of discharge alive for varying ICU lengths of stay. The left plot shows impact of having a critical care pharmacist every day of ICU stay, and the right plot shows varying pharmacist-to-patient ratios.

CMM: comprehensive medication management, ICU: intensive care unit, LOS: length of stay

CIF curves were estimated using a typical patient profile constructed from the imputed dataset, with covariates fixed at representative values:

Hospital type: Academic Medical Center

Hospital CMS star rating: 3

SOFA Score: 4

MRC-ICU Score: 10

ICU pharmacist coverage 1^st^ 24 hours: CMM delivered on interprofessional rounds

ICU type: Medical

Nurse-to-patient ratio: 2

Pharmacist-to-patient ratio (ICU patients): 17

ICU admission day of the week: Monday

Percent of assigned ICU teams pharmacist rounded with: 50

Percent days, no pharmacist or pharmacist trainee: 28.57143

Percent days, only attending physician: 0

Age: 63.82

Sex: Male

The only variable varied was CMM every day of ICU stay (left plot) and pharmacist-to-patient ratio (right plot)

CMM: comprehensive medication management, CMS: Center for Medicare & Medicaid Services, ICU: intensive care unit, MRC-ICU: medication regimen complexity-intensive care unit score, SOFA: sequential organ failure assessment

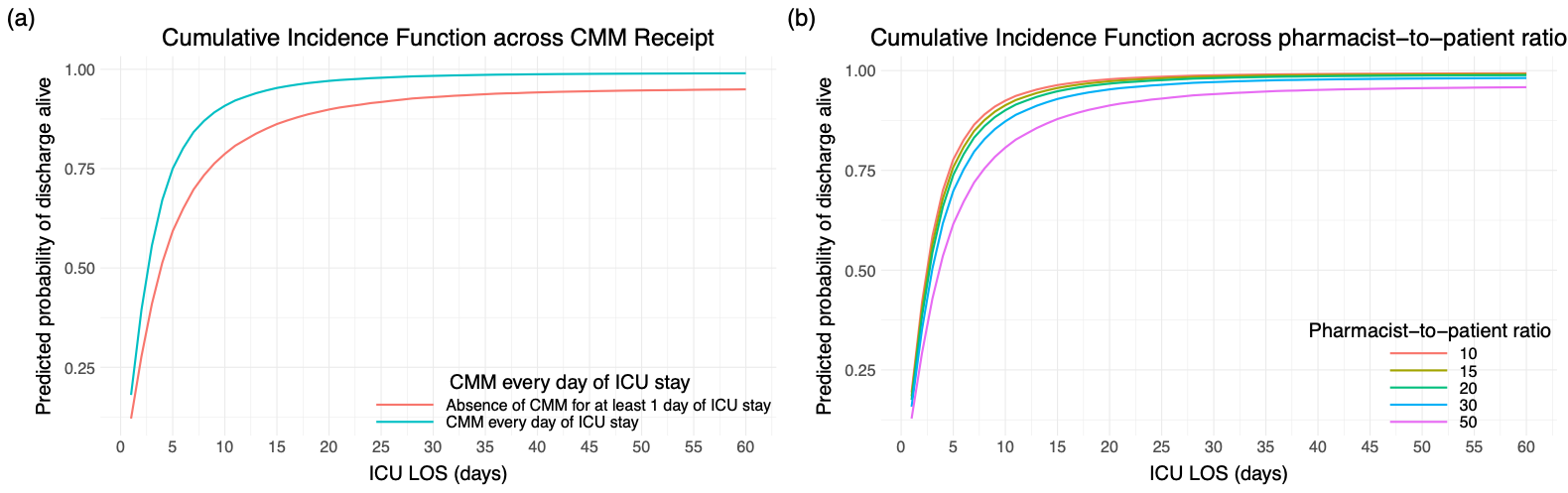

#### eFigure 6. Cumulative Incidence Function (CIF) for Probability of Discharge Alive across Pharmacist-to-patient Ratio (ICU patients) as Ranges and Displaying ICU Length of Stay

CIF curves were estimated using a typical patient profile constructed from the imputed dataset, with covariates fixed at representative values:

Hospital type: Academic Medical Center

Hospital CMS star rating: 3

SOFA Score: 4

MRC-ICU Score: 10

ICU pharmacist coverage 1^st^ 24 hours: CMM delivered on interprofessional rounds

ICU type: Medical

Nurse-to-patient ratio: 2

Pharmacist-to-patient ratio (ICU patients): 17

ICU admission day of the week: Monday

Percent of assigned ICU teams pharmacist rounded with: 50

Percent days, no pharmacist or pharmacist trainee: 28.57143

Percent days, only attending physician: 0

Age: 63.82

Sex: Male

The only variable varied was pharmacist-to-patient ratio

CMM: comprehensive medication management, CMS: Center for Medicare & Medicaid Services, ICU: intensive care unit, MRC-ICU: medication regimen complexity-intensive care unit score, SOFA: sequential organ failure assessment

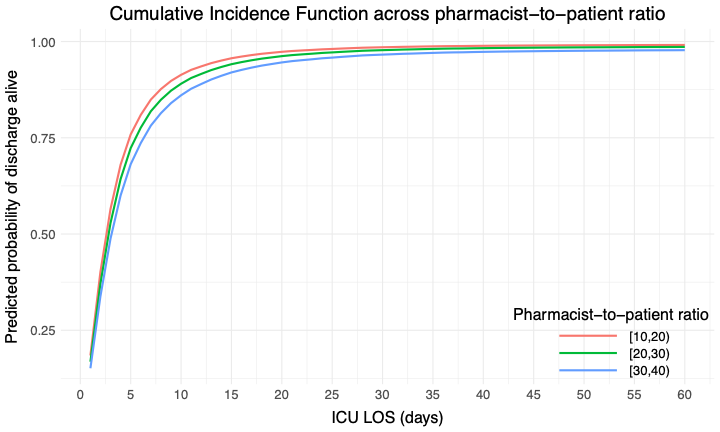

#### eFigure 7. Cumulative Incidence Function (CIF) for Probability of Discharge Alive across Pharmacist-to-patient Ratio (ICU patients) as Ranges at Number Needed to Treat Cutoff and Displaying ICU Length of Stay

CIF curves were estimated using a typical patient profile constructed from the imputed dataset, with covariates fixed at representative values:

Hospital type: Academic Medical Center

Hospital CMS star rating: 3

SOFA Score: 4

MRC-ICU Score: 10

ICU pharmacist coverage 1^st^ 24 hours: CMM delivered on interprofessional rounds

ICU type: Medical

Nurse-to-patient ratio: 2

Pharmacist-to-patient ratio (ICU patients): 17

ICU admission day of the week: Monday

Percent of assigned ICU teams pharmacist rounded with: 50

Percent days, no pharmacist or pharmacist trainee: 28.57143

Percent days, only attending physician: 0

Age: 63.82

Sex: Male

The only variable varied was pharmacist-to-patient ratio

CMM: comprehensive medication management, CMS: Center for Medicare & Medicaid Services, ICU: intensive care unit, MRC-ICU: medication regimen complexity-intensive care unit score, SOFA: sequential organ failure assessment

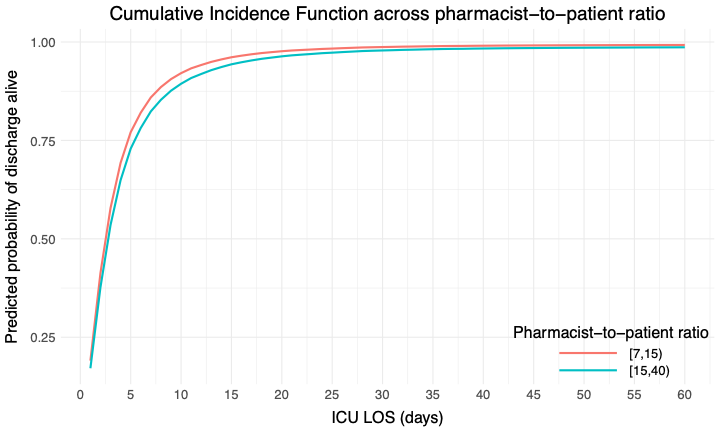

#### eFigure 8. Cumulative Incidence Function (CIF) for Probability of Discharge Alive across Pharmacist-to-patient Ratio (ICU patients) as Ranges Using 1:20 as the Cutoff and Displaying ICU Length of Stay

CIF curves were estimated using a typical patient profile constructed from the imputed dataset, with covariates fixed at representative values:

Hospital type: Academic Medical Center

Hospital CMS star rating: 3

SOFA Score: 4

MRC-ICU Score: 10

ICU pharmacist coverage 1^st^ 24 hours: CMM delivered on interprofessional rounds

ICU type: Medical

Nurse-to-patient ratio: 2

Pharmacist-to-patient ratio (ICU patients): 17

ICU admission day of the week: Monday

Percent of assigned ICU teams pharmacist rounded with: 50

Percent days, no pharmacist or pharmacist trainee: 28.57143

Percent days, only attending physician: 0

Age: 63.82

Sex: Male

The only variable varied was pharmacist-to-patient ratio

CMM: comprehensive medication management, CMS: Center for Medicare & Medicaid Services, ICU: intensive care unit, MRC-ICU: medication regimen complexity-intensive care unit score, SOFA: sequential organ failure assessment

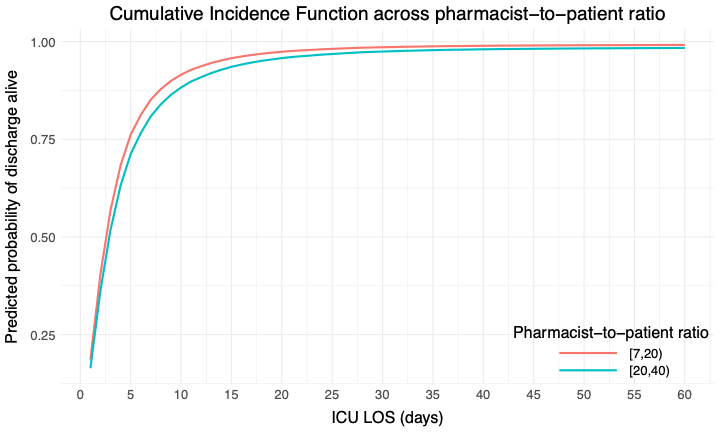

#### eFigure 9. Cumulative Incidence Function (CIF) for Probability of Discharge Alive across Critical Care Pharmacist Coverage Every Day of ICU Admission and Displaying Hospital Length of Stay

CIF curves were estimated using a typical patient profile constructed from the imputed dataset, with covariates fixed at representative values:

Hospital type: Academic Medical Center

Hospital CMS star rating: 3

SOFA Score: 4

MRC-ICU Score: 10

ICU pharmacist coverage 1^st^ 24 hours: CMM delivered on interprofessional rounds

ICU type: Medical

Nurse-to-patient ratio: 2

Pharmacist-to-patient ratio (ICU patients): 17

ICU admission day of the week: Monday

Percent of assigned ICU teams pharmacist rounded with: 50

Percent days, no pharmacist or pharmacist trainee: 28.57143

Percent days, only attending physician: 0

Age: 63.82

Sex: Male

The only variable varied was patient receiving CMM every day of ICU stay

CMM: comprehensive medication management, CMS: Center for Medicare & Medicaid Services, ICU: intensive care unit, MRC-ICU: medication regimen complexity-intensive care unit score, SOFA: sequential organ failure assessment

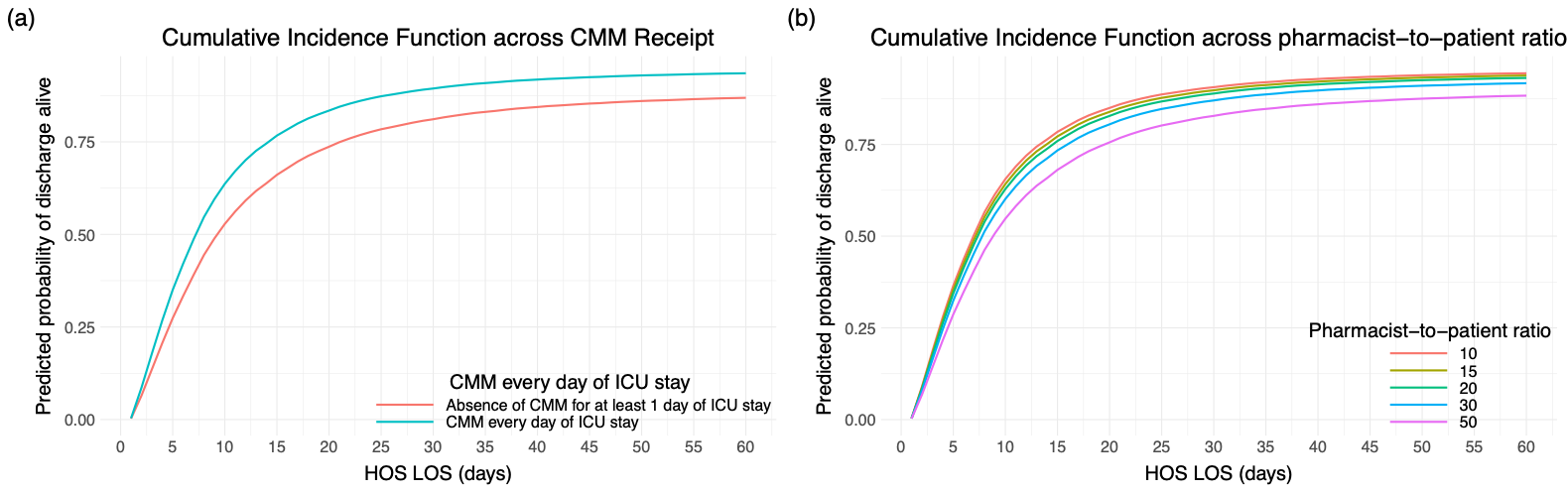

#### eFigure 10. Cumulative Incidence Function (CIF) for Probability of Discharge Alive across Pharmacist-to-patient Ratio (ICU patients) as Points and Displaying Hospital Length of Stay

CIF curves were estimated using a typical patient profile constructed from the imputed dataset, with covariates fixed at representative values:

Hospital type: Academic Medical Center

Hospital CMS star rating: 3

SOFA Score: 4

MRC-ICU Score: 10

ICU pharmacist coverage 1^st^ 24 hours: CMM delivered on interprofessional rounds

ICU type: Medical

Nurse-to-patient ratio: 2

Pharmacist-to-patient ratio (ICU patients): 17

ICU admission day of the week: Monday

Percent of assigned ICU teams pharmacist rounded with: 50

Percent days, no pharmacist or pharmacist trainee: 28.57143

Percent days, only attending physician: 0

Age: 63.82

Sex: Male

The only variable varied was pharmacist-to-patient ratio

CMM: comprehensive medication management, CMS: Center for Medicare & Medicaid Services, ICU: intensive care unit, MRC-ICU: medication regimen complexity-intensive care unit score, SOFA: sequential organ failure assessment

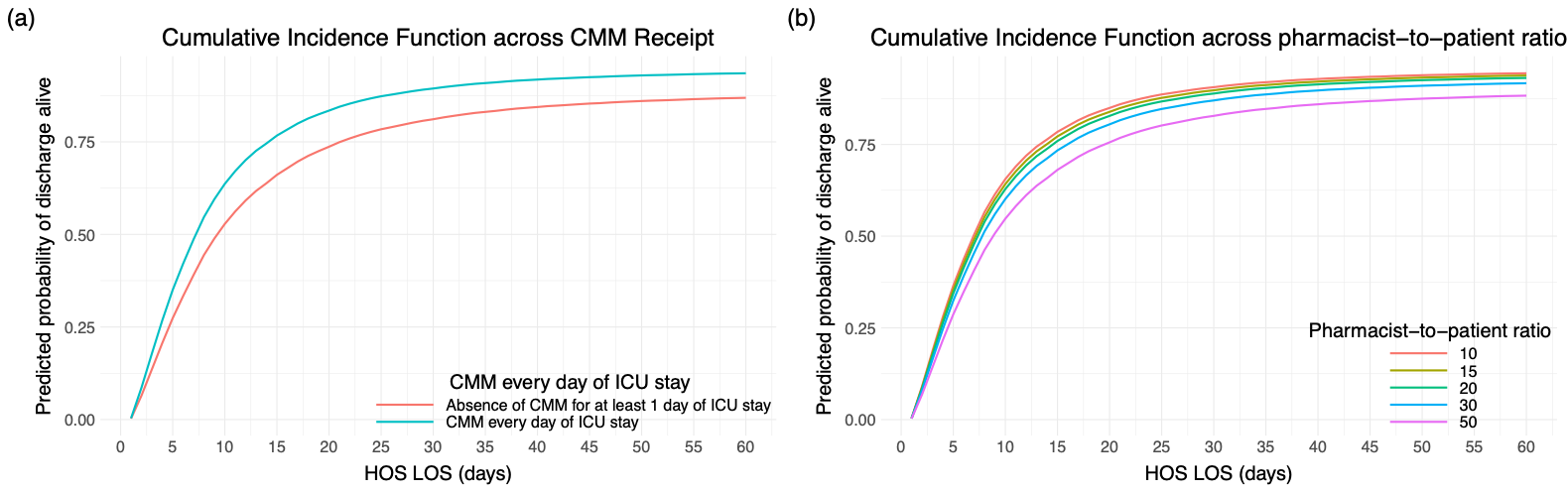

#### eFigure 11. Cumulative Incidence Function (CIF) for Probability of Discharge Alive across Pharmacist-to-patient Ratio (ICU patients) as Ranges and Displaying Hospital Length of Stay

CIF curves were estimated using a typical patient profile constructed from the imputed dataset, with covariates fixed at representative values:

Hospital type: Academic Medical Center

Hospital CMS star rating: 3

SOFA Score: 4

MRC-ICU Score: 10

ICU pharmacist coverage 1^st^ 24 hours: CMM delivered on interprofessional rounds

ICU type: Medical

Nurse-to-patient ratio: 2

Pharmacist-to-patient ratio (ICU patients): 17

ICU admission day of the week: Monday

Percent of assigned ICU teams pharmacist rounded with: 50

Percent days, no pharmacist or pharmacist trainee: 28.57143

Percent days, only attending physician: 0

Age: 63.82

Sex: Male

The only variable varied was pharmacist-to-patient ratio

CMM: comprehensive medication management, CMS: Center for Medicare & Medicaid Services, ICU: intensive care unit, MRC-ICU: medication regimen complexity-intensive care unit score, SOFA: sequential organ failure assessment

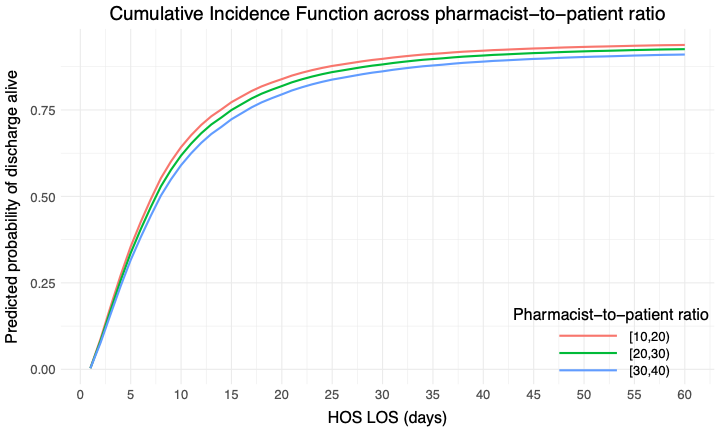

#### eFigure 12. Cumulative Incidence Function (CIF) for Probability of Discharge Alive across Pharmacist-to-patient Ratio (ICU patients) as Ranges at Number Needed to Treat Cutoff and Displaying Hospital Length of Stay

CIF curves were estimated using a typical patient profile constructed from the imputed dataset, with covariates fixed at representative values:

Hospital type: Academic Medical Center

Hospital CMS star rating: 3

SOFA Score: 4

MRC-ICU Score: 10

ICU pharmacist coverage 1^st^ 24 hours: CMM delivered on interprofessional rounds

ICU type: Medical

Nurse-to-patient ratio: 2

Pharmacist-to-patient ratio (ICU patients): 17

ICU admission day of the week: Monday

Percent of assigned ICU teams pharmacist rounded with: 50

Percent days, no pharmacist or pharmacist trainee: 28.57143

Percent days, only attending physician: 0

Age: 63.82

Sex: Male

The only variable varied was pharmacist-to-patient ratio

CMM: comprehensive medication management, CMS: Center for Medicare & Medicaid Services, ICU: intensive care unit, MRC-ICU: medication regimen complexity-intensive care unit score, SOFA: sequential organ failure assessment

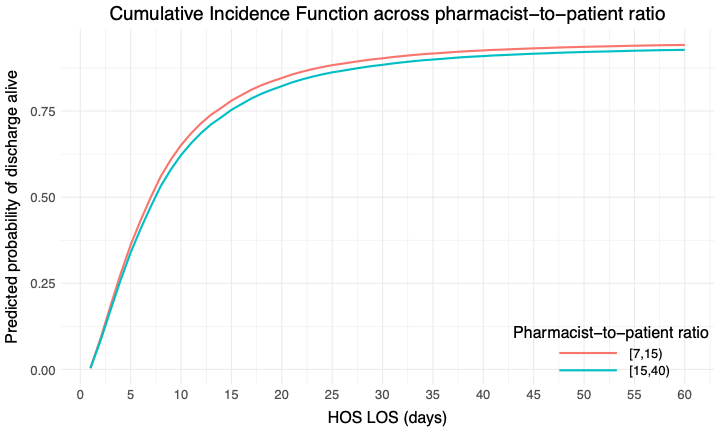

#### eFigure 13. Cumulative Incidence Function (CIF) for Probability of Discharge Alive across Pharmacist-to-patient Ratio (ICU patients) as Ranges Using 1:20 as the Cutoff and Displaying Hospital Length of Stay

CIF curves were estimated using a typical patient profile constructed from the imputed dataset, with covariates fixed at representative values:

Hospital type: Academic Medical Center

Hospital CMS star rating: 3

SOFA Score: 4

MRC-ICU Score: 10

ICU pharmacist coverage 1^st^ 24 hours: CMM delivered on interprofessional rounds

ICU type: Medical

Nurse-to-patient ratio: 2

Pharmacist-to-patient ratio (ICU patients): 17

ICU admission day of the week: Monday

Percent of assigned ICU teams pharmacist rounded with: 50

Percent days, no pharmacist or pharmacist trainee: 28.57143

Percent days, only attending physician: 0

Age: 63.82

Sex: Male

The only variable varied was pharmacist-to-patient ratio

CMM: comprehensive medication management, CMS: Center for Medicare & Medicaid Services, ICU: intensive care unit, MRC-ICU: medication regimen complexity-intensive care unit score, SOFA: sequential organ failure assessment

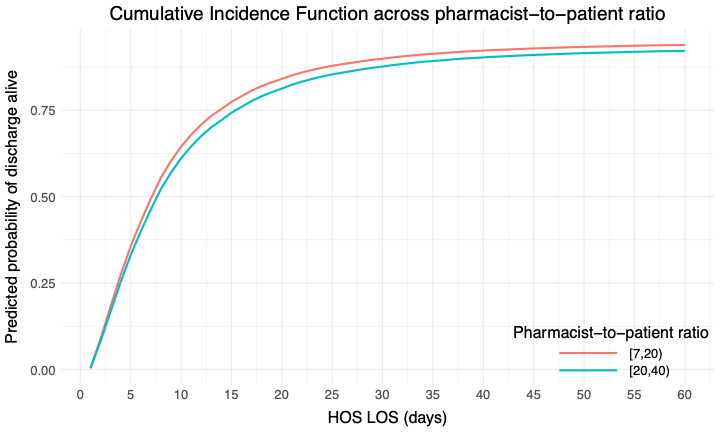

#### eFigure 14. 28-Day Cumulative Incidence Function (CIF) for Probability of Extubation Alive across CMM

CIF curves were estimated using a typical patient profile constructed from the imputed dataset, with covariates fixed at representative values:

Hospital type: Academic Medical Center

Hospital CMS star rating: 3

SOFA Score: 4

MRC-ICU Score: 10

ICU pharmacist coverage 1^st^ 24 hours: CMM delivered on interprofessional rounds

ICU type: Medical

Nurse-to-patient ratio: 2

Pharmacist-to-patient ratio (ICU patients): 17

ICU admission day of the week: Monday

Percent of assigned ICU teams pharmacist rounded with: 50

Percent days, no pharmacist or pharmacist trainee: 28.57143

Percent days, only attending physician: 0

Age: 63.82

Sex: Male

The only variable varied was patient receiving CMM every day of ICU stay

CMM: comprehensive medication management, CMS: Center for Medicare & Medicaid Services, ICU: intensive care unit, MRC-ICU: medication regimen complexity-intensive care unit score, SOFA: sequential organ failure assessment

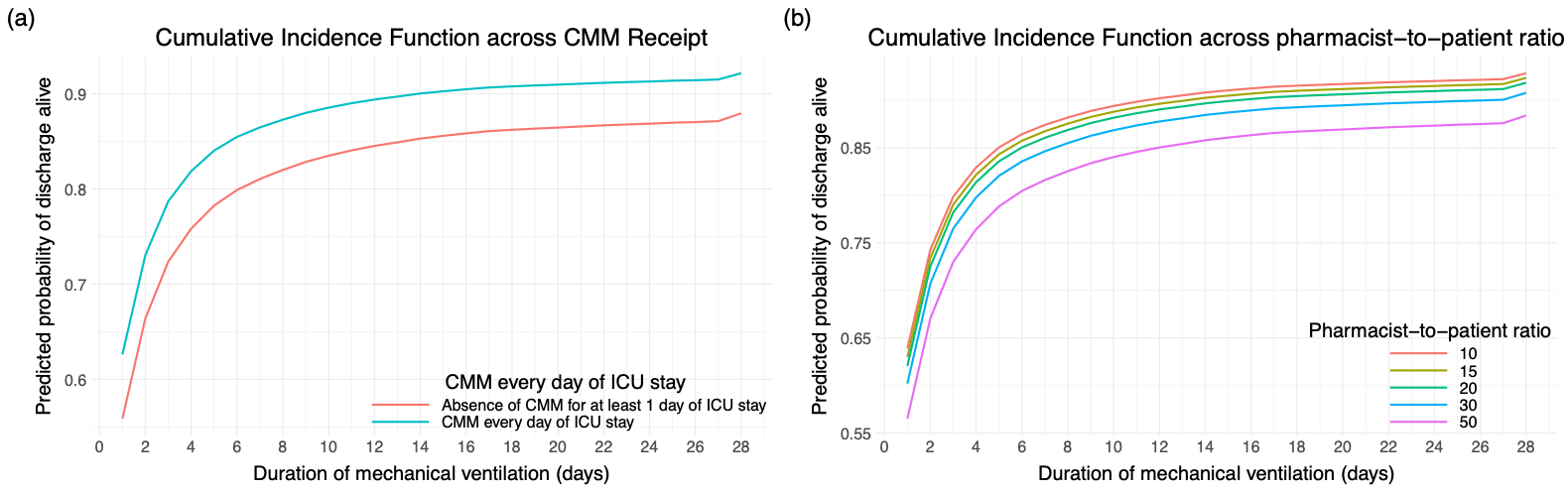

#### eFigure 15. 28-Day Cumulative Incidence Function (CIF) for Probability of Extubation Alive across Pharmacist-to-patient Ratio (ICU patients) as Points

CIF curves were estimated using a typical patient profile constructed from the imputed dataset, with covariates fixed at representative values:

Hospital type: Academic Medical Center

Hospital CMS star rating: 3

SOFA Score: 4

MRC-ICU Score: 10

ICU pharmacist coverage 1^st^ 24 hours: CMM delivered on interprofessional rounds

ICU type: Medical

Nurse-to-patient ratio: 2

Pharmacist-to-patient ratio (ICU patients): 17

ICU admission day of the week: Monday

Percent of assigned ICU teams pharmacist rounded with: 50

Percent days, no pharmacist or pharmacist trainee: 28.57143

Percent days, only attending physician: 0

Age: 63.82

Sex: Male

The only variable varied was pharmacist-to-patient ratio

CMM: comprehensive medication management, CMS: Center for Medicare & Medicaid Services, ICU: intensive care unit, MRC-ICU: medication regimen complexity-intensive care unit score, SOFA: sequential organ failure assessment

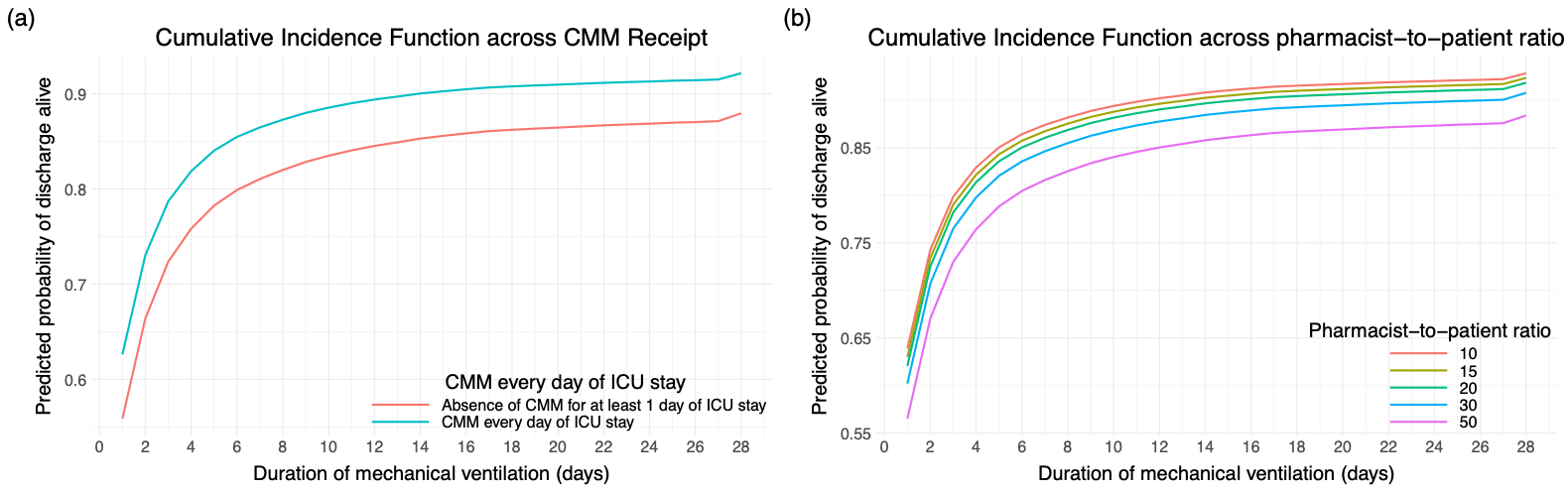

#### eFigure 16. 28-Day Cumulative Incidence Function for (CIF) Probability of Extubation Alive across Pharmacist-to-patient Ratio (ICU patients) as Ranges

CIF curves were estimated using a typical patient profile constructed from the imputed dataset, with covariates fixed at representative values:

Hospital type: Academic Medical Center

Hospital CMS star rating: 3

SOFA Score: 4

MRC-ICU Score: 10

ICU pharmacist coverage 1^st^ 24 hours: CMM delivered on interprofessional rounds

ICU type: Medical

Nurse-to-patient ratio: 2

Pharmacist-to-patient ratio (ICU patients): 17

ICU admission day of the week: Monday

Percent of assigned ICU teams pharmacist rounded with: 50

Percent days, no pharmacist or pharmacist trainee: 28.57143

Percent days, only attending physician: 0

Age: 63.82

Sex: Male

The only variable varied was pharmacist-to-patient ratio

CMM: comprehensive medication management, CMS: Center for Medicare & Medicaid Services, ICU: intensive care unit, MRC-ICU: medication regimen complexity-intensive care unit score, SOFA: sequential organ failure assessment

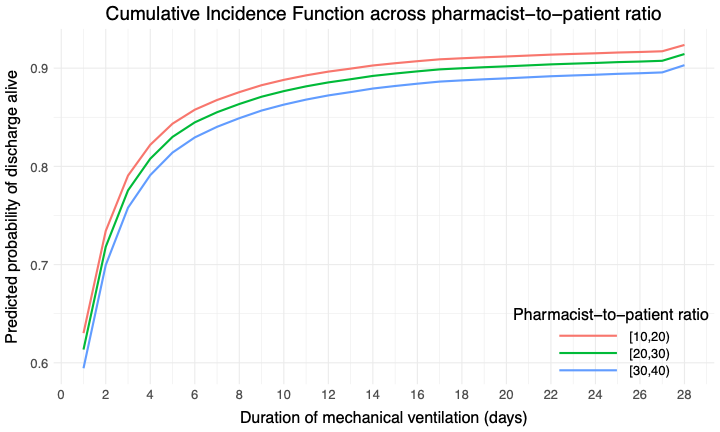

#### eFigure 17. 28-Day Cumulative Incidence Function (CIF) for Probability of Extubation Alive across Pharmacist-to-patient Ratio (ICU patients) as Ranges at Number Needed to Treat Cutoff

CIF curves were estimated using a typical patient profile constructed from the imputed dataset, with covariates fixed at representative values:

Hospital type: Academic Medical Center

Hospital CMS star rating: 3

SOFA Score: 4

MRC-ICU Score: 10

ICU pharmacist coverage 1^st^ 24 hours: CMM delivered on interprofessional rounds

ICU type: Medical

Nurse-to-patient ratio: 2

Pharmacist-to-patient ratio (ICU patients): 17

ICU admission day of the week: Monday

Percent of assigned ICU teams pharmacist rounded with: 50

Percent days, no pharmacist or pharmacist trainee: 28.57143

Percent days, only attending physician: 0

Age: 63.82

Sex: Male

The only variable varied was pharmacist-to-patient ratio

CMM: comprehensive medication management, CMS: Center for Medicare & Medicaid Services, ICU: intensive care unit, MRC-ICU: medication regimen complexity-intensive care unit score, SOFA: sequential organ failure assessment

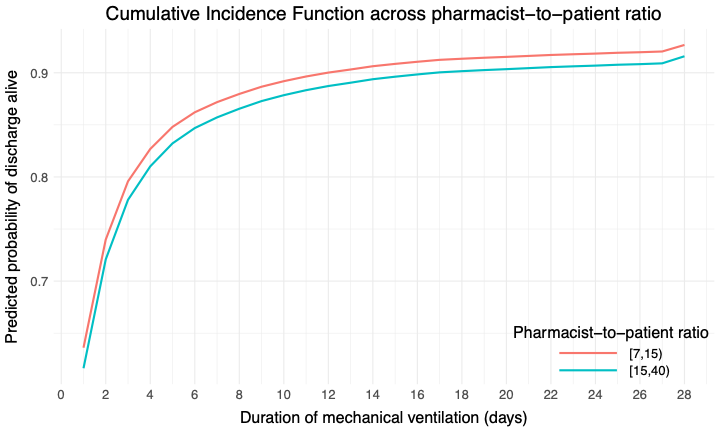

#### eFigure 18. 28-Day Cumulative Incidence Function (CIF) for Probability of Extubation Alive across Pharmacist-to-patient Ratio (ICU patients) as Ranges Using 1:20 as the Cutoff

CIF curves were estimated using a typical patient profile constructed from the imputed dataset, with covariates fixed at representative values:

Hospital type: Academic Medical Center

Hospital CMS star rating: 3

SOFA Score: 4

MRC-ICU Score: 10

ICU pharmacist coverage 1^st^ 24 hours: CMM delivered on interprofessional rounds

ICU type: Medical

Nurse-to-patient ratio: 2

Pharmacist-to-patient ratio (ICU patients): 17

ICU admission day of the week: Monday

Percent of assigned ICU teams pharmacist rounded with: 50

Percent days, no pharmacist or pharmacist trainee: 28.57143

Percent days, only attending physician: 0

Age: 63.82

Sex: Male

The only variable varied was pharmacist-to-patient ratio

CMM: comprehensive medication management, CMS: Center for Medicare & Medicaid Services, ICU: intensive care unit, MRC-ICU: medication regimen complexity-intensive care unit score, SOFA: sequential organ failure assessment

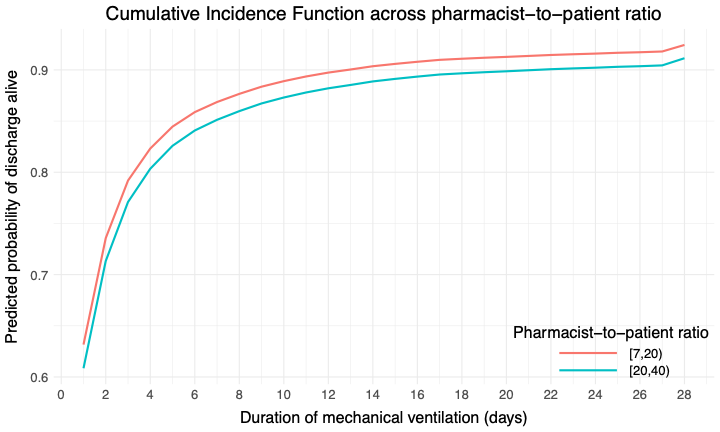

#### eFigure 19. Covariate Balance Before and After Propensity Score Matching by Pharmacist-to-Patient Ratio Without Replacement

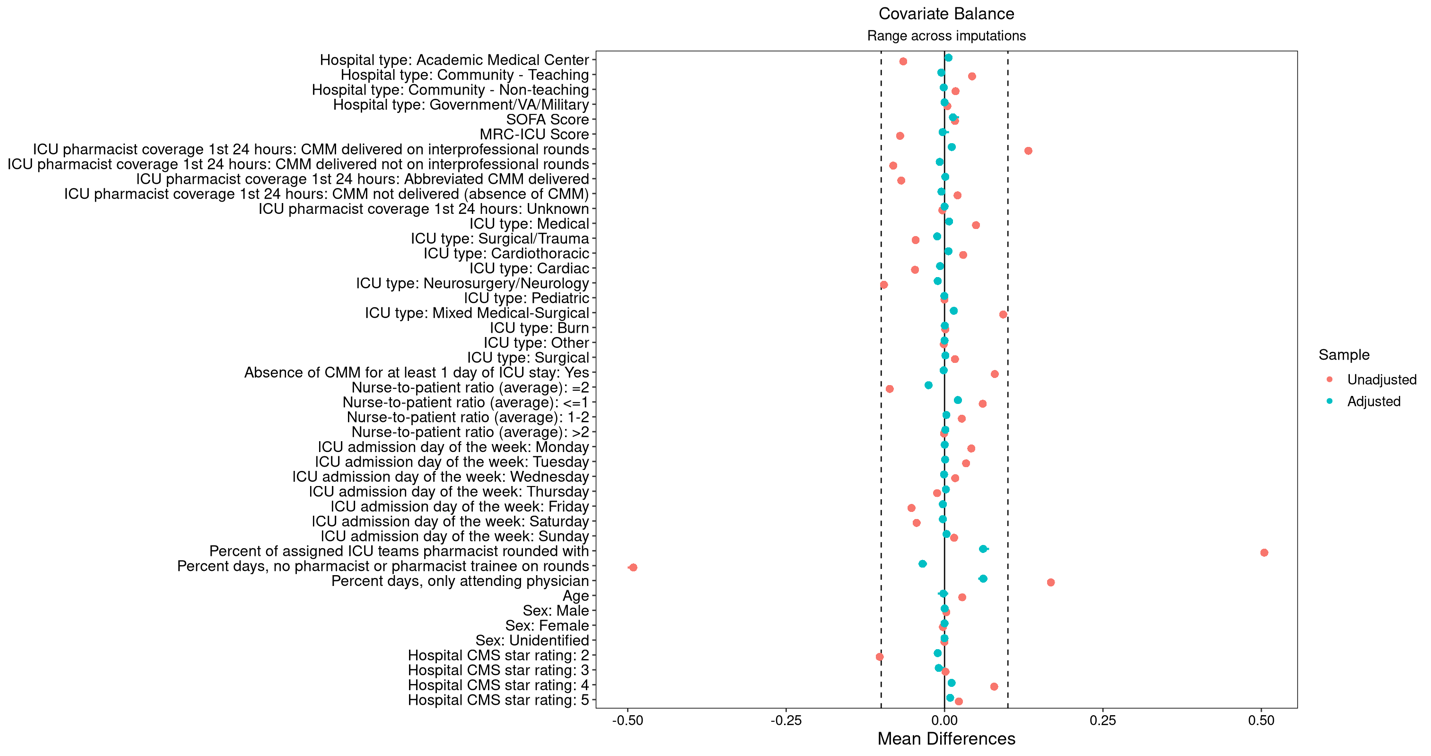

CMM: comprehensive medication management, CMS: Center for Medicare & Medicaid Services, ICU: intensive care unit, MRC-ICU: medication regiment complexity- intensive care unit, SOFA: sequential organ failure assessment

#### eFigure 20. Covariate Balance Before and After Propensity Score Matching by CMM Every Day of ICU Stay Without Replacement

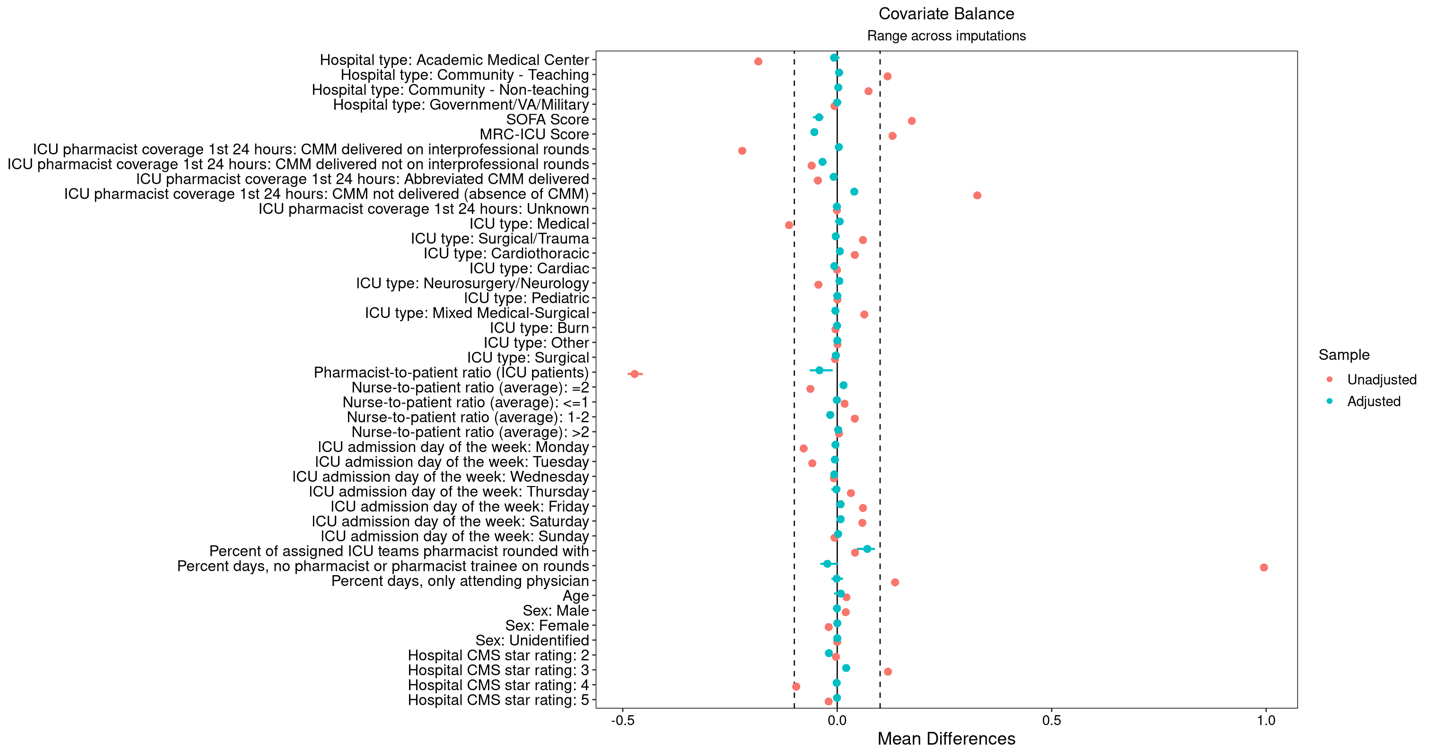

CMM: comprehensive medication management, CMS: Center for Medicare & Medicaid Services, ICU: intensive care unit, MRC-ICU: medication regiment complexity- intensive care unit, SOFA: sequential organ failure assessment

### Sensitivity Analysis- No Patients with Imputed Data

#### eTable 12. Baseline Characteristics for non-Imputed cohort (N=24420)

|  | Overall  (24,420) | Deceased  (3,387) | Alive  (21,033) | P value |
| --- | --- | --- | --- | --- |
| *Primary Variable* | | | | |
| Pharmacist-to-patient ratio (ICU patients) | 19.6 (10.2) | 17.0 (6.7) | 20.3 (10.8) | <0.001 |
| *Secondary Variable* | | | | |
| Absence of CMM for at least 1 day of ICU stay | 5,817 (23.8) | 958 (28.3) | 4,859 (23.1) | <0.001 |
| *Co-variates* | | | | |
| Age, years | 61.4 (16.9) | 66.0 (15.4) | 60.7 (17.0) | <0.001 |
| Sex, female | 10,440 (42.8) | 1,451 (42.8) | 8,989 (42.7) | >0.90 |
| SOFA Score* | 5.2 (4.1) | 8.8 (4.4) | 4.6 (3.7) | <0.001 |
| MRC-ICU Score* | 11.3 (6.6) | 14.6 (7.0) | 10.7 (6.4) | <0.001 |
| ICU admission day of the week  Monday  Tuesday  Wednesday  Thursday  Friday  Saturday  Sunday | 4,072 (16.7)  3,996 (16.4)  3,744 (15.3)  3,732 (15.3)  3,083 (12.6)  2,738 (11.2)  3,055 (12.5) | 537 (15.9)  505 (14.9)  495 (14.6)  492 (14.5)  437 (12.9)  451 (13.3)  470 (13.9) | 3,535 (16.8)  3,491 (16.6)  3,249 (15.4)  3,240 (15.4)  2,646 (12.6)  2,287 (10.9)  2,585 (12.3) | <0.001 |
| Dialysis | 2,775 (11.4) | 894 (26.4) | 1,881 (8.9) | <0.001 |
| ECMO | 230 (0.9) | 97 (2.9) | 133 (0.6) | <0.001 |
| Mechanical circulatory support | 11,571 (47.4) | 2,639 (78.0) | 8,932 (42.5) | <0.001 |
| Mechanical ventilation | 11,783 (48.3) | 2,563 (75.7) | 9,220 (43.8) | <0.001 |
| ICU type†  Medical  Surgical/Trauma  Surgical  Cardiothoracic  Cardiac  Neurosurgery/Neurology  Pediatric  Mixed Medical-Surgical  Burn  Other | 7,984 (32.7)  3,044 (12.5)  1,971 (8.1)  2,355 (9.6)  1,984 (8.1)  3,499 (14.3)  9 (0)  3,388 (13.9)  160 (0.7)  26 (0.1) | 1,492 (44.1)  326 (9.6)  198 (5.8)  160 (4.7)  290 (8.6)  408 (12.0)  0 (0)  501 (14.8)  10 (0.3)  2 (0.1) | 6,492 (30.9)  2,718 (12.9)  1,773 (8.4)  2,195 (10.4)  1,694 (8.1)  3,091 (14.7)  9 (0)  2,881 (13.7)  150 (0.7)  24 (0.1) | <0.001 |
| Hospital type  Academic Medical Center  Community - Teaching  Community - Non-teaching  Government/VA/Military | 15,915 (65.2)  6,172 (25.3)  2,188 (9.0)  145 (0.6) | 2,217 (65.5)  881 (26.0)  274 (8.1)  15 (0.4) | 13,698 (65.1)  5,291 (25.2)  1,914 (9.1)  130 (0.6) | 0.13 |
| Hospital CMS star rating  2  3  4  5 | 5,767 (23.6)  13,118 (53.7)  5,123 (21.0)  412 (1.7) | 778 (23.0)  1,835 (54.2)  724 (21.4)  50 (1.5) | 4,989 (23.7)  11,283 (53.6)  4,399 (20.9)  362 (1.7) | 0.50 |
| Pharmacist-to-patient ratio (all patients) | 27.7 (22.9) | 27.2 (19.7) | 27.8 (23.4) | 0.11 |
| ICU pharmacist coverage 1^st^ 24 hours  CMM on interprofessional rounds  CMM delivered not on interprofessional rounds  Abbreviated CMM delivered only  CMM not delivered (absence of CMM)  Unknown | 16,860 (69.0)  3,401 (13.9)  1,742 (7.1)  2,298 (9.4)  119 (0.5) | 2,297 (67.8)  437 (12.9)  244 (7.2)  387 (11.4)  22 (0.6) | 14,563 (69.2)  2,964 (14.1)  1,498 (7.1)  1,911 (9.1)  97 (0.5) | <0.001 |
| Nurse-to-patient ratio, 1:X  ≤1  1-2  2  >2 | 1,148 (4.7)  2,365 (9.7)  20,273 (83.0)  634 (2.6) | 238 (7.0)  523 (5.4)  2,545 (75.1)  81 (2.4) | 910 (4.3)  1,842 (8.8)  17,728 (84.3)  553 (2.6) | <0.001 |
| Percent of assigned ICU teams pharmacist rounded with§ | 53.7 (32.6) | 54.9 (31.2) | 53.5 (32.9) | 0.011 |
| Percent days, no pharmacist or pharmacist trainee on rounds | 30.5 (33.1) | 30.5 (29.9) | 30.5 (33.6) | 0.006 |
| Percent days, only attending physician⁂ | 8.7 (26.3) | 8.2 (24.9) | 8.8 (26.5) | 0.60 |
| *Outcomes* | | | |  |
| Hospital LOS, days | 11.6 (15.6) | 13.1 (18.9) | 11.4 (15.0) | <0.001 |
| ICU LOS, days | 5.5 (8.6) | 8.1 (12.4) | 5.1 (7.7) | <0.001 |
| ICU free days | 20.1 (9.3) | 0.0 (0.0) | 23.3 (5.3) | <0.001 |
| Duration of mechanical ventilation, days | 2.2 (19.4) | 4.8 (7.1) | 1.8 (20.7) | <0.001 |
| VFDs | 22.8 (9.9) | 0.0 (0.0) | 26.5 (4.0) | <0.001 |

CMM: comprehensive medication management, CMS: Center for Medicare & Medicaid Services, ECMO: extracorporeal membrane support, ICU: intensive care unit, LOS: length of stay, MRC-ICU: medication regimen complexity- intensive care unit, SOFA: sequential organ failure assessment, VA: Veterans Affairs, VFDs: ventilator free days

*worst score during the first 24 hours of ICU stay

**†**Inclusion criteria specified ≥18 years of age; however, some institutions allowed patients ≥18 years of age in the pediatric ICU

**‡**CMM delivered on interprofessional rounds: the pharmacist attended multidisciplinary rounds and verbally provided CMM (including review of medications and recommendations); CMM delivered not on interprofessional rounds: the pharmacist provided by CMM (reviewed medications and provided recommendations) but did not attend interdisciplinary rounds; Abbreviated CMM delivered: the pharmacist provided abbreviated CMM which includes brief medication review but does not include full patient review (e.g. progress notes, results) and recommendations were provided outside of rounds; CMM not delivered (absence of CMM): the patient received no medication review from a pharmacist outside of pharmacokinetic monitoring and prospective medication order verification

**§**Percent of assigned ICU teams pharmacist rounded is calculated by total number of ICU medical teams the pharmacist attended and provided CMM on interprofessional rounds for divided by total number of ICU medical teams providing care for patients that the pharmacist was assigned to provide CMM for

**⁂**Only attending physician refers to composition of the primary medical team. On these days, the patient received care from only the attending physician and did not receive care from any medical residents, fellows, or advanced practice providers.

#### eTable 13. Hospital Mortality Regression for non-Imputed cohort (N=24420)

|  | Univariate OR (95% CI) | p-value | Multivariate adjusted OR (95% CI) | p-value |
| --- | --- | --- | --- | --- |
| *Primary Variable* | | | | |
| Pharmacist-to-patient ratio (ICU patients) | 1.00 (1.00–1.01) | 0.11 | 1.00 (1.00–1.01) | 0.05 |
| *Secondary Variables* |  |  |  |  |
| Absence of CMM for at least 1 day of ICU stay | 1.58 (1.30–1.92) | <0.001 | 1.20 (0.98–1.46) | 0.08 |
| *Co-variates* | | | | |
| Age, years | 1.02 (1.02–1.02) | <0.001 | 1.03 (1.02–1.03) | <0.001 |
| Sex, female | 0.99 (0.91–1.07) | 0.77 | 1.03 (0.93–1.14) | 0.55 |
| SOFA Score* | 1.26 (1.25–1.28) | <0.001 | 1.31 (1.27–1.36) | <0.001 |
| MRC-ICU Score* | 1.09 (1.08–1.10) | <0.001 | 1.04 (1.02–1.06) | <0.001 |
| SOFA-MRC-ICU interaction term | 1.01 (1.01–1.01) | <0.001 | 1.00 (1.00–1.00) | <0.001 |
| ICU admission day of the week (reference: Monday)  Tuesday  Wednesday  Thursday  Friday  Saturday  Sunday | 0.95 (0.83–1.09)  0.99 (0.85–1.15)  1.00 (0.87–1.15)  1.07 (0.94–1.21)  1.26 (1.08–1.47)  1.17 (1.02–1.33) | 0.49  0.91  0.96  0.30  0.003  0.02 | 0.97 (0.84–1.11)  0.94 (0.81–1.10)  0.97 (0.83–1.13)  0.93 (0.81–1.06)  1.04 (0.87–1.25)  1.07 (0.92–1.24) | 0.62  0.43  0.67  0.37  0.67  0.40 |
| ICU type† (reference: medical ICU)  Surgical/Trauma  Surgical  Cardiothoracic  Cardiac  Neurosurgery/Neurology  Pediatric  Mixed Medical-Surgical  Burn  Other | 0.57 (0.46–0.70)  0.48 (0.38–0.61)  0.34 (0.25–0.47)  0.74 (0.59–0.94)  0.52 (0.44–0.62)  0.00 (0.00–0.00)  0.77 (0.61–0.98)  0.35 (0.19–0.64)  0.23 (0.07–0.74) | <0.001  <0.001  <0.001  0.01  <0.001  <0.001  0.03  <0.001  0.01 | 0.79 (0.65–0.97)  0.45 (0.35–0.58)  0.22 (0.14–0.36)  0.91 (0.73–1.14)  1.16 (0.93–1.46)  0.00 (0.00–0.00)  0.80 (0.60–1.05)  0.64 (0.31–1.28)  0.57 (0.19–1.73) | 0.03  <0.001  <0.001  0.41  0.19  <0.001  0.10  0.21  0.32 |
| Hospital type (reference: academic medical center)  Community - Teaching  Community - Non-teaching  Government/VA/Military | 1.06 (0.84–1.33)  0.99 (0.70–1.39)  0.73 (0.64–0.84) | 0.64  0.94  <0.001 | 0.93 (0.72–1.19)  0.83 (0.60–1.14)  1.04 (0.78–1.37) | 0.56  0.25  0.80 |
| Hospital CMS star rating (reference: 3)  2  4  5 | 0.95 (0.75–1.21)  0.99 (0.76–1.30)  0.82 (0.43–1.58) | 0.70  0.96  0.56 | 1.10 (0.90–1.34)  0.87 (0.64–1.18)  0.50 (0.30–0.82) | 0.64  0.94  0.007 |
| ICU pharmacist coverage 1st 24 hours‡ (reference: CMM delivered on interprofessional rounds)  CMM delivered not on interprofessional rounds  Abbreviated CMM delivered  CMM not delivered (absence of CMM)  Unknown | 0.97 (0.85–1.10)  1.09 (0.96–1.23)  1.31 (1.12–1.52)  1.44 (0.69–2.99) | 0.63  0.19  <0.001  0.33 | 1.05 (0.91–1.21)  1.10 (0.91–1.33)  1.09 (0.88–1.36)  1.31 (0.65–2.63) | 0.50  0.32  0.42  0.45 |
| Nurse-to-patient ratio (average)(reference: 1:2)  ≤1  1-2  >2 | 3.63 (2.55–5.17)  2.40 (2.11–2.73)  0.96 (0.68–1.35) | <0.001  <0.001  0.79 | 2.73 (1.82–4.10)  1.55 (1.27–1.89)  0.87 (0.68–1.11) | <0.001  <0.001  0.27 |
| Percent of assigned ICU teams pharmacist rounded with§ | 1.01 (0.82–1.24) | 0.93 | 1.22 (0.95–1.57) | 0.11 |
| Percent days, no pharmacist or pharmacist trainee on rounds | 1.00 (1.00–1.00) | 0.57 | 1.00 (1.00–1.00) | 0.33 |
| Percent days, only attending physician⁂ | 1.00 (0.99–1.01) | 0.4 | 1.00 (0.99–1.01) | 0.47 |

CI: confidence interval, CMM: comprehensive medication management, CMS: Center for Medicare & Medicaid Services, ICU: intensive care unit, MRC-ICU: medication regimen complexity- intensive care unit, OR: odds ratio, SOFA: sequential organ failure assessment, VA: Veterans Affairs

*worst score during the first 24 hours of ICU stay

**†**Inclusion criteria specified ≥18 years of age; however, some institutions allowed patients ≥18 years of age in the pediatric ICU

**‡**CMM delivered on interprofessional rounds: the pharmacist attended multidisciplinary rounds and verbally provided CMM (including review of medications and recommendations); CMM delivered not on interprofessional rounds: the pharmacist provided by CMM (reviewed medications and provided recommendations) but did not attend interdisciplinary rounds; Abbreviated CMM delivered: the pharmacist provided abbreviated CMM which includes brief medication review but does not include full patient review (e.g. progress notes, results) and recommendations were provided outside of rounds; CMM not delivered (absence of CMM): the patient received no medication review from a pharmacist outside of pharmacokinetic monitoring and prospective medication order verification

**§**Percent of assigned ICU teams pharmacist rounded is calculated by total number of ICU medical teams the pharmacist attended and provided CMM on interprofessional rounds for divided by total number of ICU medical teams providing care for patients that the pharmacist was assigned to provide CMM for

**⁂**Only attending physician refers to composition of the primary medical team. On these days, the patient received care from only the attending physician and did not receive care from any medical residents, fellows, or advanced practice providers.

#### eTable 14. Hospital and Intensive Care Unit Length of Stay Regression for non-Imputed cohort

|  | Hospital Length of Stay (n=24416) | | ICU Length of Stay (n=24416) | |
| --- | --- | --- | --- | --- |
|  | Hazard Ratio (95% Confidence Interval) | p-value | Hazard Ratio (95% Confidence Interval) | p-value |
| *Primary Variable* | | |  |  |
| Pharmacist-to-patient ratio (ICU patients) | 0.99 (0.99, 1.00) | <0.001 | 0.99 (0.99, 0.99) | <0.001 |
| *Secondary Variable* | | | | |
| Absence of CMM for at least 1 day of ICU stay | 0.72 (0.66, 0.78) | <0.001 | 0.58 (0.53, 0.64) | <0.001 |
| *Co-variates* | | | | |
| Age, years | 0.99 (0.99, 0.99) | <0.001 | 1.00 (1.00, 1.00) | 0.01 |
| Sex, female | 0.99 (0.96, 1.03) | 0.74 | 1.01 (0.98, 1.04) | 0.41 |
| SOFA Score | 0.90 (0.89, 0.91) | <0.001 | 0.93 (0.92, 0.94) | <0.001 |
| MRC-ICU Score | 0.98 (0.97, 0.98) | <0.001 | 0.98 (0.97, 0.98) | <0.001 |
| ICU admission day of the week (reference: Monday)  Tuesday  Wednesday  Thursday  Friday  Saturday  Sunday | 1.05 (1.00–1.11)  1.03 (0.98–1.09)  1.04 (0.99–1.10)  0.96 (0.90–1.03)  0.97 (0.90–1.05)  0.96 (0.91–1.02) | 0.07  0.22  0.14  0.24  0.43  0.19 | 1.03 (0.99–1.08)  1.03 (0.99–1.08)  1.06 (1.01–1.11)  0.91 (0.85–0.97)  0.93 (0.86–1.00)  1.00 (0.95–1.05) | 0.18  0.17  0.01  0.006  0.05  0.94 |
| ICU type† (reference: medical ICU)  Surgical/Trauma  Surgical  Cardiothoracic  Cardiac  Neurosurgery/Neurology  Pediatric  Mixed Medical-Surgical  Burn  Other | 1.08 (0.90–1.28)  1.09 (0.92–1.28)  1.62 (1.33–1.97)  1.14 (0.97–1.33)  0.94 (0.82–1.07)  1.89 (1.54–2.32)  1.10 (0.95–1.27)  0.83 (0.56–1.22)  1.44 (0.78–2.66) | 0.41  0.32  <0.001  0.12  0.34  <0.001  0.22  0.34  0.24 | 0.98 (0.84–1.13)  1.13 (0.94–1.35)  1.22 (0.99–1.50)  0.98 (0.83–1.15)  0.81 (0.70–0.93)  1.45 (0.98–2.14)  1.27 (1.37–1.50)  0.59 (0.47–0.73)  1.30 (0.96–1.77) | 0.76  0.18  0.06  0.81  0.004  0.06  0.007  <0.001  0.09 |
| Hospital type (reference: academic medical center)  Community - Teaching  Community - Non-teaching  Government/VA/Military | 1.00 (0.88–1.13)  1.85 (1.56–2.20)  3.01 (2.29–3.97) | 1  <0.001  <0.001 | 1.16 (1.03–1.30)  0.79 (0.66–0.96)  0.73 (0.60–0.88) | 0.01  0.01  <0.001 |
| Hospital CMS star rating (reference: 3)  2  4  5 | 0.87 (0.78–0.97)  0.83 (0.74–0.93)  0.76 (0.62–0.94) | 0.01  0.002  0.009 | 1.08 (0.96–1.21)  1.14 (1.00–1.30)  1.14 (0.67–1.92) | 0.21  0.06  0.63 |
| ICU pharmacist coverage 1st 24 hours‡ (reference: CMM delivered on interprofessional rounds)  CMM delivered not on interprofessional rounds  Abbreviated CMM delivered  CMM not delivered (absence of CMM)  Unknown | 0.98 (0.93–1.04)  0.94 (0.87–1.03)  1.08 (1.00–1.17)  0.78 (0.53–1.13) | 0.59  0.19  0.07  0.19 | 1.01 (0.95–1.07)  0.98 (0.89–1.08)  1.09 (1.01–1.19)  0.61 (0.47–0.77) | 0.79  0.66  0.04  <0.001 |
| Nurse-to-patient ratio (average)(reference: 1:2)  ≤1  1-2  >2 | 0.80 (0.71–0.90)  0.72 (0.67–0.78)  0.90 (0.81–1.01) | <0.001  <0.001  0.07 | 1.10 (0.93–1.30)  0.69 (0.64–0.74)  0.95 (0.77–1.17) | 0.28  <0.001  0.63 |
| Percent of assigned ICU teams pharmacist rounded with§ | 0.99 (0.85, 1.16) | 0.95 | 0.88 (0.76, 1.01) | 0.08 |
| Percent days, no pharmacist or pharmacist trainee on rounds | 1.00 (1.00, 1.00) | 0.54 | 1.00 (1.00, 1.00) | 0.13 |
| Percent days, only attending physician⁂ | 1.00 (1.00, 1.00) | 0.28 | 1.00 (1.00, 1.00) | 0.21 |

CI: confidence interval, CMM: comprehensive medication management, CMS: Center for Medicare & Medicaid Services, ICU: intensive care unit, MRC-ICU: medication regimen complexity- intensive care unit, OR: odds ratio, SOFA: sequential organ failure assessment, VA: Veterans Affairs

*worst score during the first 24 hours of ICU stay

**†**Inclusion criteria specified ≥18 years of age; however, some institutions allowed patients ≥18 years of age in the pediatric ICU

**‡**CMM delivered on interprofessional rounds: the pharmacist attended multidisciplinary rounds and verbally provided CMM (including review of medications and recommendations); CMM delivered not on interprofessional rounds: the pharmacist provided by CMM (reviewed medications and provided recommendations) but did not attend interdisciplinary rounds; Abbreviated CMM delivered: the pharmacist provided abbreviated CMM which includes brief medication review but does not include full patient review (e.g. progress notes, results) and recommendations were provided outside of rounds; CMM not delivered (absence of CMM): the patient received no medication review from a pharmacist outside of pharmacokinetic monitoring and prospective medication order verification

**§**Percent of assigned ICU teams pharmacist rounded is calculated by total number of ICU medical teams the pharmacist attended and provided CMM on interprofessional rounds for divided by total number of ICU medical teams providing care for patients that the pharmacist was assigned to provide CMM for

**⁂**Only attending physician refers to composition of the primary medical team. On these days, the patient received care from only the attending physician and did not receive care from any medical residents, fellows, or advanced practice providers.

#### eFigure 21. Cumulative Incidence Function (CIF) for Probability of Discharge Alive across CMM Every Day of ICU Admission

0- CMM every day of ICU admission
1- Absence of CMM for at least 1 day of ICU stay

CIF curves were estimated using a typical patient profile constructed from the imputed dataset, with covariates fixed at representative values:

Hospital type: Academic Medical Center

Hospital CMS star rating: 3

SOFA Score: 4

MRC-ICU Score: 10

ICU pharmacist coverage 1^st^ 24 hours: CMM delivered on interprofessional rounds

ICU type: Medical

Nurse-to-patient ratio: 2

Pharmacist-to-patient ratio (ICU patients): 17

ICU admission day of the week: Monday

Percent of assigned ICU teams pharmacist rounded with: 50

Percent days, no pharmacist or pharmacist trainee: 28.57143

Percent days, only attending physician: 0

Age: 63.82

Sex: Male

CMM: comprehensive medication management, CMS: Center for Medicare & Medicaid Services, ICU: intensive care unit, MRC-ICU: medication regimen complexity-intensive care unit score, SOFA: sequential organ failure assessment

The only variable varied was CMM every day of ICU admission, set to 0 or 1 to generate two scenarios.

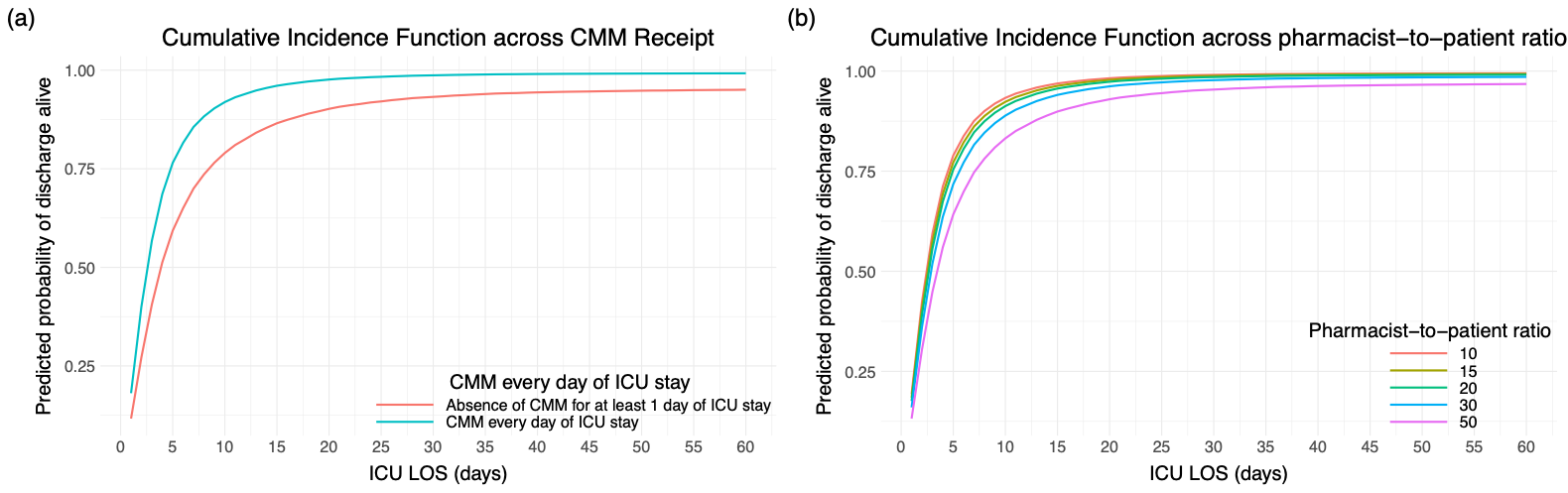

#### eFigure 22. Cumulative Incidence Function (CIF) for Probability of Discharge Alive across Pharmacist-to-patient Ratio (ICU patients) as Points

CIF curves were estimated using a typical patient profile constructed from the imputed dataset, with covariates fixed at representative values:

Hospital type: Academic Medical Center

Hospital CMS star rating: 3

SOFA Score: 4

MRC-ICU Score: 10

ICU pharmacist coverage 1^st^ 24 hours: CMM delivered on interprofessional rounds

ICU type: Medical

Nurse-to-patient ratio: 2

Pharmacist-to-patient ratio (ICU patients): 17

ICU admission day of the week: Monday

Percent of assigned ICU teams pharmacist rounded with: 50

Percent days, no pharmacist or pharmacist trainee: 28.57143

Percent days, only attending physician: 0

Age: 63.82

Sex: Male

The only variable varied was pharmacist-to-patient ratio

CMM: comprehensive medication management, CMS: Center for Medicare & Medicaid Services, ICU: intensive care unit, MRC-ICU: medication regimen complexity-intensive care unit score, SOFA: sequential organ failure assessment, CCP-to-ICU-patient ratio: critical care pharmacist to ICU patient ratio

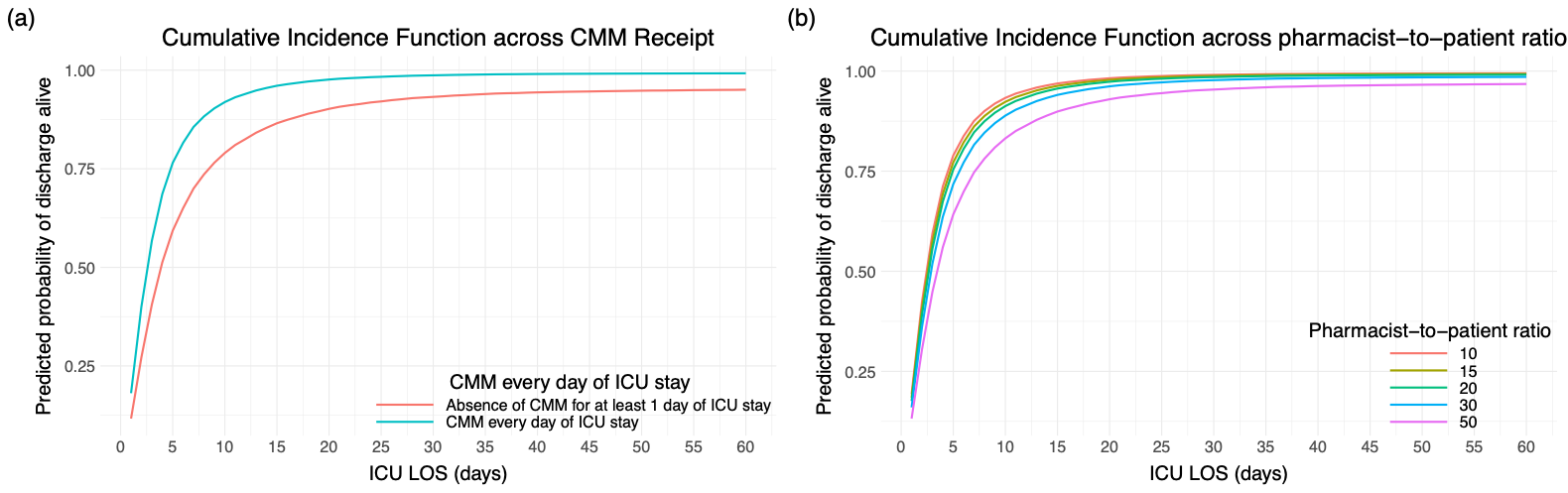

#### eFigure 23. Cumulative Incidence Function (CIF) for Probability of Discharge Alive across Pharmacist-to-patient Ratio (ICU patients) as Ranges

CIF curves were estimated using a typical patient profile constructed from the imputed dataset, with covariates fixed at representative values:

Hospital type: Academic Medical Center

Hospital CMS star rating: 3

SOFA Score: 4

MRC-ICU Score: 10

ICU pharmacist coverage 1^st^ 24 hours: CMM delivered on interprofessional rounds

ICU type: Medical

Nurse-to-patient ratio: 2

Pharmacist-to-patient ratio (ICU patients): 17

ICU admission day of the week: Monday

Percent of assigned ICU teams pharmacist rounded with: 50

Percent days, no pharmacist or pharmacist trainee: 28.57143

Percent days, only attending physician: 0

Age: 63.82

Sex: Male

The only variable varied was pharmacist-to-patient ratio

CMM: comprehensive medication management, CMS: Center for Medicare & Medicaid Services, ICU: intensive care unit, HOS LOS: hospital length of stay, MRC-ICU: medication regimen complexity-intensive care unit score, SOFA: sequential organ failure assessment, CCP-to-ICU-patient ratio: critical care pharmacist to ICU patient ratio

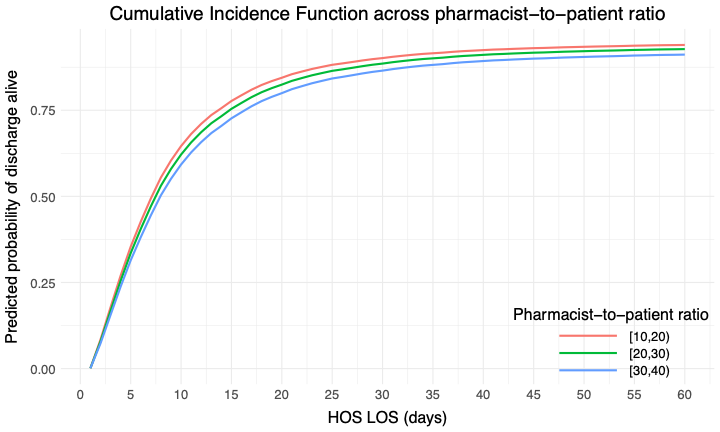

#### eFigure 24. Cumulative Incidence Function (CIF) for Probability of Discharge Alive across Pharmacist-to-patient Ratio (ICU patients) as Ranges at Number Needed to Treat Cutoff

CIF curves were estimated using a typical patient profile constructed from the imputed dataset, with covariates fixed at representative values:

Hospital type: Academic Medical Center

Hospital CMS star rating: 3

SOFA Score: 4

MRC-ICU Score: 10

ICU pharmacist coverage 1^st^ 24 hours: CMM delivered on interprofessional rounds

ICU type: Medical

Nurse-to-patient ratio: 2

Pharmacist-to-patient ratio (ICU patients): 17

ICU admission day of the week: Monday

Percent of assigned ICU teams pharmacist rounded with: 50

Percent days, no pharmacist or pharmacist trainee: 28.57143

Percent days, only attending physician: 0

Age: 63.82

Sex: Male

The only variable varied was pharmacist-to-patient ratio

CMM: comprehensive medication management, CMS: Center for Medicare & Medicaid Services, ICU: intensive care unit, HOS LOS: hospital length of stay, MRC-ICU: medication regimen complexity-intensive care unit score, SOFA: sequential organ failure assessment, CCP-to-ICU-patient ratio: critical care pharmacist to ICU patient ratio

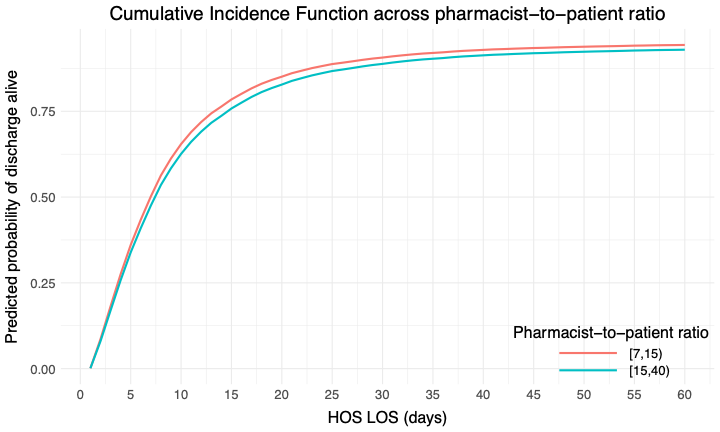

#### eFigure 25. Cumulative Incidence Function (CIF) for Probability of Discharge Alive across Pharmacist-to-patient Ratio (ICU patients) as Ranges Using 1:20 as the Cutoff

CIF curves were estimated using a typical patient profile constructed from the imputed dataset, with covariates fixed at representative values:

Hospital type: Academic Medical Center

Hospital CMS star rating: 3

SOFA Score: 4

MRC-ICU Score: 10

ICU pharmacist coverage 1^st^ 24 hours: CMM delivered on interprofessional rounds

ICU type: Medical

Nurse-to-patient ratio: 2

Pharmacist-to-patient ratio (ICU patients): 17

ICU admission day of the week: Monday

Percent of assigned ICU teams pharmacist rounded with: 50

Percent days, no pharmacist or pharmacist trainee: 28.57143

Percent days, only attending physician: 0

Age: 63.82

Sex: Male

The only variable varied was pharmacist-to-patient ratio

CMM: comprehensive medication management, CMS: Center for Medicare & Medicaid Services, ICU: intensive care unit, HOS LOS: hospital length of stay, MRC-ICU: medication regimen complexity-intensive care unit score, SOFA: sequential organ failure assessment, CCP-to-ICU-patient ratio: critical care pharmacist to ICU patient ratio

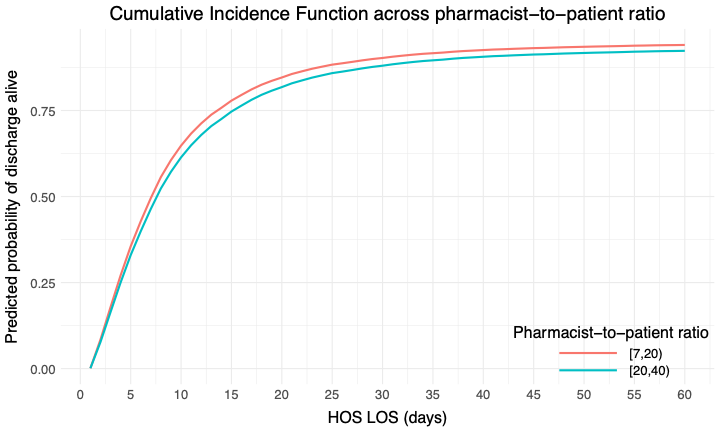

#### eTable 15. Fine-Gray Competing Risks Model for Ventilator Free Days with In-Hospital Mortality as a Competing Event (N = 24418)

Ventilator Free Days modeled using 28 days as a cut and without using 28 days as a cut

|  | **VFD Competing Model with cut (N =24418)** | | **VFD Competing Model without cut (N =24418)** | |
| --- | --- | --- | --- | --- |
|  | **Hazard Ratio (CI)** | **P-value** | **Hazard Ratio (CI)** | **P-value** |
| *Primary Variable* | | | | |
| Pharmacist-to-patient ratio (ICU patients) | 1.00 (0.99, 1.00) | <0.001 | 1.00 (0.99, 1.00) | <0.001 |
| *Secondary Variable* | | | | |
| Absence of CMM for at least 1 day of ICU stay | 0.81 (0.76, 0.86) | <0.001 | 0.81 (0.76, 0.86) | <0.001 |
| *Co-variates* | | | | |
| Age, years | 1.00 (0.99, 1.00) | <0.001 | 1.00 (0.99, 1.00) | <0.001 |
| Sex, female | 0.99 (0.96, 1.01) | 0.32 | 0.99 (0.96, 1.01) | 0.32 |
| SOFA Score* | 0.90 (0.90, 0.91) | <0.001 | 0.90 (0.90, 0.91) | <0.001 |
| MRC-ICU Score* | 0.97 (0.97, 0.97) | <0.001 | 0.97 (0.97, 0.97) | <0.001 |
| ICU admission day of the week (reference: Monday) | - | - | - | - |
| Tuesday | 1.01 (0.97, 1.06) | 0.52 | 1.01 (0.97, 1.06) | 0.52 |
| Wednesday | 1.00 (0.97, 1.04) | 0.81 | 1.00 (0.97, 1.04) | 0.81 |
| Thursday | 1.02 (0.98, 1.07) | 0.35 | 1.02 (0.98, 1.07) | 0.35 |
| Friday | 0.98 (0.93, 1.03) | 0.34 | 0.98 (0.93, 1.03) | 0.34 |
| Saturday | 0.94 (0.89, 1.00) | 0.05 | 0.94 (0.89, 1.00) | 0.05 |
| Sunday | 0.93 (0.89, 0.98) | 0.006 | 0.93 (0.89, 0.98) | 0.007 |
| ICU type† (reference: medical ICU) | - | - | - | - |
| Other | 1.31 (1.10, 1.56) | 0.003 | 1.31 (1.10, 1.56) | 0.003 |
| Surgical | 1.31 (1.20, 1.43) | <0.001 | 1.31 (1.20, 1.43) | <0.001 |
| Surgical/Trauma | 1.07 (0.99, 1.15) | 0.08 | 1.07 (0.99, 1.15) | 0.08 |
| Cardiothoracic | 1.62 (1.44, 1.83) | <0.001 | 1.62 (1.44, 1.83) | <0.001 |
| Cardiac | 1.12 (1.03, 1.22) | 0.006 | 1.12 (1.03, 1.22) | 0.006 |
| Neurosurgery/Neurology | 0.93 (0.87, 0.99) | 0.02 | 0.93 (0.87, 0.99) | 0.02 |
| Pediatric | 0.98 (0.88, 1.10) | 0.74 | 0.98 (0.88, 1.10) | 0.74 |
| Mixed Medical-Surgical | 1.14 (1.04, 1.25) | 0.006 | 1.14 (1.04, 1.25) | 0.006 |
| Burn | 1.09 (0.94, 1.27) | 0.25 | 1.09 (0.94, 1.27) | 0.24 |
| Hospital type (reference: academic medical center) | - | - | - | - |
| Community - Teaching | 0.99 (0.93, 1.05) | 0.70 | 0.99 (0.93, 1.05) | 0.69 |
| Community - Non-teaching | 1.01 (0.91, 1.13) | 0.85 | 1.01 (0.91, 1.13) | 0.85 |
| Government/VA/Military | 1.06 (0.98, 1.14) | 0.14 | 1.06 (0.98, 1.14) | 0.14 |
| Hospital CMS star rating (reference:3) | - | - | - | - |
| 2 | 1.01 (0.95, 1.07) | 0.77 | 1.01 (0.95, 1.07) | 0.77 |
| 4 | 1.04 (0.98, 1.11) | 0.21 | 1.04 (0.98, 1.11) | 0.21 |
| 5 | 1.03 (0.90, 1.19) | 0.66 | 1.03 (0.90, 1.19) | 0.66 |
| ICU pharmacist coverage 1st 24 hours‡ (reference: CMM delivered on interprofessional rounds) | - | - | - | - |
| CMM delivered not on interprofessional rounds | 0.99 (0.95, 1.04) | 0.68 | 0.99 (0.95, 1.04) | 0.68 |
| Abbreviated CMM delivered | 0.99 (0.93, 1.06) | 0.82 | 0.99 (0.93, 1.06) | 0.82 |
| CMM not delivered (absence of CMM) by a CCP | 1.03 (0.96, 1.11) | 0.34 | 1.03 (0.96, 1.11) | 0.34 |
| Unknown | 0.75 (0.61, 0.92) | 0.005 | 0.75 (0.61, 0.92) | 0.005 |
| Nurse-to-patient ratio (average)(reference: 1:2) | - | - | - | - |
| ≤1 | 0.87 (0.76, 1.01) | 0.07 | 0.87 (0.75, 1.01) | 0.06 |
| 1-2 | 0.80 (0.76, 0.85) | <0.001 | 0.80 (0.76, 0.85) | <0.001 |
| >2 | 1.03 (0.94, 1.14) | 0.49 | 1.03 (0.94, 1.14) | 0.49 |
| Percent of assigned ICU teams pharmacist rounded with§ | 0.92 (0.85, 0.98) | 0.02 | 0.92 (0.85, 0.98) | 0.02 |
| Percent days, no pharmacist or pharmacist trainee on rounds | 1.00 (1.00, 1.00) | 0.05 | 1.00 (1.00, 1.00) | 0.05 |
| Percent days, only attending physician⁂ | 1.00 (1.00, 1.00) | 0.52 | 1.00 (1.00, 1.00) | 0.52 |

CI: confidence interval, CMM: comprehensive medication management, CMS: Center for Medicare & Medicaid Services, ICU: intensive care unit, MRC-ICU: medication regimen complexity- intensive care unit, OR: odds ratio, SOFA: sequential organ failure assessment, VA: Veterans Affairs

*worst score during the first 24 hours of ICU stay

**†**Inclusion criteria specified ≥18 years of age; however, some institutions allowed patients ≥18 years of age in the pediatric ICU

**‡**CMM delivered on interprofessional rounds: the pharmacist attended multidisciplinary rounds and verbally provided CMM (including review of medications and recommendations); CMM delivered not on interprofessional rounds: the pharmacist provided by CMM (reviewed medications and provided recommendations) but did not attend interdisciplinary rounds; Abbreviated CMM delivered: the pharmacist provided abbreviated CMM which includes brief medication review but does not include full patient review (e.g. progress notes, results) and recommendations were provided outside of rounds; CMM not delivered (absence of CMM): the patient received no medication review from a pharmacist outside of pharmacokinetic monitoring and prospective medication order verification

**§**Percent of assigned ICU teams pharmacist rounded is calculated by total number of ICU medical teams the pharmacist attended and provided CMM on interprofessional rounds for divided by total number of ICU medical teams providing care for patients that the pharmacist was assigned to provide CMM for

**⁂**Only attending physician refers to composition of the primary medical team. On these days, the patient received care from only the attending physician and did not receive care from any medical residents, fellows, or advanced practice providers.

#### eFigure 26. 28-Day Cumulative Incidence Function (CIF) for Probability of Extubation Alive across CMM Every Day of ICU Admission

0- CMM every day of ICU admission
1- Absence of CMM for at least 1 day of ICU stay

CIF curves were estimated using a typical patient profile constructed from the imputed dataset, with covariates fixed at representative values:

Hospital type: Academic Medical Center

Hospital CMS star rating: 3

SOFA Score: 4

MRC-ICU Score: 10

ICU pharmacist coverage 1^st^ 24 hours: CMM delivered on interprofessional rounds

ICU type: Medical

Nurse-to-patient ratio: 2

Pharmacist-to-patient ratio (ICU patients): 17

ICU admission day of the week: Monday

Percent of assigned ICU teams pharmacist rounded with: 50

Percent days, no pharmacist or pharmacist trainee: 28.57143

Percent days, only attending physician: 0

Age: 63.82

Sex: Male

CCP: critical care pharmacist, CMM: comprehensive medication management, CMS: Center for Medicare & Medicaid Services, ICU: intensive care unit, MRC-ICU: medication regimen complexity-intensive care unit score, SOFA: sequential organ failure assessment

The only variable varied was the presence of CMM every day of ICU stay

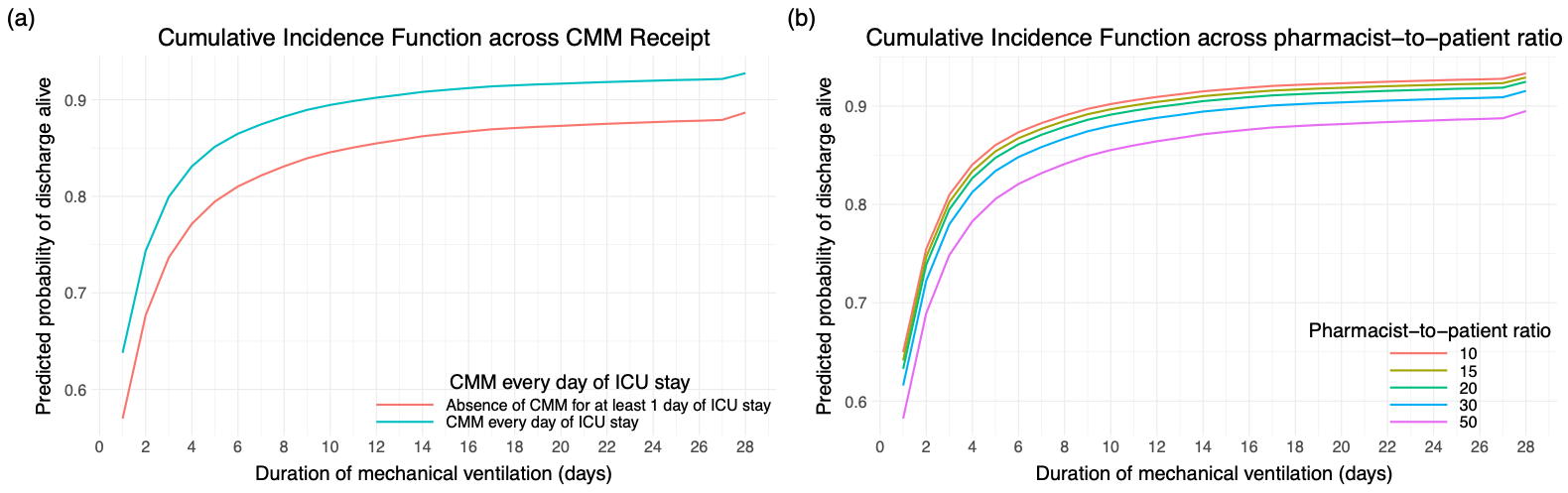

#### eFigure 27. 28-Day Cumulative Incidence Function (CIF) for Probability of Extubation Alive across Pharmacist-to-patient Ratio (ICU patients) as Points

CIF curves were estimated using a typical patient profile constructed from the imputed dataset, with covariates fixed at representative values:

Hospital type: Academic Medical Center

Hospital CMS star rating: 3

SOFA Score: 4

MRC-ICU Score: 10

ICU pharmacist coverage 1^st^ 24 hours: CMM delivered on interprofessional rounds

ICU type: Medical

Nurse-to-patient ratio: 2

Pharmacist-to-patient ratio (ICU patients): 17

ICU admission day of the week: Monday

Percent of assigned ICU teams pharmacist rounded with: 50

Percent days, no pharmacist or pharmacist trainee: 28.57143

Percent days, only attending physician: 0

Age: 63.82

Sex: Male

The only variable varied was pharmacist-to-patient ratio

CCP-to-ICU-patient ratio: critical care pharmacist to ICU patient ratio, CMM: comprehensive medication management, CMS: Center for Medicare & Medicaid Services, ICU: intensive care unit, MRC-ICU: medication regimen complexity-intensive care unit score, SOFA: sequential organ failure assessment

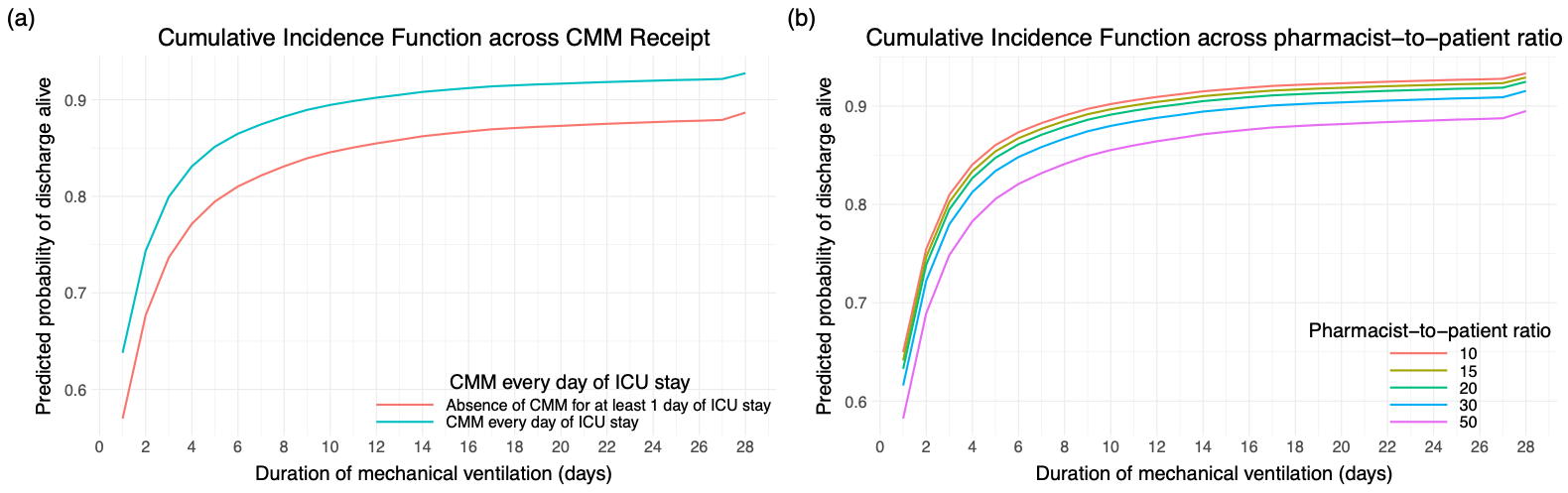

#### eFigure 28. 28-Day Cumulative Incidence Function (CIF) for Probability of Extubation Alive across Pharmacist-to-patient Ratio (ICU patients) as Ranges

CIF curves were estimated using a typical patient profile constructed from the imputed dataset, with covariates fixed at representative values:

Hospital type: Academic Medical Center

Hospital CMS star rating: 3

SOFA Score: 4

MRC-ICU Score: 10

ICU pharmacist coverage 1^st^ 24 hours: CMM delivered on interprofessional rounds

ICU type: Medical

Nurse-to-patient ratio: 2

Pharmacist-to-patient ratio (ICU patients): 17

ICU admission day of the week: Monday

Percent of assigned ICU teams pharmacist rounded with: 50

Percent days, no pharmacist or pharmacist trainee: 28.57143

Percent days, only attending physician: 0

Age: 63.82

Sex: Male

The only variable varied was pharmacist-to-patient ratio

CCP-to-ICU-patient ratio: critical care pharmacist to ICU patient ratio, CMM: comprehensive medication management, CMS: Center for Medicare & Medicaid Services, ICU: intensive care unit, MRC-ICU: medication regimen complexity-intensive care unit score, SOFA: sequential organ failure assessment,

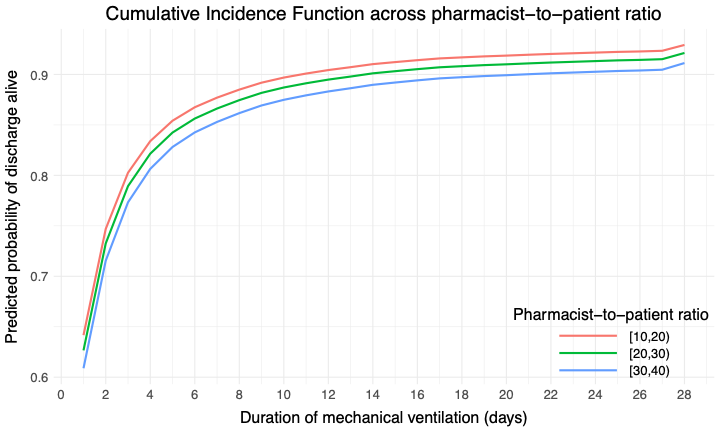

eFigure 31. 28-Day Cumulative Incidence Function for Probability of Extubation Alive across Pharmacist-to-patient Ratio (ICU patients) as Ranges at Number Needed to Treat Cutoff
CIF curves were estimated using a typical patient profile constructed from the imputed dataset, with covariates fixed at representative values:

Hospital type: Academic Medical Center

Hospital CMS star rating: 3

SOFA Score: 4

MRC-ICU Score: 10

ICU pharmacist coverage 1^st^ 24 hours: CMM delivered on interprofessional rounds

ICU type: Medical

Nurse-to-patient ratio: 2

Pharmacist-to-patient ratio (ICU patients): 17

ICU admission day of the week: Monday

Percent of assigned ICU teams pharmacist rounded with: 50

Percent days, no pharmacist or pharmacist trainee: 28.57143

Percent days, only attending physician: 0

Age: 63.82

Sex: Male

The only variable varied was pharmacist-to-patient ratio

CCP-to-ICU-patient ratio: critical care pharmacist to ICU patient ratio, CMM: comprehensive medication management, CMS: Center for Medicare & Medicaid Services, ICU: intensive care unit, MRC-ICU: medication regimen complexity-intensive care unit score, SOFA: sequential organ failure assessment,

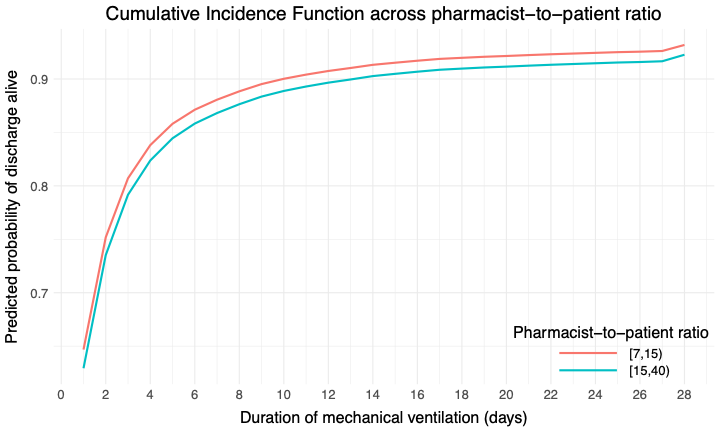

#### eFigure 29. 28-Day Cumulative Incidence Function (CIF) for Probability of Extubation Alive across Pharmacist-to-patient Ratio (ICU patients) as Ranges Using 1:20 as the Cutoff

CIF curves were estimated using a typical patient profile constructed from the imputed dataset, with covariates fixed at representative values:

Hospital type: Academic Medical Center

Hospital CMS star rating: 3

SOFA Score: 4

MRC-ICU Score: 10

ICU pharmacist coverage 1^st^ 24 hours: CMM delivered on interprofessional rounds

ICU type: Medical

Nurse-to-patient ratio: 2

Pharmacist-to-patient ratio (ICU patients): 17

ICU admission day of the week: Monday

Percent of assigned ICU teams pharmacist rounded with: 50

Percent days, no pharmacist or pharmacist trainee: 28.57143

Percent days, only attending physician: 0

Age: 63.82

Sex: Male

The only variable varied was pharmacist-to-patient ratio

CCP-to-ICU-patient ratio: critical care pharmacist to ICU patient ratio, CMM: comprehensive medication management, CMS: Center for Medicare & Medicaid Services, ICU: intensive care unit, MRC-ICU: medication regimen complexity-intensive care unit score, SOFA: sequential organ failure assessment,

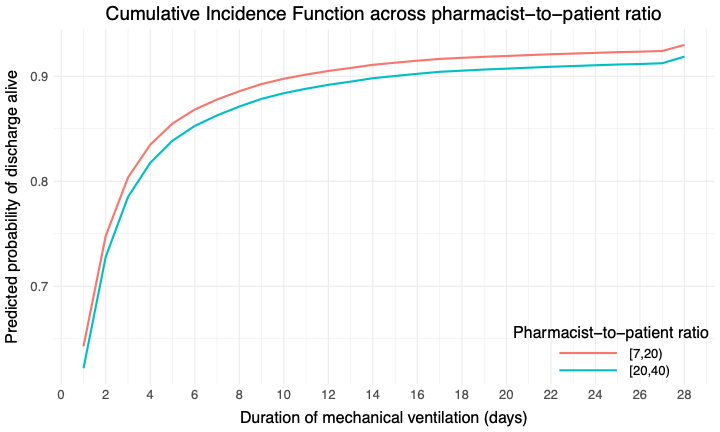

### Sensitivity Analysis- Medical Intensive Care Unit Patients (N=8850)

#### eTable 16. Patient demographics for Medical ICU Patients

|  | **Overall** | **Alive** | **Deceased** | **p-value** |
| --- | --- | --- | --- | --- |
|  | N = 8,850 | N = 7,084 | N = 1,766 |  |
| Percent of assigned ICU teams pharmacist rounded with§ | 56.9 (30.5) | 56.1 (30.6) | 60.1 (29.8) | <0.001 |
| Missing | 119 (1.3) | 101 (1.4) | 18 (1.0) |  |
| Percent days, no pharmacist or pharmacist trainee | 28.6 (33.0) | 28.8 (33.5) | 27.8 (30.8) | 0.6 |
| Missing | 211 (2.4) | 148 (2.1) | 63 (3.6) |  |
| Percent days, only attending physician⁂ | 5.7 (22.9) | 5.7 (22.9) | 5.8 (23.2) | 0.8 |
| Missing | 257 (2.9) | 174 (2.5) | 83 (4.7) |  |
| CMM every day of ICU stay |  |  |  | <0.001 |
| Yes | 7,214 (81.5) | 5,858 (82.7) | 1,356 (76.8) |  |
| No | 1,636 (18.5) | 1,226 (17.3) | 410 (23.2) |  |
| Age, years | 60.7 (16.8) | 59.9 (17.1) | 64.3 (15.1) | <0.001 |
| Sex |  |  |  | 0.3 |
| Male | 4,873 (55.1) | 3,872 (54.7) | 1,001 (56.7) |  |
| Female | 3,976 (44.9) | 3,211 (45.3) | 765 (43.3) |  |
| Unidentified | 1 (0.0) | 1 (0.0) | 0 (0.0) |  |
| SOFA Score | 6.0 (4.2) | 5.3 (3.8) | 9.0 (4.5) | <0.001 |
| Missing | 11 (0.1) | 10 (0.1) | 1 (<0.1) |  |
| Dialysis |  |  |  | <0.001 |
| No | 7,401 (83.6) | 6,123 (86.5) | 1,278 (72.4) |  |
| Yes | 1,447 (16.4) | 959 (13.5) | 488 (27.6) |  |
| Missing | 2 (<0.1) | 2 (<0.1) | 0 (0) |  |
| ECMO |  |  |  | <0.001 |
| No | 8,806 (99.5) | 7,057 (99.6) | 1,749 (99.0) |  |
| Yes | 42 (0.5) | 25 (0.4) | 17 (1.0) |  |
| Missing | 2 (<0.1) | 2 (<0.1) | 0 (0) |  |
| Mechanical circulatory support |  |  |  | <0.001 |
| No | 4,445 (50.2) | 4,066 (57.4) | 379 (21.5) |  |
| Yes | 4,403 (49.8) | 3,016 (42.6) | 1,387 (78.5) |  |
| Missing | 2 (<0.1) | 2 (<0.1) | 0 (0) |  |
| Mechanical ventilation |  |  |  | <0.001 |
| No | 4,973 (56.2) | 4,405 (62.2) | 568 (32.2) |  |
| Yes | 3,875 (43.8) | 2,677 (37.8) | 1,198 (67.8) |  |
| Missing | 2 (<0.1) | 2 (<0.1) | 0 (0) |  |
| MRC-ICU Score | 11.3 (6.5) | 10.6 (6.2) | 14.3 (7.0) | <0.001 |
| Missing | 2 (<0.1) | 2 (<0.1) | 0 (0) |  |
| ICU admission day of the week |  |  |  | 0.3 |
| Monday | 1,386 (15.7) | 1,097 (15.5) | 289 (16.4) |  |
| Tuesday | 1,314 (14.8) | 1,056 (14.9) | 258 (14.6) |  |
| Wednesday | 1,314 (14.8) | 1,044 (14.7) | 270 (15.3) |  |
| Thursday | 1,306 (14.8) | 1,046 (14.8) | 260 (14.7) |  |
| Friday | 1,236 (14.0) | 1,017 (14.4) | 219 (12.4) |  |
| Saturday | 1,099 (12.4) | 861 (12.2) | 238 (13.5) |  |
| Sunday | 1,195 (13.5) | 963 (13.6) | 232 (13.1) |  |
| Hospital LOS | 11.8 (15.9) | 11.3 (14.5) | 13.8 (20.5) | <0.001 |
| Missing | 3 (<0.1) | 3 (<0.1) | 0 (0) |  |
| ICU LOS | 5.6 (8.9) | 5.0 (7.1) | 8.1 (13.5) | <0.001 |
| ICU free days | 18.8 (10.3) | 23.3 (5.3) | 0.0 (0.0) | <0.001 |
| Missing | 73 (0.8) | 9 (0.1) | 64 (3.6) |  |
| Duration of mechanical ventilation | 2.4 (6.6) | 1.9 (6.3) | 4.5 (7.3) | <0.001 |
| Missing | 3 (<0.1) | 3 (<0.1) | 0 (0) |  |
| VFDs | 21.0 (11.2) | 26.3 (4.2) | 0.0 (0.0) | <0.001 |
| Missing | 3 (<0.1) | 3 (<0.1) | 0 (0) |  |
| Hospital Type |  |  |  | <0.001 |
| Academic medical center | 7,114 (80.4) | 5,642 (79.6) | 1,472 (83.4) |  |
| Community - Teaching | 905 (10.2) | 733 (10.3) | 172 (9.7) |  |
| Community - Non-teaching | 756 (8.5) | 646 (9.1) | 110 (6.2) |  |
| Government/VA/Military | 75 (0.8) | 63 (0.9) | 12 (0.7) |  |
| Hospital CMS star rating |  |  |  | 0.001 |
| 2 | 1,226 (14.3) | 969 (14.0) | 257 (15.6) |  |
| 3 | 5,355 (62.4) | 4,387 (63.3) | 968 (58.9) |  |
| 4 | 1,752 (20.4) | 1,371 (19.8) | 381 (23.2) |  |
| 5 | 244 (2.8) | 206 (3.0) | 38 (2.3) |  |
| Missing | 273 (3.1) | 151 (2.1) | 122 (6.9) |  |
| ICU pharmacist coverage 1^st^ 24 hours | 0 (0.0) | 0 (0.0) | 0 (0.0) |  |
| CMM delivered on interprofessional rounds |  |  |  | 0.004 |
| CMM delivered not on interprofessional rounds | 6,314 (71.4) | 5,068 (71.6) | 1,246 (70.6) |  |
| Abbreviated CMM delivered | 1,093 (12.4) | 891 (12.6) | 202 (11.4) |  |
| CMM not delivered (absence of CMM) | 734 (8.3) | 595 (8.4) | 139 (7.9) |  |
| Unknown | 663 (7.5) | 497 (7.0) | 166 (9.4) |  |
| Missing | 44 (0.5) | 31 (0.4) | 13 (0.7) |  |
| Pharmacist-to-patient ratio (all patients) | 2 (<0.1) | 2 (<0.1) | 0 (0) |  |
| Missing | 28.3 (23.2) | 28.8 (24.1) | 26.3 (19.0) | <0.001 |
| Pharmacist-to-patient ratio (ICU patients) | 105 (1.2) | 89 (1.3) | 16 (0.9) |  |
| Missing | 20.1 (11.9) | 20.2 (12.0) | 19.7 (11.3) | 0.3 |
| Nurse-to-patient ratio (average), 1:X |  |  |  | <0.001 |
| ≤1 | 676 (7.7) | 452 (6.4) | 224 (12.7) |  |
| =2 | 6,912 (78.6) | 5,691 (80.9) | 1,221 (69.4) |  |
| >2 | 335 (3.8) | 297 (4.2) | 38 (2.2) |  |
| 1-2 | 874 (9.9) | 597 (8.5) | 277 (15.7) |  |
| Missing | 53 (0.6) | 47 (0.7) | 6 (0.3) |  |

Data presented as mean (±SD) or n (%)

CMM: comprehensive medication management, CMS: Center for Medicare & Medicaid Services, ICU: intensive care unit, LOS: length of stay, MRC-ICU, medication regimen complexity- intensive care unit, SOFA: sequential organ failure assessment, VA: Veterans Affairs, VFDs: ventilator free days

*worst score during the first 24 hours of ICU stay

**†**Inclusion criteria specified ≥18 years of age; however, some institutions allowed patients ≥18 years of age in the pediatric ICU

**‡**CMM delivered on interprofessional rounds: the pharmacist attended multidisciplinary rounds and verbally provided CMM (including review of medications and recommendations); CMM delivered not on interprofessional rounds: the pharmacist provided by CMM (reviewed medications and provided recommendations) but did not attend interdisciplinary rounds; Abbreviated CMM delivered: the pharmacist provided abbreviated CMM which includes brief medication review but does not include full patient review (e.g. progress notes, results) and recommendations were provided outside of rounds; CMM not delivered (absence of CMM): the patient received no medication review from a pharmacist outside of pharmacokinetic monitoring and prospective medication order verification

**§**Percent of assigned ICU teams pharmacist rounded is calculated by total number of ICU medical teams the pharmacist attended and provided CMM on interprofessional rounds for divided by total number of ICU medical teams providing care for patients that the pharmacist was assigned to provide CMM for

**⁂**Only attending physician refers to composition of the primary medical team. On these days, the patient received care from only the attending physician and did not receive care from any medical residents, fellows, or advanced practice providers.

#### eTable 17. Univariable and Multivariable GEE Analyses for Mortality Based on Multiply Imputed Data for Medical ICU Patients (N = 8850)

| **Variable** | **Univariable** | | **Multivariable** | |
| --- | --- | --- | --- | --- |
|  | **OR (CI)** | **P-value** | **OR (CI)** | **P-value** |
| *Primary Variable* | | | | |
| Pharmacist-to-patient ratio (ICU patients) | 1.00 (0.99–1.00) | 0.16 | 1.00 (0.99–1.01) | 0.90 |
| *Secondary Variable* | | | | |
| Absence of CMM for at least 1 day of ICU stay | 1.62 (1.39–1.87) | <0.001 | 1.20 (1.01–1.42) | 0.04 |
| *Co-variates* | | | | |
| Age, years | 1.02 (1.01–1.02) | <0.001 | 1.02 (1.02–1.03) | <0.001 |
| Sex, female | 0.94 (0.83–1.06) | 0.31 | 1.03 (0.90–1.17) | 0.71 |
| SOFA Score* | 1.24 (1.22–1.26) | <0.001 | 1.27 (1.22–1.32) | <0.001 |
| MRC-ICU Score* | 1.09 (1.08–1.10) | <0.001 | 1.04 (1.01–1.07) | 0.002 |
| SOFA-MRC-ICU interaction term | 1.01 (1.01–1.01) | <0.001 | 1.00 (0.99–1.00) | 0.03 |
| ICU admission day of the week (reference: Monday) | - | - | - | - |
| Tuesday | 0.93 (0.77–1.14) | 0.49 | 0.95 (0.77–1.17) | 0.65 |
| Wednesday | 0.99 (0.82–1.19) | 0.89 | 1.00 (0.85–1.18) | 0.99 |
| Thursday | 0.93 (0.80–1.09) | 0.39 | 0.94 (0.77–1.14) | 0.53 |
| Friday | 0.85 (0.73–0.98) | 0.03 | 0.81 (0.68–0.95) | 0.01 |
| Saturday | 1.05 (0.87–1.28) | 0.61 | 0.93 (0.75–1.16) | 0.52 |
| Sunday | 0.92 (0.78–1.10) | 0.37 | 0.88 (0.72–1.07) | 0.2 |
| Hospital type (reference: academic medical center) | - | - | - | - |
| Community - Teaching | 0.89 (0.63–1.27) | 0.52 | 0.97 (0.74–1.26) | 0.8 |
| Community - Non-teaching | 0.64 (0.47–0.88) | 0.006 | 0.77 (0.49–1.21) | 0.26 |
| Government/VA/Military | 0.70 (0.59–0.83) | <0.001 | 1.14 (0.80–1.62) | 0.48 |
| Hospital CMS star rating (reference: 3) |  |  |  |  |
| 2 | 1.14 (0.86–1.52) | 0.37 | 1.03 (0.76–1.38) | 0.87 |
| 4 | 1.15 (0.78–1.70) | 0.47 | 0.72 (0.45–1.14) | 0.15 |
| 5 | 0.77 (0.53–1.11) | 0.16 | 0.45 (0.30–0.68) | <0.001 |
| ICU pharmacist coverage 1st 24 hours‡ (reference: CMM delivered on interprofessional rounds) | - | - | - | - |
| CMM delivered not on interprofessional rounds | 1.00 (0.84–1.20) | 0.96 | 1.14 (0.93–1.40) | 0.19 |
| Abbreviated CMM delivered | 1.00 (0.88–1.13) | 0.94 | 1.20 (0.97–1.49) | 0.09 |
| CMM not delivered (absence of CMM) | 1.35 (1.09–1.66) | 0.005 | 1.30 (1.04–1.64) | 0.02 |
| Unknown | 1.61 (0.62–4.23) | 0.33 | 1.10 (0.30–4.00) | 0.89 |
| Nurse-to-patient ratio (average)(reference: 1:2) | - | - | - | - |
| ≤1 | 3.51 (2.00–6.14) | <0.001 | 3.15 (1.60–6.19) | <0.001 |
| 1-2 | 2.57 (1.91–3.46) | <0.001 | 1.77 (1.26–2.47) | <0.001 |
| >2 | 0.68 (0.43–1.09) | 0.11 | 0.83 (0.56–1.25) | 0.37 |
| Percent of assigned ICU teams pharmacist rounded with§ | 1.34 (1.06–1.71) | 0.02 | 1.35 (0.98–1.86) | 0.07 |
| Percent days, no pharmacist or pharmacist trainee on rounds | 1.00 (1.00–1.00) | 0.90 | 1.00 (1.00–1.00) | 0.29 |
| Percent days, only attending physician⁂ | 1.00 (0.99–1.00) | 0.08 | 1.00 (0.99–1.00) | 0.50 |

CI: confidence interval, CMM: comprehensive medication management, CMS: Center for Medicare & Medicaid Services, ICU: intensive care unit, MRC-ICU: medication regimen complexity- intensive care unit, OR: odds ratio, SOFA: sequential organ failure assessment, VA: Veterans Affairs

*worst score during the first 24 hours of ICU stay

**†**Inclusion criteria specified ≥18 years of age; however, some institutions allowed patients ≥18 years of age in the pediatric ICU

**‡**CMM delivered on interprofessional rounds: the pharmacist attended multidisciplinary rounds and verbally provided CMM (including review of medications and recommendations); CMM delivered not on interprofessional rounds: the pharmacist provided by CMM (reviewed medications and provided recommendations) but did not attend interdisciplinary rounds; Abbreviated CMM delivered: the pharmacist provided abbreviated CMM which includes brief medication review but does not include full patient review (e.g. progress notes, results) and recommendations were provided outside of rounds; CMM not delivered (absence of CMM): the patient received no medication review from a pharmacist outside of pharmacokinetic monitoring and prospective medication order verification

**§**Percent of assigned ICU teams pharmacist rounded is calculated by total number of ICU medical teams the pharmacist attended and provided CMM on interprofessional rounds for divided by total number of ICU medical teams providing care for patients that the pharmacist was assigned to provide CMM for

**⁂**Only attending physician refers to composition of the primary medical team. On these days, the patient received care from only the attending physician and did not receive care from any medical residents, fellows, or advanced practice providers.

#### eTable 18. Fine-Gray Competing Risks Model for ICU and Hospital Length of Stay with In-ICU Mortality as a Competing Event for Medical ICU Patients (N = 8847)

| **Variable** | **Hospital Length of Stay**  **(N = 8847)** | | **ICU Length of Stay (N = 8847)** | |
| --- | --- | --- | --- | --- |
|  | **Hazard Ratio (CI)** | **P-value** | **Hazard Ratio (CI)** | **P-value** |
| *Primary Variable* | | | | |
| Pharmacist-to-patient ratio (ICU patients) | 0.99 (0.99, 1.00) | 0.04 | 0.99 (0.98, 1.00) | 0.002 |
| *Secondary Variable* | | | | |
| Absence of CMM for at least 1 day of ICU stay | 0.73 (0.63, 0.84) | <0.001 | 0.64 (0.53, 0.79) | <0.001 |
| *Co-variates* | | | | |
| Age, years | 0.99 (0.99, 0.99) | <0.001 | 1.00 (0.99, 1.00) | 0.002 |
| Sex, female | 0.99 (0.94, 1.06) | 0.84 | 1.04 (0.99, 1.10) | 0.09 |
| SOFA Score* | 0.90 (0.89, 0.91) | <0.001 | 0.94 (0.92, 0.95) | <0.001 |
| MRC-ICU Score* | 0.98 (0.97, 0.98) | <0.001 | 0.98 (0.97, 0.99) | <0.001 |
| ICU admission day of the week (reference: Monday) | - | - | - | - |
| Tuesday | 1.05 (0.97, 1.14) | 0.23 | 1.04 (0.98, 1.11) | 0.23 |
| Wednesday | 1.03 (0.96, 1.10) | 0.48 | 1.02 (0.96, 1.08) | 0.58 |
| Thursday | 1.08 (1.00, 1.16) | 0.05 | 1.02 (0.94, 1.10) | 0.63 |
| Friday | 1.10 (1.02, 1.19) | 0.02 | 0.97 (0.87, 1.08) | 0.57 |
| Saturday | 1.06 (0.95, 1.17) | 0.29 | 0.96 (0.86, 1.08) | 0.49 |
| Sunday | 1.06 (0.97, 1.16) | 0.17 | 1.08 (1.00, 1.16) | 0.04 |
| Hospital type (reference: academic medical center) | - | - | - | - |
| Community - Teaching | 1.00 (0.80, 1.26) | 1 | 1.02 (0.77, 1.37) | 0.87 |
| Community - Non-teaching | 1.83 (1.36, 2.46) | <0.001 | 0.77 (0.58, 1.03) | 0.08 |
| Government/VA/Military | 2.73 (2.20, 3.39) | <0.001 | 0.57 (0.46, 0.71) | <0.001 |
| Hospital CMS star rating (reference: 3) | - | - | - | - |
| 2 | 0.87 (0.75, 1.01) | 0.07 | 1.10 (0.83, 1.47) | 0.51 |
| 4 | 0.79 (0.66, 0.96) | 0.02 | 1.31 (0.98, 1.74) | 0.07 |
| 5 | 0.92 (0.77, 1.10) | 0.37 | 1.99 (1.48, 2.67) | <0.001 |
| ICU pharmacist coverage 1st 24 hours‡ (reference: CMM delivered on interprofessional rounds) | - | - | - | - |
| CMM delivered not on interprofessional rounds | 0.96 (0.87, 1.05) | 0.39 | 0.93 (0.85, 1.02) | 0.10 |
| Abbreviated CMM delivered | 0.91 (0.82, 1.02) | 0.09 | 0.98 (0.85, 1.15) | 0.84 |
| CMM not delivered (absence of CMM) | 1.04 (0.89, 1.21) | 0.63 | 1.09 (0.93, 1.27) | 0.30 |
| Unknown | 1.02 (0.70, 1.49) | 0.92 | 0.80 (0.52, 1.23) | 0.30 |
| Nurse-to-patient ratio (average)(reference: 1:2) | - | - | - | - |
| ≤1 | 0.69 (0.55, 0.87) | 0.002 | 0.75 (0.36, 1.55) | 0.44 |
| 1-2 | 0.70 (0.59, 0.83) | <0.001 | 0.66 (0.60, 0.74) | <0.001 |
| >2 | 0.94 (0.81, 1.08) | 0.38 | 1.02 (0.71, 1.48) | 0.91 |
| Percent of assigned ICU teams pharmacist rounded with§ | 0.90 (0.73, 1.11) | 0.32 | 0.58 (0.41, 0.83) | 0.003 |
| Percent days, no pharmacist or pharmacist trainee on rounds | 1.00 (1.00, 1.00) | 0.63 | 1.00 (1.00, 1.00) | 0.27 |
| Percent days, only attending physician⁂ | 1.00 (1.00, 1.00) | 0.93 | 1.00 (1.00, 1.00) | 0.07 |

CI: confidence interval, CMM: comprehensive medication management, CMS: Center for Medicare & Medicaid Services, ICU: intensive care unit, MRC-ICU: medication regimen complexity- intensive care unit, OR: odds ratio, SOFA: sequential organ failure assessment, VA: Veterans Affairs

*worst score during the first 24 hours of ICU stay

**†**Inclusion criteria specified ≥18 years of age; however, some institutions allowed patients ≥18 years of age in the pediatric ICU

**‡**CMM delivered on interprofessional rounds: the pharmacist attended multidisciplinary rounds and verbally provided CMM (including review of medications and recommendations); CMM delivered not on interprofessional rounds: the pharmacist provided by CMM (reviewed medications and provided recommendations) but did not attend interdisciplinary rounds; Abbreviated CMM delivered: the pharmacist provided abbreviated CMM which includes brief medication review but does not include full patient review (e.g. progress notes, results) and recommendations were provided outside of rounds; CMM not delivered (absence of CMM): the patient received no medication review from a pharmacist outside of pharmacokinetic monitoring and prospective medication order verification

**§**Percent of assigned ICU teams pharmacist rounded is calculated by total number of ICU medical teams the pharmacist attended and provided CMM on interprofessional rounds for divided by total number of ICU medical teams providing care for patients that the pharmacist was assigned to provide CMM for

**⁂**Only attending physician refers to composition of the primary medical team. On these days, the patient received care from only the attending physician and did not receive care from any medical residents, fellows, or advanced practice providers.

#### eTable 19. Fine-Gray Competing Risks Model for VFD with In-Hospital Mortality as a Competing Event and 28 days as a cut (N = 8845).

| **VFD Competing Model with cut (N = 8845)** | | |
| --- | --- | --- |
| **Variable** | **Hazard Ratio (CI)** | **P-value** |
| *Primary Variable* | | |
| Pharmacist-to-patient ratio (ICU patients) | 1.00 (0.99, 1.00) | 0.02 |
| *Secondary Variable* | | |
| Absence of CMM for at least 1 day of ICU stay | 0.81 (0.73, 0.90) | <0.001 |
| *Covariates* | | |
| Age, years | 0.99 (0.99, 1.00) | <0.001 |
| Sex, female | 1.00 (0.95, 1.05) | 0.93 |
| SOFA Score* | 0.92 (0.91, 0.92) | <0.001 |
| MRC-ICU Score* | 0.98 (0.97, 0.98) | <0.001 |
| ICU admission day of the week (reference: Monday) |  |  |
| Tuesday | 1.03 (0.95, 1.11) | 0.51 |
| Wednesday | 0.99 (0.93, 1.06) | 0.85 |
| Thursday | 1.03 (0.95, 1.10) | 0.50 |
| Friday | 1.04 (0.97, 1.10) | 0.27 |
| Saturday | 0.98 (0.90, 1.06) | 0.55 |
| Sunday | 1.03 (0.96, 1.11) | 0.36 |
| Hospital type (reference: academic medical center) | - | - |
| Community - Teaching | 1.04 (0.93, 1.15) | 0.49 |
| Community - Non-teaching | 1.10 (0.97, 1.24) | 0.13 |
| Government/VA/Military | 1.01 (0.91, 1.11) | 0.91 |
| Hospital CMS star rating (reference: 3) |  |  |
| 2 | 0.98 (0.90, 1.07) | 0.72 |
| 4 | 1.05 (0.97, 1.15) | 0.23 |
| 5 | 1.20 (1.10, 1.31) | <0.001 |
| ICU pharmacist coverage 1st 24 hours‡ (reference: CMM delivered on interprofessional rounds) | - | - |
| CMM delivered not on interprofessional rounds | 0.95 (0.89, 1.02) | 0.18 |
| Abbreviated CMM delivered | 0.95 (0.87, 1.03) | 0.22 |
| CMM not delivered (absence of CMM) | 0.99 (0.88, 1.11) | 0.83 |
| Unknown | 0.87 (0.66, 1.14) | 0.31 |
| Nurse-to-patient ratio (average)(reference: 1:2) | - | - |
| ≤1 | 0.73 (0.48, 1.12) | 0.15 |
| 1-2 | 0.76 (0.67, 0.87) | <0.001 |
| >2 | 1.05 (0.95, 1.15) | 0.34 |
| Percent of assigned ICU teams pharmacist rounded with§ | 0.82 (0.71, 0.95) | 0.009 |
| Percent days, no pharmacist or pharmacist trainee on rounds | 1.00 (1.00, 1.00) | 0.08 |
| Percent days, only attending physician⁂ | 1.00 (1.00, 1.00) | 0.81 |

CI: confidence interval, CMM: comprehensive medication management, CMS: Center for Medicare & Medicaid Services, ICU: intensive care unit, MRC-ICU: medication regimen complexity- intensive care unit, OR: odds ratio, SOFA: sequential organ failure assessment, VA: Veterans Affairs

*worst score during the first 24 hours of ICU stay

**†**Inclusion criteria specified ≥18 years of age; however, some institutions allowed patients ≥18 years of age in the pediatric ICU

**‡**CMM delivered on interprofessional rounds: the pharmacist attended multidisciplinary rounds and verbally provided CMM (including review of medications and recommendations); CMM delivered not on interprofessional rounds: the pharmacist provided by CMM (reviewed medications and provided recommendations) but did not attend interdisciplinary rounds; Abbreviated CMM delivered: the pharmacist provided abbreviated CMM which includes brief medication review but does not include full patient review (e.g. progress notes, results) and recommendations were provided outside of rounds; CMM not delivered (absence of CMM): the patient received no medication review from a pharmacist outside of pharmacokinetic monitoring and prospective medication order verification

**§**Percent of assigned ICU teams pharmacist rounded is calculated by total number of ICU medical teams the pharmacist attended and provided CMM on interprofessional rounds for divided by total number of ICU medical teams providing care for patients that the pharmacist was assigned to provide CMM for

**⁂**Only attending physician refers to composition of the primary medical team. On these days, the patient received care from only the attending physician and did not receive care from any medical residents, fellows, or advanced practice providers.

#### eTable 20. Fine-Gray Competing Risks Model for VFD with In-Hospital Mortality as a Competing Event without a cut (N = 8845).

| **VFD Competing Model without cut (N = 8845)** | | |
| --- | --- | --- |
| **Variable** | **Hazard Ratio (CI)** | **P-value** |
| *Primary Variable* | | |
| Pharmacist-to-patient ratio (ICU patients) | 1.00 (0.99, 1.00) | 0.02 |
| *Secondary Variable* | | |
| Absence of CMM for at least 1 day of ICU stay | 0.81 (0.73, 0.90) | <0.001 |
| *Covariates* | | |
| Age, years | 0.99 (0.99, 1.00) | <0.001 |
| Sex, female | 1.00 (0.95, 1.05) | 0.93 |
| SOFA Score* | 0.92 (0.91, 0.92) | <0.001 |
| MRC-ICU Score* | 0.98 (0.97, 0.98) | <0.001 |
| ICU admission day of the week (reference: Monday) | - | - |
| Tuesday | 1.03 (0.95, 1.11) | 0.51 |
| Wednesday | 0.99 (0.93, 1.06) | 0.85 |
| Thursday | 1.03 (0.95, 1.10) | 0.50 |
| Friday | 1.04 (0.97, 1.10) | 0.27 |
| Saturday | 0.98 (0.90, 1.06) | 0.55 |
| Sunday | 1.03 (0.96, 1.11) | 0.36 |
| Hospital type (reference: academic medical center) | - | - |
| Community - Teaching | 1.04 (0.93, 1.15) | 0.49 |
| Community - Non-teaching | 1.10 (0.97, 1.24) | 0.13 |
| Government/VA/Military | 1.01 (0.91, 1.11) | 0.91 |
| Hospital CMS star rating |  |  |
| 2 | 0.98 (0.90, 1.07) | 0.72 |
| 4 | 1.05 (0.97, 1.15) | 0.23 |
| 5 | 1.20 (1.10, 1.31) | <0.001 |
| ICU pharmacist coverage 1st 24 hours‡ (reference: CMM delivered on interprofessional rounds) | - | - |
| CMM delivered not on interprofessional rounds | 0.95 (0.89, 1.02) | 0.18 |
| Abbreviated CMM delivered | 0.95 (0.87, 1.03) | 0.22 |
| CMM not delivered (absence of CMM) | 0.99 (0.88, 1.11) | 0.83 |
| Unknown | 0.87 (0.66, 1.14) | 0.31 |
| Nurse-to-patient ratio (average)(reference: 1:2) | - | - |
| ≤1 | 0.73 (0.48, 1.12) | 0.15 |
| 1-2 | 0.76 (0.67, 0.87) | <0.001 |
| >2 | 1.05 (0.95, 1.15) | 0.34 |
| Percent of assigned ICU teams pharmacist rounded with§ | 0.82 (0.71, 0.95) | 0.009 |
| Percent days, no pharmacist or pharmacist trainee on rounds | 1.00 (1.00, 1.00) | 0.08 |
| Percent days, only attending physician⁂ | 1.00 (1.00, 1.00) | 0.81 |

CI: confidence interval, CMM: comprehensive medication management, CMS: Center for Medicare & Medicaid Services, ICU: intensive care unit, MRC-ICU: medication regimen complexity- intensive care unit, OR: odds ratio, SOFA: sequential organ failure assessment, VA: Veterans Affairs

*worst score during the first 24 hours of ICU stay

**†**Inclusion criteria specified ≥18 years of age; however, some institutions allowed patients ≥18 years of age in the pediatric ICU

**‡**CMM delivered on interprofessional rounds: the pharmacist attended multidisciplinary rounds and verbally provided CMM (including review of medications and recommendations); CMM delivered not on interprofessional rounds: the pharmacist provided by CMM (reviewed medications and provided recommendations) but did not attend interdisciplinary rounds; Abbreviated CMM delivered: the pharmacist provided abbreviated CMM which includes brief medication review but does not include full patient review (e.g. progress notes, results) and recommendations were provided outside of rounds; CMM not delivered (absence of CMM): the patient received no medication review from a pharmacist outside of pharmacokinetic monitoring and prospective medication order verification

**§**Percent of assigned ICU teams pharmacist rounded is calculated by total number of ICU medical teams the pharmacist attended and provided CMM on interprofessional rounds for divided by total number of ICU medical teams providing care for patients that the pharmacist was assigned to provide CMM for

**⁂**Only attending physician refers to composition of the primary medical team. On these days, the patient received care from only the attending physician and did not receive care from any medical residents, fellows, or advanced practice providers.

#### eFigure 30. Cumulative Incidence Function (CIF) for Probability of Discharge Alive across CMM Every Day of ICU Admission

0- CMM every day of ICU admission
1- Absence of CMM for at least 1 day of ICU stay

CIF curves were estimated using a typical patient profile constructed from the imputed dataset, with covariates fixed at representative values:

Hospital type: Academic Medical Center

Hospital CMS star rating: 3

SOFA Score: 4

MRC-ICU Score: 10

ICU pharmacist coverage 1^st^ 24 hours: CMM delivered on interprofessional rounds

ICU type: Medical

Nurse-to-patient ratio: 2

Pharmacist-to-patient ratio (ICU patients): 17

ICU admission day of the week: Monday

Percent of assigned ICU teams pharmacist rounded with: 50

Percent days, no pharmacist or pharmacist trainee: 28.57143

Percent days, only attending physician: 0

Age: 63.82

Sex: Male

The only variable varied was CMM every day of ICU admission

CCP: critical care pharmacist, CMM: comprehensive medication management, CMS: Center for Medicare & Medicaid Services, ICU: intensive care unit, MRC-ICU: medication regimen complexity-intensive care unit score, SOFA: sequential organ failure assessment

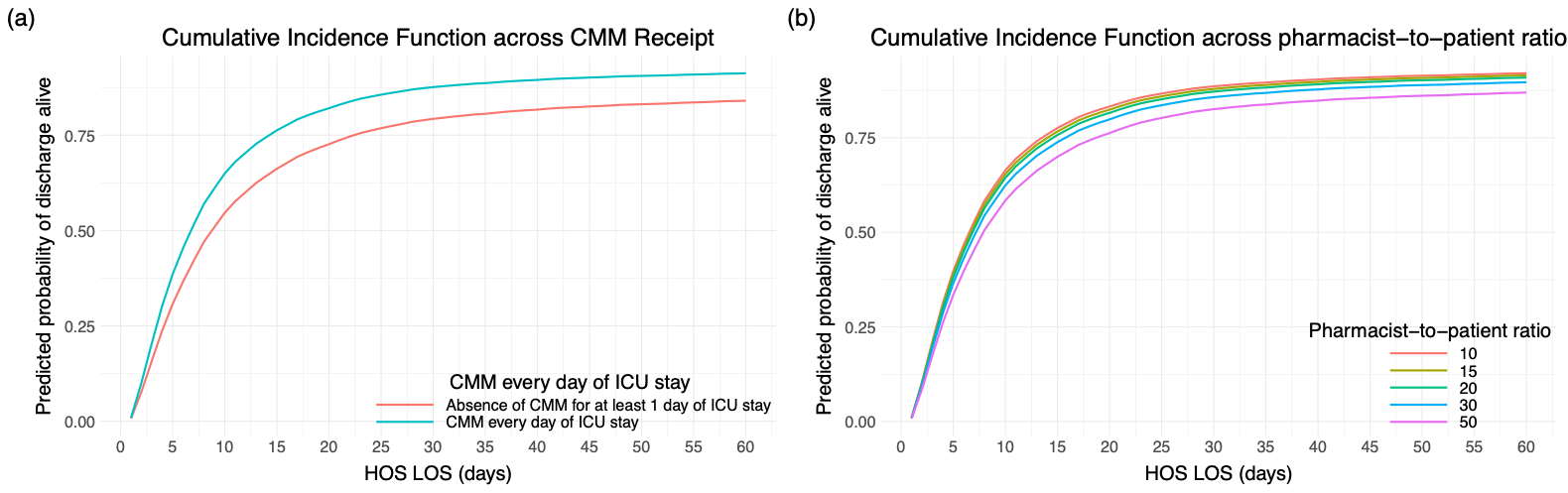

#### eFigure 31. Cumulative Incidence Function (CIF) for Probability of Discharge Alive across Pharmacist-to-patient Ratio (ICU patients) as Points

CIF curves were estimated using a typical patient profile constructed from the imputed dataset, with covariates fixed at representative values:

Hospital type: Academic Medical Center

Hospital CMS star rating: 3

SOFA Score: 4

MRC-ICU Score: 10

ICU pharmacist coverage 1^st^ 24 hours: CMM delivered on interprofessional rounds

ICU type: Medical

Nurse-to-patient ratio: 2

Pharmacist-to-patient ratio (ICU patients): 17

ICU admission day of the week: Monday

Percent of assigned ICU teams pharmacist rounded with: 50

Percent days, no pharmacist or pharmacist trainee: 28.57143

Percent days, only attending physician: 0

Age: 63.82

Sex: Male

The only variable varied was pharmacist-to-patient ratio

CCP-to-ICU-patient ratio: critical care pharmacist to ICU patient ratio, CMM: comprehensive medication management, CMS: Center for Medicare & Medicaid Services, ICU: intensive care unit, HOS LOS: hospital length of stay, MRC-ICU: medication regimen complexity-intensive care unit score, SOFA: sequential organ failure assessment

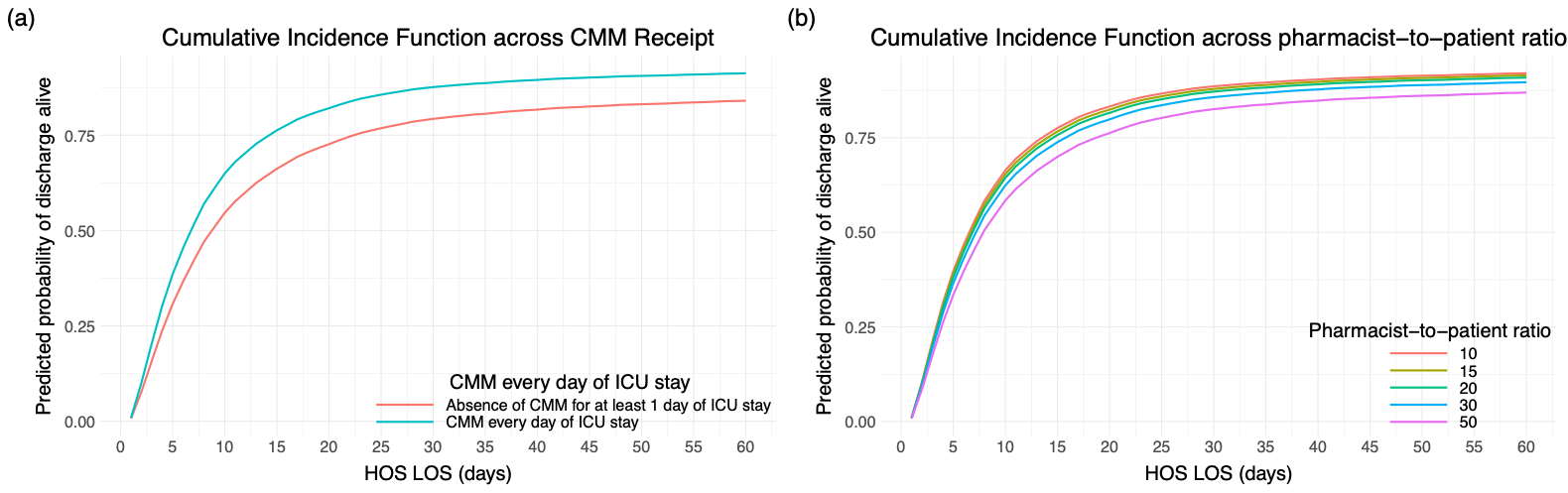

#### eFigure 32. Cumulative Incidence Function (CIF) for Probability of Discharge Alive across Pharmacist-to-patient Ratio (ICU patients) as Ranges

CIF curves were estimated using a typical patient profile constructed from the imputed dataset, with covariates fixed at representative values:

Hospital type: Academic Medical Center

Hospital CMS star rating: 3

SOFA Score: 4

MRC-ICU Score: 10

ICU pharmacist coverage 1^st^ 24 hours: CMM delivered on interprofessional rounds

ICU type: Medical

Nurse-to-patient ratio: 2

Pharmacist-to-patient ratio (ICU patients): 17

ICU admission day of the week: Monday

Percent of assigned ICU teams pharmacist rounded with: 50

Percent days, no pharmacist or pharmacist trainee: 28.57143

Percent days, only attending physician: 0

Age: 63.82

Sex: Male

The only variable varied was pharmacist-to-patient ratio

CCP-to-ICU-patient ratio: critical care pharmacist to ICU patient ratio, CMM: comprehensive medication management, CMS: Center for Medicare & Medicaid Services, ICU: intensive care unit, HOS LOS: hospital length of stay, MRC-ICU: medication regimen complexity-intensive care unit score, SOFA: sequential organ failure assessment

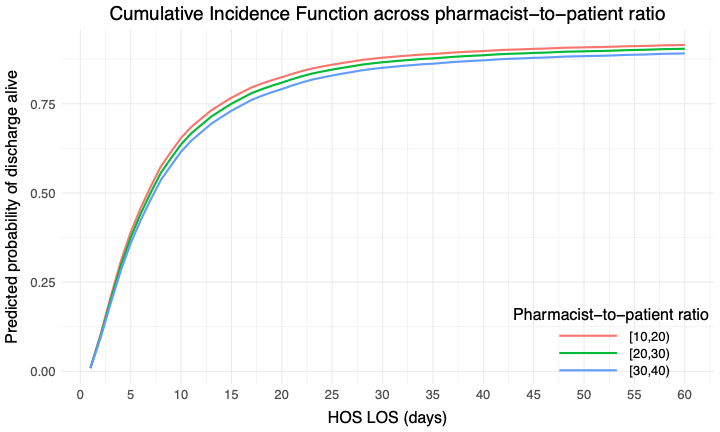

#### eFigure 33. Cumulative Incidence Function (CIF) for Probability of Discharge Alive across Pharmacist-to-patient Ratio (ICU patients) as Ranges at Number Needed to Treat Cutoff

CIF curves were estimated using a typical patient profile constructed from the imputed dataset, with covariates fixed at representative values:

Hospital type: Academic Medical Center

Hospital CMS star rating: 3

SOFA Score: 4

MRC-ICU Score: 10

ICU pharmacist coverage 1^st^ 24 hours: CMM delivered on interprofessional rounds

ICU type: Medical

Nurse-to-patient ratio: 2

Pharmacist-to-patient ratio (ICU patients): 17

ICU admission day of the week: Monday

Percent of assigned ICU teams pharmacist rounded with: 50

Percent days, no pharmacist or pharmacist trainee: 28.57143

Percent days, only attending physician: 0

Age: 63.82

Sex: Male

The only variable varied was pharmacist-to-patient ratio

CCP-to-ICU-patient ratio: critical care pharmacist to ICU patient ratio, CMM: comprehensive medication management, CMS: Center for Medicare & Medicaid Services, ICU: intensive care unit, HOS LOS: hospital length of stay, MRC-ICU: medication regimen complexity-intensive care unit score, SOFA: sequential organ failure assessment

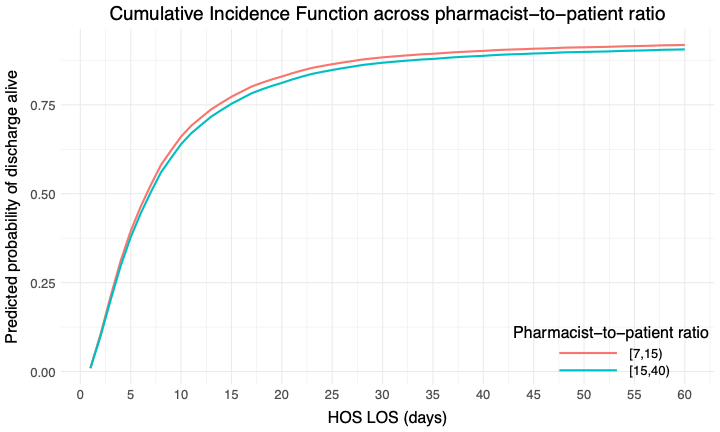

#### eFigure 34. Cumulative Incidence Function (CIF) for Probability of Discharge Alive across Pharmacist-to-patient Ratio (ICU patients) as Ranges Using 1:20 as the Cutoff

CIF curves were estimated using a typical patient profile constructed from the imputed dataset, with covariates fixed at representative values:

Hospital type: Academic Medical Center

Hospital CMS star rating: 3

SOFA Score: 4

MRC-ICU Score: 10

ICU pharmacist coverage 1^st^ 24 hours: CMM delivered on interprofessional rounds

ICU type: Medical

Nurse-to-patient ratio: 2

Pharmacist-to-patient ratio (ICU patients): 17

ICU admission day of the week: Monday

Percent of assigned ICU teams pharmacist rounded with: 50

Percent days, no pharmacist or pharmacist trainee: 28.57143

Percent days, only attending physician: 0

Age: 63.82

Sex: Male

The only variable varied was pharmacist-to-patient ratio

CCP-to-ICU-patient ratio: critical care pharmacist to ICU patient ratio, CMM: comprehensive medication management, CMS: Center for Medicare & Medicaid Services, ICU: intensive care unit, HOS LOS: hospital length of stay, MRC-ICU: medication regimen complexity-intensive care unit score, SOFA: sequential organ failure assessment

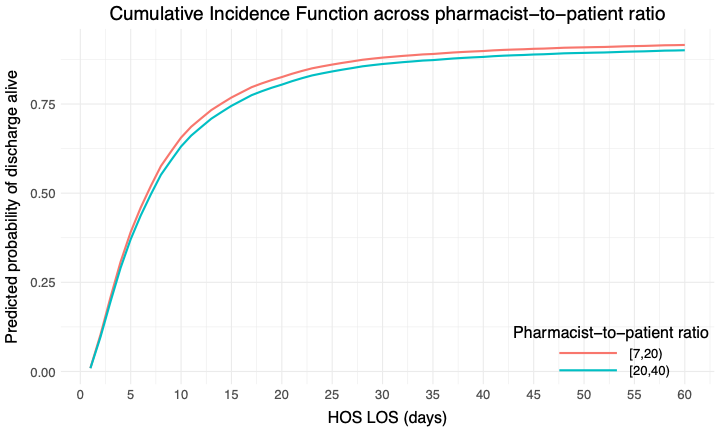

#### eFigure 35. 28-Day Cumulative Incidence Function (CIF) for Probability of Extubation Alive across CMM Every Day of ICU Admission

0- CMM every day of ICU admission
1- Absence of CMM for at least 1 day of ICU stay

CIF curves were estimated using a typical patient profile constructed from the imputed dataset, with covariates fixed at representative values:

Hospital type: Academic Medical Center

Hospital CMS star rating: 3

SOFA Score: 4

MRC-ICU Score: 10

ICU pharmacist coverage 1^st^ 24 hours: CMM delivered on interprofessional rounds

ICU type: Medical

Nurse-to-patient ratio: 2

Pharmacist-to-patient ratio (ICU patients): 17

ICU admission day of the week: Monday

Percent of assigned ICU teams pharmacist rounded with: 50

Percent days, no pharmacist or pharmacist trainee: 28.57143

Percent days, only attending physician: 0

Age: 63.82

Sex: Male

The only variable varied was pharmacist-to-patient ratio

CCP: critical care pharmacist, CMM: comprehensive medication management, CMS: Center for Medicare & Medicaid Services, ICU: intensive care unit, MRC-ICU: medication regimen complexity-intensive care unit score, SOFA: sequential organ failure assessment

The only variable varied was the presence of CMM every day of ICU stay

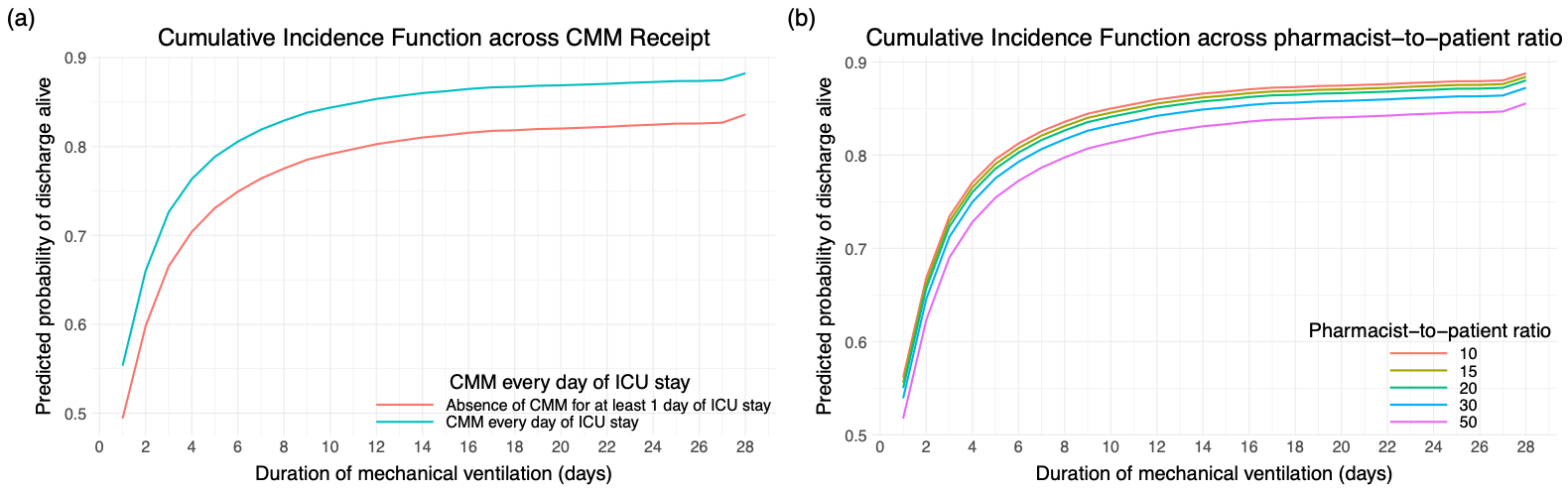

#### eFigure 36. 28-Day Cumulative Incidence Function (CIF) for Probability of Extubation Alive across Pharmacist-to-patient Ratio (ICU patients) as Points

CIF curves were estimated using a typical patient profile constructed from the imputed dataset, with covariates fixed at representative values:

Hospital type: Academic Medical Center

Hospital CMS star rating: 3

SOFA Score: 4

MRC-ICU Score: 10

ICU pharmacist coverage 1^st^ 24 hours: CMM delivered on interprofessional rounds

ICU type: Medical

Nurse-to-patient ratio: 2

Pharmacist-to-patient ratio (ICU patients): 17

ICU admission day of the week: Monday

Percent of assigned ICU teams pharmacist rounded with: 50

Percent days, no pharmacist or pharmacist trainee: 28.57143

Percent days, only attending physician: 0

Age: 63.82

Sex: Male

The only variable varied was pharmacist-to-patient ratio

CCP-to-ICU-patient ratio: critical care pharmacist to ICU patient ratio, CMM: comprehensive medication management, CMS: Center for Medicare & Medicaid Services, ICU: intensive care unit, MRC-ICU: medication regimen complexity-intensive care unit score, SOFA: sequential organ failure assessment

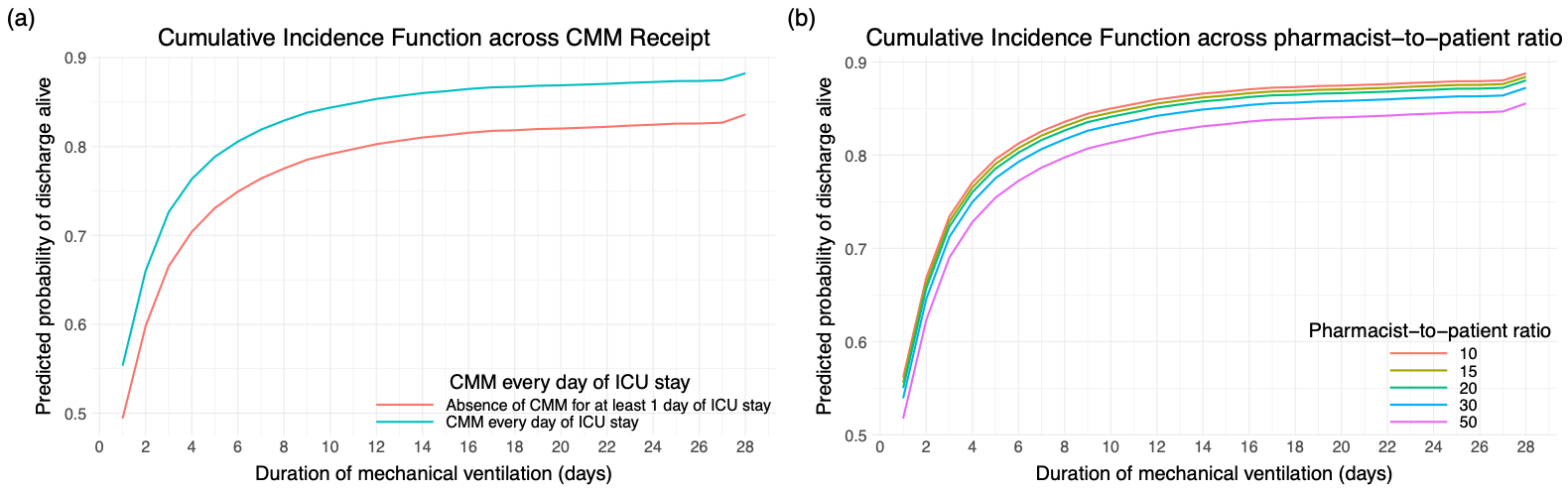

#### eFigure 37. 28-Day Cumulative Incidence Function (CIF) for Probability of Extubation Alive across Pharmacist-to-patient Ratio (ICU patients) as Ranges

CIF curves were estimated using a typical patient profile constructed from the imputed dataset, with covariates fixed at representative values:

Hospital type: Academic Medical Center

Hospital CMS star rating: 3

SOFA Score: 4

MRC-ICU Score: 10

ICU pharmacist coverage 1^st^ 24 hours: CMM delivered on interprofessional rounds

ICU type: Medical

Nurse-to-patient ratio: 2

Pharmacist-to-patient ratio (ICU patients): 17

ICU admission day of the week: Monday

Percent of assigned ICU teams pharmacist rounded with: 50

Percent days, no pharmacist or pharmacist trainee: 28.57143

Percent days, only attending physician: 0

Age: 63.82

Sex: Male

The only variable varied was pharmacist-to-patient ratio

CCP-to-ICU-patient ratio: critical care pharmacist to ICU patient ratio, CMM: comprehensive medication management, CMS: Center for Medicare & Medicaid Services, ICU: intensive care unit, MRC-ICU: medication regimen complexity-intensive care unit score, SOFA: sequential organ failure assessment

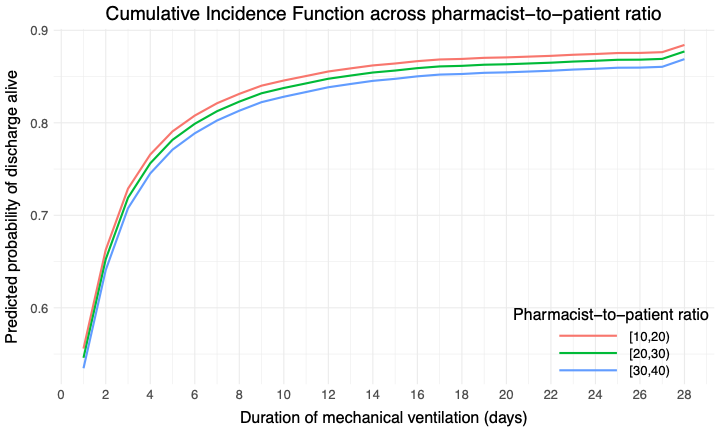

#### eFigure 38. 28-Day Cumulative Incidence Function (CIF) for Probability of Extubation Alive across Pharmacist-to-patient Ratio (ICU patients) as Ranges at Number Needed to Treat Cutoff

CIF curves were estimated using a typical patient profile constructed from the imputed dataset, with covariates fixed at representative values:

Hospital type: Academic Medical Center

Hospital CMS star rating: 3

SOFA Score: 4

MRC-ICU Score: 10

ICU pharmacist coverage 1^st^ 24 hours: CMM delivered on interprofessional rounds

ICU type: Medical

Nurse-to-patient ratio: 2

Pharmacist-to-patient ratio (ICU patients): 17

ICU admission day of the week: Monday

Percent of assigned ICU teams pharmacist rounded with: 50

Percent days, no pharmacist or pharmacist trainee: 28.57143

Percent days, only attending physician: 0

Age: 63.82

Sex: Male

The only variable varied was pharmacist-to-patient ratio

CCP-to-ICU-patient ratio: critical care pharmacist to ICU patient ratio, CMM: comprehensive medication management, CMS: Center for Medicare & Medicaid Services, ICU: intensive care unit, MRC-ICU: medication regimen complexity-intensive care unit score, SOFA: sequential organ failure assessment

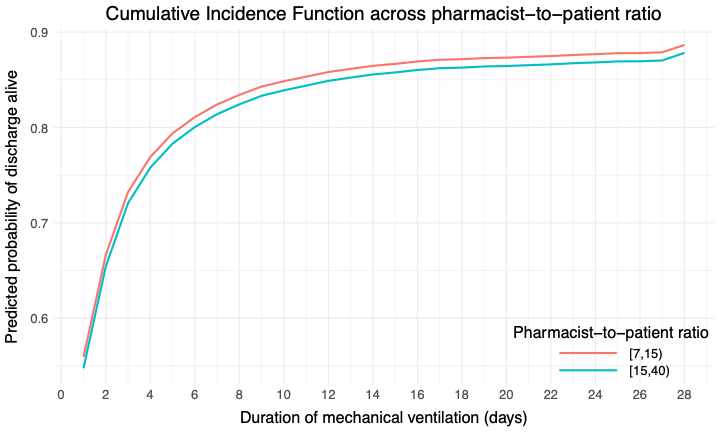

#### eFigure 39. 28-Day Cumulative Incidence Function (CIF) for Probability of Extubation Alive across Pharmacist-to-patient Ratio (ICU patients) as Ranges Using 1:20 as the Cutoff

CIF curves were estimated using a typical patient profile constructed from the imputed dataset, with covariates fixed at representative values:

Hospital type: Academic Medical Center

Hospital CMS star rating: 3

SOFA Score: 4

MRC-ICU Score: 10

ICU pharmacist coverage 1^st^ 24 hours: CMM delivered on interprofessional rounds

ICU type: Medical

Nurse-to-patient ratio: 2

Pharmacist-to-patient ratio (ICU patients): 17

ICU admission day of the week: Monday

Percent of assigned ICU teams pharmacist rounded with: 50

Percent days, no pharmacist or pharmacist trainee: 28.57143

Percent days, only attending physician: 0

Age: 63.82

Sex: Male

The only variable varied was pharmacist-to-patient ratio

CCP-to-ICU-patient ratio: critical care pharmacist to ICU patient ratio, CMM: comprehensive medication management, CMS: Center for Medicare & Medicaid Services, ICU: intensive care unit, MRC-ICU: medication regimen complexity-intensive care unit score, SOFA: sequential organ failure assessment

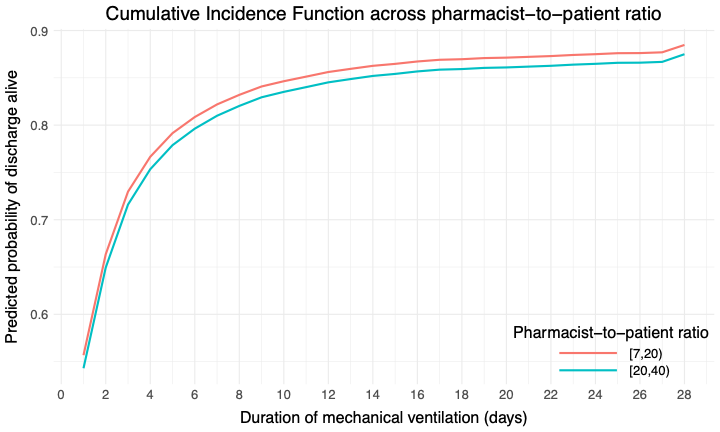

### Sensitivity Analysis- Academic Medical Center Patients (N=19243)

#### eTable 21. Patient demographics for Academic Medical Center Patients

|  | **Overall** | **Alive** | **Deceased** | **p-value** |
| --- | --- | --- | --- | --- |
|  | N = 19,243 | N = 16,375 | N = 2,868 |  |
| Percent of assigned ICU teams pharmacist rounded with§ | 60.2 (32.0) | 60.0 (32.2) | 61.7 (30.6) | 0.024 |
| Missing | 281 (1.5) | 248 (1.5) | 33 (1.2) |  |
| Percent days, no pharmacist or pharmacist trainee | 32.4 (34.0) | 32.5 (34.4) | 31.6 (31.6) | 0.7 |
| Missing | 782 (4.1) | 614 (3.7) | 168 (5.9) |  |
| Percent days, only attending physician⁂ | 0.8 (8.8) | 0.7 (7.9) | 1.8 (12.9) | <0.001 |
| Missing | 1,319 (6.9) | 1,067 (6.5) | 252 (8.8) |  |
| CMM every day of ICU stay |  |  |  | <0.001 |
| Yes | 15,250 (79.2) | 13,091 (79.9) | 2,159 (75.3) |  |
| No | 3,993 (20.8) | 3,284 (20.1) | 709 (24.7) |  |
| Age, years | 60.2 (17.1) | 59.5 (17.2) | 64.3 (15.7) | <0.001 |
| Sex |  |  |  | 0.5 |
| Male | 11,225 (58.3) | 9,534 (58.2) | 1,691 (59.0) |  |
| Female | 8,009 (41.6) | 6,832 (41.7) | 1,177 (41.0) |  |
| Unidentified | 8 (0.0) | 8 (0.0) | 0 (0.0) |  |
| Missing | 1 (<0.1) | 1 (<0.1) | 0 (0) |  |
| SOFA Score* | 5.3 (4.2) | 4.7 (3.8) | 8.6 (4.5) | <0.001 |
| Missing | 20 (0.1) | 16 (<0.1) | 4 (0.1) |  |
| Dialysis |  |  |  | <0.001 |
| No | 17,007 (88.4) | 14,929 (91.2) | 2,078 (72.5) |  |
| Yes | 2,236 (11.6) | 1,446 (8.8) | 790 (27.5) |  |
| ECMO |  |  |  | <0.001 |
| No | 18,997 (98.7) | 16,240 (99.2) | 2,757 (96.1) |  |
| Yes | 246 (1.3) | 135 (0.8) | 111 (3.9) |  |
| Mechanical circulatory support |  |  |  | <0.001 |
| No | 9,919 (51.5) | 9,246 (56.5) | 673 (23.5) |  |
| Yes | 9,324 (48.5) | 7,129 (43.5) | 2,195 (76.5) |  |
| Mechanical ventilation |  |  |  | <0.001 |
| No | 9,768 (50.8) | 9,045 (55.2) | 723 (25.2) |  |
| Yes | 9,475 (49.2) | 7,330 (44.8) | 2,145 (74.8) |  |
| MRC-ICU Score* | 11.6 (6.7) | 11.1 (6.5) | 14.6 (7.2) | <0.001 |
| Missing | 2 (<0.1) | 2 (<0.1) | 0 (0) |  |
| ICU admission day of the week |  |  |  | 0.009 |
| Monday | 3,086 (16.0) | 2,613 (16.0) | 473 (16.5) |  |
| Tuesday | 3,033 (15.8) | 2,613 (16.0) | 420 (14.6) |  |
| Wednesday | 2,905 (15.1) | 2,490 (15.2) | 415 (14.5) |  |
| Thursday | 2,975 (15.5) | 2,551 (15.6) | 424 (14.8) |  |
| Friday | 2,591 (13.5) | 2,225 (13.6) | 366 (12.8) |  |
| Saturday | 2,341 (12.2) | 1,943 (11.9) | 398 (13.9) |  |
| Sunday | 2,310 (12.0) | 1,938 (11.8) | 372 (13.0) |  |
| Missing | 2 (<0.1) | 2 (<0.1) | 0 (0) |  |
| Hospital LOS | 13.4 (18.9) | 13.0 (18.5) | 15.2 (21.3) | <0.001 |
| Missing | 3 (<0.1) | 3 (<0.1) | 0 (0) |  |
| ICU LOS | 6.1 (10.5) | 5.6 (9.6) | 9.2 (14.2) | <0.001 |
| ICU free days | 19.7 (9.6) | 23.0 (5.7) | 0.0 (0.0) | <0.001 |
| Missing | 200 (1.0) | 52 (0.3) | 148 (5.2) |  |
| Duration of mechanical ventilation | 2.3 (6.3) | 1.8 (5.8) | 5.1 (8.1) | <0.001 |
| Missing | 1 (<0.1) | 1 (<0.1) | 0 (0) |  |
| VFDs | 22.5 (10.1) | 26.4 (4.1) | 0.0 (0.0) | <0.001 |
| Missing | 1 (<0.1) | 1 (<0.1) | 0 (0) |  |
| ICU pharmacist coverage 1^st^ 24 hours |  |  |  | 0.04 |
| CMM delivered on interprofessional rounds | 12,995 (67.5) | 11,060 (67.6) | 1,935 (67.5) |  |
| CMM delivered not on interprofessional rounds | 2,694 (14.0) | 2,323 (14.2) | 371 (12.9) |  |
| Abbreviated CMM delivered | 1,710 (8.9) | 1,462 (8.9) | 248 (8.6) |  |
| CMM not delivered (absence of CMM) | 1,722 (9.0) | 1,426 (8.7) | 296 (10.3) |  |
| Unknown | 117 (0.6) | 99 (0.6) | 18 (0.6) |  |
| Missing | 5 (<0.1) | 5 (<0.1) | 0 (0) |  |
| ICU type† |  |  |  | <0.001 |
| Medical | 7,114 (37.0) | 5,642 (34.5) | 1,472 (51.3) |  |
| Surgical/Trauma | 3,286 (17.1) | 2,919 (17.8) | 367 (12.8) |  |
| Surgical | 1,975 (10.3) | 1,770 (10.8) | 205 (7.1) |  |
| Cardiothoracic | 2,099 (10.9) | 1,947 (11.9) | 152 (5.3) |  |
| Cardiac | 1,436 (7.5) | 1,210 (7.4) | 226 (7.9) |  |
| Neurosurgery/Neurology | 2,502 (13.0) | 2,188 (13.4) | 314 (10.9) |  |
| Pediatric | 35 (0.2) | 33 (0.2) | 2 (0.1) |  |
| Mixed Medical-Surgical | 612 (3.2) | 493 (3.0) | 119 (4.1) |  |
| Burn | 177 (0.9) | 166 (1.0) | 11 (0.4) |  |
| Other | 7 (0.0) | 7 (0.0) | 0 (0.0) |  |
| Hospital CMS star rating |  |  |  | 0.09 |
| 2 | 4,284 (23.1) | 3,706 (23.3) | 578 (21.8) |  |
| 3 | 9,116 (49.2) | 7,765 (48.9) | 1,351 (51.0) |  |
| 4 | 4,710 (25.4) | 4,040 (25.4) | 670 (25.3) |  |
| 5 | 424 (2.3) | 374 (2.4) | 50 (1.9) |  |
| Pharmacist-to-patient ratio (all patients) | 25.0 (15.2) | 25.1 (15.4) | 24.8 (14.6) | >0.9 |
| Missing | 251 (1.3) | 220 (1.3) | 31 (1.1) |  |
| Pharmacist-to-patient ratio (ICU patients) | 20.4 (11.0) | 20.4 (11.0) | 20.5 (11.1) | 0.3 |
| Missing | 251 (1.3) | 220 (1.3) | 31 (1.1) |  |
| Nurse-to-patient ratio |  |  |  | <0.001 |
| ≤1 | 1,615 (8.7) | 1,227 (7.8) | 388 (13.9) |  |
| =2 | 14,613 (79.1) | 12,691 (80.9) | 1,922 (68.9) |  |
| >2 | 241 (1.3) | 215 (1.4) | 26 (0.9) |  |
| 1-2 | 2,001 (10.8) | 1,548 (9.9) | 453 (16.2) |  |
| Missing | 773 (4.0) | 694 (4.2) | 79 (2.8) |  |

Data presented as mean (±SD) or n (%)

CMM: comprehensive medication management, CMS: Center for Medicare & Medicaid Services, ECMO: extracorporeal membrane oxygenation, ICU: intensive care unit, LOS: length of stay, MRC-ICU, medication regimen complexity- intensive care unit, SOFA: sequential organ failure assessment, VA: Veterans Affairs, VFDs: ventilator free days

*worst score during the first 24 hours of ICU stay

**†**Inclusion criteria specified ≥18 years of age; however, some institutions allowed patients ≥18 years of age in the pediatric ICU

**‡**CMM delivered on interprofessional rounds: the pharmacist attended multidisciplinary rounds and verbally provided CMM (including review of medications and recommendations); CMM delivered not on interprofessional rounds: the pharmacist provided by CMM (reviewed medications and provided recommendations) but did not attend interdisciplinary rounds; Abbreviated CMM delivered: the pharmacist provided abbreviated CMM which includes brief medication review but does not include full patient review (e.g. progress notes, results) and recommendations were provided outside of rounds; CMM not delivered (absence of CMM): the patient received no medication review from a pharmacist outside of pharmacokinetic monitoring and prospective medication order verification

**§**Percent of assigned ICU teams pharmacist rounded is calculated by total number of ICU medical teams the pharmacist attended and provided CMM on interprofessional rounds for divided by total number of ICU medical teams providing care for patients that the pharmacist was assigned to provide CMM for

**⁂**Only attending physician refers to composition of the primary medical team. On these days, the patient received care from only the attending physician and did not receive care from any medical residents, fellows, or advanced practice providers.

#### eTable 22. Univariable and Multivariable GEE for Mortality Based on Multiply Imputed Data (N = 19,243)

| **Mortality Model (N=19243)** | | | | |
| --- | --- | --- | --- | --- |
| **Variable** | **Univariable** | | **Multivariable** | |
|  | **OR (CI)** | **P-value** | **OR (CI)** | **P-value** |
| *Primary Variable* | | | | |
| Pharmacist-to-patient ratio (ICU patients) | 1.01 (1.00–1.01) | 0.06 | 1.01 (1.00–1.01) | 0.008 |
| *Secondary Variable* | | | | |
| Absence of CMM for at least 1 day of ICU stay | 1.49 (1.34–1.65) | <0.001 | 1.27 (1.08–1.49) | 0.004 |
| *Co-variates* | | | | |
| Age, years | 1.02 (1.01–1.02) | <0.001 | 1.02 (1.02–1.03) | <0.001 |
| Sex, female | 0.96 (0.88–1.05) | 0.4 | 1.00 (0.91–1.11) | 0.93 |
| SOFA Score* | 1.23 (1.21–1.26) | <0.001 | 1.29 (1.24–1.35) | <0.001 |
| MRC-ICU Score* | 1.08 (1.07–1.09) | <0.001 | 1.04 (1.02–1.06) | <0.001 |
| SOFA-MRC-ICU interaction term | 1.01 (1.01–1.01) | <0.001 | 1.00 (0.99–1.00) | 0.001 |
| ICU admission day of the week (reference: Monday) |  |  |  |  |
| Tuesday | 0.89 (0.77–1.02) | 0.09 | 0.92 (0.79–1.06) | 0.25 |
| Wednesday | 0.93 (0.81–1.07) | 0.31 | 0.89 (0.78–1.02) | 0.09 |
| Thursday | 0.91 (0.80–1.04) | 0.17 | 0.88 (0.76–1.01) | 0.06 |
| Friday | 0.92 (0.81–1.04) | 0.18 | 0.84 (0.73–0.95) | 0.007 |
| Saturday | 1.10 (0.98–1.25) | 0.11 | 0.95 (0.81–1.12) | 0.57 |
| Sunday | 1.03 (0.89–1.19) | 0.68 | 0.94 (0.80–1.11) | 0.49 |
| ICU type† (reference: medical) | - | - | - | - |
| Surgical/Trauma | 0.54 (0.44–0.66) | <0.001 | 0.73 (0.60–0.88) | 0.001 |
| Surgical | 0.43 (0.32–0.58) | <0.001 | 0.46 (0.33–0.65) | <0.001 |
| Cardiothoracic | 0.32 (0.22–0.46) | <0.001 | 0.19 (0.11–0.34) | <0.001 |
| Cardiac | 0.62 (0.48–0.80) | <0.001 | 0.77 (0.59–1.02) | 0.07 |
| Neurosurgery/Neurology | 0.56 (0.49–0.64) | <0.001 | 1.17 (0.97–1.40) | 0.1 |
| Pediatric | 0.22 (0.12–0.42) | <0.001 | 0.60 (0.27–1.33) | 0.21 |
| Mixed Medical-Surgical | 0.86 (0.59–1.24) | 0.42 | 0.82 (0.60–1.13) | 0.23 |
| Burn | 0.35 (0.22–0.54) | <0.001 | 0.58 (0.33–1.03) | 0.06 |
| Hospital CMS star rating (reference: 3) |  |  |  |  |
| 2 | 0.94 (0.71–1.23) | 0.65 | 1.11 (0.86–1.44) | 0.42 |
| 4 | 1.05 (0.72–1.53) | 0.8 | 0.73 (0.50–1.07) | 0.1 |
| 5 | 0.86 (0.43–1.71) | 0.66 | 0.52 (0.30–0.89) | 0.02 |
| ICU pharmacist coverage 1st 24 hours‡ (reference: CMM delivered on interprofessional rounds) | - | - | - | - |
| CMM delivered not on interprofessional rounds | 0.94 (0.84–1.07) | 0.35 | 1.06 (0.92–1.23) | 0.43 |
| Abbreviated CMM delivered | 1.04 (0.90–1.21) | 0.57 | 1.06 (0.89–1.27) | 0.49 |
| CMM not delivered (absence of CMM) | 1.15 (0.98–1.34) | 0.08 | 1.07 (0.89–1.30) | 0.48 |
| Unknown | 1.01 (0.57–1.79) | 0.98 | 0.89 (0.42–1.86) | 0.75 |
| Nurse-to-patient ratio (average)(reference: 1:2) | - | - | - | - |
| ≤1 | 3.07 (2.41–3.92) | <0.001 | 2.72 (1.77–4.17) | <0.001 |
| 1-2 | 2.37 (1.96–2.86) | <0.001 | 1.63 (1.22–2.19) | 0.001 |
| >2 | 0.82 (0.49–1.38) | 0.46 | 0.82 (0.49–1.35) | 0.44 |
| Percent of assigned ICU teams pharmacist rounded with§ | 1.00 (0.76–1.32) | 0.98 | 1.19 (0.90–1.57) | 0.22 |
| Percent days, no pharmacist or pharmacist trainee on rounds | 1.00 (1.00–1.00) | 0.7 | 1.00 (1.00–1.00) | 0.26 |
| Percent days, only attending physician⁂ | 1.00 (1.00–1.00) | 0.34 | 1.00 (0.99–1.00) | 0.26 |

CI: confidence interval, CMM: comprehensive medication management, CMS: Center for Medicare & Medicaid Services, ICU: intensive care unit, MRC-ICU: medication regimen complexity- intensive care unit, OR: odds ratio, SOFA: sequential organ failure assessment, VA: Veterans Affairs

*worst score during the first 24 hours of ICU stay

**†**Inclusion criteria specified ≥18 years of age; however, some institutions allowed patients ≥18 years of age in the pediatric ICU

**‡**CMM delivered on interprofessional rounds: the pharmacist attended multidisciplinary rounds and verbally provided CMM (including review of medications and recommendations); CMM delivered not on interprofessional rounds: the pharmacist provided by CMM (reviewed medications and provided recommendations) but did not attend interdisciplinary rounds; Abbreviated CMM delivered: the pharmacist provided abbreviated CMM which includes brief medication review but does not include full patient review (e.g. progress notes, results) and recommendations were provided outside of rounds; CMM not delivered (absence of CMM): the patient received no medication review from a pharmacist outside of pharmacokinetic monitoring and prospective medication order verification

**§**Percent of assigned ICU teams pharmacist rounded is calculated by total number of ICU medical teams the pharmacist attended and provided CMM on interprofessional rounds for divided by total number of ICU medical teams providing care for patients that the pharmacist was assigned to provide CMM for

**⁂**Only attending physician refers to composition of the primary medical team. On these days, the patient received care from only the attending physician and did not receive care from any medical residents, fellows, or advanced practice providers.

#### eTable 23. Fine-Gray Competing Risks Model for ICU and Hospital Length of Stay with In-ICU Mortality as a Competing Event (N = 19240)

| **Variable** | **Hospital Length of Stay**  **(N = 19240)** | | **ICU Length of Stay**  **(N = 19240)** | |
| --- | --- | --- | --- | --- |
|  | **Hazard Ratio (CI)** | **P-value** | **Variable** | **Hazard Ratio (CI)** |
| *Primary Variable* | | | | |
| Pharmacist-to-patient ratio (ICU patients) | 0.99 (0.99, 1.00) | <0.001 | 0.99 (0.98, 0.99) | <0.001 |
| *Secondary Variable* | | | | |
| Absence of CMM for at least 1 day of ICU stay | 0.76 (0.69, 0.84) | <0.001 | 0.63 (0.56, 0.70) | <0.001 |
| *Co-variates* | | | | |
| Age, years | 0.99 (0.99, 0.99) | <0.001 | 1.00 (1.00, 1.00) | 0.07 |
| Sex, female | 1.00 (0.97, 1.04) | 0.87 | 1.01 (0.98, 1.05) | 0.42 |
| SOFA Score* | 0.90 (0.89, 0.92) | <0.001 | 0.93 (0.92, 0.95) | <0.001 |
| MRC-ICU Score* | 0.98 (0.97, 0.98) | <0.001 | 0.98 (0.98, 0.99) | <0.001 |
| ICU admission day of the week (reference: Monday) | - | - | - | - |
| Tuesday | 1.04 (0.98, 1.10) | 0.19 | 1.02 (0.97, 1.07) | 0.51 |
| Wednesday | 1.02 (0.97, 1.07) | 0.48 | 1.02 (0.98, 1.07) | 0.31 |
| Thursday | 1.04 (0.99, 1.10) | 0.15 | 1.01 (0.96, 1.07) | 0.72 |
| Friday | 1.00 (0.93, 1.07) | 0.97 | 0.93 (0.87, 1.01) | 0.07 |
| Saturday | 0.99 (0.92, 1.07) | 0.88 | 0.93 (0.85, 1.01) | 0.07 |
| Sunday | 0.96 (0.89, 1.03) | 0.28 | 0.97 (0.91, 1.04) | 0.38 |
| ICU type† (reference: medical) | - | - | - | - |
| Surgical/Trauma | 1.07 (0.89, 1.28) | 0.49 | 0.94 (0.80, 1.10) | 0.45 |
| Surgical | 1.08 (0.90, 1.30) | 0.41 | 1.13 (0.93, 1.37) | 0.22 |
| Cardiothoracic | 1.70 (1.37, 2.13) | <0.001 | 1.22 (0.95, 1.55) | 0.12 |
| Cardiac | 1.20 (0.99, 1.46) | 0.06 | 0.92 (0.74, 1.15) | 0.48 |
| Neurosurgery/Neurology | 0.96 (0.83, 1.11) | 0.55 | 0.79 (0.66, 0.94) | 0.008 |
| Pediatric | 1.20 (0.89, 1.63) | 0.23 | 1.05 (0.81, 1.37) | 0.69 |
| Mixed Medical-Surgical | 0.90 (0.78, 1.04) | 0.16 | 1.24 (0.99, 1.57) | 0.06 |
| Burn | 0.84 (0.56, 1.27) | 0.42 | 0.59 (0.46, 0.74) | <0.001 |
| Other | 5.25 (4.19, 6.58) | <0.001 | 1.64 (1.42, 1.90) | <0.001 |
| Hospital CMS star rating (reference: 3) |  |  |  |  |
| 2 | 0.91 (0.80, 1.05) | 0.19 | 1.19 (1.04, 1.38) | 0.01 |
| 4 | 0.81 (0.71, 0.93) | 0.002 | 1.23 (1.06, 1.43) | 0.006 |
| 5 | 0.82 (0.66, 1.01) | 0.06 | 1.41 (0.91, 2.17) | 0.12 |
| ICU pharmacist coverage 1st 24 hours‡ (reference: CMM delivered on interprofessional rounds) | - | - | - | - |
| CMM delivered not on interprofessional rounds | 0.94 (0.87, 1.01) | 0.1 | 0.98 (0.91, 1.04) | 0.44 |
| Abbreviated CMM delivered | 0.95 (0.87, 1.05) | 0.34 | 0.99 (0.90, 1.10) | 0.92 |
| CMM not delivered (absence of CMM) | 1.08 (0.99, 1.19) | 0.08 | 1.11 (1.02, 1.22) | 0.02 |
| Unknown | 0.85 (0.55, 1.29) | 0.44 | 0.63 (0.48, 0.83) | <0.001 |
| Nurse-to-patient ratio (average)(reference: 1:2) | - | - | - | - |
| ≤1 | 0.75 (0.63, 0.90) | 0.002 | 0.84 (0.56, 1.27) | 0.41 |
| 1-2 | 0.73 (0.67, 0.81) | <0.001 | 0.69 (0.63, 0.74) | <0.001 |
| >2 | 1.06 (0.92, 1.23) | 0.40 | 1.18 (0.80, 1.74) | 0.42 |
| Percent of assigned ICU teams pharmacist rounded with§ | 0.88 (0.74, 1.04) | 0.14 | 0.85 (0.71, 1.02) | 0.07 |
| Percent days, no pharmacist or pharmacist trainee on rounds | 1.00 (1.00, 1.00) | 0.85 | 1.00 (1.00, 1.00) | 0.04 |
| Percent days, only attending physician⁂ | 1.00 (1.00, 1.00) | 0.57 | 0.99 (0.99, 1.00) | <0.001 |

CI: confidence interval, CMM: comprehensive medication management, CMS: Center for Medicare & Medicaid Services, ICU: intensive care unit, MRC-ICU: medication regimen complexity- intensive care unit, OR: odds ratio, SOFA: sequential organ failure assessment, VA: Veterans Affairs

*worst score during the first 24 hours of ICU stay

**†**Inclusion criteria specified ≥18 years of age; however, some institutions allowed patients ≥18 years of age in the pediatric ICU

**‡**CMM delivered on interprofessional rounds: the pharmacist attended multidisciplinary rounds and verbally provided CMM (including review of medications and recommendations); CMM delivered not on interprofessional rounds: the pharmacist provided by CMM (reviewed medications and provided recommendations) but did not attend interdisciplinary rounds; Abbreviated CMM delivered: the pharmacist provided abbreviated CMM which includes brief medication review but does not include full patient review (e.g. progress notes, results) and recommendations were provided outside of rounds; CMM not delivered (absence of CMM): the patient received no medication review from a pharmacist outside of pharmacokinetic monitoring and prospective medication order verification

**§**Percent of assigned ICU teams pharmacist rounded is calculated by total number of ICU medical teams the pharmacist attended and provided CMM on interprofessional rounds for divided by total number of ICU medical teams providing care for patients that the pharmacist was assigned to provide CMM for

**⁂**Only attending physician refers to composition of the primary medical team. On these days, the patient received care from only the attending physician and did not receive care from any medical residents, fellows, or advanced practice providers.

#### eTable 24. Fine-Gray Competing Risks Model for VFD with In-Hospital Mortality as a Competing Event and 28 days as a cut (N = 19240).

| **VFD Competing Model without cut (N = 19240)** | | |
| --- | --- | --- |
| **Variable** | **Hazard Ratio (CI)** | **P-value** |
| *Primary Variable* | | |
| Pharmacist-to-patient ratio (ICU patients) | 0.99 (0.99, 1.00) | <0.001 |
| *Secondary Variable* | | |
| Absence of CMM for at least 1 day of ICU stay | 0.82 (0.76, 0.88) | <0.001 |
| *Covariates* | | |
| Age, years | 1.00 (0.99, 1.00) | <0.001 |
| Sex, female | 1.00 (0.98, 1.03) | 0.77 |
| SOFA Score* | 0.91 (0.90, 0.92) | <0.001 |
| MRC-ICU Score* | 0.97 (0.97, 0.98) | <0.001 |
| ICU admission day of the week (reference: Monday) | - | - |
| Tuesday | 1.01 (0.96, 1.06) | 0.71 |
| Wednesday | 1.01 (0.98, 1.05) | 0.49 |
| Thursday | 1.04 (0.99, 1.09) | 0.14 |
| Friday | 1.01 (0.96, 1.07) | 0.65 |
| Saturday | 0.96 (0.90, 1.02) | 0.19 |
| Sunday | 0.95 (0.90, 1.02) | 0.14 |
| ICU type† (reference: medical ICU) | - | - |
| Other | 1.49 (1.41, 1.58) | <0.001 |
| Surgical | 1.31 (1.17, 1.48) | <0.001 |
| Surgical/Trauma | 1.07 (0.99, 1.15) | 0.09 |
| Cardiothoracic | 1.72 (1.50, 1.97) | <0.001 |
| Cardiac | 1.15 (1.05, 1.26) | 0.002 |
| Neurosurgery/Neurology | 0.94 (0.88, 1.00) | 0.04 |
| Pediatric | 1.15 (0.98, 1.35) | 0.09 |
| Mixed Medical-Surgical | 1.09 (0.96, 1.24) | 0.17 |
| Burn | 1.08 (0.86, 1.36) | 0.51 |
| Hospital CMS star rating (reference: 3) | - | - |
| 2 | 1.07 (1.01, 1.14) | 0.03 |
| 4 | 1.08 (1.00, 1.15) | 0.04 |
| 5 | 1.11 (0.92, 1.34) | 0.29 |
| ICU pharmacist coverage 1st 24 hours‡ (reference: CMM delivered on interprofessional rounds) | - | - |
| CMM delivered not on interprofessional rounds | 0.97 (0.92, 1.02) | 0.26 |
| Abbreviated CMM delivered | 1.00 (0.94, 1.07) | 0.90 |
| CMM not delivered (absence of CMM) | 1.05 (0.98, 1.13) | 0.19 |
| Unknown | 0.82 (0.69, 0.97) | 0.02 |
| Nurse-to-patient ratio (average)(reference: 1:2) | - | - |
| ≤1 | 0.82 (0.65, 1.03) | 0.08 |
| 1-2 | 0.79 (0.73, 0.85) | <0.001 |
| >2 | 1.11 (0.96, 1.29) | 0.16 |
| Percent of assigned ICU teams pharmacist rounded with§ | 0.89 (0.82, 0.96) | 0.004 |
| Percent days, no pharmacist or pharmacist trainee on rounds | 1.00 (1.00, 1.00) | 0.07 |
| Percent days, only attending physician⁂ | 1.00 (1.00, 1.00) | 0.21 |

CI: confidence interval, CMM: comprehensive medication management, CMS: Center for Medicare & Medicaid Services, ICU: intensive care unit, MRC-ICU: medication regimen complexity- intensive care unit, OR: odds ratio, SOFA: sequential organ failure assessment, VA: Veterans Affairs

*worst score during the first 24 hours of ICU stay

**†**Inclusion criteria specified ≥18 years of age; however, some institutions allowed patients ≥18 years of age in the pediatric ICU

**‡**CMM delivered on interprofessional rounds: the pharmacist attended multidisciplinary rounds and verbally provided CMM (including review of medications and recommendations); CMM delivered not on interprofessional rounds: the pharmacist provided by CMM (reviewed medications and provided recommendations) but did not attend interdisciplinary rounds; Abbreviated CMM delivered: the pharmacist provided abbreviated CMM which includes brief medication review but does not include full patient review (e.g. progress notes, results) and recommendations were provided outside of rounds; CMM not delivered (absence of CMM): the patient received no medication review from a pharmacist outside of pharmacokinetic monitoring and prospective medication order verification

**§**Percent of assigned ICU teams pharmacist rounded is calculated by total number of ICU medical teams the pharmacist attended and provided CMM on interprofessional rounds for divided by total number of ICU medical teams providing care for patients that the pharmacist was assigned to provide CMM for

**⁂**Only attending physician refers to composition of the primary medical team. On these days, the patient received care from only the attending physician and did not receive care from any medical residents, fellows, or advanced practice providers.

#### eTable 25. Fine-Gray Competing Risks Model for VFD with In-Hospital Mortality as a Competing Event without a cut (N = 19240).

| **VFD Competing Model with cut (N = 19240)** | | |
| --- | --- | --- |
| **Variable** | **Hazard Ratio (CI)** | **P-value** |
| *Primary Variable* | | |
| Pharmacist-to-patient ratio (ICU patients) | 1.00 (0.99, 1.00) | <0.001 |
| *Secondary Variable* | | |
| Absence of CMM for at least 1 day of ICU stay | 0.82 (0.76, 0.88) | <0.001 |
| *Covariates* | | |
| Age, years | 1.00 (0.99, 1.00) | <0.001 |
| Sex, female | 1.00 (0.98, 1.03) | 0.77 |
| SOFA Score* | 0.91 (0.90, 0.92) | <0.001 |
| MRC-ICU Score* | 0.97 (0.97, 0.98) | <0.001 |
| ICU admission day of the week (reference: Monday) | - | - |
| Tuesday | 1.01 (0.96, 1.06) | 0.71 |
| Wednesday | 1.01 (0.98, 1.05) | 0.49 |
| Thursday | 1.04 (0.99, 1.09) | 0.14 |
| Friday | 1.01 (0.96, 1.07) | 0.65 |
| Saturday | 0.96 (0.90, 1.02) | 0.19 |
| Sunday | 0.95 (0.90, 1.02) | 0.14 |
| ICU type† (reference: medical ICU) | - | - |
| Other | 1.49 (1.41, 1.58) | <0.001 |
| Surgical | 1.31 (1.17, 1.48) | <0.001 |
| Surgical/Trauma | 1.07 (0.99, 1.15) | 0.09 |
| Cardiothoracic | 1.72 (1.50, 1.97) | <0.001 |
| Cardiac | 1.15 (1.05, 1.26) | 0.002 |
| Neurosurgery/Neurology | 0.94 (0.88, 1.00) | 0.04 |
| Pediatric | 1.15 (0.98, 1.35) | 0.09 |
| Mixed Medical-Surgical | 1.09 (0.96, 1.24) | 0.17 |
| Burn | 1.08 (0.86, 1.36) | 0.51 |
| Hospital CMS star rating (reference: 3) | - | - |
| 2 | 1.07 (1.01, 1.14) | 0.03 |
| 4 | 1.08 (1.00, 1.15) | 0.04 |
| 5 | 1.11 (0.92, 1.34) | 0.29 |
| ICU pharmacist coverage 1st 24 hours‡ (reference: CMM delivered on interprofessional rounds) | - | - |
| CMM delivered not on interprofessional rounds | 0.97 (0.92, 1.02) | 0.26 |
| Abbreviated CMM delivered | 1.00 (0.94, 1.07) | 0.9 |
| CMM not delivered (absence of CMM) | 1.05 (0.98, 1.13) | 0.19 |
| Unknown | 0.82 (0.69, 0.97) | 0.02 |
| Nurse-to-patient ratio (average)(reference: 1:2) | - | - |
| ≤1 | 0.82 (0.65, 1.03) | 0.08 |
| 1-2 | 0.79 (0.73, 0.85) | <0.001 |
| >2 | 1.11 (0.96, 1.29) | 0.16 |
| Percent of assigned ICU teams pharmacist rounded with§ | 0.89 (0.82, 0.96) | 0.004 |
| Percent days, no pharmacist or pharmacist trainee on rounds | 1.00 (1.00, 1.00) | 0.07 |
| Percent days, only attending physician⁂ | 1.00 (1.00, 1.00) | 0.21 |

CI: confidence interval, CMM: comprehensive medication management, CMS: Center for Medicare & Medicare Services, ICU: intensive care unit, MRC-ICU: medication regimen complexity- intensive care unit, OR: odds ratio, SOFA: sequential organ failure assessment, VA: Veterans Affairs

*worst score during the first 24 hours of ICU stay

**†**Inclusion criteria specified ≥18 years of age; however, some institutions allowed patients ≥18 years of age in the pediatric ICU

**‡**CMM delivered on interprofessional rounds: the pharmacist attended multidisciplinary rounds and verbally provided CMM (including review of medications and recommendations); CMM delivered not on interprofessional rounds: the pharmacist provided by CMM (reviewed medications and provided recommendations) but did not attend interdisciplinary rounds; Abbreviated CMM delivered: the pharmacist provided abbreviated CMM which includes brief medication review but does not include full patient review (e.g. progress notes, results) and recommendations were provided outside of rounds; CMM not delivered (absence of CMM): the patient received no medication review from a pharmacist outside of pharmacokinetic monitoring and prospective medication order verification

**§**Percent of assigned ICU teams pharmacist rounded is calculated by total number of ICU medical teams the pharmacist attended and provided CMM on interprofessional rounds for divided by total number of ICU medical teams providing care for patients that the pharmacist was assigned to provide CMM for

**⁂**Only attending physician refers to composition of the primary medical team. On these days, the patient received care from only the attending physician and did not receive care from any medical residents, fellows, or advanced practice providers.

#### eFigure 40. Cumulative Incidence Function (CIF) for Probability of Discharge Alive across CMM Every Day of ICU Admission

0- CMM every day of ICU admission
1- Absence of CMM for at least 1 day of ICU stay

CIF curves were estimated using a typical patient profile constructed from the imputed dataset, with covariates fixed at representative values:

Hospital type: Academic Medical Center

Hospital CMS star rating: 3

SOFA Score: 4

MRC-ICU Score: 10

ICU pharmacist coverage 1^st^ 24 hours: CMM delivered on interprofessional rounds

ICU type: Medical

Nurse-to-patient ratio: 2

Pharmacist-to-patient ratio (ICU patients): 17

ICU admission day of the week: Monday

Percent of assigned ICU teams pharmacist rounded with: 50

Percent days, no pharmacist or pharmacist trainee: 28.57143

Percent days, only attending physician: 0

Age: 63.82

Sex: Male

CCP: critical care pharmacist, CMM: comprehensive medication management, CMS: Center for Medicare & Medicaid Services, ICU: intensive care unit, MRC-ICU: medication regimen complexity-intensive care unit score, SOFA: sequential organ failure assessment

The only variable varied was the presence of CMM every day of ICU stay

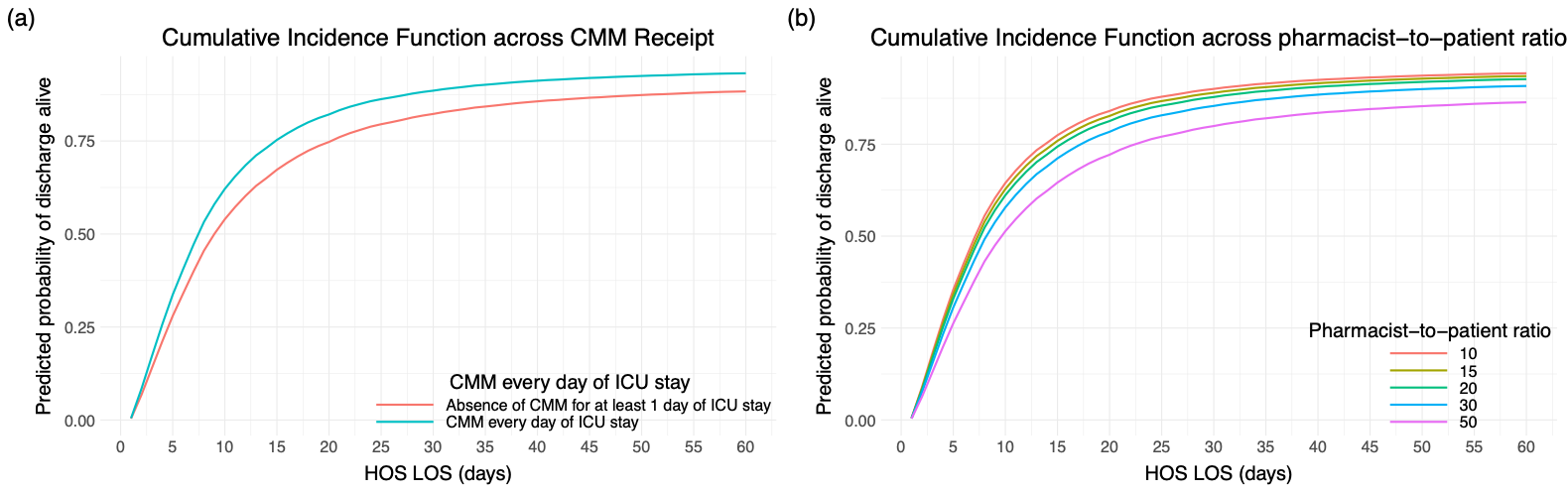

#### eFigure 41. Cumulative Incidence Function (CIF) for Probability of Discharge Alive across Pharmacist-to-patient Ratio (ICU patients) as Points

CIF curves were estimated using a typical patient profile constructed from the imputed dataset, with covariates fixed at representative values:

Hospital type: Academic Medical Center

Hospital CMS star rating: 3

SOFA Score: 4

MRC-ICU Score: 10

ICU pharmacist coverage 1^st^ 24 hours: CMM delivered on interprofessional rounds

ICU type: Medical

Nurse-to-patient ratio: 2

Pharmacist-to-patient ratio (ICU patients): 17

ICU admission day of the week: Monday

Percent of assigned ICU teams pharmacist rounded with: 50

Percent days, no pharmacist or pharmacist trainee: 28.57143

Percent days, only attending physician: 0

Age: 63.82

Sex: Male

The only variable varied was pharmacist-to-patient ratio

CCP-to-ICU-patient ratio: critical care pharmacist to ICU patient ratio, CMM: comprehensive medication management, CMS: Center for Medicare & Medicaid Services, ICU: intensive care unit, ICU LOS: ICU length of stay, MRC-ICU: medication regimen complexity-intensive care unit score, SOFA: sequential organ failure assessment

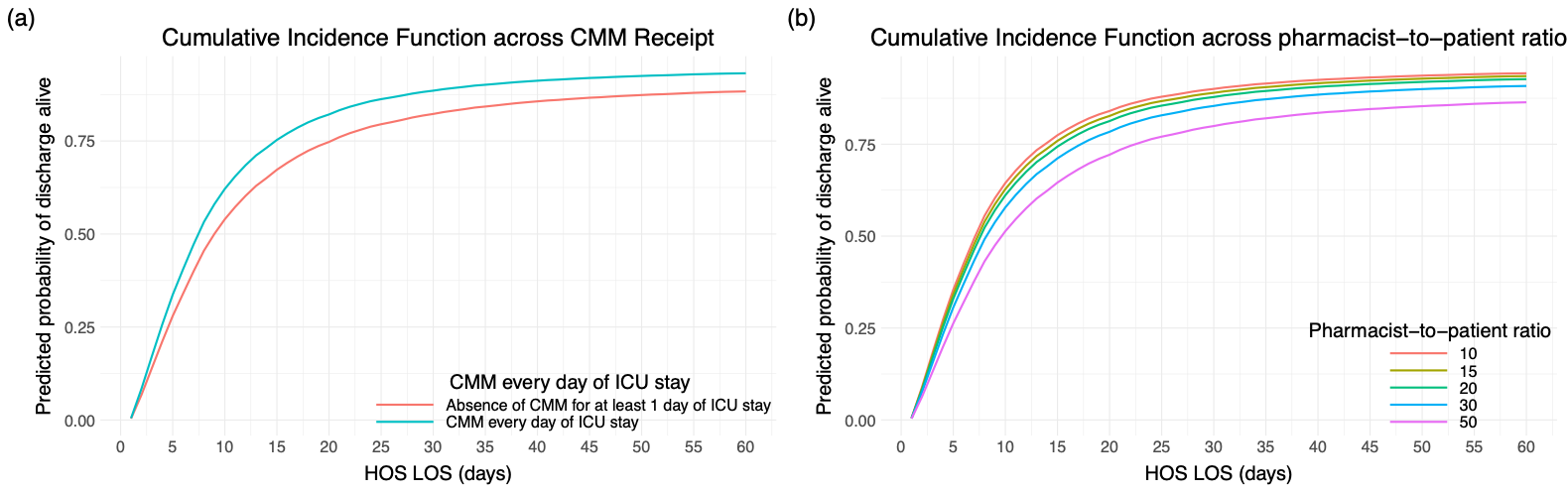

#### eFigure 42. Cumulative Incidence Function (CIF) for Probability of Discharge Alive across Pharmacist-to-patient Ratio (ICU patients) as Ranges

CIF curves were estimated using a typical patient profile constructed from the imputed dataset, with covariates fixed at representative values:

Hospital type: Academic Medical Center

Hospital CMS star rating: 3

SOFA Score: 4

MRC-ICU Score: 10

ICU pharmacist coverage 1^st^ 24 hours: CMM delivered on interprofessional rounds

ICU type: Medical

Nurse-to-patient ratio: 2

Pharmacist-to-patient ratio (ICU patients): 17

ICU admission day of the week: Monday

Percent of assigned ICU teams pharmacist rounded with: 50

Percent days, no pharmacist or pharmacist trainee: 28.57143

Percent days, only attending physician: 0

Age: 63.82

Sex: Male

The only variable varied was pharmacist-to-patient ratio

CCP-to-ICU-patient ratio: critical care pharmacist to ICU patient ratio, CMM: comprehensive medication management, CMS: Center for Medicare & Medicaid Services, ICU: intensive care unit, HOS LOS: Hospital length of stay, MRC-ICU: medication regimen complexity-intensive care unit score, SOFA: sequential organ failure assessment

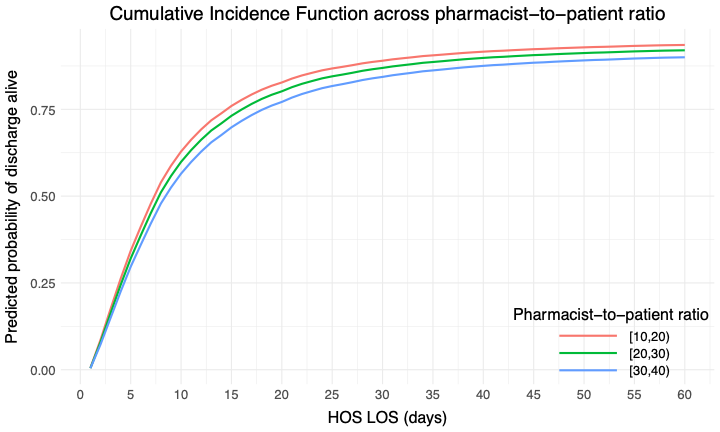

#### eFigure 43. Cumulative Incidence Function (CIF) for Probability of Discharge Alive across Pharmacist-to-patient Ratio (ICU patients) as Ranges at Number Needed to Treat Cutoff

CIF curves were estimated using a typical patient profile constructed from the imputed dataset, with covariates fixed at representative values:

Hospital type: Academic Medical Center

Hospital CMS star rating: 3

SOFA Score: 4

MRC-ICU Score: 10

ICU pharmacist coverage 1^st^ 24 hours: CMM delivered on interprofessional rounds

ICU type: Medical

Nurse-to-patient ratio: 2

Pharmacist-to-patient ratio (ICU patients): 17

ICU admission day of the week: Monday

Percent of assigned ICU teams pharmacist rounded with: 50

Percent days, no pharmacist or pharmacist trainee: 28.57143

Percent days, only attending physician: 0

Age: 63.82

Sex: Male

The only variable varied was pharmacist-to-patient ratio

CCP-to-ICU-patient ratio: critical care pharmacist to ICU patient ratio, CMM: comprehensive medication management, CMS: Center for Medicare & Medicaid Services, ICU: intensive care unit, HOS LOS: Hospital length of stay, MRC-ICU: medication regimen complexity-intensive care unit score, SOFA: sequential organ failure assessment

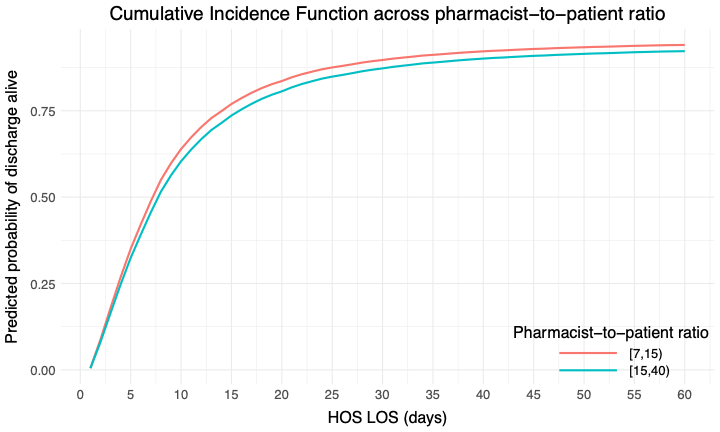

#### eFigure 44. Cumulative Incidence Function (CIF) for Probability of Discharge Alive across Pharmacist-to-patient Ratio (ICU patients) as Ranges Using 1:20 as the Cutoff

CIF curves were estimated using a typical patient profile constructed from the imputed dataset, with covariates fixed at representative values:

Hospital type: Academic Medical Center

Hospital CMS star rating: 3

SOFA Score: 4

MRC-ICU Score: 10

ICU pharmacist coverage 1^st^ 24 hours: CMM delivered on interprofessional rounds

ICU type: Medical

Nurse-to-patient ratio: 2

Pharmacist-to-patient ratio (ICU patients): 17

ICU admission day of the week: Monday

Percent of assigned ICU teams pharmacist rounded with: 50

Percent days, no pharmacist or pharmacist trainee: 28.57143

Percent days, only attending physician: 0

Age: 63.82

Sex: Male

The only variable varied was pharmacist-to-patient ratio

CCP-to-ICU-patient ratio: critical care pharmacist to ICU patient ratio, CMM: comprehensive medication management, CMS: Center for Medicare & Medicaid Services, ICU: intensive care unit, HOS LOS: Hospital length of stay, MRC-ICU: medication regimen complexity-intensive care unit score, SOFA: sequential organ failure assessment

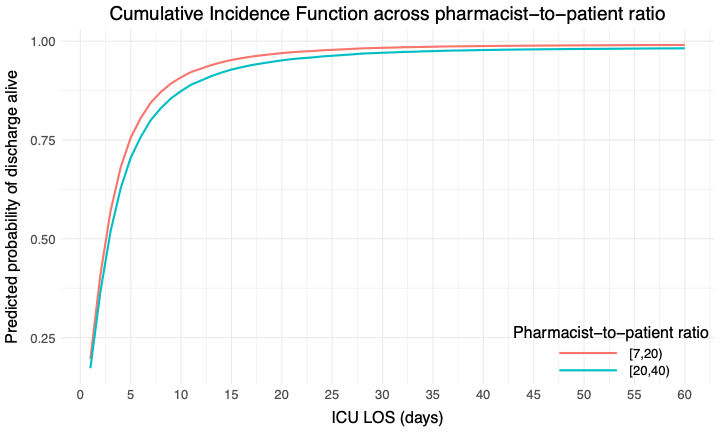

#### eFigure 45. 28-Day Cumulative Incidence Function (CIF) for Probability of Extubation Alive across CMM Every Day of ICU Admission

0- CMM every day of ICU admission
1- Absence of CMM for at least 1 day of ICU stay

CIF curves were estimated using a typical patient profile constructed from the imputed dataset, with covariates fixed at representative values:

Hospital type: Academic Medical Center

Hospital CMS star rating: 3

SOFA Score: 4

MRC-ICU Score: 10

ICU pharmacist coverage 1^st^ 24 hours: CMM delivered on interprofessional rounds

ICU type: Medical

Nurse-to-patient ratio: 2

Pharmacist-to-patient ratio (ICU patients): 17

ICU admission day of the week: Monday

Percent of assigned ICU teams pharmacist rounded with: 50

Percent days, no pharmacist or pharmacist trainee: 28.57143

Percent days, only attending physician: 0

Age: 63.82

Sex: Male

The only variable varied was presence of CMM every day of ICU stay

CCP: critical care pharmacist, CMM: comprehensive medication management, CMS: Center for Medicare & Medicaid Services, ICU: intensive care unit, MRC-ICU: medication regimen complexity-intensive care unit score, SOFA: sequential organ failure assessment

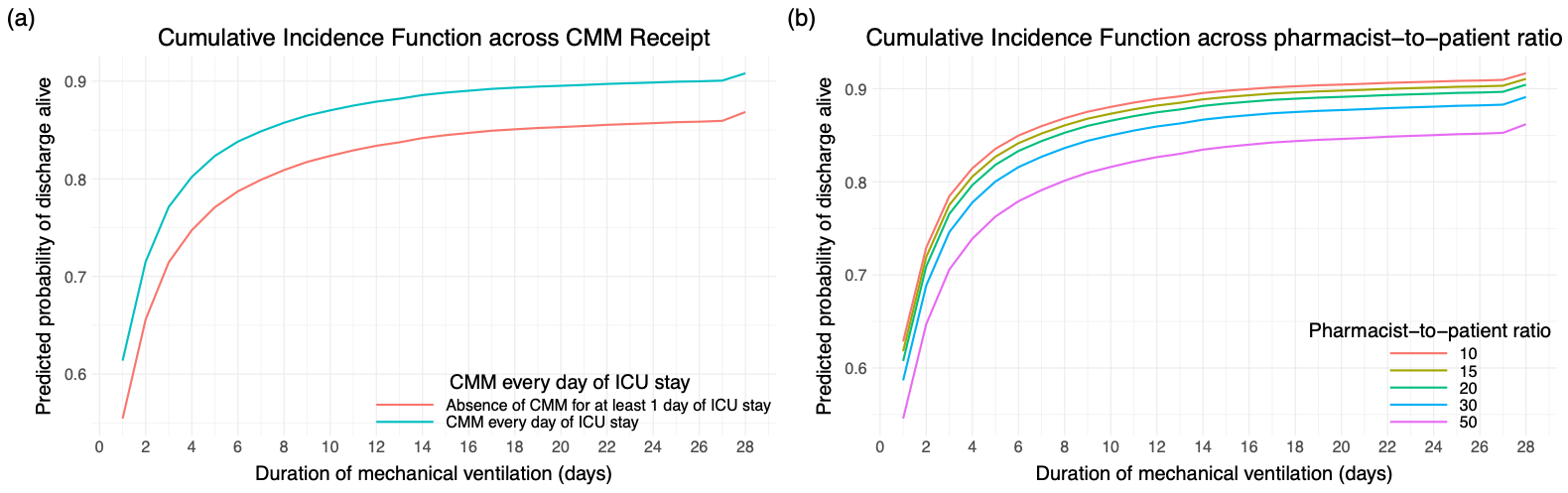

#### eFigure 46. 28-Day Cumulative Incidence Function (CIF) for Probability of Extubation Alive across Pharmacist-to-patient Ratio (ICU patients) as Points

CIF curves were estimated using a typical patient profile constructed from the imputed dataset, with covariates fixed at representative values:

Hospital type: Academic Medical Center

Hospital CMS star rating: 3

SOFA Score: 4

MRC-ICU Score: 10

ICU pharmacist coverage 1^st^ 24 hours: CMM delivered on interprofessional rounds

ICU type: Medical

Nurse-to-patient ratio: 2

Pharmacist-to-patient ratio (ICU patients): 17

ICU admission day of the week: Monday

Percent of assigned ICU teams pharmacist rounded with: 50

Percent days, no pharmacist or pharmacist trainee: 28.57143

Percent days, only attending physician: 0

Age: 63.82

Sex: Male

The only variable varied was pharmacist-to-patient ratio

CCP-to-ICU-patient ratio: critical care pharmacist to ICU patient ratio, CMM: comprehensive medication management, CMS: Center for Medicare & Medicaid Services, ICU: intensive care unit, MRC-ICU: medication regimen complexity-intensive care unit score, SOFA: sequential organ failure assessment

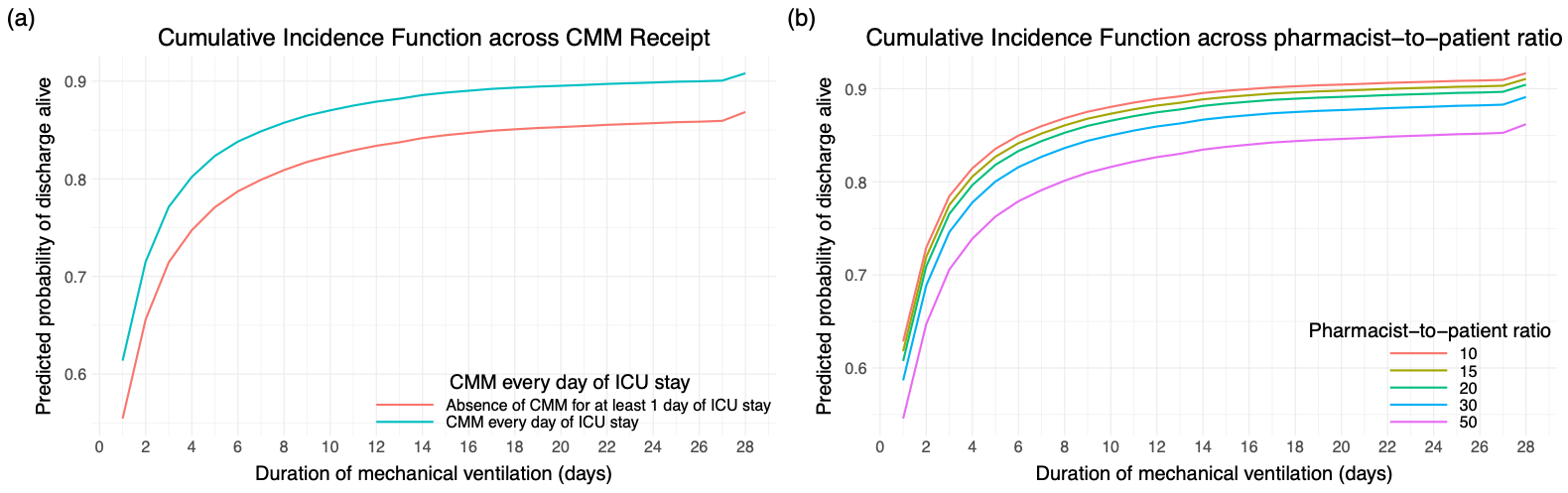

#### eFigure 47. 28-Day Cumulative Incidence Function (CIF) for Probability of Extubation Alive across Pharmacist-to-patient Ratio (ICU patients) as Ranges

CIF curves were estimated using a typical patient profile constructed from the imputed dataset, with covariates fixed at representative values:

Hospital type: Academic Medical Center

Hospital CMS star rating: 3

SOFA Score: 4

MRC-ICU Score: 10

ICU pharmacist coverage 1^st^ 24 hours: CMM delivered on interprofessional rounds

ICU type: Medical

Nurse-to-patient ratio: 2

Pharmacist-to-patient ratio (ICU patients): 17

ICU admission day of the week: Monday

Percent of assigned ICU teams pharmacist rounded with: 50

Percent days, no pharmacist or pharmacist trainee: 28.57143

Percent days, only attending physician: 0

Age: 63.82

Sex: Male

The only variable varied was pharmacist-to-patient ratio

CCP-to-ICU-patient ratio: critical care pharmacist to ICU patient ratio, CMM: comprehensive medication management, CMS: Center for Medicare & Medicaid Services, ICU: intensive care unit, MRC-ICU: medication regimen complexity-intensive care unit score, SOFA: sequential organ failure assessment

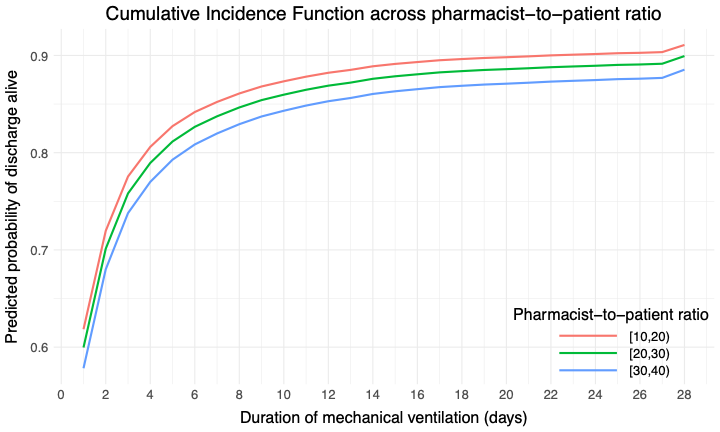

#### eFigure 48. 28-Day Cumulative Incidence Function (CIF) for Probability of Extubation Alive across Pharmacist-to-patient Ratio (ICU patients) as Ranges at Number Needed to Treat Cutoff

CIF curves were estimated using a typical patient profile constructed from the imputed dataset, with covariates fixed at representative values:

Hospital type: Academic Medical Center

Hospital CMS star rating: 3

SOFA Score: 4

MRC-ICU Score: 10

ICU pharmacist coverage 1^st^ 24 hours: CMM delivered on interprofessional rounds

ICU type: Medical

Nurse-to-patient ratio: 2

Pharmacist-to-patient ratio (ICU patients): 17

ICU admission day of the week: Monday

Percent of assigned ICU teams pharmacist rounded with: 50

Percent days, no pharmacist or pharmacist trainee: 28.57143

Percent days, only attending physician: 0

Age: 63.82

Sex: Male

The only variable varied was pharmacist-to-patient ratio

CCP-to-ICU-patient ratio: critical care pharmacist to ICU patient ratio, CMM: comprehensive medication management, CMS: Center for Medicare & Medicaid Services, ICU: intensive care unit, MRC-ICU: medication regimen complexity-intensive care unit score, SOFA: sequential organ failure assessment

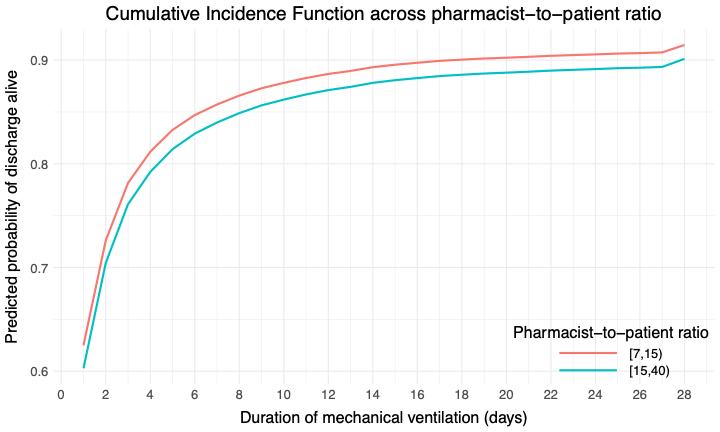

#### eFigure 49. 28-Day Cumulative Incidence Function (CIF) for Probability of Extubation Alive across Pharmacist-to-patient Ratio (ICU patients) as Ranges Using 1:20 as the Cutoff

CIF curves were estimated using a typical patient profile constructed from the imputed dataset, with covariates fixed at representative values:

Hospital type: Academic Medical Center

Hospital CMS star rating: 3

SOFA Score: 4

MRC-ICU Score: 10

ICU pharmacist coverage 1^st^ 24 hours: CMM delivered on interprofessional rounds

ICU type: Medical

Nurse-to-patient ratio: 2

Pharmacist-to-patient ratio (ICU patients): 17

ICU admission day of the week: Monday

Percent of assigned ICU teams pharmacist rounded with: 50

Percent days, no pharmacist or pharmacist trainee: 28.57143

Percent days, only attending physician: 0

Age: 63.82

Sex: Male

The only variable varied was pharmacist-to-patient ratio

CCP-to-ICU-patient ratio: critical care pharmacist to ICU patient ratio, CMM: comprehensive medication management, CMS: Center for Medicare & Medicaid Services, ICU: intensive care unit, MRC-ICU: medication regimen complexity-intensive care unit score, SOFA: sequential organ failure assessment

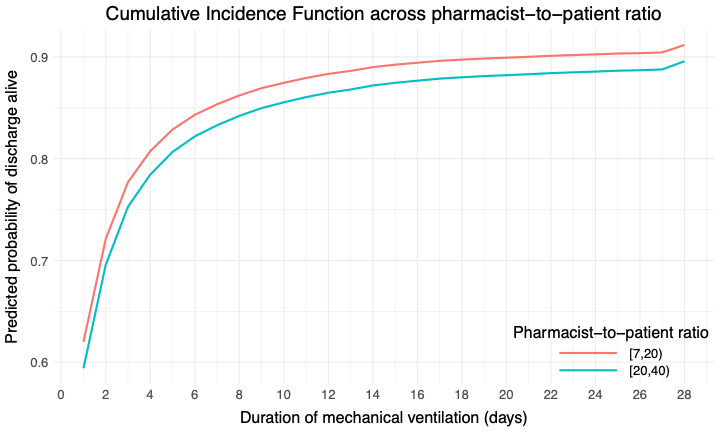

#### eTable 26. Odds Ratios for Categorical Pharmacist-to-patient ratio for mortality for Academic Medical Center Patients

| **CCP-to-ICU-patient ratio** | **Adjusted OR (95% CI)** |
| --- | --- |
| 15.1-46 compared to 7-15 | 1.29 (1.14–1.46) |
| 15.1-40 compared to 4-15 | 1.28 (1.13–1.45) |
| 15.1-40 compared to 10-15 | 1.29 (1.14–1.45) |

### Sensitivity Analysis- First Day Analysis

#### eTable 27. Patient Demographics for First Day Analysis

|  | **Overall** | **Alive** | **Deceased** | **p-value** |
| --- | --- | --- | --- | --- |
|  | **N = 28,795** | **N = 24,571** | **N = 4,224** |  |
| Percent of assigned ICU teams pharmacist rounded with**§*** | 62.0 (36.5) | 61.6 (36.5) | 64.0 (36.0) | <0.001 |
| Missing | 2,636 (9.2) | 2,243 (9.1) | 393 (9.3) |  |
| Percent days, no pharmacist or pharmacist trainee on rounds | 32.2 (34.1) | 32.4 (34.6) | 31.1 (30.7) | 0.6 |
| Missing | 1,045 (3.6) | 818 (3.3) | 227 (5.4) |  |
| Percent days, only attending physician**⁂** | 8.9 (26.7) | 8.9 (26.8) | 8.7 (25.9) | 0.3 |
| Missing | 1,768 (6.1) | 1,412 (5.7) | 356 (8.4) |  |
| Any days with only a physician |  |  |  | 0.046 |
| No | 25,165 (88.3) | 21,551 (88.5) | 3,614 (87.4) |  |
| Yes | 3,321 (11.7) | 2,801 (11.5) | 520 (12.6) |  |
| Missing | 309 (1.1) | 219 (0.9) | 90 (2.1) |  |
| CMM delivered every day of ICU stay |  |  |  | <0.001 |
| Yes | 21,416 (74.4) | 18,459 (75.1) | 2,957 (70.0) |  |
| No | 7,379 (25.6) | 6,112 (24.9) | 1,267 (30.0) |  |
| Age, years | 61.23 (16.98) | 60.49 (17.12) | 65.49 (15.49) | <0.001 |
| Sex | 61.2 (17.0) | 60.5 (17.1) | 65.5 (15.5) | <0.001 |
| Male |  |  |  | 0.5 |
| Female | 16,640 (57.8) | 14,186 (57.7) | 2,454 (58.1) |  |
| Unidentified | 12,144 (42.2) | 10,374 (42.2) | 1,770 (41.9) |  |
| Missing | 10 (0.0) | 10 (0.0) | 0 (0.0) |  |
| SOFA Score** | 1 (<0.1) | 1 (<0.1) | 0 (0) |  |
| Missing | 5.2 (4.1) | 4.6 (3.8) | 8.7 (4.4) | <0.001 |
| Dialysis |  |  |  | <0.001 |
| No | 25,419 (88.3) | 22,358 (91.0) | 3,061 (72.5) |  |
| Yes | 3,371 (11.7) | 2,210 (9.0) | 1,161 (27.5) |  |
| Missing | 5 (<0.1) | 3 (<0.1) | 2 (<0.1) |  |
| ECMO |  |  |  | <0.001 |
| No | 28,468 (98.9) | 24,389 (99.3) | 4,079 (96.6) |  |
| Yes | 324 (1.1) | 180 (0.7) | 144 (3.4) |  |
| Missing | 3 (<0.1) | 2 (<0.1) | 1 (<0.1) |  |
| Mechanical circulatory support |  |  |  | <0.001 |
| No | 14,813 (51.5) | 13,898 (56.6) | 915 (21.7) |  |
| Yes | 13,972 (48.5) | 10,665 (43.4) | 3,307 (78.3) |  |
| Missing | 10 (<0.1) | 8 (<0.1) | 2 (<0.1) |  |
| Mechanical ventilation |  |  |  | <0.001 |
| No | 14,643 (50.9) | 13,602 (55.4) | 1,041 (24.6) |  |
| Yes | 14,150 (49.1) | 10,967 (44.6) | 3,183 (75.4) |  |
| Missing | 2 (<0.1) | 2 (<0.1) | 0 (0) |  |
| MRC-ICU Score** | 11.4 (6.6) | 10.9 (6.4) | 14.6 (7.1) | <0.001 |
| Missing | 4 (<0.1) | 4 (<0.1) | 0 (0) |  |
| ICU admission day of the week |  |  |  | 0.01 |
| Monday | 4,634 (16.1) | 3,965 (16.1) | 669 (15.8) |  |
| Tuesday | 4,599 (16.0) | 3,959 (16.1) | 640 (15.2) |  |
| Wednesday | 4,364 (15.2) | 3,749 (15.3) | 615 (14.6) |  |
| Thursday | 4,386 (15.2) | 3,763 (15.3) | 623 (14.7) |  |
| Friday | 3,852 (13.4) | 3,294 (13.4) | 558 (13.2) |  |
| Saturday | 3,408 (11.8) | 2,845 (11.6) | 563 (13.3) |  |
| Sunday | 3,549 (12.3) | 2,993 (12.2) | 556 (13.2) |  |
| Missing | 3 (<0.1) | 3 (<0.1) | 0 (0) |  |
| Hospital LOS | 12.3 (17.0) | 12.0 (16.5) | 14.1 (19.6) | <0.001 |
| Missing | 4 (<0.1) | 4 (<0.1) | 0 (0) |  |
| ICU LOS | 5.8 (9.6) | 5.3 (8.8) | 8.7 (12.8) | <0.001 |
| ICU free days | 19.8 (9.5) | 23.1 (5.5) | 0.0 (0.0) | <0.001 |
| Missing | 245 (0.9) | 58 (0.2) | 187 (4.4) |  |
| Duration of mechanical ventilation | 2.4 (18.3) | 1.9 (19.4) | 5.1 (8.0) | <0.001 |
| Missing | 4 (<0.1) | 4 (<0.1) | 0 (0) |  |
| VFDs | 22.5 (10.1) | 26.4 (4.1) | 0.0 (0.0) | <0.001 |
| Missing | 4 (<0.1) | 4 (<0.1) | 0 (0) |  |
| Hospital Type |  |  |  | 0.02 |
| Academic medical center | 19,243 (66.8) | 16,375 (66.6) | 2,868 (67.9) |  |
| Community - Teaching | 7,006 (24.3) | 5,972 (24.3) | 1,034 (24.5) |  |
| Community - Non-teaching | 2,398 (8.3) | 2,092 (8.5) | 306 (7.2) |  |
| Government/VA/Military | 148 (0.5) | 132 (0.5) | 16 (0.4) |  |
| Hospital CMS star rating |  |  |  | 0.2 |
| 2 | 6,345 (22.6) | 5,475 (22.7) | 870 (21.7) |  |
| 3 | 14,958 (53.3) | 12,807 (53.2) | 2,151 (53.7) |  |
| 4 | 6,354 (22.6) | 5,423 (22.5) | 931 (23.3) |  |
| 5 | 424 (1.5) | 374 (1.6) | 50 (1.2) |  |
| Missing | 714 (2.5) | 492 (2.0) | 222 (5.3) |  |
| ICU type† * |  |  |  | <0.001 |
| Medical | 8,856 (30.8) | 7,100 (28.9) | 1,756 (41.6) |  |
| Surgical/Trauma | 3,923 (13.6) | 3,503 (14.3) | 420 (9.9) |  |
| Cardiothoracic | 2,574 (8.9) | 2,384 (9.7) | 190 (4.5) |  |
| Cardiac | 2,539 (8.8) | 2,119 (8.6) | 420 (9.9) |  |
| Neurosurgery/Neurology | 3,882 (13.5) | 3,420 (13.9) | 462 (10.9) |  |
| Pediatric | 35 (0.1) | 33 (0.1) | 2 (0.0) |  |
| Mixed Medical-Surgical | 4,192 (14.6) | 3,532 (14.4) | 660 (15.6) |  |
| Burn | 187 (0.6) | 175 (0.7) | 12 (0.3) |  |
| Other | 41 (0.1) | 33 (0.1) | 8 (0.2) |  |
| Surgical | 2,563 (8.9) | 2,269 (9.2) | 294 (7.0) |  |
| Missing | 3 (<0.1) | 3 (<0.1) | 0 (0) |  |
| ICU pharmacist coverage 1^st^ 24 hours* |  |  |  | 0.005 |
| CMM delivered on interprofessional rounds | 19,547 (67.9) | 16,662 (67.8) | 2,885 (68.3) |  |
| CMM delivered not on interprofessional rounds | 3,995 (13.9) | 3,468 (14.1) | 527 (12.5) |  |
| Abbreviated CMM delivered | 2,087 (7.2) | 1,791 (7.3) | 296 (7.0) |  |
| CMM not delivered (absence of CMM) | 3,016 (10.5) | 2,527 (10.3) | 489 (11.6) |  |
| Unknown | 142 (0.5) | 116 (0.5) | 26 (0.6) |  |
| Missing | 8 (<0.1%) | 7 (<0.1%) | 1 (<0.1%) |  |
| Pharmacist-to-patient ratio (all patients)* | 27.4 (31.3) | 27.4 (31.1) | 27.0 (32.8) | 0.001 |
| Missing | 2,613 (9.1) | 2,223 (9.0) | 390 (9.2) |  |
| Pharmacist-to-patient ratio (ICU patients)* | 19.5 (13.8) | 19.4 (13.6) | 19.7 (15.0) | 0.3 |
| Missing | 2,618 (9.1) | 2,226 (9.1) | 392 (9.3) |  |
| Nurse-to-patient ratio* |  |  |  | <0.001 |
| ≤1 | 1,785 (6.4) | 1,324 (5.6) | 461 (11.2) |  |
| =2 | 22,388 (80.5) | 19,510 (82.3) | 2,878 (69.8) |  |
| >2 | 674 (2.4) | 587 (2.5) | 87 (2.1) |  |
| 1-2 | 2,970 (10.7) | 2,274 (9.6) | 696 (16.9) |  |
| Missing | 978 (3.4) | 876 (3.6) | 102 (2.4) |  |
| Pharmacy Trainee Present* |  |  |  | 0.7 |
| No | 16,980 (60.4) | 14,467 (60.4) | 2,513 (60.6) |  |
| Yes | 11,132 (39.6) | 9,500 (39.6) | 1,632 (39.4) |  |
| Missing | 683 (2.4) | 604 (2.5) | 79 (1.9) |  |
| Medical Team Coverage* |  |  |  | <0.001 |
| Attending Physician Only | 2,512 (8.9) | 2,142 (8.9) | 370 (9.0) |  |
| Attending Physician and APP | 6,499 (23.1) | 5,662 (23.5) | 837 (20.3) |  |
| Attending Physician and Medical Residents/Fellows | 9,625 (34.2) | 8,097 (33.7) | 1,528 (37.1) |  |
| Attending Physician and Medical Residents/Fellows and APP | 9,526 (33.8) | 8,142 (33.9) | 1,384 (33.6) |  |
| Missing | 633 (2.2) | 528 (2.1) | 105 (2.5) |  |

*indicates that this is reflective of only the value on the first day of ICU stay

Data presented as mean (±SD) or n (%)

APP: advanced practice provider, CMM: comprehensive medication management, CMS: Center for Medicare & Medicaid Services, ECMO: extracorporeal membrane oxygenation, ICU: intensive care unit, LOS: length of stay, MRC-ICU, medication regimen complexity- intensive care unit, SOFA: sequential organ failure assessment, VA: Veterans Affairs, VFDs: ventilator free days

**worst score during the first 24 hours of ICU stay

**†**Inclusion criteria specified ≥18 years of age; however, some institutions allowed patients ≥18 years of age in the pediatric ICU

**‡**CMM delivered on interprofessional rounds: the pharmacist attended multidisciplinary rounds and verbally provided CMM (including review of medications and recommendations); CMM delivered not on interprofessional rounds: the pharmacist provided by CMM (reviewed medications and provided recommendations) but did not attend interdisciplinary rounds; Abbreviated CMM delivered: the pharmacist provided abbreviated CMM which includes brief medication review but does not include full patient review (e.g. progress notes, results) and recommendations were provided outside of rounds; CMM not delivered (absence of CMM): the patient received no medication review from a pharmacist outside of pharmacokinetic monitoring and prospective medication order verification

**§**Percent of assigned ICU teams pharmacist rounded is calculated by total number of ICU medical teams the pharmacist attended and provided CMM on interprofessional rounds for divided by total number of ICU medical teams providing care for patients that the pharmacist was assigned to provide CMM for

#### eTable 28. Univariable and Multivariable GEE for Mortality in First Day Analysis Based on Multiply Imputed Data (N = 28,795)

| **Mortality model (N=28,795)** |  | |  | |
| --- | --- | --- | --- | --- |
| **Variable** | Univariable | | Multivariable | |
|  | OR (CI) | P-value | OR (CI) | P-value |
| *Primary Variable* | | | | |
| Pharmacist-to-patient ratio* (ICU patients) | 1.00 (1.00–1.01) | 0.26 | 1.00 (1.00–1.01) | 0.10 |
| *Co-variates* | | | | |
| Age, years | 1.02 (1.02–1.02) | <0.001 | 1.03 (1.02–1.03) | <0.001 |
| Sex, female | 0.98 (0.91–1.05) | 0.58 | 1.02 (0.94–1.12) | 0.59 |
| SOFA Score** | 1.25 (1.23–1.27) | <0.001 | 1.31 (1.26–1.35) | <0.001 |
| MRC-ICU Score** | 1.09 (1.08–1.10) | <0.001 | 1.04 (1.03–1.06) | <0.001 |
| SOFA-MRC-ICU interaction term | 1.01 (1.01–1.01) | <0.001 | 1.00 (1.00–1.00) | <0.001 |
| ICU admission day of the week (reference: Monday) | - | - | - | - |
| Tuesday | 0.96 (0.85–1.07) | 0.45 | 0.98 (0.87–1.11) | 0.78 |
| Wednesday | 0.97 (0.86–1.09) | 0.62 | 0.95 (0.84–1.07) | 0.38 |
| Thursday | 0.97 (0.86–1.10) | 0.68 | 0.99 (0.87–1.12) | 0.84 |
| Friday | 1.01 (0.89–1.14) | 0.90 | 0.94 (0.82–1.06) | 0.32 |
| Saturday | 1.15 (1.01–1.31) | 0.04 | 1.02 (0.88–1.18) | 0.77 |
| Sunday | 1.08 (0.95–1.22) | 0.23 | 1.02 (0.89–1.17) | 0.80 |
| Hospital type (reference: academic medical center) | - | - | - | - |
| Community - Teaching | 0.98 (0.76–1.26) | 0.88 | 0.93 (0.75–1.14) | 0.47 |
| Community – nonteaching | 0.89 (0.64–1.24) | 0.49 | 0.79 (0.63–0.99) | 0.04 |
| Government/VA/Military | 0.68 (0.58–0.80) | <0.001 | 0.90 (0.70–1.15) | 0.39 |
| Hospital CMS star rating (reference: 3) |  |  |  |  |
| 2 | 0.92 (0.74–1.15) | 0.47 | 1.03 (0.85–1.23) | 0.79 |
| 4 | 1.06 (0.80–1.40) | 0.67 | 0.93 (0.71–1.22) | 0.62 |
| 5 | 0.87 (0.45–1.69) | 0.68 | 0.64 (0.40–1.04) | 0.07 |
| Pharmacy trainee present* | 0.98 (0.91–1.05) | 0.54 | 0.99 (0.91–1.07) | 0.71 |
| Medical team coverage* (reference: only attending physician) | - | - | - | - |
| Attending physician and APP | 1.01 (0.81–1.26) | 0.92 | 0.86 (0.70–1.05) | 0.14 |
| Attending physician and medical residents/fellows | 1.24 (0.95–1.63) | 0.11 | 0.92 (0.72–1.19) | 0.53 |
| Attending physician and medical residents/fellows and APP | 1.08 (0.81–1.43) | 0.59 | 0.93 (0.73–1.20) | 0.59 |
| ICU type† * (reference: medical) | - | - | - | - |
| Surgical/Trauma | 0.54 (0.45–0.64) | <0.001 | 0.76 (0.64–0.91) | 0.003 |
| Surgical | 0.52 (0.41–0.67) | <0.001 | 0.53 (0.41–0.68) | <0.001 |
| Cardiothoracic | 0.36 (0.25–0.50) | <0.001 | 0.24 (0.15–0.39) | <0.001 |
| Cardiac | 0.77 (0.64–0.93) | 0.007 | 0.98 (0.82–1.18) | 0.84 |
| Neurosurgery/Neurology | 0.51 (0.44–0.61) | <0.001 | 1.10 (0.92–1.32) | 0.31 |
| Pediatric | 0.23 (0.12–0.46) | <0.001 | 0.93 (0.43–2.01) | 0.85 |
| Mixed Medical-Surgical | 0.75 (0.59–0.94) | 0.01 | 0.79 (0.64–0.98) | 0.03 |
| Burn | 0.34 (0.22–0.54) | <0.001 | 0.66 (0.43–1.00) | 0.05 |
| Other | 0.81 (0.56–1.17) | 0.27 | 1.15 (0.83–1.59) | 0.41 |
| ICU pharmacist coverage 1st 24 hours‡ (reference: CMM delivered on interprofessional rounds) | - | - | - | - |
| CMM delivered not on interprofessional rounds | 0.93 (0.83–1.05) | 0.24 | 1.04 (0.93–1.16) | 0.50 |
| Abbreviated CMM delivered | 1.04 (0.90–1.19) | 0.61 | 1.07 (0.87–1.31) | 0.53 |
| CMM not delivered (absence of CMM) | 1.14 (0.97–1.33) | 0.11 | 1.17 (0.98–1.39) | 0.08 |
| Unknown | 1.32 (0.71–2.45) | 0.38 | 1.11 (0.62–1.99) | 0.74 |
| Nurse-to-patient ratio* (reference: 1:2) | - | - | - | - |
| ≤1 | 1.90 (1.52–2.37) | <0.001 | 1.41 (1.09–1.82) | 0.008 |
| 1-2 | 0.62 (0.37–1.03) | 0.06 | 0.75 (0.42–1.36) | 0.34 |
| >2 | 0.64 (0.47–0.85) | 0.003 | 0.82 (0.62–1.09) | 0.18 |
| Percent of assigned ICU teams pharmacist rounded with§ * | 1.00 (1.00–1.00) | 0.25 | 1.00 (1.00–1.00) | 0.02 |

*indicates that this is reflective of only the value on the first day of ICU stay

APP: advanced practice provider, CI: confidence interval, CMM: comprehensive medication management, CMS: Center for Medicare & Medicaid Services, ICU: intensive care unit, MRC-ICU: medication regimen complexity- intensive care unit, OR: odds ratio, SOFA: sequential organ failure assessment, VA: Veterans Affairs

**worst score during the first 24 hours of ICU stay

**†**Inclusion criteria specified ≥18 years of age; however, some institutions allowed patients ≥18 years of age in the pediatric ICU

**‡**CMM delivered on interprofessional rounds: the pharmacist attended multidisciplinary rounds and verbally provided CMM (including review of medications and recommendations); CMM delivered not on interprofessional rounds: the pharmacist provided by CMM (reviewed medications and provided recommendations) but did not attend interdisciplinary rounds; Abbreviated CMM delivered: the pharmacist provided abbreviated CMM which includes brief medication review but does not include full patient review (e.g. progress notes, results) and recommendations were provided outside of rounds; CMM not delivered (absence of CMM): the patient received no medication review from a pharmacist outside of pharmacokinetic monitoring and prospective medication order verification

**§**Percent of assigned ICU teams pharmacist rounded is calculated by total number of ICU medical teams the pharmacist attended and provided CMM on interprofessional rounds for divided by total number of ICU medical teams providing care for patients that the pharmacist was assigned to provide CMM for

#### eTable 29. Fine-Gray Competing Risks Model for ICU and Hospital Length of Stay with In-ICU Mortality as a Competing Event (N = 28,791)

| **Variable** | **Length of ICU Stay Competing Model (N = 28,791)** | | **Length of Hospital Stay Competing Model (N = 28,791)** | |
| --- | --- | --- | --- | --- |
|  | **Hazard Ratio (CI)** | **P-value** | **Hazard Ratio (CI)** | **P-value** |
| *Primary Variable* | | | | |
| Pharmacist-to-patient ratio* (ICU patients) | 1.00 (1.00, 1.00) | 0.03 | 1.00 (1.00, 1.00) | 0.09 |
| *Co-variates* | | | | |
| Age, years | 1.00 (1.00, 1.00) | 0.03 | 0.99 (0.99, 0.99) | <0.001 |
| Sex, female | 1.02 (0.99, 1.04) | 0.2 | 1.00 (0.97, 1.03) | 0.91 |
| SOFA Score** | 0.93 (0.92, 0.94) | <0.001 | 0.90 (0.89, 0.91) | <0.001 |
| MRC-ICU Score** | 0.98 (0.97, 0.98) | <0.001 | 0.98 (0.97, 0.98) | <0.001 |
| ICU admission day of the week (reference: Monday) | - | - | - | - |
| Tuesday | 1.01 (0.96, 1.05) | 0.8 | 1.01 (0.96, 1.06) | 0.67 |
| Wednesday | 0.99 (0.96, 1.03) | 0.79 | 1.00 (0.96, 1.04) | 0.89 |
| Thursday | 0.99 (0.96, 1.03) | 0.79 | 0.99 (0.94, 1.03) | 0.54 |
| Friday | 0.92 (0.87, 0.98) | 0.008 | 0.95 (0.90, 1.01) | 0.13 |
| Saturday | 0.94 (0.88, 1.01) | 0.11 | 0.98 (0.92, 1.05) | 0.62 |
| Sunday | 0.99 (0.94, 1.04) | 0.63 | 0.96 (0.91, 1.01) | 0.16 |
| Hospital type (reference: academic medical center) | - | - | - | - |
| Community - Teaching | 1.09 (0.99, 1.20) | 0.09 | 0.99 (0.90, 1.09) | 0.86 |
| Community – nonteaching | 0.83 (0.71, 0.95) | 0.01 | 1.80 (1.56, 2.09) | <0.001 |
| Government/VA/Military | 0.91 (0.75, 1.11) | 0.35 | 3.29 (2.44, 4.44) | <0.001 |
| Hospital CMS star rating (reference: 3) |  |  |  |  |
| 2 | 1.10 (0.99, 1.23) | 0.09 | 0.86 (0.77, 0.96) | 0.006 |
| 4 | 1.18 (1.08, 1.30) | <0.001 | 0.83 (0.75, 0.91) | <0.001 |
| 5 | 1.44 (1.05, 1.97) | 0.02 | 0.81 (0.69, 0.96) | 0.01 |
| Pharmacy trainee present* | 1.01 (0.96, 1.06) | 0.81 | 1.00 (0.95, 1.05) | 0.94 |
| Medical team coverage* (reference: only attending physician) | - | - | - | - |
| Attending physician and APP | 1.09 (1.00, 1.20) | 0.05 | 1.04 (0.94, 1.14) | 0.46 |
| Attending physician and medical residents/fellows | 1.04 (0.95, 1.15) | 0.39 | 1.04 (0.93, 1.16) | 0.49 |
| Attending physician and medical residents/fellows and APP | 1.14 (1.01, 1.28) | 0.03 | 1.06 (0.95, 1.20) | 0.3 |
| ICU type† * (reference: medical) | - | - | - | - |
| Surgical/Trauma | 0.92 (0.79, 1.07) | 0.27 | 1.07 (0.91, 1.25) | 0.42 |
| Surgical | 1.08 (0.91, 1.29) | 0.37 | 1.02 (0.87, 1.20) | 0.77 |
| Cardiothoracic | 1.16 (0.99, 1.37) | 0.07 | 1.61 (1.39, 1.87) | <0.001 |
| Cardiac | 0.91 (0.78, 1.06) | 0.24 | 1.05 (0.89, 1.23) | 0.55 |
| Neurosurgery/Neurology | 0.83 (0.71, 0.97) | 0.02 | 0.96 (0.85, 1.09) | 0.53 |
| Pediatric | 0.88 (0.64, 1.21) | 0.43 | 1.04 (0.77, 1.40) | 0.79 |
| Mixed Medical-Surgical | 1.26 (1.08, 1.46) | 0.002 | 1.05 (0.93, 1.20) | 0.43 |
| Burn | 0.58 (0.44, 0.76) | <0.001 | 0.82 (0.53, 1.27) | 0.37 |
| Other | 0.78 (0.46, 1.33) | 0.36 | 1.03 (0.65, 1.63) | 0.91 |
| ICU pharmacist coverage 1st 24 hours‡ (reference: CMM delivered on interprofessional rounds) | - | - | - | - |
| CMM delivered not on interprofessional rounds | 1.02 (0.96, 1.08) | 0.5 | 0.97 (0.91, 1.03) | 0.32 |
| Abbreviated CMM delivered | 1.01 (0.91, 1.12) | 0.88 | 0.95 (0.86, 1.04) | 0.25 |
| CMM not delivered (absence of CMM) | 0.83 (0.76, 0.90) | <0.001 | 0.88 (0.80, 0.96) | 0.006 |
| Unknown | 0.66 (0.54, 0.80) | <0.001 | 0.79 (0.56, 1.11) | 0.18 |
| Nurse-to-patient ratio* (reference: 1:2) | - | - | - | - |
| ≤1 | 0.88 (0.72, 1.07) | 0.19 | 0.84 (0.75, 0.94) | 0.003 |
| 1-2 | 0.76 (0.64, 0.90) | 0.002 | 0.84 (0.72, 0.99) | 0.03 |
| >2 | 1.45 (1.18, 1.79) | <0.001 | 1.09 (0.96, 1.23) | 0.17 |
| Percent of assigned ICU teams pharmacist rounded with§ * | 1.00 (1.00, 1.00) | 0.02 | 1.00 (1.00, 1.00) | 0.24 |

*indicates that this is reflective of only the value on the first day of ICU stay

APP: advanced practice provider, CI: confidence interval, CMM: comprehensive medication management, CMS: Center for Medicare & Medicaid Services, ICU: intensive care unit, MRC-ICU: medication regimen complexity- intensive care unit, OR: odds ratio, SOFA: sequential organ failure assessment, VA: Veterans Affairs

**worst score during the first 24 hours of ICU stay

**†**Inclusion criteria specified ≥18 years of age; however, some institutions allowed patients ≥18 years of age in the pediatric ICU

**‡**CMM delivered on interprofessional rounds: the pharmacist attended multidisciplinary rounds and verbally provided CMM (including review of medications and recommendations); CMM delivered not on interprofessional rounds: the pharmacist provided by CMM (reviewed medications and provided recommendations) but did not attend interdisciplinary rounds; Abbreviated CMM delivered: the pharmacist provided abbreviated CMM which includes brief medication review but does not include full patient review (e.g. progress notes, results) and recommendations were provided outside of rounds; CMM not delivered (absence of CMM): the patient received no medication review from a pharmacist outside of pharmacokinetic monitoring and prospective medication order verification

**§**Percent of assigned ICU teams pharmacist rounded is calculated by total number of ICU medical teams the pharmacist attended and provided CMM on interprofessional rounds for divided by total number of ICU medical teams providing care for patients that the pharmacist was assigned to provide CMM for

#### eTable 30. Fine-Gray Competing Risks Model for VFD with In-Hospital Mortality as a Competing Event and 28 days as a cut (N = 28789)

| **VFD Competing Model with cut (N = 28789)** | | | |
| --- | --- | --- | --- |
| **Variable** | | **Hazard Ratio (CI)** | **P-value** |
| *Primary Variable* | | | |
| Pharmacist-to-patient ratio* (ICU patients) | 1.00 (1.00, 1.00) | | 0.009 |
| *Covariates* | | | |
| Age, years | 1.00 (0.99, 1.00) | | <0.001 |
| Sex, female | 1.00 (0.97, 1.02) | | 0.76 |
| SOFA Score** | 0.90 (0.90, 0.91) | | <0.001 |
| MRC-ICU Score** | 0.97 (0.97, 0.97) | | <0.001 |
| ICU admission day of the week (reference: Monday) | - | | - |
| Tuesday | 1.00 (0.96, 1.04) | | 0.92 |
| Wednesday | 0.99 (0.96, 1.03) | | 0.69 |
| Thursday | 1.00 (0.96, 1.04) | | 0.97 |
| Friday | 0.98 (0.93, 1.03) | | 0.34 |
| Saturday | 0.95 (0.90, 1.00) | | 0.07 |
| Sunday | 0.95 (0.90, 0.99) | | 0.02 |
| ICU type† * (reference: medical ICU) | - | | - |
| Other | 0.96 (0.73, 1.28) | | 0.8 |
| Surgical | 1.25 (1.12, 1.40) | | <0.001 |
| Surgical/Trauma | 1.05 (0.98, 1.12) | | 0.19 |
| Cardiothoracic | 1.61 (1.44, 1.80) | | <0.001 |
| Cardiac | 1.07 (0.99, 1.15) | | 0.1 |
| Neurosurgery/Neurology | 0.94 (0.88, 1.00) | | 0.07 |
| Pediatric | 1.03 (0.81, 1.29) | | 0.83 |
| Mixed Medical-Surgical | 1.14 (1.05, 1.24) | | 0.003 |
| Burn | 1.04 (0.80, 1.35) | | 0.78 |
| Institution type (reference: academic medical center) | - | | - |
| Community - Teaching | 0.98 (0.92, 1.04) | | 0.53 |
| Community - nonteaching | 1.04 (0.95, 1.14) | | 0.38 |
| Government/VA/military | 1.20 (1.11, 1.29) | | <0.001 |
| Hospital CMS star rating (reference: 3) |  | |  |
| 2 | 1.02 (0.97, 1.08) | | 0.41 |
| 4 | 1.02 (0.97, 1.08) | | 0.4 |
| 5 | 1.07 (0.95, 1.22) | | 0.27 |
| ICU pharmacist coverage 1st 24 hours‡ (reference: CMM delivered on interprofessional rounds) | - | | - |
| CMM delivered not on interprofessional rounds | 1.01 (0.96, 1.05) | | 0.75 |
| Abbreviated CMM delivered | 1.01 (0.94, 1.08) | | 0.85 |
| CMM not delivered (absence of CMM) | 0.92 (0.86, 0.98) | | 0.008 |
| Unknown | 0.78 (0.66, 0.92) | | 0.003 |
| Nurse-to-patient ratio* (reference: 1:2) | - | | - |
| ≤1 | 0.89 (0.79, 1.02) | | 0.09 |
| 1-2 | 1.04 (0.85, 1.28) | | 0.7 |
| >2 | 1.21 (1.12, 1.32) | | <0.001 |
| Percent of assigned ICU teams pharmacist rounded with§* | 1.00 (1.00, 1.00) | | 0.001 |
| Pharmacy trainee present* | 1.00 (0.97, 1.04) | | 0.84 |
| Medical team coverage* (reference: attending physician only) | - | | - |
| Attending physician and APP | 1.09 (1.00, 1.18) | | 0.05 |
| Attending physician and medical residents/fellows | 1.06 (0.97, 1.16) | | 0.22 |
| Attending physician and medical residents/fellows and APP | 1.06 (0.96, 1.16) | | 0.26 |

*indicates that this is reflective of only the value on the first day of ICU stay

APP: advanced practice provider, CI: confidence interval, CMM: comprehensive medication management, CMS: Center for Medicare & Medicaid Services, ICU: intensive care unit, MRC-ICU: medication regimen complexity- intensive care unit, OR: odds ratio, SOFA: sequential organ failure assessment, VA: Veterans Affairs

**worst score during the first 24 hours of ICU stay

**†**Inclusion criteria specified ≥18 years of age; however, some institutions allowed patients ≥18 years of age in the pediatric ICU

**‡**CMM delivered on interprofessional rounds: the pharmacist attended multidisciplinary rounds and verbally provided CMM (including review of medications and recommendations); CMM delivered not on interprofessional rounds: the pharmacist provided by CMM (reviewed medications and provided recommendations) but did not attend interdisciplinary rounds; Abbreviated CMM delivered: the pharmacist provided abbreviated CMM which includes brief medication review but does not include full patient review (e.g. progress notes, results) and recommendations were provided outside of rounds; CMM not delivered (absence of CMM): the patient received no medication review from a pharmacist outside of pharmacokinetic monitoring and prospective medication order verification

**§**Percent of assigned ICU teams pharmacist rounded is calculated by total number of ICU medical teams the pharmacist attended and provided CMM on interprofessional rounds for divided by total number of ICU medical teams providing care for patients that the pharmacist was assigned to provide CMM for

#### eTable 31. Fine-Gray Competing Risks Model for VFD with In-Hospital Mortality as a Competing Event without a cut (N = 28789)

| **VFD Competing Model without cut (N = 28789)** | | |
| --- | --- | --- |
| **Variable** | **Hazard Ratio (CI)** | **P-value** |
| *Primary Variable* | | |
| Pharmacist-to-patient ratio* (ICU patients) | 1.00 (1.00, 1.00) | 0.009 |
| *Covariates* | | |
| Age, years | 1.00 (0.99, 1.00) | <0.001 |
| Sex, female | 1.00 (0.97, 1.02) | 0.76 |
| SOFA Score** | 0.90 (0.90, 0.91) | <0.001 |
| MRC-ICU Score** | 0.97 (0.97, 0.97) | <0.001 |
| ICU admission day of the week (reference: Monday) | - | - |
| Tuesday | 1.00 (0.96, 1.04) | 0.93 |
| Wednesday | 0.99 (0.96, 1.03) | 0.69 |
| Thursday | 1.00 (0.96, 1.04) | 0.97 |
| Friday | 0.98 (0.93, 1.03) | 0.34 |
| Saturday | 0.95 (0.90, 1.00) | 0.07 |
| Sunday | 0.95 (0.90, 0.99) | 0.02 |
| ICU type† * (reference: medical ICU) | - | - |
| Other | 0.96 (0.73, 1.28) | 0.80 |
| Surgical | 1.25 (1.12, 1.40) | <0.001 |
| Surgical/Trauma | 1.05 (0.98, 1.12) | 0.19 |
| Cardiothoracic | 1.61 (1.44, 1.80) | <0.001 |
| Cardiac | 1.07 (0.99, 1.15) | 0.10 |
| Neurosurgery/Neurology | 0.94 (0.88, 1.00) | 0.07 |
| Pediatric | 1.03 (0.81, 1.29) | 0.83 |
| Mixed Medical-Surgical | 1.14 (1.05, 1.24) | 0.003 |
| Burn | 1.04 (0.80, 1.35) | 0.77 |
| Institution type (reference: academic medical center) | - | - |
| Community - Teaching | 0.98 (0.92, 1.04) | 0.53 |
| Community - nonteaching | 1.04 (0.95, 1.14) | 0.38 |
| Government/VA/military | 1.20 (1.11, 1.29) | <0.001 |
| Hospital CMS star rating (reference: 3) |  |  |
| 2 | 1.02 (0.97, 1.08) | 0.41 |
| 4 | 1.02 (0.97, 1.08) | 0.40 |
| 5 | 1.07 (0.95, 1.22) | 0.27 |
| ICU pharmacist coverage 1st 24 hours‡ (reference: CMM delivered on interprofessional rounds) | - | - |
| CMM delivered not on interprofessional rounds | 1.01 (0.96, 1.05) | 0.75 |
| Abbreviated CMM delivered | 1.01 (0.94, 1.08) | 0.85 |
| CMM not delivered (absence of CMM) | 0.92 (0.86, 0.98) | 0.008 |
| Unknown | 0.78 (0.66, 0.92) | 0.003 |
| Nurse-to-patient ratio* (reference: 1:2) | - | - |
| ≤1 | 0.89 (0.79, 1.02) | 0.09 |
| 1-2 | 1.04 (0.85, 1.28) | 0.70 |
| >2 | 1.21 (1.12, 1.32) | <0.001 |
| Percent of assigned ICU teams pharmacist rounded with§* | 1.00 (1.00, 1.00) | 0.001 |
| Pharmacy trainee present* | 1.00 (0.97, 1.04) | 0.84 |
| Medical team coverage* (reference: attending physician only) | - | - |
| Attending physician and APP | 1.09 (1.00, 1.18) | 0.06 |
| Attending physician and medical residents/fellows | 1.06 (0.97, 1.16) | 0.22 |
| Attending physician and medical residents/fellows and APP | 1.06 (0.96, 1.16) | 0.27 |

*indicates that this is reflective of only the value on the first day of ICU stay

APP: advanced practice provider, CI: confidence interval, CMM: comprehensive medication management, CMS: Center for Medicare & Medicaid Services, ICU: intensive care unit, MRC-ICU: medication regimen complexity- intensive care unit, OR: odds ratio, SOFA: sequential organ failure assessment, VA: Veterans Affairs

**worst score during the first 24 hours of ICU stay

**†**Inclusion criteria specified ≥18 years of age; however, some institutions allowed patients ≥18 years of age in the pediatric ICU

**‡**CMM delivered on interprofessional rounds: the pharmacist attended multidisciplinary rounds and verbally provided CMM (including review of medications and recommendations); CMM delivered not on interprofessional rounds: the pharmacist provided by CMM (reviewed medications and provided recommendations) but did not attend interdisciplinary rounds; Abbreviated CMM delivered: the pharmacist provided abbreviated CMM which includes brief medication review but does not include full patient review (e.g. progress notes, results) and recommendations were provided outside of rounds; CMM not delivered (absence of CMM): the patient received no medication review from a pharmacist outside of pharmacokinetic monitoring and prospective medication order verification

**§**Percent of assigned ICU teams pharmacist rounded is calculated by total number of ICU medical teams the pharmacist attended and provided CMM on interprofessional rounds for divided by total number of ICU medical teams providing care for patients that the pharmacist was assigned to provide CMM for

#### Sensitivity Analysis- only mechanically ventilated patients

#### eTable 32. Patient Demographics for Mechanical Ventilation Analysis (N=14150)

|  | **Overall** | **Alive** | **Deceased** | **p-value** |
| --- | --- | --- | --- | --- |
|  | N = 14,150 | N = 10,967 | N = 3,183 |  |
| Percent of assigned ICU teams pharmacist rounded with**§** | 0.6 (0.3) | 0.5 (0.3) | 0.6 (0.3) | 0.014 |
| Missing | 160 (1.1) | 127 (1.2) | 33 (1.0) |  |
| Percent days, no pharmacist or pharmacist trainee on rounds | 31.8 (30.8) | 31.9 (31.2) | 31.4 (29.4) | 0.9 |
| Missing | 648 (4.6) | 461 (4.2) | 187 (5.9) |  |
| Percent days, only attending physician**⁂** | 7.5 (24.1) | 7.3 (23.7) | 8.4 (25.4) | 0.016 |
| Missing | 1,130 (8.0) | 833 (7.6) | 297 (9.3) |  |
| Any days with only a physician |  |  |  | 0.007 |
| No | 12,378 (88.7) | 9,654 (89.1) | 2,724 (87.4) |  |
| Yes | 1,571 (11.3) | 1,178 (10.9) | 393 (12.6) |  |
| Missing | 201 (1.4) | 135 (1.2) | 66 (2.1) |  |
| CMM every day of ICU stay |  |  |  | 0.001 |
| Yes | 9,957 (70.4) | 7,790 (71.0) | 2,167 (68.1) |  |
| No | 4,193 (29.6) | 3,177 (29.0) | 1,016 (31.9) |  |
| Age, years | 59.5 (16.7) | 58.3 (16.9) | 63.7 (15.6) | <0.001 |
| Sex |  |  |  | 0.6 |
| Male | 8,499 (60.1) | 6,606 (60.2) | 1,893 (59.5) |  |
| Female | 5,647 (39.9) | 4,357 (39.7) | 1,290 (40.5) |  |
| Unidentified | 3 (0.0) | 3 (0.0) | 0 (0.0) |  |
| Missing | 1 (<0.1) | 1 (<0.1) | 0 (0) |  |
| SOFA Score* | 7.2 (4.2) | 6.6 (3.9) | 9.4 (4.4) | <0.001 |
| Missing | 14 (<0.1) | 11 (0.1) | 3 (<0.1) |  |
| Dialysis |  |  |  | <0.001 |
| No | 12,036 (85.1) | 9,843 (89.8) | 2,193 (68.9) |  |
| Yes | 2,112 (14.9) | 1,124 (10.2) | 988 (31.1) |  |
| Missing | 2 (<0.1) | 0 (0) | 2 (<0.1) |  |
| ECMO |  |  |  | <0.001 |
| No | 13,844 (97.8) | 10,801 (98.5) | 3,043 (95.6) |  |
| Yes | 305 (2.2) | 166 (1.5) | 139 (4.4) |  |
| Missing | 1 (<0.1) | 0 (0) | 1 (<0.1) |  |
| Mechanical circulatory support |  |  |  | <0.001 |
| No | 4,583 (32.4) | 4,107 (37.5) | 476 (15.0) |  |
| Yes | 9,560 (67.6) | 6,855 (62.5) | 2,705 (85.0) |  |
| Missing | 7 (<0.1) | 5 (<0.1) | 2 (<0.1) |  |
| Mechanical ventilation |  |  |  | >0.9 |
| No | 0 (0.0) | 0 (0.0) | 0 (0.0) |  |
| Yes | 14,150 (100.0) | 10,967 (100.0) | 3,183 (100.0) |  |
| MRC-ICU Score* | 15.5 (6.2) | 15.2 (6.0) | 16.3 (6.9) | <0.001 |
| Missing | 1 (<0.1) | 1 (<0.1) | 0 (0) |  |
| ICU admission day of the week |  |  |  | 0.004 |
| Monday | 2,225 (15.7) | 1,722 (15.7) | 503 (15.8) |  |
| Tuesday | 2,177 (15.4) | 1,712 (15.6) | 465 (14.6) |  |
| Wednesday | 2,196 (15.5) | 1,746 (15.9) | 450 (14.1) |  |
| Thursday | 2,171 (15.3) | 1,688 (15.4) | 483 (15.2) |  |
| Friday | 1,964 (13.9) | 1,535 (14.0) | 429 (13.5) |  |
| Saturday | 1,714 (12.1) | 1,281 (11.7) | 433 (13.6) |  |
| Sunday | 1,703 (12.0) | 1,283 (11.7) | 420 (13.2) |  |
| Hospital LOS | 16.2 (20.1) | 16.5 (20.0) | 15.0 (20.2) | <0.001 |
| Missing | 3 (<0.1) | 3 (<0.1) | 0 (0) |  |
| ICU LOS | 8.4 (12.2) | 8.0 (11.8) | 9.9 (13.5) | <0.001 |
| ICU free days | 16.4 (10.5) | 20.9 (6.9) | 0.0 (0.0) | <0.001 |
| Missing | 218 (1.5) | 43 (0.4) | 175 (5.5) |  |
| Duration of mechanical ventilation | 4.8 (25.8) | 4.2 (28.9) | 6.8 (8.6) | <0.001 |
| Missing | 2 (<0.1) | 2 (<0.1) | 0 (0) |  |
| VFDs | 18.9 (11.3) | 24.4 (5.5) | 0.0 (0.0) | <0.001 |
| Missing | 2 (<0.1) | 2 (<0.1) | 0 (0) |  |
| Hospital Type |  |  |  | 0.70 |
| Academic medical center | 9,475 (67.0) | 7,330 (66.8) | 2,145 (67.4) |  |
| Community - Teaching | 3,590 (25.4) | 2,797 (25.5) | 793 (24.9) |  |
| Community - Non-teaching | 1,066 (7.5) | 827 (7.5) | 239 (7.5) |  |
| Government/VA/Military | 19 (0.1) | 13 (0.1) | 6 (0.2) |  |
| Hospital CMS star rating |  |  |  | <0.001 |
| 2 | 2,991 (21.5) | 2,311 (21.4) | 680 (22.2) |  |
| 3 | 7,226 (52.0) | 5,564 (51.4) | 1,662 (54.2) |  |
| 4 | 3,431 (24.7) | 2,737 (25.3) | 694 (22.6) |  |
| 5 | 235 (1.7) | 206 (1.9) | 29 (0.9) |  |
| Missing | 267 (1.9) | 149 (1.4) | 118 (3.7) |  |
| ICU type† |  |  |  | <0.001 |
| Medical | 3,875 (27.4) | 2,677 (24.4) | 1,198 (37.6) |  |
| Surgical/Trauma | 2,176 (15.4) | 1,801 (16.4) | 375 (11.8) |  |
| Cardiothoracic | 1,914 (13.5) | 1,745 (15.9) | 169 (5.3) |  |
| Cardiac | 1,005 (7.1) | 706 (6.4) | 299 (9.4) |  |
| Neurosurgery/Neurology | 1,863 (13.2) | 1,466 (13.4) | 397 (12.5) |  |
| Pediatric | 14 (0.1) | 12 (0.1) | 2 (0.1) |  |
| Mixed Medical-Surgical | 1,858 (13.1) | 1,367 (12.5) | 491 (15.4) |  |
| Burn | 72 (0.5) | 63 (0.6) | 9 (0.3) |  |
| Other | 5 (0.0) | 4 (0.0) | 1 (0.0) |  |
| Surgical | 1,368 (9.7) | 1,126 (10.3) | 242 (7.6) |  |
| Missing | 3 (<0.1%) | 3 (<0.1%) | 0 (0%) |  |
| ICU pharmacist coverage 1^st^ 24 hours |  |  |  | 0.03 |
| CMM delivered on interprofessional rounds | 9,640 (68.1) | 7,503 (68.4) | 2,137 (67.2) |  |
| CMM delivered not on interprofessional rounds | 1,901 (13.4) | 1,492 (13.6) | 409 (12.9) |  |
| Abbreviated CMM delivered | 1,013 (7.2) | 783 (7.1) | 230 (7.2) |  |
| CMM not delivered (absence of CMM) | 1,505 (10.6) | 1,118 (10.2) | 387 (12.2) |  |
| Unknown | 87 (0.6) | 68 (0.6) | 19 (0.6) |  |
| Missing | 4 (<0.1) | 3 (<0.1) | 1 (<0.1) |  |
| Pharmacist-to-patient ratio (all patients) | 27.3 (20.8) | 27.5 (21.3) | 26.6 (18.8) | 0.13 |
| Missing | 131 (0.9) | 101 (0.9) | 30 (0.9) |  |
| Pharmacist-to-patient ratio (ICU patients) | 19.8 (9.7) | 19.8 (9.7) | 19.7 (9.8) | 0.70 |
| Missing | 131 (0.9) | 101 (0.9) | 30 (0.9) |  |
| Nurse-to-patient ratio (average) |  |  |  | <0.001 |
| ≤1 | 995 (7.3) | 661 (6.3) | 334 (10.7) |  |
| =2 | 10,328 (75.6) | 8,198 (77.6) | 2,130 (68.5) |  |
| >2 | 259 (1.9) | 196 (1.9) | 63 (2.0) |  |
| 1-2 | 2,087 (15.3) | 1,504 (14.2) | 583 (18.7) |  |
| Missing | 481 (3.4) | 408 (3.7) | 73 (2.3) |  |

Data presented as mean (±SD) or n (%)

APP: advanced practice provider, CMM: comprehensive medication management, CMS: Center for Medicare & Medicaid Services, ECMO: extracorporeal membrane oxygenation, ICU: intensive care unit, LOS: length of stay, MRC-ICU, medication regimen complexity- intensive care unit, SOFA: sequential organ failure assessment, VA: Veterans Affairs, VFDs: ventilator free days

*worst score during the first 24 hours of ICU stay

**†**Inclusion criteria specified ≥18 years of age; however, some institutions allowed patients ≥18 years of age in the pediatric ICU

**‡**CMM delivered on interprofessional rounds: the pharmacist attended multidisciplinary rounds and verbally provided CMM (including review of medications and recommendations); CMM delivered not on interprofessional rounds: the pharmacist provided by CMM (reviewed medications and provided recommendations) but did not attend interdisciplinary rounds; Abbreviated CMM delivered: the pharmacist provided abbreviated CMM which includes brief medication review but does not include full patient review (e.g. progress notes, results) and recommendations were provided outside of rounds; CMM not delivered (absence of CMM): the patient received no medication review from a pharmacist outside of pharmacokinetic monitoring and prospective medication order verification

**§**Percent of assigned ICU teams pharmacist rounded is calculated by total number of ICU medical teams the pharmacist attended and provided CMM on interprofessional rounds for divided by total number of ICU medical teams providing care for patients that the pharmacist was assigned to provide CMM for

**⁂**Only attending physician refers to composition of the primary medical team. On these days, the patient received care from only the attending physician and did not receive care from any medical residents, fellows, or advanced practice providers.

#### eTable 33. Fine-Gray Competing Risks Model for VFD with In-Hospital Mortality as a Competing Event and 28 days as a cut (N = 14147)

| **VFD Competing Model with cut (N = 14147)** | | |
| --- | --- | --- |
| **Variable** | **Hazard Ratio (CI)** | **P-value** |
| *Primary variable* | | |
| Pharmacist-to-patient ratio (ICU patients) | 0.99 (0.99, 1.00) | 0.003 |
| *Secondary variable* | | |
| Absence of CMM for at least 1 day of ICU stay | 0.77 (0.70, 0.84) | <0.001 |
| *Covariates* | | |
| Age, years | 0.99 (0.99, 0.99) | <0.001 |
| Sex, female | 0.99 (0.99, 0.99) | <0.001 |
| SOFA Score* | 0.90 (0.88, 0.91) | <0.001 |
| MRC-ICU Score* | 1.00 (1.00, 1.01) | 0.27 |
| ICU admission day of the week (reference: Monday) | - | - |
| Tuesday | 1.01 (0.95, 1.08) | 0.72 |
| Wednesday | 1.09 (1.02, 1.17) | 0.01 |
| Thursday | 1.02 (0.95, 1.10) | 0.58 |
| Friday | 0.97 (0.88, 1.06) | 0.46 |
| Saturday | 0.91 (0.82, 1.01) | 0.08 |
| Sunday | 0.88 (0.80, 0.97) | 0.01 |
| ICU type† (reference: medical ICU) | - | - |
| Other | 1.10 (0.46, 2.59) | 0.83 |
| Surgical | 1.69 (1.33, 2.15) | <0.001 |
| Surgical/Trauma | 1.30 (1.14, 1.49) | <0.001 |
| Cardiothoracic | 3.20 (2.47, 4.13) | <0.001 |
| Cardiac | 1.16 (0.98, 1.36) | 0.08 |
| Neurosurgery/Neurology | 1.01 (0.86, 1.18) | 0.92 |
| Pediatric | 1.20 (0.81, 1.77) | 0.37 |
| Mixed Medical-Surgical | 1.13 (0.95, 1.35) | 0.16 |
| Burn | 1.04 (0.82, 1.32) | 0.74 |
| Hospital type (reference: academic medical center) | - | - |
| Community - Teaching | 1.16 (1.00, 1.35) | 0.06 |
| Community - nonteaching | 1.05 (0.88, 1.26) | 0.61 |
| Government/VA/military | 0.97 (0.62, 1.50) | 0.88 |
| Hospital CMS star rating (reference: 3) |  |  |
| 2 | 0.96 (0.84, 1.11) | 0.61 |
| 4 | 1.14 (0.97, 1.35) | 0.10 |
| 5 | 1.37 (1.05, 1.77) | 0.02 |
| ICU pharmacist coverage 1st 24 hours‡ (reference: CMM delivered on interprofessional rounds) | - | - |
| CMM delivered not on interprofessional rounds | 1.02 (0.94, 1.12) | 0.58 |
| Abbreviated CMM delivered | 0.98 (0.87, 1.10) | 0.73 |
| CMM not delivered (absence of CMM) | 1.05 (0.95, 1.16) | 0.32 |
| Unknown | 0.72 (0.52, 1.00) | 0.05 |
| Nurse-to-patient ratio (average)(reference: 1:2) | - | - |
| ≤1 | 0.64 (0.48, 0.85) | 0.002 |
| 1-2 | 0.69 (0.62, 0.77) | <0.001 |
| >2 | 0.92 (0.80, 1.06) | 0.23 |
| Percent of assigned ICU teams pharmacist rounded with§ | 0.88 (0.72, 1.07) | 0.19 |
| Percent days, no pharmacist or pharmacist trainee on rounds | 1.00 (1.00, 1.00) | 0.73 |
| Percent days, only attending physician⁂ | 1.00 (1.00, 1.00) | 0.10 |

APP: advanced practice provider, CI: confidence interval, CMM: comprehensive medication management, CMS: Center for Medicare & Medicaid Services, ICU: intensive care unit, MRC-ICU: medication regimen complexity- intensive care unit, OR: odds ratio, SOFA: sequential organ failure assessment, VA: Veterans Affairs

**†**Inclusion criteria specified ≥18 years of age; however, some institutions allowed patients ≥18 years of age in the pediatric ICU

**‡**CMM delivered on interprofessional rounds: the pharmacist attended multidisciplinary rounds and verbally provided CMM (including review of medications and recommendations); CMM delivered not on interprofessional rounds: the pharmacist provided by CMM (reviewed medications and provided recommendations) but did not attend interdisciplinary rounds; Abbreviated CMM delivered: the pharmacist provided abbreviated CMM which includes brief medication review but does not include full patient review (e.g. progress notes, results) and recommendations were provided outside of rounds; CMM not delivered (absence of CMM): the patient received no medication review from a pharmacist outside of pharmacokinetic monitoring and prospective medication order verification

**§**Percent of assigned ICU teams pharmacist rounded is calculated by total number of ICU medical teams the pharmacist attended and provided CMM on interprofessional rounds for divided by total number of ICU medical teams providing care for patients that the pharmacist was assigned to provide CMM for

**⁂**Only attending physician refers to composition of the primary medical team. On these days, the patient received care from only the attending physician and did not receive care from any medical residents, fellows, or advanced practice providers.

#### eTable 34. Fine-Gray Competing Risks Model for VFD with In-Hospital Mortality as a Competing Event without a cut (N = 14147)

| **VFD Competing Model with cut (N = 14147)** | | |
| --- | --- | --- |
| **Variable** | **Hazard Ratio (CI)** | **P-value** |
| *Primary variable* | | |
| Pharmacist-to-patient ratio (ICU patients) | 0.99 (0.99, 1.00) | 0.003 |
| *Secondary variable* | | |
| Absence of CMM for at least 1 day of ICU stay | 0.77 (0.70, 0.84) | <0.001 |
| *Covariates* | | |
| Age, years | 0.99 (0.99, 0.99) | <0.001 |
| Sex, female | 0.99 (0.95, 1.03) | 0.64 |
| SOFA Score* | 0.90 (0.88, 0.91) | <0.001 |
| MRC-ICU Score* | 1.00 (1.00, 1.01) | 0.27 |
| ICU admission day of the week (reference: Monday) | - | - |
| Tuesday | 1.01 (0.95, 1.08) | 0.72 |
| Wednesday | 1.09 (1.02, 1.17) | 0.01 |
| Thursday | 1.02 (0.95, 1.10) | 0.58 |
| Friday | 0.97 (0.88, 1.06) | 0.46 |
| Saturday | 0.91 (0.82, 1.01) | 0.08 |
| Sunday | 0.88 (0.80, 0.97) | 0.01 |
| ICU type† (reference: medical ICU) | - | - |
| Other | 1.10 (0.46, 2.59) | 0.83 |
| Surgical | 1.69 (1.33, 2.15) | <0.001 |
| Surgical/Trauma | 1.30 (1.14, 1.49) | <0.001 |
| Cardiothoracic | 3.20 (2.47, 4.13) | <0.001 |
| Cardiac | 1.16 (0.98, 1.36) | 0.08 |
| Neurosurgery/Neurology | 1.01 (0.86, 1.18) | 0.92 |
| Pediatric | 1.20 (0.81, 1.77) | 0.37 |
| Mixed Medical-Surgical | 1.13 (0.95, 1.35) | 0.16 |
| Burn | 1.04 (0.82, 1.32) | 0.74 |
| Hospital type (reference: academic medical center) | - | - |
| Community - Teaching | 1.16 (1.00, 1.35) | 0.06 |
| Community - nonteaching | 1.05 (0.88, 1.26) | 0.61 |
| Government/VA/military | 0.97 (0.62, 1.50) | 0.88 |
| Hospital CMS star rating (reference: 3) | - | - |
| 2 | 0.96 (0.84, 1.11) | 0.61 |
| 4 | 1.14 (0.97, 1.35) | 0.10 |
| 5 | 1.37 (1.05, 1.77) | 0.02 |
| ICU pharmacist coverage 1st 24 hours‡ (reference: CMM delivered on interprofessional rounds) | - | - |
| CMM delivered not on interprofessional rounds | 1.02 (0.94, 1.12) | 0.58 |
| Abbreviated CMM delivered | 0.98 (0.87, 1.10) | 0.73 |
| CMM not delivered (absence of CMM) | 1.05 (0.95, 1.16) | 0.32 |
| Unknown | 0.72 (0.52, 1.00) | 0.05 |
| Nurse-to-patient ratio (average)(reference: 1:2) | - | - |
| ≤1 | 0.64 (0.48, 0.85) | 0.002 |
| 1-2 | 0.69 (0.62, 0.77) | <0.001 |
| >2 | 0.92 (0.80, 1.06) | 0.23 |
| Percent of assigned ICU teams pharmacist rounded with§ | 0.88 (0.72, 1.07) | 0.19 |
| Percent days, no pharmacist or pharmacist trainee on rounds | 1.00 (1.00, 1.00) | 0.73 |
| Percent days, only attending physician⁂ | 1.00 (1.00, 1.00) | 0.10 |

APP: advanced practice provider, CI: confidence interval, CMM: comprehensive medication management, CMS: Center for Medicare & Medicaid Services, ICU: intensive care unit, MRC-ICU: medication regimen complexity- intensive care unit, OR: odds ratio, SOFA: sequential organ failure assessment, VA: Veterans Affairs

**†**Inclusion criteria specified ≥18 years of age; however, some institutions allowed patients ≥18 years of age in the pediatric ICU

**‡**CMM delivered on interprofessional rounds: the pharmacist attended multidisciplinary rounds and verbally provided CMM (including review of medications and recommendations); CMM delivered not on interprofessional rounds: the pharmacist provided by CMM (reviewed medications and provided recommendations) but did not attend interdisciplinary rounds; Abbreviated CMM delivered: the pharmacist provided abbreviated CMM which includes brief medication review but does not include full patient review (e.g. progress notes, results) and recommendations were provided outside of rounds; CMM not delivered (absence of CMM): the patient received no medication review from a pharmacist outside of pharmacokinetic monitoring and prospective medication order verification

**§**Percent of assigned ICU teams pharmacist rounded is calculated by total number of ICU medical teams the pharmacist attended and provided CMM on interprofessional rounds for divided by total number of ICU medical teams providing care for patients that the pharmacist was assigned to provide CMM for

**⁂**Only attending physician refers to composition of the primary medical team. On these days, the patient received care from only the attending physician and did not receive care from any medical residents, fellows, or advanced practice providers.
